# Proteomic analysis of blood neuronal and glial extracellular vesicles reveals neuroprotective effects of the angiotensin type-1 blocker candesartan in Parkinson’s disease patients

**DOI:** 10.1101/2025.05.02.25326639

**Authors:** L Camacho-Meño, CM Labandeira, SB Bravo, M.V Torres, H Bejr-Kasem, A Molina-Crespo, M Atienza, JL Lanciego, JL Cantero, J Kulisevsky, JL Labandeira-García, AI Rodríguez-Perez

**Author notes:** Correspondence: Jose L. Labandeira-Garcia M.D., Ph.D., or Ana I. Rodriguez-Perez Ph.D., Research Center for Molecular Medicine and Chronic diseases (CIMUS), University of Santiago de Compostela, 15782 Santiago de Compostela, Spain. E. mail. These authors contributed equally to this work (L Camacho-Meño, CM Labandeira).

## Abstract

**Background:** The renin-angiotensin system (RAS) is a major regulator of cell homeostasis, oxidative stress, and inflammatory responses in different tissues, including the brain. In Parkinson’s disease (PD) models, activation of angiotensin type-1 receptors (AT1) increased the vulnerability of dopaminergic neurons, which was inhibited by AT1 blockers (ARBs). This is consistent with recent studies in human brains showing the highest vulnerability of the dopaminergic neurons with the highest rate of the AT1 gene (*AGTR1*) expression, and retrospective cohort studies showing a reduction of PD risk in patients receiving ARB treatment. Neuroprotective effects of ARBs have also been suggested for Alzheimer’s disease.

**Methods:** We recently performed a randomized phase-II 28-week clinical trial of the ARB candesartan in PD. However, the molecular changes induced by ARBs with BBB-penetrating properties such as candesartan in the brain of PD patients remain unexplored. In PD patients, we used a minimally invasive approach to characterize cell– type–specific molecular changes induced by candesartan in the brain. We obtained extracellular vesicles (EVs) from neurons and different types of glial cells within the same blood sample, and the protein content was compared before and after candesartan treatment.

**Results:** Candesartan induced the upregulation or downregulation of many proteins relevant to PD progression. These changes were detected in EVs derived from neurons (46 dysregulated proteins), microglia/macrophages (48 dysregulated proteins), astrocytes (22 dysregulated proteins), and oligodendrocytes (92 dysregulated proteins). Altogether, the proteomic changes indicate a coordinated neuroprotective response, involving improved redox and mitochondrial function, enhanced proteostasis and gene regulation, and attenuation of synaptic, metabolic, and inflammatory stress pathways.

**Conclusion:** Our proteomic analysis indicates the neuroprotective effects of ARBs such as candesartan on neurons and glial cells in PD patients, highlighting its therapeutic potential and suggesting the need of larger clinical trials for repurposing of these drugs. The results also reveal the potential of the present minimally invasive approach to detect brain cell-type–specific molecular changes and points to a shift in neurodegenerative disease research and monitoring.

## Background

The renin-angiotensin system (RAS) was initially associated with blood pressure regulation. However, RAS was later identified as a major regulator of cell homeostasis, oxidative stress, and inflammatory responses in many tissues. The tissue RAS exerts its functional effects through two opposing axes: The first axis, primarily formed by angiotensin II (AngII) binding to angiotensin type 1 (AT1) receptors, promotes inflammatory, oxidative, vasoconstrictive, fibrotic, and thrombotic effects. The second or compensatory axis, formed by AngII acting on AT2 receptors and angiotensin 1-7 (Ang1-7) acting on Mas receptors (MasR), counteracts the effects of AT1 activation [1, 2], and both axes must be correctly balanced under physiological conditions. However, this balance may be disrupted under pathological circumstances that favor the AT1-related pro-oxidative, pro-inflammatory axis. In the brain, all major components of the RAS have been observed in rodents and primates, including humans [3–5]. For more than a decade, our laboratory and several others have shown the presence and functional effects of major RAS components in the nigrostriatal neurons and glial cells using in vivo and in vitro models of Parkinson’s disease (PD; see for review [6]). Interestingly, an intracellular RAS was also observed in cells, particularly dopaminergic neurons, which, in addition to plasma membrane RAS receptors, showed different RAS receptors in intracellular compartments such as the nucleus and the mitochondria [2, 7].

In a series of studies in animal and vitro models, we observed that activation of AT1 receptors increased the vulnerability of dopaminergic neurons that was inhibited by AT1 receptor blockers (ARBs), and that ARBs that were able to cross the brain-blood barrier (BBB) such as candesartan or telmisartan were particularly effective in animal models [9, 10]. In PD models, activation of AT1 receptors exacerbated major mechanisms involved in dopaminergic neuron degeneration and PD progression, which was inhibited by ARB treatment or activation of components of the compensatory RAS system [11–13].

Recent studies in humans are consistent with the results in experimental models. Using single-nucleus RNA sequencing and unbiased clustering analysis, Kamath et al. [5] showed that the most vulnerable population of nigral dopaminergic neurons expressed the highest rate of the AT1 receptor gene (*AGTR1*), which was confirmed by several additional studies [14, 15]. Furthermore, recent retrospective cohort studies showed that ARB treatment was associated with a marked reduction of PD risk in hypertensive patients, and that ARBs with BBB-penetrating properties and a high cumulative duration of treatment were particularly effective [16, 17]. We have recently performed a randomized phase-II 28-week clinical trial of candesartan to explore the effects on cognitive impairment (CI) in PD patients [18]. Results showed that candesartan treatment was safe and well-tolerated and revealed improvement in apathy, a highly debilitating non-motor symptom experienced by PD patients [19]. However, to our knowledge, *in vivo* molecular brain changes induced by the administration of candesartan remain unexplored in PD patients. Interestingly, neuroprotective effects of ARBs have also been suggested for Alzheimer’s disease [20, 21].

Extracellular vesicles (EVs) are lipid bilayer-enclosed nanoparticles secreted by virtually all cell types, which mediate intercellular communication through the transfer of proteins, lipids, and nucleic acids [22]. Small EVs can cross the BBB and, owing to their biogenesis, their molecular cargo reflects the physiological state of their origin cell [23]. These properties position EVs as a powerful tool for investigating central nervous system (CNS) processes through peripheral biofluids. In the present study, we isolated EVs enriched from neurons, astrocytes, oligodendrocytes, and microglia/macrophage cell origin in blood samples from five randomly selected PD patients of the above-mentioned clinical trial, before and after the candesartan treatment, which were then subjected to proteomic analysis to identify molecular effects induced by candesartan in the patient brain. We observed remarkable effects of candesartan on proteins and cellular processes involved in dopaminergic degeneration and PD progression. Notably, this minimally invasive approach enabled the characterization of CNS cell type–specific molecular changes induced by the administration of ARBs such as candesartan, potentially constituting a paradigm shift in the investigation and clinical monitoring of neurodegenerative disorders.

## Methods

### Study Design and Participants

This study analyzed serum samples from five patients diagnosed with Parkinson’s disease (PD) (two females and three males) who were recruited from a single-center, 28-week, prospective, randomized, double-blind, placebo-controlled, parallel-group phase II clinical trial conducted at the Movement Disorders Unit of the Sant Pau University Hospital in Barcelona. The trial aimed to evaluate the effects of candesartan compared to placebo in PD patients [18]. The study protocol was approved by the Ethics Committee of Sant Pau University Hospital, Barcelona, and was registered in the European Clinical Trials Register under EudraCT number 2016-000679-25 (https://www.clinicaltrialsregister.eu/ctr-search/trial/2016-000679-25/ES ). The trial was conducted following the Declaration of Helsinki and the guidelines of Good Clinical Practice. PD patients were included if they met the following inclusion criteria: neuroimaging evidence within the last 18 months compatible with PD; Hoehn and Yahr (H&Y) stage I-III; stable dopaminergic treatment for at least four weeks before enrollment; and Montreal Cognitive Assessment (MoCA) scores of ≥20 and ≤25 for PD patients with mild cognitive impairment or dementia, or ≥26 for those PD patients with subjective cognitive decline.

Exclusion criteria included illiteracy; significant visual or hearing impairment; any severe and/or uncontrolled medical condition deemed clinically relevant for study participation; uncontrolled motor fluctuations or disabling dyskinesia; history of deep brain stimulation; severe white matter involvement on CT or MRI; active psychosis or major hallucinations; severe depression or delirium; or past or current drug or alcohol abuse. Additional exclusion criteria specific to the investigational medication included symptomatic orthostatic hypotension, hypovolemia, or contraindications for antihypertensive drugs; dependence on the renin-angiotensin system (e.g., heart failure class III or IV according to the NYHA classification, unilateral or bilateral renal artery stenosis); current or prior use of ARBs, angiotensin-converting enzyme inhibitors, or potassium-sparing diuretics; previous treatment with candesartan; hereditary galactose intolerance, glucose-galactose malabsorption; and concomitant use of lithium, acetylcholinesterase inhibitors, anticholinergic agents, or dopamine receptor blockers. Patients initiated candesartan treatment at a dose of 4 mg/day for the first four weeks. The dose was then titrated to 8 mg/day for the remainder of the study. In cases of adverse events, the study physician could reduce the dose back to 4 mg/day. Treatment was maintained at the target dose until week 24, after which it was tapered to 4 mg/day for four weeks and discontinued at the end of week 28. The specific characteristics of the patients enrolled in this study can be found in Supplemental Table S1 (in Supplementary Material).

Blood samples were obtained from each patient before the initiation of treatment and upon its completion. Venipuncture was performed, and blood was collected in Vacutainer SST II Advance Serum Separator Gel tubes (8.5 mL; Ref. 366468). The samples were centrifuged at 1,500 × g for 10 minutes to isolate serum. The resulting serum samples were immediately frozen and stored at −80°C until further analysis. For this study, we analyzed a total of 10 serum samples from 5 PD patients: 5 samples collected before initiating candesartan treatment and 5 obtained at the end of the six-month treatment period.

### Isolation of total extracellular vesicles using size exclusion chromatography

Total EVs were isolated from serum samples that had undergone centrifugation at 10,000 × g, for 10 minutes at 4°C to ensure the removal of debris. For EV isolation, qEV Original 70 nm Gen2 columns were pre-equilibrated with 17 mL of filtered 1X PBS (0.22 µm filter) before loading a 500 µL sample. A buffer volume of 2.9 mL was used to discard the void volume before EV collection. Six fractions of 0.4 mL each were collected using an Automatic Fraction Collector (AFC) (IZON SCIENCE LTD, New Zealand). Then, the EV-enriched fractions were pooled and concentrated using 100 kDa ultrafiltration devices (Amicon Ultra, Millipore) by centrifugation at 12,000 × g at 4°C until reaching the desired volume (200 μL). The concentrated EVs were finally transferred to a sterile tube and stored at −80°C until further analysis. For each individual and time point, two independent 500 µL samples were processed separately and subsequently pooled.

### Isolation of brain**□**derived extracellular vesicles

From an initial 100 µL concentrate of total EVs isolated from 1 mL of serum, a sequential separation was performed to obtain EVs derived from different brain cell types, including neurons, astrocytes, oligodendrocytes, and microglia. Initially, 100 µL of total EVs were incubated overnight at 4°C under continuous agitation with 4 µg of L1CAM (L1 Cell Adhesion Molecule )/CD171-Biotin (Ref. 13-1919-82, ThermoFisher Scientific) in 300 µL of PBS containing 1.3% BSA for the isolation of neuronal EVs (nEVs). The following day, 200 µL of streptavidin magnetic beads (ThermoFisher Scientific, Waltham, MA, USA) were added and incubated at room temperature for 3 hours under agitation. After incubation, the immunocomplexes (containing nEVs) were captured using a magnetic separator, while the supernatant (depleted of nEVs) was collected for subsequent extractions of specific EV subpopulations.

To remove unbound components, the bead-bound nEV immunocomplexes were washed three times with 1000 µL of PBS containing 0.1% BSA. The immunocomplexes were then resuspended in 100 µL of filtered PBS elution buffer and immediately frozen at -80°C for further characterization and analysis.

The supernatant was sequentially processed to isolate additional brain cell-origin EV subpopulations. First, it was incubated with 4 µg of biotinylated Glast (Glutamate Aspartate Transporter)-antibody (Ref. NB100-1869B, Novus Biologicals) to isolate astrocyte-derived EVs (aEVs), following the same workflow. Next, the remaining supernatant was incubated with 4 µg of biotinylated Human/Mouse MOG (Myelin Oligodendrocyte Glycoprotein)-antibody (Ref. BAM2439, R&D Systems) for oligodendrocyte-derived EVs (oEVs) isolation. Finally, the residual supernatant underwent incubation with 4 µg of biotinylated TMEM119 (Transmembrane protein 119)-antibody (Ref. 1023426, Novus Biologicals) to isolate microglia/macrophage-derived EVs (m/mEVs). Each step involved incubation with streptavidin magnetic beads, magnetic separation, and collection of the supernatant for the next extraction. A diagram illustrating the steps involved in isolating EVs enriched from different brain cell types is presented in Figure S1.

### Extracellular vesicles characterization

#### Confirmation of the presence of EV marker proteins

Total EVs isolated from 1 mL of serum, along with serum samples, were analyzed using specific ELISA assays to detect established EV markers: CD81 (Ref. 18095935), CD63 (Ref. 16853020), and ALIX (Ref. 30240446). Calnexin (Ref. 18234959), an endoplasmic reticulum resident protein generally absent from EVs, was included as a negative control to assess potential non-vesicular contamination.

All ELISA kits were sourced from Invitrogen. Serum samples were processed according to the manufacturer’s instructions, and assays with EVs were performed by resuspending 50 µL of total EVs in 50 µL of RIPA buffer, followed by four cycles of sonication (10 seconds each, 10% amplitude) on ice. The samples were then centrifuged at 10,000 × g for 10 minutes at 4°C. A 6 µL aliquot of the supernatant was used for total protein quantification via the Pierce BCA Protein Assay, while 90 µL were loaded onto the corresponding ELISA plates.

From this point, all procedures were strictly followed according to the manufacturer’s instructions. Absorbance was measured at 450/620 nm using an Infinite M200 multiwell plate reader (TECAN, Männedorf, Switzerland). Tetraspanin and calnexin concentrations were quantified by extrapolating from a standard curve generated using a four-parameter logistic (4PL) curve fit.

#### Nanoparticle Tracking analysis (NTA)

Quantification of the hydrodynamic diameter distribution and concentration of extracellular vesicles was performed using the NanoSight PRO (Malvern Instruments) equipped with a blue (488 nm) laser and a high-sensitivity camera. Samples were diluted 1:10.000 in filtered PBS to achieve a particle concentration within the optimal detection range (1 × 10□ – 1 × 10□ particles/mL). Each sample was measured by recording two 60-second videos at a detection threshold of exposure time 13, contrast gain 3, 0. Automatic settings were applied for blur, minimum track length, and maximum jump distance. The collected videos were analyzed using NTA software (NanoSight NTA 3.4), providing particle size distribution and mean, mode, and median particle diameters. Data were expressed as mean particle size (nm) ± standard deviation and particle concentration (particles/mL).

#### Transmission electron microscopy (TEM)

Morphology, size, and structural integrity were evaluated by transmission electron microscopy (TEM). At this end, enriched EVs were diluted 1:10 in distilled water. A 10 μL aliquot of the diluted sample was placed onto a carbon-coated copper grid and allowed to adsorb for 1 min. Excess liquid was carefully removed using filter paper. The grid was then placed on a drop of 1% phosphotungstic acid solution prepared in Milli-Q® water for 1 min, followed by blotting to remove excess stain. Finally, EV morphology was examined using a JEOL JEM 2010 transmission electron microscope operating at 200 kV

#### Characterization of brain cell-origin EV subpopulations

To assess neuronal enrichment, neuron-specific enolase (NSE), a glycolytic enzyme predominantly expressed in neurons, was quantified in nEVs using the human neuron specific Enolase ELISA Kit (Ref. ab217778, Abcam). Astrocytic enrichment was evaluated using the Glial Fibrillary Acidic Protein (GFAP) ELISA kit (Ref. EEL079, Invitrogen), as GFAP is a specific intermediate filament protein predominantly expressed in astrocytes.

For the assessment of oligodendrocytic and microglial enrichment, the Human Myelin Basic Protein (MBP) ELISA kit (Ref. MBS2502574, MyBioSource) and the Ionized Calcium-Binding Adapter Molecule 1 (IBA-1) ELISA kit (Ref. ABIN6953617, Antibodies Online) were used, respectively. MBP, a key structural component of the myelin sheath, is exclusively expressed by oligodendrocytes and serves as a well-established marker for oligodendrocyte-derived EVs. IBA-1, a cytoplasmic protein involved in actin remodeling and phagocytosis, is specifically expressed in microglia, making it a reliable marker for microglia-derived EVs.

For each ELISA assay, 50 µL of EV samples from the different subpopulations were resuspended in 50 µL of RIPA buffer and subjected to four sonication cycles (10 seconds each, 10% amplitude) on ice. The samples were then centrifuged at 10,000 × g for 10 minutes at 4°C. A 6 µL aliquot of the supernatant was used to determine total protein concentration using the Pierce BCA Protein Assay, while 90 µL were loaded onto the corresponding ELISA plates.

From this point, all assays were performed strictly following the manufacturer’s instructions. Absorbance was measured at 450/620 nm using an Infinite M200 multiwell plate reader (TECAN, Männedorf, Switzerland). Protein marker concentrations were quantified by extrapolation from a four-parameter logistic (4PL) standard curve.

### Quantitative Proteomics Analysis by SWATH

Proteomic analysis was performed as previously described [24, 25]. In brief, 15 μg of EVs proteins were mixed with a lysis buffer in a ratio of 1:1, boiled and concentrated in a resolving 10% SDS-PAGE gel [26]. The protein band was stained (Sypro Ruby fluorescent staining; Lonza, Basel, Switzerland) and excised. Gel pieces were reduced (10 mM DTT; Sigma-Aldrich, St. Louis, MO, USA) and alkylated (55 mM iodoacetamide; Sigma-Aldrich, St. Louis, MO, USA). Then, we performed in-gel tryptic digestion, as described elsewhere [27]. Peptides were extracted [50% ACN/0.1% TFA (×3) and ACN (×1)], pooled, concentrated in a SpeedVac, and stored at −20 °C. For label-free quantitative proteomics, we performed an MS analysis by sequential window acquisition of all theoretical mass spectra (SWATH-MS), as previously described [28, 29] . First, 6 peptide pooles were created by mixing equal amounts of peptides from each sample type (MOG, TMEN, L1CAM, GLAST, total sEV and Plasma). These pooles were analysed by LC-MS/MS on a TripleTOF® 6600 LC-MS/MS system via a data-dependent acquisition (DDA) method in order to create a SWATH-MS spectral library. Only proteins and peptides with <1% FDR were included in this library [30]. Then, 4 µg of peptides derived from the individual samples from MOG, TMEN, L1CAM, GLAST, total sEV and serum after and before candesartan treatment were analysed by SWATH-MS method. SWATH-MS acquisition was performed on a TripleTOF® 6600 LC-MS/MS system via a data-independent acquisition (DIA) method. The whole 400 to 1250 m/z range was covered in 100 steps with spectral windows of variable width (1 m/z overlap). Peaks extraction was carried out with PeakView software (version 2.2; Sciex, Redwood City, CA, USA) and scored using the PeakView SWATH Acquistion MicroApp (version 2.0; Sciex, Redwood City, CA, USA). The integrated peak areas were exported to the MarkerView software (version 1.3, Sciex, Redwood City, CA, USA; accessed on 3 September 2022). To ensure a more accurate comparison between samples, well-known endogenous peptides were used during data alignment to compensate for small variations in both mass and retention times. The amount of each protein in every EVs sample was calculated as the averaged area sums of 10 peptides per protein and 7 transitions per peptide. Then, an averaged MS peak area of each protein was calculated. Data normalisation was carried out with the most likely ratio normalisation (MLR) method. As part of the initial analysis, the MarkerView software (version 1.3, Sciex, Redwood City, CA, USA) also allowed a principal component analysis (PCA) to see how well each protein distinguishes between groups. The mass spectrometry proteomics data have been deposited in the ProteomeXchange Consortium via the PRIDE (31) partner repository with the dataset identifier PXD063198. R (version 4.1.2) was used to perform all bioinformatic analyses. The data was log-transformed, quantile normalized, and differentially enriched proteins were identified using linear models (limma, version 3.50.3). Proteins were considered differentially enriched if *P* < 0.05 and |log_2_ FC| > 1; FC > 2. Heatmaps and volcano plots were generated using heatmap3 (version 1.1.9), GraphPad Prism 8 (GraphPad, Inc., San Diego, CA, USA), and Glimma (version 2.4.0) packages. Functional annotation was performed using clusterProfiler (version 4.2.2) and rrvgo (version 1.6.0) packages. Identified proteins were analyzed using STRING v[12.0] (https://string-db.org) to evaluate predicted protein–protein interactions. The analysis was performed for *Homo sapiens* with a minimum required interaction score of 0.7 (high confidence). Gene Ontology (GO) enrichment analysis was conducted within STRING. Interaction networks were visualized and exported for further interpretation. More detailed information about the method can be found in Supplementary Methods (in Supplementary Material).

All information regarding the function, subcellular localization, and tissue-specific expression of differentially expressed proteins identified in extracellular vesicles was obtained from UniProt (https://www.uniprot.org) and the Human Protein Atlas (https://www.proteinatlas.org).

## RESULTS

### Characterization of the obtained extracellular vesicles

Nanoparticle tracking analysis (NTA; Figs. 1A–B) confirmed the presence of particles with a mean diameter of 115.6 nm ± 1.965 (SEM) in EVs isolated before candesartan treatment and 119.2 nm ± 2.354 (SEM) in EVs isolated after treatment. A paired t-test revealed no statistically significant difference in EV size before and after candesartan administration. Similarly, no differences were observed in EV concentration between treatment conditions (3.764 × 10¹² particles/mL± 2.62×10^11^[SEM] *vs.* 3.598 × 10¹² particles/mL ± 2.88×10^11^ [SEM])

**Fig. 1.**
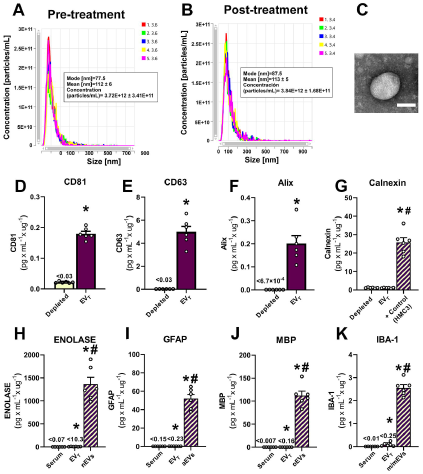
Characterization of serum-derived extracellular vesicles (EVs) and validation of CNS cell-type–specific markers. **(A–B)** Nanoparticle tracking analysis (NTA) showing size distribution and particle concentration of total EVs before and after candesartan treatment in each patient. (**C)** Transmission electron microscopy (TEM) image of a representative EV, confirming typical vesicular morphology (scale bar = 100[nm). **(D– F)** Quantification of canonical EV markers CD81, CD63, and Alix by ELISA, demonstrating EV enrichment compared to EV-depleted serum fractions. (**G)** Calnexin levels were used as a negative control to assess the absence of cellular contamination. An extract from the human microglial cell line HMC3 (ATCC® CRL-3304) was employed as a positive control. (**H–K)** Quantification of neuronal (Enolase), astrocytic (Glial Fibrillary Acidic Protein; GFAP), oligodendrocytic (Myelin Basic Protein; MBP), and microglial (Ionized calcium-binding adapter molecule 1; IBA-1) markers in serum, total extracellular vesicles (EV_T_), and in extracellular vesicles (EVs) isolated from distinct brain cell types confirmed the presence of CNS–derived EVs. Data are presented as mean ± SEM. *p < 0.05 vs. EV-depleted (D-G) or serum (H-K); #p < 0.05 vs. EV_T_.

Transmission electron microscopy (TEM) confirmed the presence of vesicles with typical rounded morphology and diameters within the expected nanometric range (Fig. 1C).

Initially, the concentration of canonical EV markers was compared in 3 pre-candesartan treatment samples and 3 post-candesartan samples. As no concentration differences were detected, the samples were pooled. Protein profiling of total EV (EV_T_) preparations showed robust enrichment of canonical EV markers. The tetraspanins CD81 and CD63 and the cytoplasmic protein involved in exosome biogenesis through the ESCRT pathway Alix were significantly elevated in EV_T_-enriched fractions compared to EV-depleted samples (Fig. 1D and F). Calnexin, an endoplasmic reticulum marker indicative of cellular contamination, was absent in EV_T_ fractions and only detectable in the positive control (the human microglial cell line HMC3 [ATCC® CRL-3304] cell lysate), supporting the purity of the EV isolation (Fig. 1G).

To confirm the CNS origin of EVs, we performed quantitative ELISA analyses targeting cell-type-specific protein markers, each representative of a distinct EV subpopulation. nEVs were identified by the presence of neuron-specific enolase, a glycolytic enzyme abundantly expressed in mature neurons (Fig. 1H). aEVs were characterized by Glial Fibrillary Acidic Protein (GFAP; Fig. 1I), an intermediate filament protein selectively expressed in astrocytes. oEVs were detected using Myelin Basic Protein (MBP; Fig. 1J), a structural component and significant constituent of the myelin sheath. m/mEVs were identified by Ionized Calcium-Binding Adapter Molecule 1 (IBA-1; Fig. 1K), a cytoplasmic protein expressed in microglia and peripheral macrophages. Three pre-treatment samples and three post-treatment samples were compared using a t-test to assess whether the concentrations of NSE, GFAP,IBA-1, and MBP varied following candesartan treatment. No differences were found (Data Sheet in Supplementary Material), and the results were polled to show the enrichment of these markers after immunocapture. All four EV subtypes showed significantly higher levels of their respective markers in the enriched EV fractions compared to whole serum and EV-depleted fractions. These results confirm the successful isolation and characterization of EVs enriched for neuronal, astrocytic, oligodendrocytic, and microglial/macrophage origin.

### Dysregulated proteins of isolated EVs are associated with brain tissue

A quantitative analysis was performed using the SWATH-MS method to compare the protein expression profiles of patients before and after treatment. This study identified 1,141 proteins, 10,416 peptides, and 95,129 spectra, with an FDR <1%. Over these proteins, 874 were quantified. The most dysregulated proteins were identified based on a log fold change (FC) ≥ 0.6 or ≤ -0.7 and p < 0.05, with a more stringent threshold of p < 0.01 applied to highlight the most significantly dysregulated proteins. To validate the characterization data of the obtained extracellular vesicles, the 874 quantified proteins were analyzed using STRING. The results revealed that most of these proteins are associated with brain tissue. Fig. 2A illustrates these findings: red bubbles indicate expression in the brain tissue, blue bubbles represent expression in brain cell lines, green bubbles indicate expression in the brainstem, and yellow bubbles correspond to expression in the brain ventricles. In the protein set enrichment analysis (Fig. 2B), brain tissue and brain cell line exhibit large bubble sizes and low FDR values, suggesting a high degree of statistical significance. Collectively, these findings support the relevance of these analyzed extracellular vesicles in neurological contexts. The protein content of a) nEVs; b) aEVs; c) oEVs; d) m/mEVs was compared before and after candesartan treatment.

**Fig. 2.**
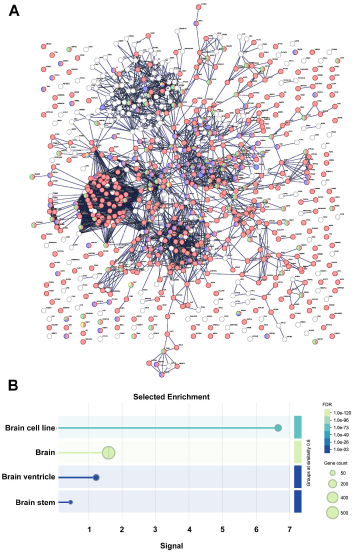
Dysregulated proteins of isolated EVs are associated with brain tissue. **(A)** Protein-protein interaction (PPI) network of the identified proteins generated using STRING. Each node (circle) represents a protein, and edges (lines) indicate known or predicted interactions. Red bubbles indicate tissue expression in the brain, blue bubbles represent expression in brain cell lines, green bubbles denote expression in the brainstem and yellow bubbles correspond to expression in the brain ventricles. Edge colors indicate interaction types: blue for co-occurrence across species, green for functional associations based on text mining, purple for experimentally validated interactions, and yellow for database-derived interactions. The network reveals multiple interconnected protein clusters, suggesting functional modules and highlighting potential key regulatory proteins involved in neurological and cellular processes. **(B)** Gene set enrichment analysis of the identified proteins. The y-axis lists enriched biological pathways, including Brain cell line, Brain, Brainstem, and Brain ventricle, while the x-axis represents statistical significance thresholds. The color gradient indicates the false discovery rate (FDR), with lighter green representing highly significant pathways (FDR < 1.0e-120) and darker blue indicating less significant pathways (FDR ∼ 1.0e-03). The bubble size corresponds to the number of genes associated with each pathway, with larger bubbles indicating pathways with a higher number of identified proteins. Among the most enriched pathways, Brain and Brain cell line exhibit large bubble sizes and low FDR values, suggesting a strong statistical association. These findings indicate a predominant involvement of brain-related pathways in the analyzed dataset.

### Candesartan-induced protein changes in EVs enriched by neuronal origin

In the first analysis (post vs pre-candesartan treatment) in nEVs, 46 dysregulated proteins were identified, of which 19 were upregulated and 27 were downregulated (Supplemental Table S2 in Supplementary Material). The volcano plot analysis of nEVs (Fig. 3A) shows the main upregulated proteins (p<0.05; log_2_FC >1). An in-depth analysis of the dataset shows that RL21, PARK7/DJ-1, FUS, TCPZ, H1X, and CAYP1 were upregulated with p < 0.01, all linked to key neuronal recovery mechanisms. RL21 indicates increased protein synthesis, while PARK7/DJ-1 reflects the activation of antioxidant and mitochondrial protective pathways. CAYP1 may suggest restored intracellular transport. FUS is involved in RNA metabolism, DNA repair, and synaptic maintenance. TCPZ supports proteostasis through the proper folding of cytoskeletal proteins, and H1X may reflect chromatin remodeling in response to treatment. In parallel, several proteins were significantly downregulated (p < 0.01), including HS90B, LANC1, DEST, PLCD1, NRCAM, PUR2, LV39, and AMPL. HS90B, a chaperone involved in protein folding and transcriptional regulation, and LANC1, a glutathione transferase, were downregulated, suggesting diminished proteotoxic and oxidative stress. Lower levels of DEST and PLCD1 suggest the stabilization of cytoskeletal architecture and calcium signaling. NRCAM downregulation may reflect the completion of axonal remodeling, while reduced PUR2 expression implies the normalization of purine metabolism. The decrease in AMPL, a Zn²[-dependent metallopeptidase involved in glutathione S-conjugate degradation and redox regulation, further supports reduced oxidative stress. Finally, lower LV39 levels may indicate decreased immune activation. Altogether, this proteomic profile suggests a coordinated neuroprotective response, involving improved redox and mitochondrial function, enhanced proteostasis and gene regulation, and attenuation of synaptic, metabolic, and inflammatory stress pathways, highlighting the therapeutic potential of the intervention in PD.

**Fig. 3.**
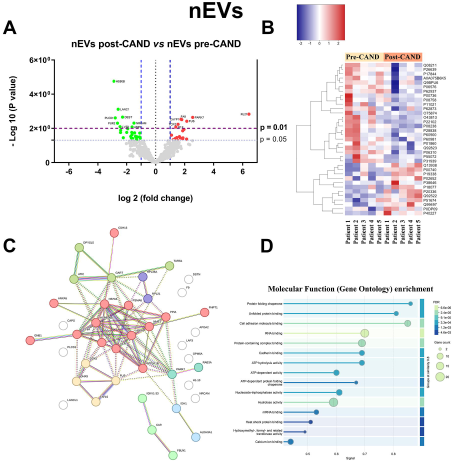
Candesartan-induced protein changes in EVs enriched by neuronal origin. **(A)** Volcano plot showing the differential protein expression analysis of nEVs post-CAND *vs* nEVs pre-CAND. The x-axis represents the log□ fold change, indicating the magnitude of upregulation (right) or downregulation (left), while the y-axis displays the −log□□(p-value), reflecting statistical significance. Upregulated genes (red) and downregulated genes (blue) meet the significance threshold, while non-significant genes (gray) remain unchanged. (**B)** Heatmap representing the expression levels of differentially expressed proteins in nEVs after candesartan treatment. The x-axis corresponds to individual patients, with the first five samples representing the pre-treatment condition and the last five the post-treatment condition. The y-axis lists the proteins that exhibit significant changes in expression. The color scale indicates relative protein abundance, with blue representing downregulation, red representing upregulation, and white denoting intermediate expression levels. **(C)** Protein-protein interaction network generated using the STRING database, illustrating functional associations among key dysregulated proteins. Line colors represent different types of functional associations. **(D)** Gene Ontology (GO) pathway enrichment analysis. The bubble plot illustrates significantly enriched molecular functions. The Y-axis lists the enriched GO categories, while the X-axis represents the signal strength, indicating the enrichment level of each category. Bubble size corresponds to the number of genes associated with each category, and the background color reflects the False Discovery Rate (FDR), where lighter shades indicate higher statistical significance. Larger bubbles denote a greater number of associated genes, and black lines separate different functional groups. This analysis highlights key molecular functions affected in the study and their statistical relevance.

Clustering protein expression patterns are visualized in the heat map (Fig. 3B), illustrating distinct protein expression profiles in nEVs after treatment. Notably, P02652 (APOA2) and Q99497 (PARK7/DJ-1) were upregulated after candesartan treatment, suggesting the activation of neuroprotective pathways. APOA2 may contribute to membrane stabilization, while PARK7DJ-1 supports mitochondrial function and redox homeostasis. Conversely, Q08211 (DHX9), P26639 (SYTC), and P17844 (DDX5) were downregulated after candesartan treatment, possibly reflecting reduced transcriptional stress and improved proteostasis. These proteins are involved in RNA metabolism and translation, and their decreased expression may indicate a normalization of neuronal homeostasis. The consistency of these alterations across individuals supports a reproducible and systemic therapeutic effect on the nEV proteome in Parkinson’s disease.

The protein-protein interaction network generated using STRING (Fig. 3C) highlights key functional associations of all significantly dysregulated proteins (up and downregulated). Central nodes include HSP90AB1, HSPA5, and VCP, which are involved in protein homeostasis, and PARK7DJ-1, which interacts with genes involved in ribosomal proteins (RPL35A, RPL21) and chaperones, linking translational regulation to protein quality control. The presence of PSMA6 (proteasomal subunit) and RNA-binding proteins (FUS, DHX9, DDX5) suggests a connection between protein degradation and post-transcriptional regulation. Notably, PARK7/DJ-1 and RAB3A indicate a role in vesicular trafficking. The strong connectivity of HSPA5 and HSP90 underscores the relevance of proteotoxic stress, while PARK7/DJ-1 emerges as a key regulator and potential biomarker in these processes.

Gene ontology (GO) molecular function enrichment analysis (Fig. 3D) identified four main functional categories associated with dysregulated proteins: (i) Protein Folding and Cellular Stress Response, which includes chaperones and heat shock proteins essential for maintaining protein quality control; (ii) Cell Adhesion and Protein-Protein Interaction, crucial for intercellular communication and structural organization; (iii) Enzymatic Regulation and Energy Metabolism, encompassing ATP-dependent enzymatic activities relevant to biochemical transformations and nucleotide metabolism; and (iv) Nucleic Acid and Ion Binding, regulating gene expression, post-transcriptional processes, and calcium-dependent signaling

The treemap reveals processes related to Candesartan-induced dysregulated proteins in nEVs. The visualization of upregulated biological processes (Figure S2) highlights key biological processes relevant to neuroprotection. Among the upregulated processes, we will mention the following: Increased protein localization to Cajal bodies implies enhanced RNA metabolism and ribonucleoprotein (RNP) assembly, which are crucial for accurate RNA processing and protein synthesis, both essential for maintaining neuronal integrity. Regulation of lyase activity and cytidine triphosphate (CTP) metabolic processes point toward improved neurotransmitter biosynthesis and cellular energy homeostasis, crucial factors for counteracting mitochondrial dysfunction in PD. Enhanced dopamine transport and insulin regulation of neurotransmitter levels indicate improved dopaminergic signaling and glucose homeostasis, which are critical for maintaining synaptic function and preventing metabolic stress. Processes related to detoxification of copper ions and sterol esterification indicate a response to metal-induced oxidative stress and lipid metabolism dysregulation, which have been implicated in dopaminergic neurodegeneration. The regulation of short-term neuronal synaptic plasticity reflects a potential improvement in synaptic efficiency and adaptability, which could contribute to better motor and cognitive function in PD patients. The positive regulation of cellular respiration indicates an enhancement of mitochondrial function, potentially leading to a better ATP supply and reduced oxidative damage. The regulation of the autophagy of mitochondria highlights an increase in mitochondrial quality control, which is crucial for removing dysfunctional mitochondria and maintaining cellular energy efficiency. The regulation of dopamine metabolic processes suggests an adaptive response to optimize dopamine turnover and availability, central to PD symptom relief. These findings suggest that candesartan treatment for PD patients may activate endogenous defense mechanisms that optimize cellular metabolism, enhance synaptic function, suppress neuroinflammation, and improve proteostasis, potentially delaying disease progression and improving patient outcomes.

The treemap visualization of downregulated GO biological processes (Figure S3) after candesartan treatment suggests a shift toward reduced cellular stress, oxidative damage, and neuroinflammation, contributing to improved neuronal homeostasis and survival. The decrease in Inosine Monophosphate (IMP) biosynthesis and nucleobase catabolism may reflect a reduction in the metabolic burden and oxidative stress, alleviating mitochondrial dysfunction. Similarly, the downregulation of unfolded protein response (UPR) and protein folding pathways indicates a lower demand for proteostasis mechanisms, suggesting that treatment has mitigated the accumulation of misfolded proteins. The reduction in NF-kappaB transcription factor activity and interferon-gamma response regulation points toward decreased neuroinflammatory signaling, which is crucial for limiting chronic glial activation and neuronal damage. A decrease in nucleocytoplasmic transport may prevent the mislocalization of toxic proteins such as α-synuclein, while the downregulation of ribose phosphate biosynthetic processes may indicate a reduced reliance on compensatory antioxidant mechanisms, reflecting an overall decline in oxidative stress. Notably, the observed suppression of aerobic respiration pathways suggests improved mitochondrial efficiency, which may limit excessive reactive oxygen species (ROS) production. The decline in cerebellar development-related pathways may represent a restoration of motor circuit function, minimizing compensatory mechanisms that arise due to basal ganglia dysfunction. Altogether, these findings suggest that candesartan treatment in PD reduces pathological stress responses, enhances mitochondrial efficiency, limits neuroinflammation, and improves proteostasis, ultimately fostering a more stable neuronal environment that may slow disease progression and improve patient outcomes.

### Candesartan-induced protein changes in EVs enriched by astrocytic origin

Analysis of astrocyte-derived extracellular vesicles (aEV) proteomic cargo showed 22 dysregulated proteins, of which 10 were upregulated and 12 were downregulated after candesartan treatment (Supplemental Table S3 in Supplementary Material). The volcano plot analysis of aEVs (Fig. 4A) identified the principal dysregulated proteins (p < 0.05; log_2_FC >1). Among those upregulated, the proteins with the highest statistical significance (p < 0.01) included PSB4, LSAMP, PHLD, and SYFB, all associated with cellular maintenance and neuroprotective functions. PSB4, a component of the 20S proteasome complex, may indicate enhanced proteasomal activity and improved proteostasis, which are essential for the clearance of misfolded or damaged proteins in neurodegenerative conditions. LSAMP, involved in cell adhesion and synaptic organization, may contribute to stabilizing astrocyte–neuron interactions and synaptic architecture. PHLD upregulation may indicate enhanced astrocytic clearance of toxic GPI-anchored proteins, suggesting a neuroprotective response induced by the treatment. SYFB, which is involved in mitochondrial protein synthesis, may reflect improved mitochondrial efficiency and metabolic support within astrocytes, both critical for sustaining neuronal health in PD.

**Fig. 4.**
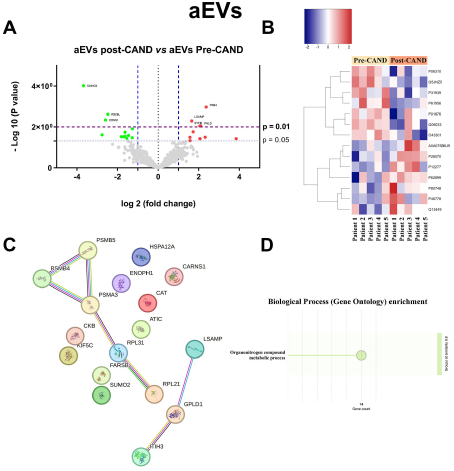
Candesartan-induced protein changes in EVs enriched by astrocytic origin. **(A)** Volcano plot showing the differential protein expression analysis of aEVs post-CAND *vs* aEVs pre-CAND. The x-axis represents the log□ fold change, indicating the magnitude of upregulation (right) or downregulation (left), while the y-axis displays the −log□□(p-value), reflecting statistical significance. Upregulated genes (red) and downregulated genes (blue) meet the significance threshold, while non-significant genes (gray) remain unchanged. **(B)** Heatmap representing the expression levels of differentially expressed proteins in aEVs after candesartan treatment. The x-axis corresponds to individual patients, with the first five samples representing the pre-treatment condition and the last five the post-treatment condition. The y-axis lists the proteins that exhibit significant changes in expression. The color scale indicates relative protein abundance, with blue representing downregulation, red representing upregulation, and white denoting intermediate expression levels. **(C)** Protein-protein interaction network generated using the STRING database, illustrating functional associations among key dysregulated proteins. Line colors represent different types of functional associations. **(D)** Gene Ontology (GO) pathway enrichment analysis. The bubble plot illustrates significantly enriched molecular functions. The Y-axis lists the enriched GO categories, while the X-axis represents the signal strength, indicating the enrichment level of each category. Bubble size corresponds to the number of genes associated with each category, and the background color reflects the False Discovery Rate (FDR), where lighter shades indicate higher statistical significance. Larger bubbles denote a greater number of associated genes, and black lines separate different functional groups. This analysis highlights key molecular functions affected in the study an their statistical relevance.

Conversely, the treatment induced the downregulation (p<0.01) of stress-related proteins, including SUMO2, RS26L, and CRNS1. SUMO2, a central component of the SUMOylation pathway, is often elevated in response to oxidative and proteotoxic stress; its decreased expression may reflect reduced cellular stress and restored homeostasis. RS26L, typically upregulated under stress conditions to regulate translation, may be downregulated due to normalized protein synthesis activity. CRNS1 (Carnosinase 1), involved in dipeptide metabolism and redox regulation, showed reduced levels, potentially indicating a shift toward a more balanced oxidative state and metabolic profile in astrocytes. Together, these proteomic alterations suggest a dual mechanism of action, in which treatment promotes cellular maintenance and metabolic support while concurrently attenuating astrocytic stress responses, contributing to a neuroprotective environment in the context of PD.

The heatmap (Fig. 4B) shows distinct protein expression changes in aEVs pre- and post-treatment, with clustering indicating coordinated regulation. Notably, P31939 (PUR9) and O43301 (HS12A) were significantly downregulated after candesartan treatment, suggesting reduced astrocytic stress and metabolic burden, consistent with a protective effect on the neural environment. In contrast, P28070 (PSB4) and P46778 (RPL21) were upregulated after candesartan treatment, indicating enhanced proteostasis and protein synthesis. These changes suggest a transition to a less reactive, more supportive astrocytic phenotype, potentially contributing to neuroprotection in PD. Their consistency across samples supports a reproducible treatment effect.

The PPI network generated with all significantly dysregulated proteins using STRING (Fig. 4C) revealed key functional associations among differentially expressed proteins, highlighting processes such as protein homeostasis, energy metabolism, and neuroinflammatory regulation. Central nodes include PSMB4, PSMB5, and PSMA3, indicating an enhanced ubiquitin-proteasome system (UPS) that may reduce proteotoxic stress and α-synuclein accumulation. Upregulated RPL21 and RPL31 may suggest enhanced ribosomal activity and protein synthesis associated with cellular recovery processes, while CKB and FARSB may indicate enhanced astrocytic metabolic support and protein synthesis capacity. A decreased expression of HSPA12A, SUMO2, and ATIC may reflect reduced cellular stress and metabolic burden in astrocytes. HSPA12A is a stress-inducible chaperone involved in proteostasis. SUMO2 participates in post-translational stress responses, and ATIC is a key enzyme in purine biosynthesis, often upregulated during inflammation and cellular stress. Furthermore, the decrease in immunomodulatory proteins such as ITIH3 indicates a shift toward a less inflammatory state. Altogether, these changes in aEV cargo indicate that candesartan elicits a cellular response characterized by enhanced mechanisms of cellular maintenance and metabolic support, alongside a reduction in astrocytic stress-related pathways. These changes suggest that candesartan promotes a neuroprotective phenotype in astrocytes within the context of Parkinson’s disease.

The GO biological process analysis (Fig. 4D) identifies “organonitrogen compound metabolic process” which suggests active involvement of astrocytes in the metabolism of nitrogen-containing neurotransmitters, amino acids, and related compounds, reflecting their role in neurotransmitter clearance, glutamine synthesis, and ammonia detoxification potentially contributing to improved proteostasis, metabolic efficiency, and neuroprotection in the context of PD.

The treemap (Figure S4) containing the most significant enriched gene ontology terms (biological processes) suggests a multifaceted neuroprotective response after candesartan treatment. An increased expression of genes associated with processes such as mitochondrial ATP synthesis, regulation of protein translation, RNA splicing, purine and vitamin metabolism, and cellular respiration suggests enhanced metabolic support, proteostasis, and intercellular communication. Notably, upregulation of genes involved in interleukin-7 response, substantia nigra development, intermediate filament organization, and midbrain development may indicate activation of neurodevelopmental and structural support pathways, contributing to astrocyte-mediated resilience and neurorepair. These molecular signatures reflect a shift toward a reparative astrocytic phenotype, improved bioenergetic balance, and reduced neuroinflammation, collectively supporting a neuroprotective response induced by the treatment in PD patients.

Figure S5 shows the tree map containing the downregulated Gene ontology terms (biological processes) after candesartan treatment. It highlights several functional pathways whose suppression suggests a neuroprotective response. Among the most relevant downregulated processes are the acute inflammatory response, lipid catabolic processes, complement activation, regulation of endopeptidase activity, cytolysis, protein oxidation, cell killing, and the positive regulation of cytokine production. The downregulation of immune-related processes, including inflammation and cytokine signaling, suggests a reduction in neuroinflammatory activity, a key driver of neurodegeneration in PD. Likewise, the attenuation of complement activation may reflect decreased neurovascular damage and immune-mediated cytotoxicity. Reduced activity in cytolytic, oxidative, and cell death pathways points toward diminished cellular stress and enhanced neuronal survival. Additionally, downregulation of lipid degradation and remodeling processes may indicate improved membrane stability and reduced susceptibility to lipid peroxidation. Collectively, these downregulated processes point to a therapeutic shift toward metabolic efficiency, synaptic stability, reduced neuroinflammation, and oxidative stress control.

**Fig. 5.**
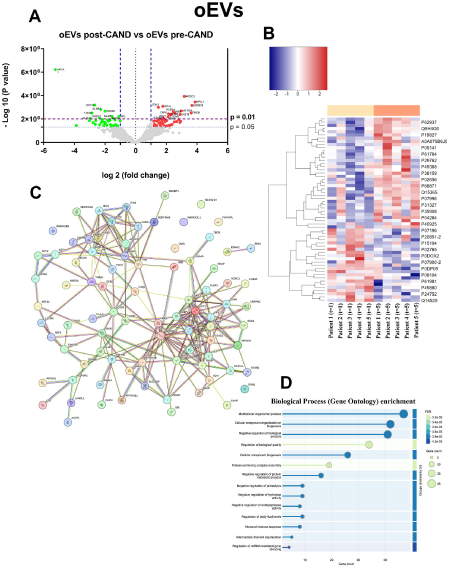
Candesartan-induced protein changes in EVs enriched by oligodendroglia origin**. (A)** Volcano plot showing the differential protein expression analysis of oEVs post-CAND *vs* oEVs pre-CAND. The x-axis represents the log□ fold change, indicating the magnitude of upregulation (right) or downregulation (left), while the y-axis displays the −log□□(p-value), reflecting statistical significance. Upregulated genes (red) and downregulated genes (blue) meet the significance threshold, while non-significant genes (gray) remain unchanged. **(B)** Heatmap representing the expression levels of differentially expressed proteins in oEVs after candesartan treatment. The x-axis corresponds to individual patients, with the first five samples representing the pre-treatment condition and the last five the post-treatment condition. The y-axis lists the proteins that exhibit significant changes in expression. The color scale indicates relative protein abundance, with blue representing downregulation, red representing upregulation, and white denoting intermediate expression levels. (**C)** Protein-protein interaction network generated using the STRING database, illustrating functional associations among key dysregulated proteins. Line colors represent different types of functional associations. (**D)** Gene Ontology (GO) pathway enrichment analysis. The bubble plot illustrates significantly enriched molecular functions. The Y-axis lists the enriched GO categories, while the X-axis represents the signal strength, indicating the enrichment level of each category. Bubble size corresponds to the number of genes associated with each category, and the background color reflects the False Discovery Rate (FDR), where lighter shades indicate higher statistical significance. Larger bubbles denote a greater number of associated genes, and black lines separate different functional groups.

### Candesartan-induced protein changes in EVs enriched by oligodendroglia origin

Oligodendrocyte-derived extracellular vesicles (oEV) proteomic cargo showed 92 dysregulated proteins, of which 55 were upregulated and 37 were downregulated after candesartan treatment (Supplemental Table S4 in Supplementary Material). The volcano plot analysis of oEVs revealed significant alterations in their proteomic cargo following candesartan treatment (Fig. 5A), suggesting a neuroprotective effect in PD aimed at enhancing myelin repair, metabolic homeostasis, oxidative stress resistance, immune modulation, and neuronal support. An increase (p<0.01; log_2_FC > 1) in the expression of APOC3, NP1L1, and SORCN supports lipid metabolism, neurogenesis, and calcium homeostasis. APOC3 may aid membrane repair via triglyceride modulation. NP1L1 could enhance DNA repair and neurogenesis, supporting oligodendrocyte regeneration. SORCN may improve cellular resilience through calcium regulation. PPIA, DHX9, and PCBP1 are involved in RNA processing, protein quality control, and cellular stress responses, supporting proteostasis and reducing toxic aggregate formation. ATP5H and CPSM contribute to mitochondrial function and ammonia detoxification, enhancing metabolic efficiency. TBCB and DBNL regulate cytoskeletal dynamics and axonal growth, potentially facilitating neuronal repair and connectivity. STXB1 and L1CAM are critical for synaptic vesicle fusion and cell adhesion, respectively, indicating improved synaptic function and neuronal communication. Collectively, the enrichment of these proteins in oEVs suggests that oligodendrocytes contribute to a neuroprotective environment through enhanced support of neuronal homeostasis, synaptic integrity, and cellular resilience.

Conversely, the downregulation (p<0.01; log_2_FC < -0.7) of AK1A1, a key enzyme for detoxifying reactive aldehydes such as methylglyoxal and acrolein, suggests a lower oxidative burden in treated cells. A decline in CALR, a chaperone involved in ER stress and calcium homeostasis, points to improved proteostasis, while reduced CLIC4 levels imply attenuation of redox-sensitive signaling and inflammatory processes. The downregulation of KLKB1, a protease activating the kallikrein-kinin system, indicates dampened oligodendrocyte-mediated inflammation. The lower abundance of CAD13 may reflect an environment favoring axonal regeneration or remyelination. Reduction of ENOG, a neurotrophic factor, may indicate diminished compensatory stress responses. Overall, these changes suggest that the treatment promotes a more homeostatic and neuroprotective oligodendrocyte phenotype in PD patients.

The heatmap analysis of differentially expressed proteins in oEVs after candesartan treatment (Fig. 5B) reveals distinct proteomic shifts indicative of a neuroprotective response. Elevated expression of Q9H4G0 (E41L1) may reflect an effective modulation in neuron-oligodendroglial interaction, while upregulation of P61764 (STXB1) points to potential restoration of synaptic vesicle trafficking and neurotransmission. The presence of P40925 (MDHC) indicates an improvement in metabolic function and redox homeostasis. Conversely, proteins such as P24752 (THIL) and P61981 (1433G) display downregulation after candesartan treatment, which suggests a reduction in mitochondrial β-oxidation activity and improved mitochondrial protein quality control, respectively. The observed proteomic shifts suggest that the cargo of oEVs reflects the neuroprotective effects of candesartan, primarily through the modulation of neuron-glia communication, the promotion of metabolic homeostasis, and the attenuation of oxidative stress.

The protein-protein interaction (PPI) network analysis generated using STRING (Fig. 5C) reveals a highly coordinated molecular adaptation, suggesting a shift toward a neuroprotective status. The presence of functionally distinct clusters indicates specific cellular adaptations, including enhanced mitochondrial function and metabolic efficiency (ATP5PD, ATP5F1B, SLC25A5, VDAC2, ALDH9A1), optimized RNA processing and translation regulation (EIF4G1, RBMX, ELAVL1, PCBP1, FUBP1, RPS12, RPL27A), and reinforcement of synaptic stability and axonal support (MAP1A, SNAP25, STXBP1, TUBA4A, L1CAM, NEFL). The upregulation of oxidative stress regulators (CAT, PPIA, CTSD, LANCL1) and the downregulation of HSP90AA1 suggest an adaptive response to neurotoxic stress, while the modulation of immune-related proteins (C1QC, SERPINA5, CAMP, ITIH1, ITIH3, THBS1) indicates a controlled neuroinflammatory response, reducing potential oligodendrocyte-mediated toxicity. The presence of highly connected central hubs, such as HSP90AA1, ATP5F1B, and EIF4G1, further underscores the critical role of cellular energy balance, protein homeostasis, and synaptic modulation in oligodendrocyte-mediated neuroprotection.

GO biological process analysis (Fig. 5D) identified several major functional categories enriched among the dysregulated proteins in oEVs, including pathways related to structural organization, protein complex assembly, and regulation of metabolic and degradative processes. These biological functions are particularly relevant in PD, as they reflect the glial mechanisms involved in maintaining tissue homeostasis and neural support.

The treemap showed the candesartan-induced upregulation of key biological processes in oligodendrocytes suggesting neuroprotection (Figure S6). The increased purine nucleoside triphosphate biosynthetic process, regulation of RNA export from the nucleus, and synapse organization indicate enhanced molecular processing, transcriptional regulation, and synaptic maintenance, all crucial for oligodendrocyte function and neuron-glia communication. Negative regulation of proteolysis and cellular catabolic processes suggests an effort to preserve essential myelin and structural proteins, preventing excessive degradation linked to PD pathology. The upregulation of apoptotic cell clearance and antigen processing and presentation suggests a balanced neuroimmune response, enhancing cell survival while reducing neuroinflammation. Furthermore, the activation of the hydrogen peroxide catabolic process, aerobic respiration, and ATP metabolic pathways suggests an enhancement in energy metabolism. Collectively, these upregulated pathways suggest an adaptive and protective role of oligodendrocytes, enhancing myelination, synaptic function, metabolic efficiency, and neuroimmune balance, ultimately supporting neuronal survival and functional recovery post-treatment.

Following candesartan treatment, the downregulation of key biological processes in oligodendrocytes (Figure S7) also suggests a neuroprotective effect. The inhibition of alpha-amino acid catabolic processes, carboxylic acid metabolic processes, and organic acid catabolic processes indicates a decrease in metabolic overload, preventing excessive oxidative stress and cellular damage. Similarly, the negative regulation of phagocytosis and complement activation via the lectin pathway suggests a reduction in excessive immune activation, preventing chronic inflammation that could contribute to oligodendrocyte dysfunction in PD. Overall, these downregulated processes suggest a neuroprotective adaptation in oligodendrocytes, reducing metabolic and oxidative stress, stabilizing gene expression, limiting excessive immune responses, and preventing unnecessary cell proliferation. This shift likely contributes to a more supportive and stable neural environment, ultimately enhancing neuronal survival and functional recovery in PD following treatment.

#### Candesartan-induced protein changes in EVs enriched by microglia/macrophage origin

Analysis of microglia/macrophage-derived extracellular vesicles (m/mEV) proteomic cargo showed 48 dysregulated proteins, of which 23 were upregulated and 25 were downregulated (Supplemental Table S5 in Supplementary Material). The volcano plot analysis of m/mEVs reveals significant alterations in their proteomic cargo (Fig. 6A), suggesting a neuroprotective response mediated by enhanced immune regulation, metabolic adaptation, oxidative stress resistance, and synaptic support. The increased expression (p<0.01; log_2_FC > 1) of NSF1C, APOA2, E41L1, PP1R7, RAB14, SNAP25, and ITIH1 after candesartan treatment suggests a shift toward a reparative immune phenotype. NSF1C supports proteostasis via VCP modulation; APOA2 and ITIH1 are linked to anti-inflammatory lipid and matrix regulation; E41L1 and SNAP25 may reflect enhanced neuroimmune communication and synaptic support; PP1R7 and RAB14 indicate improved signaling and vesicle trafficking. Together, these changes suggest reduced neuroinflammation and enhanced microglial support for neuronal homeostasis. Conversely, A decreased expression (p<0.01; log_2_FC < -0.7) of ODO1, PCP4, PDIA6, RS10, and HSP72 may indicate reduced microglial stress, inflammatory signaling, and maladaptive activation. ODO1 downregulation could reflect lower mitochondrial metabolic overactivation, limiting ROS production. Reduced PCP4 may indicate dampened calcium/calmodulin signaling, associated with microglial reactivity. Lower PDIA6 and HSP72 levels suggest decreased endoplasmic reticulum stress and protein misfolding burden, respectively. Reduced RS10 could reflect a shift away from excessive protein synthesis, often seen in pro-inflammatory states. Together, these changes are consistent with a transition toward a less inflammatory, homeostatic microglial phenotype, potentially supporting neuronal survival.

**Fig. 6.**
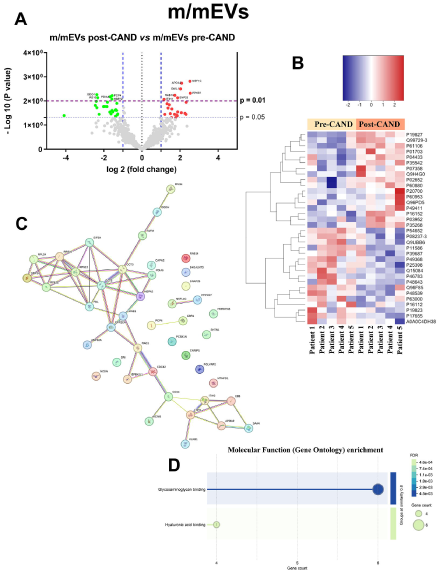
Candesartan-induced protein changes in EVs enriched by microglia/macrophage origin. **(A)** Volcano plot showing the differential protein expression analysis of m/mEVs post-CAND *vs* m/mEVs pre-CAND. The x-axis represents the log□ fold change, indicating the magnitude of upregulation (right) or downregulation (left), while the y-axis displays the −log□□(p-value), reflecting statistical significance. Upregulated genes (red) and downregulated genes (blue) meet the significance threshold, while non-significant genes (gray) remain unchanged. **(B)** Heatmap representing the expression levels of differentially expressed proteins in m/mEVs after candesartan treatment. The x-axis corresponds to individual patients, with the first five samples representing the pre-treatment condition and the last five the post-treatment condition. The y-axis lists the proteins that exhibit significant changes in expression. The color scale indicates relative protein abundance, with blue representing downregulation, red representing upregulation, and white denoting intermediate expression levels. **(C)** Protein-protein interaction network generated using the STRING database, illustrating functional associations among key dysregulated proteins. Line colors represent different types of functional associations. **(D)** Gene Ontology (GO) pathway enrichment analysis. The bubble plot illustrates significantly enriched molecular functions. The Y-axis lists the enriched GO categories, while the X-axis represents the signal strength, indicating the enrichment level of each category. Bubble size corresponds to the number of genes associated with each category, and the background color reflects the False Discovery Rate (FDR), where lighter shades indicate higher statistical significance. Larger bubbles denote a greater number of associated genes, and black lines separate different functional groups.

The heat map of differentially expressed proteins in m/mEVs (Fig. 6B) reveals a distinct proteomic shift indicative of a therapeutic effect, suggesting a coordinated molecular response to treatment. After candesartan treatment, the increased concentration of proteins involved in synaptic maintenance P60880 (SNP25), vesicle trafficking P61106 (RAB14), RNA regulation Q99729 (HNRNPAB), immune modulation P04433, P01703, P07358 (KV311, LV140 CO8B), extracellular matrix stability P19827 (ITIH1), and lipid metabolism P35542 (SAA4) may indicate enhanced microglial support of neuronal function and homeostasis, consistent with treatment-induced neuroprotection. Conversely, proteins associated with ER stress Q15084 (PDIA6), altered protein synthesis P46783 (RS10), cytoskeletal destabilization P48643 (TCPE), and neurotoxic proteolysis P17655 (CAN2) display downregulation after candesartan treatment, suggesting a shift toward a homeostatic, neuroprotective microglial state.

The PPI network analysis of differentially expressed proteins in microglia post-treatment (Fig. 6C) reveals a highly coordinated molecular adaptation, suggesting a shift toward a neuroprotective and homeostatic phenotype. The central cluster, comprising ribosomal and translational regulators (RPS10, RPS12, RPL22, RPL24, EIF3A, FBL, SRP14), suggests an upregulation of protein synthesis machinery, potentially supporting the production of proteins essential for neuronal maintenance and immune regulation. Strong connectivity between cytoskeletal regulators (CDC42, RAC1, EPB41L1, CD44, LMNB1) and translational machinery indicates a coordinated response that enhances microglial structural plasticity while preventing excessive activation. Additionally, the upregulation of ITIH1, ITIH2, C8B, APOA2, and SAA4 suggests a modulation of immune signaling and complement regulation, contributing to controlled inflammatory responses and reduced neurotoxicity. Overall, these findings indicate that the candesartan treatment induces a coordinated microglial response, optimizing translational activity, cytoskeletal remodeling, extracellular matrix stability, and metabolic efficiency, ultimately fostering a neuroprotective microenvironment in PD.

The GO functional enrichment analysis of differentially expressed genes in microglia (Fig. 6D) reveals a significant modulation of biological processes associated with glycosaminoglycan and hyaluronic acid binding, both crucial for extracellular matrix remodeling, immune signaling, and neuroprotection.

The treemap (Figure S8) showed upregulations of protein-containing complex remodeling, Golgi localization, and phospholipid efflux, suggesting intracellular trafficking and lipid transport increase, which are essential for membrane maintenance, vesicle trafficking, and cholesterol homeostasis in microglia. The upregulation of steroid esterification, regulation of cholesterol esterification, and high-density lipoprotein particle clearance highlight a critical role in lipid metabolism and transport, potentially reducing lipid accumulation and oxidative stress. Additionally, the positive regulation of pseudopodium assembly, cell migration, and vesicle organization suggests an increase in microglial motility and endocytic processing, which may enhance the clearance of neurotoxic protein aggregates and apoptotic debris. Altogether, these upregulated processes suggest that microglia/macrophages are transitioning toward a homeostatic and neuroprotective phenotype, enhancing cellular clearance, lipid homeostasis, synaptic stability, and neuronal support.

The downregulation (Figure S9) of protein localization to Cajal bodies and regulation of RNA localization suggest a shift toward transcriptional stability, preventing unnecessary metabolic burden on microglia. The decrease in regulation of phospholipid biosynthetic processes and dicarboxylic acid metabolic processes may indicate an effort to reduce lipid metabolic stress, preventing lipotoxicity that can impair microglial function. Similarly, the inhibition of calcium ion transmembrane transport and the regulation of electrical coupling suggest the stabilization of intracellular calcium levels, preventing calcium dysregulation, a key contributor to oxidative stress and neuroinflammation in PD. The downregulation of endopeptidase activity and negative regulation of inclusion body assembly suggest a reduction in proteolytic activity and protein aggregation management, which may reflect a decreased load of misfolded proteins, reducing the need for excessive degradation pathways. The downregulation of engulfment of apoptotic cells suggests a controlled microglial phagocytic response, limiting excessive synaptic pruning and preserving neuronal connections. The negative regulation of intrinsic apoptotic signaling in response to DNA damage suggests a reduced need for microglial apoptosis regulation, potentially indicating lower cellular stress and improved survival rates of beneficial microglial subtypes. These downregulated processes suggest a transition of microglia/macrophages toward a homeostatic, neuroprotective state, reducing excessive immune activation, metabolic stress, and extracellular remodeling, ultimately contributing to a stabilized neural environment and enhanced neuronal survival.

## Discussion

The present methodology allows for the first time the demonstration of functional effects exerted by the oral administration of angiotensin AT1 receptor antagonists (ARBs) in PD patients. Treatment with candesartan induced the upregulation or downregulation of a large number of proteins relevant to PD progression. These changes were detected not only in EVs derived from neurons (46 dysregulated proteins) and microglia/macrophages (48 dysregulated proteins), but also in astrocytes (22 dysregulated proteins) and oligodendrocytes (92 dysregulated proteins).

PD has traditionally been considered a gray matter/neuronal disease, with glial alterations interpreted as consequences or collateral damage of the neurodegenerative process. However, recent studies have identified novel glial mechanisms that could play significant roles in neurodegeneration. These mechanisms include a glial role beyond the microglial neuroinflammatory response and astrocyte regulation of the microglial response and neuroinflammation. Astrocytes are essential for providing metabolic support to neurons and preventing neurodegeneration [32]. The role of oligodendrocytes extends beyond their involvement in myelin formation and demyelinating diseases. Recent multi-omic studies have shown that oligodendrocyte-related genes are significantly altered across all stages of PD in the midbrain, even in early stages, suggesting a role for oligodendrocytes in PD pathogenesis rather than as passive participants in neurodegeneration and neuroinflammation, for instance, by providing intercellular metabolic support to axons through the transfer of energy metabolites [33].

The isolation and analysis of EVs from neurons and different types of glial cells within the same blood sample provides an unprecedented tool for obtaining information on the effects occurring in the brain during pathological processes, and in particular, for monitoring the effects of potential neuroprotective treatments. In EVs derived from neurons, candesartan induced a series of changes affecting major mechanisms involved in PD progression. The data suggest neuroprotective effects associated with processes such as the reduction of oxidative stress, ER stress, and mitochondrial dysfunction, reduction of proteotoxic stress burden, enhanced cellular resilience, and synaptic stability. There was also significant enrichment in processes related to dopamine metabolism and transport. Additionally, the downregulation of inflammatory and immune response pathways, such as the regulation of NF-κB transcription activity and response to interferon-gamma, suggests a decrease in the stimulation of inflammatory responses, a major contributor to neuronal degeneration in PD. Changes in aEVs contribute to a more supportive astroglial environment, potentially slowing PD progression and improving neuronal function. Astroglial changes suggest a shift toward reduced neuroinflammation, oxidative stress control, metabolic efficiency, and reduced neuroinflammation, which may contribute to reduced neurodegeneration and improved neuronal resilience in PD patients. Changes in oEVs suggest an adaptive and protective role of oligodendrocytes, enhancing myelination, synaptic function, metabolic efficiency, and neuroimmune balance to prevent excessive oligodendrocyte-mediated inflammation, ultimately supporting neuronal survival and functional recovery post-treatment. Candesartan-induced protein changes in EVs enriched by microglia/macrophage origin (m/mEVs) promote a shift toward a neuroprotective phenotype and a microglial-mediated neuroprotective environment, including cytoskeletal remodeling, metabolic stabilization, diminished oxidative stress, and controlled immune activation, ultimately contributing to a stabilized neural environment and enhanced neuronal survival.

The results observed in EVs from PD patients in the present study are consistent with numerous previous studies in PD animal and cell culture models showing the neuroprotective effects of ARBs by counteracting several major mechanisms of dopaminergic neuron degeneration [6]. In rodent models and primary mesencephalic cultures, ARBs such as candesartan and telmisartan reduced dopaminergic neuron death induced by neurotoxins such as 6-OHDA [12, 13], MPTP [34, 35] or overexpression of alpha-synuclein [9, 11] through inhibition of NADPH-oxidase derived oxidative stress and the microglial neuroinflammatory response. AT1 blockers modulated mitochondrial respiration and mitochondrial-derived oxidative stress [7, 36, 37] and regulated mitochondrial dynamics by inhibiting mitochondrial fission [38]. Similar effects on mitochondrial dynamics were observed by upregulation of the compensatory RAS through administration of Angiotensin 1-7 [38]. In a recent study, we also observed that AT1 promotes oxidative stress and intracellular calcium increase, leading to alpha-synuclein aggregation, which is inhibited by AT1 blockers [11, 39]. Angiotensin/AT1 signaling also modulated cholesterol homeostasis in dopaminergic neurons and astrocytes [40]. Furthermore, mutual regulation between brain angiotensin and dopamine systems has been shown in experimental models in our laboratory [41–43] and several others [6, 44].

## Conclusion

Despite limitations such as the short duration of candesartan treatment (6 months) and a limited number of patients, the present proteomic analysis demonstrates remarkable neuroprotective effects of ARBs crossing the BBB, such as candesartan, on PD patient neurons and glial cells. The results suggest that larger clinical trials should be conducted using ARBs or other RAS modifier drugs as an early therapeutic strategy for neuroprotection against the progression of PD.

## Supporting information

The Supplementary Material contains Supplementary Tables (S1 to S5), Supplementary Methods, Data sheet relative to Figure 1 and Supplementary Figures

## Abbreviations

Ang1-7: Angiotensin 1-7
AngII: Angiotensin II
AT1: Angiotensin type 1
aEVs: Astrocyte-derived EVs
ARBs: AT1 receptor blockers
*AGTR1*: AT1 receptor gene
AFC: Automatic Fraction Collector
BBB: Brain-blood-barrier
CRNS1: Carnosinase 1
CNS: Central nervous system
CI: Cognitive impairment
CTP: Cytidine triphosphate
DDA: Data-dependent acquisition
DIA: Data-independent acquisition
EVs: Extracellular vesicles
FC: Fold change
4PL: Four-parameter logistic
GO: Gene Ontology
GFAP: Glial Fibrillary Acidic Protein
Glast: Glutamate Aspartate Transporter.
H&Y: Hoehn and Yahr
IMP: Inosine Monophosphate
IBA-1: Ionized Calcium-Binding Adapter Molecule 1
L1CAM: L1 Cell Adhesion Molecule
MasR: Mas receptors
m/mEVs: Microglia/macrophage-derived EVs
MoCA: Montreal Cognitive Assessment
MLR: Most likely ratio
MBP: Myelin Basic Protein
MOG: Myelin Oligodendrocyte Glycoprotein
NTA: Nanoparticle Tracking analysis
nEVs: Neuronal EVs
NSE: Neuron-specific enolase
oEVs: Oligodendrocyte-derived EVs
PD: Parkinson’s disease
PCA: Principal component analysis
PPI: Protein-protein interaction
ROS: Reactive oxygen species
RAS: Renin-Angiotensin System
RNP: Ribonucleoprotein
SEM: Standard Error of the Mean
EVT: Total EV
TMEM119: Transmembrane protein 119
TEM: Transmission electron microscopy
UPS: Ubiquitin-proteasome system
UPR: Unfolded protein response

## Supplementary Information

The Supplementary Material contains: Supplementary Tables (S1 to S5), Supplementary Methods, Data sheet relative to Figure 1 and Supplementary Figures (S1 to S9)

## Acknowledgements

The patients participating in the study are gratefully acknowledged. The authors would like to thank Pilar Aldrey, Iria Novoa, and Cristina Gianzo for their valuable technical assistance. We also thank EGO Genomics (https://egogenomics.com/) for their support and collaboration throughout the study.

## Author contributions

JL-LG and AI-RP conceptualized and designed the study, obtained funding, performed data analysis, and drafted the manuscript. L-CM, M-VT, and A-MC conducted the isolation and characterization of extracellular vesicles and contributed to the optimization of experimental protocols and data interpretation related to vesicle biology. SB-B carried out the proteomic analyses and supported the integration of proteomic data with clinical and experimental findings. H-BK and J-K designed the clinical study, recruited patients, and collected all clinical data. CM-L, JL-L, M-A, and JL-C contributed to study design and data analysis, and participated in the critical revision of the manuscript. All authors contributed to the critical review and final approval of the manuscript.

## Data availability

The mass spectrometry proteomics data have been deposited to the ProteomeXchange Consortium via the PRIDE [1] partner repository with the dataset identifier PXD063198. The data supporting the conclusions of this article is included within the article and its Supplementary Information.

## Declarations

### Ethics approval and consent to participate

This study was conducted following the EU Regulation 2016/679, and the Spanish Organic Law 3/2018 on the protection of personal data. Ethical approval was obtained from by the Ethics Committee of Sant Pau University Hospital, Barcelona, and was registered in the European Clinical Trials Register under EudraCT number 2016-000679-25 (https://www.clinicaltrialsregister.eu/ctr-search/trial/2016-000679-25/ES) All participants were informed about the purpose and procedures of the study, and written informed consent was obtained before their inclusion.

### Consent for publication

All authors have approved the contents of this manuscript and provided consent for publication

