## Supplementary material for "Proteomic analysis of blood neuronal and glial extracellular vesicles reveals neuroprotective effects of the angiotensin type-1 blocker candesartan in Parkinson’s disease patients": The Supplementary Material contains Supplementary Tables (S1 to S5), Supplementary Methods, Data sheet relative to Figure 1 and Supplementary Figures: Supplementary Material_05_2025.pdf

**Supplemental Table 1.** Main characteristics of the Parkinsonian patients in the study.

| **Patient Code** | **Sex** | **Age Range** (years) | **Nationality** | **Education** | **UPDRS III (Pre)** | **H&Y (Pre)** | **UPDRS III (Post)** | **H&Y (Post)** |
|---|---|---|---|---|---|---|---|---|
| 01 | F | 66-70 | Spanish | 6 years | 32 | 2 | 31 | 2 |
| 02 | M | 60-65 | Spanish | 18 years | 36 | 2 | 31 | 2 |
| 03 | M | 60-65 | Spanish | 8 years | 28 | 2 | 27 | 2 |
| 04 | F | 66-70 | Spanish | 7 years | 22 | 2 | 27 | 2 |
| 05 | M | 66-70 | Spanish | 18 years | 20 | 2 | 26 | 3 |

DOB = Date of Birth; H&Y = Hoehn and Yahr stage; UPDRS III: motor examination section of the Unified Parkinson's Disease Rating Scale. Pre/Post indicate scores before and after treatment.

**Supplemental Table 2.** Differentially expressed proteins in neuronal extracellular vesicles (nEVs) isolated from patients following CAND treatment compared to the pre-CAND condition and the complete list of all proteins identified in the proteomic analysis.

Each row corresponds to a protein identified by its UniProt accession number, gene symbol, and protein symbol. Expression changes are reported as $\log_2$ fold change (logFC); positive values indicate upregulation after CAND treatment, whereas negative values indicate downregulation. AveExpr represents the average expression across all samples. The t-statistic and associated P value were derived from moderated t-tests. The B statistic reflects the log-odds that a given protein is differentially expressed.

#### Supplemental Table 2. Differentially Expressed Proteins in nEVs

| UniProt | Gene Symbol | Protein Name | logFC | AveExpr | t | P.Value | B |
|---|---|---|---|---|---|---|---|
| P46778 | RPL21 | RL21 | 6.430273765 | 9.748979406 | 3.363691046 | 0.001473651 | -1.265541025 |
| Q99497 | PARK7 | PARK7 | 2.556061761 | 9.508329594 | 3.213247719 | 0.002267459 | -1.551439503 |
| P35637 | FUS | FUS | 2.145018408 | 9.460307707 | 3.020538693 | 0.003838571 | -1.982090763 |
| P18077 | RPL35A | RL35A | 2.13320939 | 11.39346411 | 2.07903997 | 0.042430602 | -3.741001708 |
| Q9NRX4 | PHPT1 | PHP14 | 1.952491539 | 8.126823235 | 2.657806068 | 0.011609861 | -2.778090376 |
| P49189 | ALDH9A1 | AL9A1 | 1.881719038 | 8.956597096 | 2.153484788 | 0.035900912 | -3.610108727 |
| P20336 | RAB3A | RAB3A | 1.865997996 | 9.798318055 | 2.094283649 | 0.040906403 | -3.715400772 |
| P01594 | IGKV1-33 | KV133 | 1.864613652 | 13.40636808 | 2.549460321 | 0.013875674 | -2.9250108 |
| P00740 | F9 | FA9 | 1.797619151 | 11.14518565 | 3.097697927 | 0.003059806 | -1.783472679 |
| Q13938 | CAPS | CAYP1 | 1.748181751 | 11.20708462 | 3.112992895 | 0.002928219 | -1.749978083 |
| P51674 | GPM6A | GPM6A | 1.699523899 | 9.977344172 | 2.145937384 | 0.036273141 | -3.629437694 |
| P0DP09 | IGKV1D-13 | KV113 | 1.642127756 | 13.65181592 | 2.228869763 | 0.030219938 | -3.494066287 |
| P40227 | CCT6A | TCPZ | 1.544325414 | 12.26876077 | 2.868735929 | 0.005820961 | -2.271675529 |
| P19338 | NCL | NUCL | 1.484126594 | 14.240002 | 2.028629437 | 0.047312171 | -3.820267402 |
| P15531 | NME1 | NDKA | 1.456301029 | 8.297834232 | 2.182651104 | 0.034516078 | -3.570234365 |
| Q92522 | H1-10 | H1X | 1.395640369 | 9.944587487 | 2.734569573 | 0.008371482 | -2.545662495 |
| P02652 | APOA2 | APOA2 | 1.341106856 | 13.89357777 | 2.10313014 | 0.040008642 | -3.700102625 |
| P23142 | FBLN1 | FBLN1 | 1.217786786 | 9.47254383 | 2.046261497 | 0.045860481 | -3.781813964 |
| P38646 | HSPA9 | GRP75 | 1.09362884 | 14.21458655 | 2.3030727 | 0.025052351 | -3.360161367 |
| P62937 | PPIA | PPIA | -1.064997521 | 11.22550632 | -2.127188393 | 0.037869941 | -3.660532173 |
| P17844 | DDX5 | DDX5 | -1.097688396 | 11.72587252 | -2.416511471 | 0.018989982 | -3.156397772 |
| P11021 | HSPA5 | BIP | -1.112787495 | 14.82700771 | -2.200972348 | 0.031921602 | -3.536877754 |
| Q9BPU6 | DPYSL5 | DPYL5 | -1.277472957 | 10.87507514 | -2.205027225 | 0.031619963 | -3.529982737 |
| P08758 | ANXA5 | ANXA5 | -1.310282222 | 12.5208057 | -2.054272912 | 0.044677693 | -3.77931354 |
| P55072 | VCP | TERA | -1.361284526 | 13.8859349 | -2.193868072 | 0.032456196 | -3.548933223 |
| P62873 | GNB1 | GBB1 | -1.386189383 | 12.21737201 | -2.24039291 | 0.02909385 | -3.469411367 |
| P26639 | TARS1 | SYTC | -1.452602625 | 9.377103521 | -2.430687115 | 0.018395295 | -3.132119559 |
| Q92823 | NRCAM | NRCAM | -1.492534884 | 13.18445558 | -2.93097586 | 0.004901114 | -2.141474737 |

| | | | | | | | |
|---|---|---|---|---|---|---|---|
| Q08211 | DHX9 | DHX9 | -1.581087625 | 13.43428184 | -2.114539238 | 0.038981626 | -3.681383474 |
| P28838 | LAP3 | AMPL | -1.621096508 | 11.12429347 | -2.740620497 | 0.008237236 | -2.533506773 |
| P06576 | ATP5F1B | ATPB | -1.630639715 | 11.92407979 | -2.326182999 | 0.023693181 | -3.319275505 |
| P06310 | IGKV2-30 | KV230 | -1.649017815 | 13.49282589 | -2.452770146 | 0.017350956 | -3.089661977 |
| P55290 | CDH13 | CAD13 | -1.661684628 | 9.449083416 | -2.03149106 | 0.047492911 | -3.805593386 |
| P00736 | C1R | C1R | -1.66925474 | 13.74472205 | -2.131716637 | 0.037478758 | -3.653042788 |
| Q92804 | TAF15 | RBP56 | -1.716167034 | 9.172633519 | -2.238189075 | 0.029842382 | -3.475668167 |
| P31939 | ATIC | PUR9 | -1.926605071 | 12.54945522 | -2.647469258 | 0.010539889 | -2.718476783 |
| P01860 | IGHG3 | IGHG3 | -1.996337459 | 14.1756181 | -2.455768173 | 0.01722131 | -3.084109685 |
| A0A075B6I9 | IGLV7-46 | LV746 | -2.229648914 | 8.760973137 | -2.161328697 | 0.035443774 | -3.599568184 |
| P60981 | DSTN | DEST | -2.283187244 | 11.21887308 | -3.21457334 | 0.002198593 | -1.530479274 |
| O75874 | IDH1 | IDHC | -2.411163335 | 10.43066861 | -2.572450795 | 0.012806505 | -2.86401488 |
| P60900 | PSMA6 | PSA6 | -2.415898911 | 10.92947557 | -2.448041424 | 0.017557231 | -3.098408878 |
| A0A075B6K5 | IGLV3-9 | LV39 | -2.456656756 | 12.2728433 | -2.751077488 | 0.008009893 | -2.51245426 |
| O43813 | LANCL1 | LANC1 | -2.582167583 | 12.55751351 | -3.56146683 | 0.000766388 | -0.723331895 |
| P22102 | GART | PUR2 | -2.631213321 | 10.2812548 | -2.927628388 | 0.005009725 | -2.156363162 |
| P51178-2 | PLCD1 | PLCD1 | -2.782028265 | 9.24043183 | -3.189426969 | 0.002451847 | -1.645434207 |
| P08238 | HSP90AB1 | HS90B | -2.884911646 | 13.56843396 | -4.701192645 | 1.75622E-05 | 2.184252907 |

#### Supplemental Table 2. All Quantified proteins

| UniProt | Gene Symbol | logFC | AveExpr | t | P.Value | adj.P.Val | B |
|---|---|---|---|---|---|---|---|
| P08238 | HSP90AB1 | -2.88491 | 13.56843 | -4.70119 | 1.76E-05 | 0.015279 | 2.184253 |
| O43813 | LANCL1 | -2.58217 | 12.55751 | -3.56147 | 0.000766 | 0.332754 | -0.72333 |
| P46778 | RPL21 | 6.430274 | 9.748979 | 3.363691 | 0.001474 | 0.332754 | -1.26554 |
| P60981 | DSTN | -2.28319 | 11.21887 | -3.21457 | 0.002199 | 0.332754 | -1.53048 |
| Q99497 | PARK7 | 2.556062 | 9.50833 | 3.213248 | 0.002267 | 0.332754 | -1.55144 |
| P51178-2 | #N/D | -2.78203 | 9.240432 | -3.18943 | 0.002452 | 0.332754 | -1.64543 |
| Q13938 | #N/D | 1.748182 | 11.20708 | 3.112993 | 0.002928 | 0.332754 | -1.74998 |
| P00740 | F9 | 1.797619 | 11.14519 | 3.097698 | 0.00306 | 0.332754 | -1.78347 |
| P35637 | FUS | 2.145018 | 9.460308 | 3.020539 | 0.003839 | 0.371062 | -1.98209 |
| Q92823 | NRCAM | -1.49253 | 13.18446 | -2.93098 | 0.004901 | 0.396224 | -2.14147 |
| P22102 | GART | -2.63121 | 10.28125 | -2.92763 | 0.00501 | 0.396224 | -2.15636 |
| P40227 | CCT6A | 1.544325 | 12.26876 | 2.868736 | 0.005821 | 0.42202 | -2.27168 |
| A0A075B6I | #N/D | -2.45666 | 12.27284 | -2.75108 | 0.00801 | 0.485546 | -2.51245 |
| P28838 | LAP3 | -1.6211 | 11.12429 | -2.74062 | 0.008237 | 0.485546 | -2.53351 |
| Q92522 | H1-10 | 1.39564 | 9.944587 | 2.73457 | 0.008371 | 0.485546 | -2.54566 |
| P31939 | ATIC | -1.92661 | 12.54946 | -2.64747 | 0.01054 | 0.573106 | -2.71848 |
| Q9NRX4 | PHPT1 | 1.952492 | 8.126823 | 2.657806 | 0.01161 | 0.594152 | -2.77809 |
| O75874 | IDH1 | -2.41116 | 10.43067 | -2.57245 | 0.012807 | 0.618981 | -2.86401 |
| P01594 | #N/D | 1.864614 | 13.40637 | 2.54946 | 0.013876 | 0.63536 | -2.92501 |
| P01860 | #N/D | -1.99634 | 14.17562 | -2.45577 | 0.017221 | 0.688387 | -3.08411 |
| P06310 | #N/D | -1.64902 | 13.49283 | -2.45277 | 0.017351 | 0.688387 | -3.08966 |
| P60900 | PSMA6 | -2.4159 | 10.92948 | -2.44804 | 0.017557 | 0.688387 | -3.09841 |
| P26639 | TARS1 | -1.4526 | 9.377104 | -2.43069 | 0.018395 | 0.688387 | -3.13212 |
| P17844 | DDX5 | -1.09769 | 11.72587 | -2.41651 | 0.01899 | 0.688387 | -3.1564 |
| P06576 | ATP5F1B | -1.63064 | 11.92408 | -2.32618 | 0.023693 | 0.824523 | -3.31928 |
| P38646 | HSPA9 | 1.093629 | 14.21459 | 2.303073 | 0.025052 | 0.825278 | -3.36016 |
| P62873 | GNB1 | -1.38619 | 12.21737 | -2.24039 | 0.029094 | 0.825278 | -3.46941 |
| Q92804 | TAF15 | -1.71617 | 9.172634 | -2.23819 | 0.029842 | 0.825278 | -3.47567 |

| | | | | | | | |
|---|---|---|---|---|---|---|---|
| P0DP09 | #N/D | 1.642128 | 13.65182 | 2.22887 | 0.03022 | 0.825278 | -3.49407 |
| Q9BPU6 | DPYSL5 | -1.27747 | 10.87508 | -2.20503 | 0.03162 | 0.825278 | -3.52998 |
| P11021 | HSPA5 | -1.11279 | 14.82701 | -2.20097 | 0.031922 | 0.825278 | -3.53688 |
| P55072 | VCP | -1.36128 | 13.88593 | -2.19387 | 0.032456 | 0.825278 | -3.54893 |
| P15531 | NME1 | 1.456301 | 8.297834 | 2.182651 | 0.034516 | 0.825278 | -3.57023 |
| A0A075B6I | #N/D | -2.22965 | 8.760973 | -2.16133 | 0.035444 | 0.825278 | -3.59957 |
| P49189 | ALDH9A1 | 1.881719 | 8.956597 | 2.153485 | 0.035901 | 0.825278 | -3.61011 |
| P51674 | GPM6A | 1.699524 | 9.977344 | 2.145937 | 0.036273 | 0.825278 | -3.62944 |
| P00736 | C1R | -1.66925 | 13.74472 | -2.13172 | 0.037479 | 0.825278 | -3.65304 |
| P62937 | PPIA | -1.065 | 11.22551 | -2.12719 | 0.03787 | 0.825278 | -3.66053 |
| Q08211 | DHX9 | -1.58109 | 13.43428 | -2.11454 | 0.038982 | 0.825278 | -3.68138 |
| P02652 | APOA2 | 1.341107 | 13.89358 | 2.10313 | 0.040009 | 0.825278 | -3.7001 |
| P20336 | RAB3A | 1.865998 | 9.798318 | 2.094284 | 0.040906 | 0.825278 | -3.7154 |
| P18077 | RPL35A | 2.133209 | 11.39346 | 2.07904 | 0.042431 | 0.825278 | -3.741 |
| P08758 | ANXA5 | -1.31028 | 12.52081 | -2.05427 | 0.044678 | 0.825278 | -3.77931 |
| P23142 | FBLN1 | 1.217787 | 9.472544 | 2.046261 | 0.04586 | 0.825278 | -3.78181 |
| P19338 | NCL | 1.484127 | 14.24 | 2.028629 | 0.047312 | 0.825278 | -3.82027 |
| P55290 | CDH13 | -1.66168 | 9.449083 | -2.03149 | 0.047493 | 0.825278 | -3.80559 |
| O43301 | HSPA12A | 1.241831 | 10.41348 | 1.993854 | 0.051284 | 0.825278 | -3.86118 |
| Q9UDR5 | AASS | 2.542155 | 10.55739 | 1.980886 | 0.052576 | 0.825278 | -3.89536 |
| A0A0C4DH | #N/D | 1.443328 | 9.696796 | 1.975453 | 0.053205 | 0.825278 | -3.90381 |
| P83731 | RPL24 | -1.55708 | 8.556325 | -1.98709 | 0.053396 | 0.825278 | -3.86998 |
| P0DOX3 | #N/D | 0.859642 | 13.40279 | 1.973717 | 0.053408 | 0.825278 | -3.90651 |
| P11766 | ADH5 | -1.7276 | 10.15171 | -1.97288 | 0.053506 | 0.825278 | -3.90781 |
| P06733 | ENO1 | -0.81586 | 12.59836 | -1.95473 | 0.055667 | 0.825278 | -3.93587 |
| A0A075B6I | #N/D | 0.845034 | 13.46962 | 1.947798 | 0.056512 | 0.825278 | -3.94652 |
| O00429 | DNM1L | 1.227878 | 7.713053 | 1.956777 | 0.057138 | 0.825278 | -3.92694 |
| P80748 | #N/D | 1.38119 | 13.59399 | 1.937177 | 0.057828 | 0.825278 | -3.96279 |
| Q13838 | DDX39B | -1.81438 | 13.4483 | -1.93358 | 0.05828 | 0.825278 | -3.96829 |
| P09543 | CNP | -0.8087 | 12.18182 | -1.93064 | 0.058651 | 0.825278 | -3.97276 |
| Q08209-3 | #N/D | -1.1371 | 10.11222 | -1.92624 | 0.059304 | 0.825278 | -3.97999 |

| | | | | | | | |
|---|---|---|---|---|---|---|---|
| Q07954 | LRP1 | -1.38531 | 9.068573 | -1.91098 | 0.061483 | 0.825278 | -4.00419 |
| Q9UHG2 | PCSK1N | -1.53774 | 7.955461 | -1.91831 | 0.061984 | 0.825278 | -3.95663 |
| P51659 | HSD17B4 | 0.963802 | 12.08701 | 1.90075 | 0.062541 | 0.825278 | -4.018 |
| P84103 | SRSF3 | 1.300193 | 7.821272 | 1.897797 | 0.064234 | 0.825278 | -3.98232 |
| P37840 | SNCA | 1.55265 | 9.699817 | 1.888823 | 0.06435 | 0.825278 | -4.03687 |
| P01766 | #N/D | 1.36372 | 9.036312 | 1.889628 | 0.064682 | 0.825278 | -4.01925 |
| P23515 | OMG | -1.01445 | 10.03831 | -1.87827 | 0.06561 | 0.825278 | -4.05163 |
| Q9NRV9 | HEBP1 | -1.6609 | 10.05472 | -1.8786 | 0.066205 | 0.825278 | -4.05421 |
| Q9GZV7 | HAPLN2 | -1.08556 | 13.03861 | -1.86907 | 0.066901 | 0.825278 | -4.06529 |
| P02768 | ALB | 1.615821 | 17.61783 | 1.868342 | 0.067004 | 0.825278 | -4.06637 |
| Q9Y6R7 | FCGBP | 0.964453 | 13.98588 | 1.860171 | 0.068172 | 0.825278 | -4.07845 |
| P99999 | CYCS | -1.29323 | 7.341751 | -1.8809 | 0.06959 | 0.825278 | -4.02807 |
| Q15485 | FCN2 | -1.82909 | 12.63067 | -1.8408 | 0.071008 | 0.825278 | -4.10691 |
| P04004 | VTN | -1.06522 | 14.98777 | -1.83148 | 0.072406 | 0.825278 | -4.1205 |
| P04843 | RPN1 | -1.00991 | 8.447986 | -1.82863 | 0.073148 | 0.825278 | -4.12589 |
| P06396 | GSN | 1.148982 | 14.7431 | 1.825446 | 0.073324 | 0.825278 | -4.12928 |
| P35580 | MYH10 | 1.350564 | 9.704816 | 1.822772 | 0.073734 | 0.825278 | -4.13316 |
| P20851-2 | #N/D | 1.401946 | 11.52893 | 1.815153 | 0.074913 | 0.825278 | -4.14419 |
| P67775 | PPP2CA | 1.596206 | 8.996608 | 1.809822 | 0.076664 | 0.825278 | -4.10942 |
| Q9UHD8 | SEPTIN9 | -1.05082 | 7.179257 | -1.82268 | 0.076803 | 0.825278 | -4.10035 |
| O75083 | WDR1 | -0.8886 | 11.68371 | -1.79958 | 0.077372 | 0.825278 | -4.1666 |
| P01703 | #N/D | 1.58571 | 13.56328 | 1.796629 | 0.077846 | 0.825278 | -4.17083 |
| Q14974 | KPNB1 | -2.02045 | 12.05301 | -1.79653 | 0.077861 | 0.825278 | -4.17097 |
| P30084 | ECHS1 | 0.83458 | 10.15609 | 1.778509 | 0.080806 | 0.825278 | -4.19667 |
| P21796 | VDAC1 | -1.47587 | 8.076572 | -1.78005 | 0.082233 | 0.825278 | -4.17812 |
| Q99880 | H2BC13 | 1.292302 | 12.3185 | 1.769225 | 0.082359 | 0.825278 | -4.20981 |
| P61247 | RPS3A | -0.77169 | 12.29605 | -1.76806 | 0.082555 | 0.825278 | -4.21146 |
| P49588 | AARS1 | 2.268776 | 10.01352 | 1.756554 | 0.085067 | 0.825278 | -4.22933 |
| P0DOY3 | #N/D | 1.511527 | 15.80861 | 1.752523 | 0.085215 | 0.825278 | -4.23331 |
| P61626 | LYZ | 1.378607 | 13.1144 | 1.73785 | 0.087792 | 0.825278 | -4.2538 |
| P01023 | A2M | 1.25439 | 15.60344 | 1.737525 | 0.08785 | 0.825278 | -4.25425 |

| | | | | | | | |
|---|---|---|---|---|---|---|---|
| Q9NRW1 | RAB6B | 1.251203 | 9.368933 | 1.730849 | 0.089252 | 0.825278 | -4.24066 |
| P49207 | RPL34 | -1.36214 | 9.206154 | -1.72849 | 0.089571 | 0.825278 | -4.26706 |
| P15814 | IGLL1 | 2.351696 | 11.9045 | 1.727178 | 0.090259 | 0.825278 | -4.2701 |
| Q9BW30 | TPPP3 | 1.008086 | 7.409834 | 1.735282 | 0.091639 | 0.825278 | -4.18823 |
| Q92686 | NRGN | 1.410011 | 9.047773 | 1.711876 | 0.092831 | 0.825278 | -4.2905 |
| P14174 | MIF | 1.484711 | 11.07338 | 1.699848 | 0.094767 | 0.825278 | -4.30614 |
| P01031 | C5 | 0.654215 | 12.98874 | 1.698593 | 0.095005 | 0.825278 | -4.30786 |
| A0A0C4DH | #N/D | 1.276332 | 12.36129 | 1.696615 | 0.095381 | 0.825278 | -4.31055 |
| O75340 | PDCD6 | -1.44722 | 7.338176 | -1.72227 | 0.095522 | 0.825278 | -4.20802 |
| P53680 | AP2S1 | -1.09557 | 7.833872 | -1.70009 | 0.096436 | 0.825278 | -4.30994 |
| P01871 | #N/D | 1.127864 | 16.51568 | 1.689983 | 0.09665 | 0.825278 | -4.31957 |
| P07737 | PFN1 | -1.72217 | 12.48645 | -1.69232 | 0.096757 | 0.825278 | -4.29317 |
| Q07020 | RPL18 | -1.60169 | 11.58711 | -1.67477 | 0.099616 | 0.841413 | -4.34014 |
| P0DMV9 | HSPA1A | -0.73222 | 12.81602 | -1.66507 | 0.101543 | 0.849446 | -4.35315 |
| P42704 | LRPPRC | 1.521426 | 13.36178 | 1.650453 | 0.104508 | 0.858062 | -4.37265 |
| P04406 | GAPDH | -1.1357 | 14.71391 | -1.65027 | 0.104546 | 0.858062 | -4.3729 |
| P43243 | MATR3 | -1.73556 | 7.788896 | -1.64091 | 0.109427 | 0.872248 | -4.28591 |
| P02745 | C1QA | 1.457295 | 10.24676 | 1.627231 | 0.109802 | 0.872248 | -4.40409 |
| P30153 | PPP2R1A | -1.27299 | 10.68953 | -1.62332 | 0.110197 | 0.872248 | -4.40844 |
| P48047 | ATP5PO | 1.723735 | 12.76757 | 1.619653 | 0.110985 | 0.872248 | -4.41324 |
| Q16799 | RTN1 | 1.137802 | 10.36556 | 1.616507 | 0.111664 | 0.872248 | -4.41735 |
| P09382 | LGALS1 | -1.17168 | 9.996774 | -1.60718 | 0.113699 | 0.872248 | -4.42948 |
| Q14204 | DYNC1H1 | -0.76957 | 13.22936 | -1.59534 | 0.116323 | 0.872248 | -4.44479 |
| P04181 | OAT | -1.14737 | 9.027395 | -1.59592 | 0.116407 | 0.872248 | -4.44437 |
| P51149 | RAB7A | -0.78379 | 8.780581 | -1.58432 | 0.119137 | 0.872248 | -4.45942 |
| P62805 | H4C9 | 1.228832 | 11.74498 | 1.581205 | 0.119523 | 0.872248 | -4.46294 |
| O00425 | IGF2BP3 | 1.235616 | 8.291206 | 1.582616 | 0.120938 | 0.872248 | -4.40111 |
| P01704 | #N/D | 0.975983 | 11.22917 | 1.570966 | 0.121884 | 0.872248 | -4.476 |
| P15880 | RPS2 | 1.434915 | 12.70417 | 1.568982 | 0.122345 | 0.872248 | -4.47852 |
| Q9UQM7 | CAMK2A | 1.230647 | 11.74729 | 1.562019 | 0.123977 | 0.872248 | -4.48735 |
| P12814 | ACTN1 | -1.25882 | 7.963896 | -1.57151 | 0.124484 | 0.872248 | -4.41574 |

| | | | | | | | |
|---|---|---|---|---|---|---|---|
| Q99878 | H2AC14 | 1.315814 | 8.686371 | 1.562333 | 0.125003 | 0.872248 | -4.46026 |
| O95445 | APOM | 1.679751 | 11.61754 | 1.55633 | 0.125323 | 0.872248 | -4.49453 |
| P39687 | ANP32A | -1.11736 | 9.050789 | -1.55359 | 0.126416 | 0.872248 | -4.47019 |
| O75347 | TBCA | 1.065897 | 9.096358 | 1.547223 | 0.127502 | 0.872248 | -4.50598 |
| P08865 | RPSA | -0.94773 | 13.49287 | -1.53897 | 0.129504 | 0.872248 | -4.51631 |
| P02788 | LTF | 1.565916 | 9.520811 | 1.538689 | 0.129784 | 0.872248 | -4.51689 |
| P48539 | PCP4 | 0.838332 | 8.670542 | 1.534763 | 0.130745 | 0.872248 | -4.52178 |
| P08133 | ANXA6 | -0.66604 | 11.75849 | -1.53176 | 0.131272 | 0.872248 | -4.52529 |
| Q13228 | SELENBP1 | -0.8141 | 10.91175 | -1.53119 | 0.131412 | 0.872248 | -4.52599 |
| P30038 | ALDH4A1 | -0.78601 | 11.58862 | -1.5188 | 0.134499 | 0.872248 | -4.54133 |
| P25705 | ATP5F1A | 1.518147 | 13.38486 | 1.518301 | 0.134623 | 0.872248 | -4.54194 |
| P11166 | SLC2A1 | -1.18442 | 9.8587 | -1.51538 | 0.135359 | 0.872248 | -4.54554 |
| P06727 | #N/D | 0.668077 | 14.15249 | 1.514525 | 0.135576 | 0.872248 | -4.54659 |
| P18669 | PGAM1 | -0.82686 | 13.03714 | -1.50744 | 0.137378 | 0.872248 | -4.55529 |
| Q5IS61 | #N/D | 1.051465 | 8.614239 | 1.509413 | 0.137561 | 0.872248 | -4.52405 |
| P35542 | SAA4 | 0.773971 | 12.48858 | 1.503204 | 0.138465 | 0.872248 | -4.56047 |
| P17600 | SYN1 | 1.145721 | 12.09097 | 1.4963 | 0.14025 | 0.872248 | -4.56889 |
| Q15424 | SAFB | -0.88795 | 10.29881 | -1.47738 | 0.145339 | 0.872248 | -4.59185 |
| P15104 | GLUL | 1.086461 | 13.57473 | 1.470491 | 0.147084 | 0.872248 | -4.60004 |
| P45880 | VDAC2 | -0.80026 | 11.73893 | -1.46396 | 0.148856 | 0.872248 | -4.60785 |
| Q92598-4 | #N/D | 1.343908 | 8.188924 | 1.472089 | 0.148938 | 0.872248 | -4.52997 |
| P35998 | PSMC2 | 1.45843 | 10.28438 | 1.464294 | 0.149086 | 0.872248 | -4.57707 |
| P02656 | APOC3 | 1.119032 | 12.92227 | 1.462588 | 0.149229 | 0.872248 | -4.60948 |
| P07195 | LDHB | -0.65689 | 13.41831 | -1.4585 | 0.150348 | 0.872248 | -4.61435 |
| P06681 | C2 | -0.88267 | 10.88017 | -1.45927 | 0.150457 | 0.872248 | -4.61363 |
| Q00610 | CLTC | 1.167318 | 13.78944 | 1.456409 | 0.150922 | 0.872248 | -4.61683 |
| P46777 | RPL5 | -1.07595 | 8.455774 | -1.45987 | 0.15121 | 0.872248 | -4.58283 |
| Q14123 | PDE1C | 1.765082 | 9.472294 | 1.45347 | 0.151944 | 0.872248 | -4.62044 |
| P09651 | HNRNPA1 | 0.949652 | 12.21855 | 1.451868 | 0.152177 | 0.872248 | -4.62221 |
| P00492 | HPRT1 | 1.288752 | 10.76362 | 1.451986 | 0.152247 | 0.872248 | -4.62213 |
| Q02878 | RPL6 | 1.242924 | 11.19865 | 1.445576 | 0.153928 | 0.872248 | -4.62964 |

| | | | | | | | |
|---|---|---|---|---|---|---|---|
| Q96F85 | CNRIP1 | 0.832135 | 10.63719 | 1.444305 | 0.154284 | 0.872248 | -4.63114 |
| Q16629 | SRSF7 | 1.186821 | 9.577583 | 1.443457 | 0.154957 | 0.872248 | -4.63238 |
| P01019 | AGT | -1.17828 | 13.11805 | -1.4381 | 0.156029 | 0.872248 | -4.63843 |
| P06331 | #N/D | 0.992861 | 11.07987 | 1.435416 | 0.156789 | 0.872248 | -4.64158 |
| P22626 | HNRNPA2B1 | -0.66356 | 12.6985 | -1.43325 | 0.157406 | 0.872248 | -4.64412 |
| P11940 | PABPC1 | -0.94732 | 9.661148 | -1.42792 | 0.159136 | 0.876258 | -4.61897 |
| P51148 | RAB5C | 0.917245 | 8.07766 | 1.428927 | 0.160349 | 0.877381 | -4.59249 |
| P13533 | MYH6 | -0.7761 | 10.46117 | -1.41776 | 0.161859 | 0.88011 | -4.66214 |
| P40925 | MDH1 | -0.77858 | 13.09244 | -1.41012 | 0.164093 | 0.88323 | -4.67097 |
| Q99426 | TBCB | 0.996474 | 8.295429 | 1.410318 | 0.165738 | 0.88323 | -4.61304 |
| P62081 | RPS7 | -0.86368 | 10.42682 | -1.40237 | 0.166384 | 0.88323 | -4.67989 |
| Q06830 | PRDX1 | -0.73433 | 12.88336 | -1.39022 | 0.17002 | 0.88323 | -4.69375 |
| P00918 | CA2 | 0.89479 | 9.133649 | 1.386773 | 0.171271 | 0.88323 | -4.66528 |
| P19367 | HK1 | -0.66488 | 11.89524 | -1.38599 | 0.171304 | 0.88323 | -4.69856 |
| P39023 | RPL3 | -1.73655 | 11.33349 | -1.38518 | 0.171547 | 0.88323 | -4.69947 |
| P02655 | APOC2 | 1.060126 | 11.22537 | 1.383795 | 0.172177 | 0.88323 | -4.66858 |
| P19823 | ITIH2 | 0.813246 | 14.69691 | 1.382334 | 0.172415 | 0.88323 | -4.7027 |
| P62750 | RPL23A | 0.85137 | 11.64551 | 1.381226 | 0.172754 | 0.88323 | -4.70395 |
| Q13813 | SPTAN1 | -0.50602 | 11.99901 | -1.37846 | 0.1736 | 0.88323 | -4.70708 |
| P04196 | HRG | -1.4288 | 14.14042 | -1.37198 | 0.175599 | 0.883847 | -4.71438 |
| Q15185 | PTGES3 | -1.53261 | 14.08679 | -1.37181 | 0.175754 | 0.883847 | -4.71459 |
| P10720 | PF4V1 | 1.159096 | 11.14838 | 1.367282 | 0.177159 | 0.885793 | -4.71967 |
| Q9P258 | RCC2 | -0.74425 | 11.42629 | -1.35724 | 0.180206 | 0.895881 | -4.73085 |
| O00410 | IPO5 | -1.08108 | 9.76488 | -1.33941 | 0.185906 | 0.918968 | -4.75057 |
| Q9UBB6 | NCDN | -1.09247 | 8.646641 | -1.33252 | 0.188346 | 0.92577 | -4.75814 |
| P43487 | RANBP1 | -0.96947 | 11.29012 | -1.32135 | 0.19247 | 0.932648 | -4.73637 |
| O94856 | NFASC | -0.82119 | 13.27301 | -1.31861 | 0.192725 | 0.932648 | -4.77327 |
| P68871 | HBB | -1.08772 | 14.57697 | -1.31525 | 0.193841 | 0.932648 | -4.77689 |
| P68366 | TUBA4A | -0.73796 | 8.182302 | -1.31316 | 0.195319 | 0.932648 | -4.74504 |
| Q92777 | SYN2 | 1.255983 | 10.07691 | 1.307798 | 0.196341 | 0.932648 | -4.78493 |
| Q9H4G4 | GLIPR2 | -0.90123 | 9.978347 | -1.30383 | 0.197779 | 0.932648 | -4.78918 |

| | | | | | | | |
|---|---|---|---|---|---|---|---|
| P17096 | HMGA1 | 1.286469 | 10.9518 | 1.302387 | 0.198171 | 0.932648 | -4.79073 |
| P06454 | PTMA | 0.971143 | 9.310123 | 1.301518 | 0.198466 | 0.932648 | -4.79166 |
| P15313 | ATP6V1B1 | -0.83631 | 8.509029 | -1.29807 | 0.200057 | 0.932648 | -4.79532 |
| P01714 | #N/D | 1.119817 | 10.84083 | 1.287406 | 0.203304 | 0.932648 | -4.80669 |
| P60201 | PLP1 | 0.706866 | 12.86375 | 1.272712 | 0.208434 | 0.932648 | -4.82217 |
| P08559-4 | #N/D | -0.8575 | 10.42252 | -1.26922 | 0.209666 | 0.932648 | -4.82582 |
| P23528 | CFL1 | -0.75747 | 12.52033 | -1.26871 | 0.209847 | 0.932648 | -4.82635 |
| A0A0J9YXX | #N/D | 0.977144 | 10.65969 | 1.266506 | 0.21063 | 0.932648 | -4.82866 |
| Q5IS67 | #N/D | 1.258265 | 8.456854 | 1.267393 | 0.211475 | 0.932648 | -4.79245 |
| P39748 | FEN1 | -1.54476 | 9.947938 | -1.26234 | 0.212113 | 0.932648 | -4.833 |
| P68104 | EEF1A1 | -0.88568 | 9.086982 | -1.26135 | 0.212563 | 0.932648 | -4.83401 |
| Q15334 | LLGL1 | 0.881187 | 8.598936 | 1.260726 | 0.21269 | 0.932648 | -4.83467 |
| Q9UMF0 | ICAM5 | 0.813602 | 9.084601 | 1.248623 | 0.217053 | 0.932648 | -4.84719 |
| P05091 | ALDH2 | -0.73998 | 12.45982 | -1.24698 | 0.21765 | 0.932648 | -4.84888 |
| P30040 | ERP29 | -1.28394 | 8.785068 | -1.24421 | 0.220097 | 0.932648 | -4.81582 |
| P84077 | ARF1 | -0.89872 | 9.006425 | -1.23624 | 0.221678 | 0.932648 | -4.85985 |
| P29972 | AQP1 | 0.969752 | 8.612235 | 1.234554 | 0.222835 | 0.932648 | -4.82563 |
| P48735 | IDH2 | -0.81322 | 11.01398 | -1.22773 | 0.224737 | 0.932648 | -4.86852 |
| P61026 | RAB10 | 0.907104 | 8.602059 | 1.231319 | 0.224832 | 0.932648 | -4.82866 |
| Q15631 | TSN | -1.03152 | 8.819731 | -1.22757 | 0.225094 | 0.932648 | -4.86861 |
| P61956 | SUMO2 | -1.07032 | 9.670243 | -1.22638 | 0.225336 | 0.932648 | -4.86987 |
| P09661 | SNRPA1 | 1.505491 | 11.24558 | 1.224571 | 0.225917 | 0.932648 | -4.87172 |
| P04350 | TUBB4A | -0.83942 | 11.70512 | -1.22436 | 0.225998 | 0.932648 | -4.87194 |
| P49006 | MARCKSL1 | 1.237088 | 8.963973 | 1.223704 | 0.226434 | 0.932648 | -4.87254 |
| P14314 | PRKCSH | 0.954106 | 12.48603 | 1.223748 | 0.22652 | 0.932648 | -4.87247 |
| P01718 | #N/D | 0.808163 | 12.55539 | 1.222826 | 0.226571 | 0.932648 | -4.87349 |
| Q9NZL9 | MAT2B | -1.03521 | 9.370227 | -1.22249 | 0.226791 | 0.932648 | -4.8738 |
| P50395 | GDI2 | -0.64823 | 11.20433 | -1.21928 | 0.227902 | 0.932648 | -4.87706 |
| P00738 | HP | 0.620282 | 16.23776 | 1.219105 | 0.227968 | 0.932648 | -4.87724 |
| P01861 | #N/D | 0.693718 | 9.667126 | 1.213119 | 0.230523 | 0.932648 | -4.88316 |
| P22314 | UBA1 | -0.80496 | 12.03395 | -1.2093 | 0.231681 | 0.932648 | -4.88706 |

| | | | | | | | |
|---|---|---|---|---|---|---|---|
| Q16851 | UGP2 | -0.47634 | 12.72477 | -1.20742 | 0.232399 | 0.932648 | -4.88894 |
| P62241 | RPS8 | -0.70156 | 9.619243 | -1.20552 | 0.233124 | 0.932648 | -4.89083 |
| Q15257-2 | #N/D | 1.192803 | 9.172995 | 1.204023 | 0.234204 | 0.932648 | -4.85572 |
| P36354 | #N/D | 0.997362 | 8.62113 | 1.205548 | 0.234521 | 0.932648 | -4.89034 |
| P48147 | PREP | -0.87543 | 7.680943 | -1.2021 | 0.235992 | 0.932648 | -4.82633 |
| Q14894 | CRYM | 0.731241 | 12.35016 | 1.191851 | 0.238399 | 0.932648 | -4.90438 |
| O95336 | PGLS | -0.94773 | 10.2894 | -1.18047 | 0.242855 | 0.932648 | -4.91554 |
| Q7L099-4 | #N/D | -0.72006 | 10.43573 | -1.17297 | 0.245824 | 0.932648 | -4.92284 |
| P10620 | MGST1 | -0.90493 | 10.88042 | -1.17174 | 0.246316 | 0.932648 | -4.92404 |
| O43761 | SYNGR3 | 1.110012 | 10.57735 | 1.169011 | 0.247403 | 0.932648 | -4.92668 |
| Q13177 | PAK2 | -0.86772 | 8.338763 | -1.17246 | 0.247406 | 0.932648 | -4.88572 |
| P67936 | TPM4 | 1.080868 | 9.11576 | 1.170075 | 0.247944 | 0.932648 | -4.85658 |
| P80108 | GPLD1 | 0.775463 | 9.224428 | 1.167362 | 0.248347 | 0.932648 | -4.92816 |
| P62899 | RPL31 | -1.22062 | 12.22519 | -1.16138 | 0.250467 | 0.932648 | -4.93404 |
| A0A075B6I | #N/D | 1.270762 | 11.3767 | 1.156893 | 0.252561 | 0.932648 | -4.90083 |
| Q9NR30 | DDX21 | 0.818808 | 8.743727 | 1.155007 | 0.253326 | 0.932648 | -4.87074 |
| P61020 | RAB5B | -0.68291 | 8.987505 | -1.15164 | 0.254795 | 0.932648 | -4.9057 |
| Q00839 | HNRNPU | -0.60488 | 12.5168 | -1.15043 | 0.254908 | 0.932648 | -4.94453 |
| P00748 | F12 | -0.56756 | 11.60206 | -1.14526 | 0.257023 | 0.932648 | -4.94944 |
| P61088 | UBE2N | 0.8563 | 11.45261 | 1.14481 | 0.257392 | 0.932648 | -4.94978 |
| P17174 | GOT1 | -0.77613 | 13.06163 | -1.14398 | 0.257549 | 0.932648 | -4.95065 |
| P16401 | H1-5 | 0.880689 | 10.04102 | 1.143847 | 0.257604 | 0.932648 | -4.95078 |
| A0A0C4DH | #N/D | 1.039723 | 9.855679 | 1.143421 | 0.258373 | 0.932648 | -4.91326 |
| Q14624 | ITIH4 | 1.083734 | 9.94507 | 1.140826 | 0.259333 | 0.932648 | -4.91571 |
| A0A0B4J1V | #N/D | 0.811002 | 12.65679 | 1.139114 | 0.259556 | 0.932648 | -4.95526 |
| P15169 | CPN1 | 0.88386 | 9.464541 | 1.138508 | 0.260086 | 0.932648 | -4.9557 |
| P29401 | TKT | -0.52359 | 12.31095 | -1.12947 | 0.263563 | 0.932648 | -4.96432 |
| P37837 | TALDO1 | -0.58289 | 13.06675 | -1.12218 | 0.266624 | 0.932648 | -4.97113 |
| Q6PCE3 | PGM2L1 | -0.91895 | 10.75219 | -1.11992 | 0.26758 | 0.932648 | -4.97324 |
| Q14520 | HABP2 | -1.0167 | 9.292678 | -1.11893 | 0.268272 | 0.932648 | -4.93589 |
| P16152 | CBR1 | -0.63593 | 14.15987 | -1.11552 | 0.269441 | 0.932648 | -4.97731 |

| | | | | | | | |
|---|---|---|---|---|---|---|---|
| P07900-2 | #N/D | 0.791858 | 14.11273 | 1.114804 | 0.269746 | 0.932648 | -4.97798 |
| P60174 | TPI1 | -1.14758 | 13.33968 | -1.11377 | 0.270185 | 0.932648 | -4.97893 |
| Q99747 | NAPG | 0.568065 | 8.259074 | 1.112333 | 0.270885 | 0.932648 | -4.98022 |
| P11279 | LAMP1 | -0.7158 | 9.296891 | -1.11301 | 0.270981 | 0.932648 | -4.97938 |
| P38117 | ETFB | -0.89772 | 8.695554 | -1.11164 | 0.271782 | 0.932648 | -4.98053 |
| P25398 | RPS12 | 0.714017 | 8.911054 | 1.109608 | 0.272233 | 0.932648 | -4.94433 |
| O43175 | PHGDH | -0.49876 | 13.11601 | -1.10874 | 0.272329 | 0.932648 | -4.98357 |
| P26373 | RPL13 | 0.648683 | 10.14526 | 1.108401 | 0.272476 | 0.932648 | -4.98389 |
| P16070 | CD44 | -0.81219 | 8.029181 | -1.10345 | 0.275644 | 0.932648 | -4.98785 |
| P06748 | NPM1 | -1.25924 | 9.025254 | -1.10384 | 0.276036 | 0.932648 | -4.87089 |
| Q15365 | PCBP1 | 0.895499 | 11.13094 | 1.098139 | 0.277069 | 0.932648 | -4.95467 |
| Q6YN16 | HSDL2 | -1.04884 | 9.242755 | -1.09734 | 0.277605 | 0.932648 | -4.95528 |
| P13611 | VCAN | -0.4121 | 12.41909 | -1.09056 | 0.280188 | 0.932648 | -5.00018 |
| P02747 | C1QC | 1.109112 | 14.27094 | 1.083606 | 0.283232 | 0.932648 | -5.00645 |
| P30086 | PEBP1 | -0.51569 | 12.34105 | -1.07785 | 0.28577 | 0.932648 | -5.01162 |
| Q5IS74 | #N/D | -0.84651 | 8.979446 | -1.07752 | 0.286004 | 0.932648 | -5.01187 |
| Q5U7I5 | #N/D | 1.251939 | 12.18705 | 1.075828 | 0.286751 | 0.932648 | -5.01338 |
| P45974 | USP5 | -0.62181 | 9.969402 | -1.07515 | 0.286966 | 0.932648 | -5.01404 |
| P09493-4 | #N/D | 0.719255 | 12.6635 | 1.071208 | 0.288719 | 0.932648 | -5.01756 |
| P62277 | RPS13 | 0.648097 | 10.17611 | 1.070647 | 0.288969 | 0.932648 | -5.01806 |
| P10515 | DLAT | 0.731756 | 8.7176 | 1.0691 | 0.289832 | 0.932648 | -4.98028 |
| P63241 | EIF5A | -0.70371 | 8.213411 | -1.06873 | 0.290491 | 0.932648 | -5.01935 |
| A0A0C4DH | #N/D | 1.023087 | 10.49513 | 1.067646 | 0.290668 | 0.932648 | -4.98143 |
| P62942 | FKBP1A | 1.038813 | 10.83055 | 1.065538 | 0.291253 | 0.932648 | -5.02259 |
| Q99536 | VAT1 | 0.624165 | 11.20367 | 1.062188 | 0.292757 | 0.932648 | -5.02556 |
| O60641 | SNAP91 | -0.59107 | 12.93002 | -1.06136 | 0.293128 | 0.932648 | -5.02629 |
| Q13491-4 | #N/D | 0.721411 | 9.101016 | 1.061413 | 0.293278 | 0.932648 | -4.98695 |
| Q9Y2J2 | EPB41L3 | 0.797274 | 8.157606 | 1.062082 | 0.294071 | 0.932648 | -4.9522 |
| P10768 | ESD | -0.76681 | 10.75161 | -1.05795 | 0.294668 | 0.932648 | -5.02929 |
| P61353 | RPL27 | -0.5658 | 9.636353 | -1.05498 | 0.296014 | 0.932648 | -5.03191 |
| O00264 | PGRMC1 | 0.829457 | 9.27944 | 1.055403 | 0.296273 | 0.932648 | -4.99196 |

| | | | | | | | |
|---|---|---|---|---|---|---|---|
| A0A0C4DH | #N/D | 0.88487 | 14.43238 | 1.053297 | 0.296946 | 0.932648 | -5.03327 |
| P21283 | ATP6V1C1 | -0.987 | 9.48756 | -1.04298 | 0.301657 | 0.940651 | -5.00276 |
| Q96KP4 | CNDP2 | -0.51965 | 11.48894 | -1.04261 | 0.301657 | 0.940651 | -5.0427 |
| Q6UWR7 | ENPP6 | -0.56205 | 11.19948 | -1.03534 | 0.305007 | 0.947699 | -5.04899 |
| P12268 | IMPDH2 | -0.81499 | 9.058515 | -1.02621 | 0.309895 | 0.957369 | -5.0166 |
| P01780 | #N/D | 0.862633 | 11.11587 | 1.021959 | 0.311241 | 0.957369 | -5.06046 |
| P14550 | AKR1A1 | 0.882106 | 7.585267 | 1.014235 | 0.315964 | 0.957369 | -4.89215 |
| A5YM72 | CARNS1 | 0.832296 | 9.654885 | 1.01259 | 0.316395 | 0.957369 | -5.06788 |
| Q99439 | CNN2 | 0.691243 | 9.43566 | 1.006623 | 0.318571 | 0.957369 | -5.03321 |
| P62269 | RPS18 | -0.4622 | 10.80738 | -1.00103 | 0.321244 | 0.957369 | -5.07805 |
| P29622 | SERPINA4 | -0.75748 | 8.812058 | -1.00046 | 0.322629 | 0.957369 | -5.0375 |
| P05090 | APOD | 0.558841 | 14.6869 | 0.992442 | 0.325296 | 0.957369 | -5.08524 |
| O14818 | PSMA7 | -0.59847 | 8.736898 | -0.98785 | 0.3276 | 0.957369 | -5.08898 |
| Q8N163 | CCAR2 | -0.7455 | 8.405871 | -0.98456 | 0.329362 | 0.957369 | -5.05104 |
| P62701 | RPS4X | 0.434472 | 10.53558 | 0.98208 | 0.330329 | 0.957369 | -5.09378 |
| P0C0L5 | C4B | -0.56648 | 12.58969 | -0.97835 | 0.33223 | 0.957369 | -5.09677 |
| Q00765 | REEP5 | -0.70198 | 8.66685 | -0.978 | 0.333039 | 0.957369 | -5.05597 |
| P09417 | QDPR | 0.709859 | 11.79198 | 0.973598 | 0.334487 | 0.957369 | -5.1007 |
| P13797 | PLS3 | -0.62329 | 9.880516 | -0.97095 | 0.33579 | 0.957369 | -5.10285 |
| P0DOX2 | #N/D | 0.388292 | 13.86052 | 0.968694 | 0.336907 | 0.957369 | -5.10468 |
| Q14203 | DCTN1 | -1.65712 | 7.611697 | -0.97577 | 0.337136 | 0.957369 | -4.7043 |
| A0A0B4J1U | #N/D | 0.636149 | 9.305291 | 0.967549 | 0.338179 | 0.957369 | -5.10507 |
| P23284 | PPIB | -0.73919 | 11.31656 | -0.96438 | 0.339284 | 0.957369 | -5.10798 |
| P09874 | PARP1 | 0.432394 | 12.84743 | 0.96253 | 0.339965 | 0.957369 | -5.10965 |
| P40429 | RPL13A | 0.601419 | 11.00395 | 0.961557 | 0.340449 | 0.957369 | -5.11043 |
| P09012 | SNRPA | -0.87445 | 9.436966 | -0.95837 | 0.343066 | 0.957369 | -5.07127 |
| O75636 | FCN3 | 0.495019 | 11.6985 | 0.956031 | 0.343209 | 0.957369 | -5.11485 |
| P08195 | SLC3A2 | 0.729505 | 10.25962 | 0.955887 | 0.343357 | 0.957369 | -5.11491 |
| O43143 | DHX15 | 0.808151 | 7.908015 | 0.952958 | 0.346276 | 0.957369 | -4.99066 |
| P29966 | MARCKS | 0.759432 | 12.49708 | 0.947166 | 0.347666 | 0.957369 | -5.1219 |
| P07358 | C8B | -0.62802 | 13.46551 | -0.94645 | 0.348026 | 0.957369 | -5.12247 |

| | | | | | | | |
|---|---|---|---|---|---|---|---|
| P00367 | GLUD1 | -0.50037 | 13.41429 | -0.94534 | 0.348591 | 0.957369 | -5.12335 |
| A0A075B6I | #N/D | 0.524114 | 14.94383 | 0.942168 | 0.350196 | 0.957369 | -5.12585 |
| P62753 | RPS6 | 0.700034 | 9.50014 | 0.940553 | 0.351016 | 0.957369 | -5.12712 |
| P27824 | CANX | -0.73383 | 11.05687 | -0.93932 | 0.351643 | 0.957369 | -5.12809 |
| P28072 | PSMB6 | -0.62578 | 9.15565 | -0.93562 | 0.353527 | 0.957369 | -5.13099 |
| P63244 | RACK1 | -0.61968 | 12.16177 | -0.93536 | 0.353659 | 0.957369 | -5.13119 |
| P02774-3 | #N/D | -0.73513 | 14.29976 | -0.93436 | 0.354171 | 0.957369 | -5.13197 |
| P30044 | PRDX5 | -1.09839 | 11.42963 | -0.93214 | 0.35531 | 0.957369 | -5.13371 |
| P51649 | ALDH5A1 | -0.42779 | 11.25326 | -0.93115 | 0.355816 | 0.957369 | -5.13448 |
| P12956 | XRCC6 | 0.669926 | 12.11524 | 0.926833 | 0.358032 | 0.957369 | -5.13783 |
| Q16695 | H3-4 | 0.608886 | 10.26812 | 0.923712 | 0.359641 | 0.957369 | -5.14025 |
| P38159 | RBMX | 0.917219 | 13.33236 | 0.918276 | 0.362454 | 0.957369 | -5.14444 |
| P63000 | RAC1 | -0.62309 | 11.00234 | -0.91702 | 0.363106 | 0.957369 | -5.1454 |
| P07384 | CAPN1 | -0.68239 | 8.309043 | -0.911 | 0.366386 | 0.957369 | -5.14988 |
| Q14152 | EIF3A | -0.7053 | 8.131085 | -0.91303 | 0.366779 | 0.957369 | -5.10553 |
| P40939 | HADHA | -0.83547 | 9.753015 | -0.90522 | 0.369342 | 0.957369 | -5.15434 |
| P21926 | CD9 | 0.712937 | 10.94564 | 0.90399 | 0.369914 | 0.957369 | -5.15533 |
| P04040 | CAT | -0.44122 | 8.854698 | -0.90486 | 0.370313 | 0.957369 | -5.07614 |
| P07741 | APRT | -1.08414 | 8.592073 | -0.90167 | 0.371696 | 0.957369 | -5.07869 |
| Q01082 | SPTBN1 | -0.35163 | 11.78071 | -0.89502 | 0.374645 | 0.957369 | -5.16208 |
| P05154 | SERPINA5 | -0.51704 | 11.45435 | -0.89361 | 0.375397 | 0.957369 | -5.16314 |
| P27797 | CALR | 0.479549 | 10.33747 | 0.888075 | 0.378339 | 0.957369 | -5.16727 |
| Q16143 | SNCB | 0.534998 | 11.41947 | 0.884545 | 0.380224 | 0.957369 | -5.16989 |
| Q00325 | SLC25A3 | 0.780231 | 11.25621 | 0.883694 | 0.380679 | 0.957369 | -5.17052 |
| O76021 | RSL1D1 | -0.71568 | 9.23965 | -0.88179 | 0.381915 | 0.957369 | -5.1296 |
| P00742 | F10 | -0.73689 | 12.22352 | -0.87832 | 0.383631 | 0.957369 | -5.17443 |
| P02743 | APCS | 1.227045 | 14.22881 | 0.873168 | 0.38634 | 0.957369 | -5.17827 |
| Q14103 | HNRNPD | -0.90355 | 10.83965 | -0.87262 | 0.386635 | 0.957369 | -5.17867 |
| Q96GD0 | PDXP | 0.585796 | 10.49093 | 0.868173 | 0.389044 | 0.957369 | -5.18191 |
| P80723 | BASP1 | 0.774712 | 10.51359 | 0.867822 | 0.389234 | 0.957369 | -5.18217 |
| P25786 | PSMA1 | -0.83328 | 8.552348 | -0.86787 | 0.390026 | 0.957369 | -5.08074 |

| | | | | | | | |
|---|---|---|---|---|---|---|---|
| P49753 | ACOT2 | -0.59071 | 10.06568 | -0.8603 | 0.393333 | 0.957369 | -5.18762 |
| Q14195-2 | #N/D | -0.44105 | 12.44951 | -0.85875 | 0.394179 | 0.957369 | -5.18874 |
| P69905 | HBA1 | -0.43229 | 14.01226 | -0.85564 | 0.39588 | 0.957369 | -5.19097 |
| Q9UMS4 | PRPF19 | -0.70736 | 9.289913 | -0.8544 | 0.396564 | 0.957369 | -5.19186 |
| A0A0C4DH | #N/D | 0.754197 | 14.37611 | 0.852598 | 0.397552 | 0.957369 | -5.19315 |
| A0A0C4DH | #N/D | 0.925382 | 10.33687 | 0.849474 | 0.39934 | 0.957369 | -5.19532 |
| Q08722 | CD47 | 0.52155 | 9.746025 | 0.848463 | 0.39983 | 0.957369 | -5.1961 |
| O43426 | SYNJ1 | -1.77354 | 13.74991 | -0.84754 | 0.400399 | 0.957369 | -5.19671 |
| A0A0B4J1X | #N/D | -1.06743 | 10.0226 | -0.84572 | 0.401415 | 0.957369 | -5.19799 |
| Q96C19 | EFHD2 | 0.785124 | 8.816843 | 0.845912 | 0.402037 | 0.957369 | -5.15457 |
| P52272 | HNRNPM | -0.47732 | 12.38158 | -0.84185 | 0.403491 | 0.957369 | -5.20079 |
| P60953 | CDC42 | 0.708328 | 9.514288 | 0.841631 | 0.403892 | 0.957369 | -5.15797 |
| P50502 | ST13 | 0.687145 | 12.29466 | 0.836015 | 0.406941 | 0.957369 | -5.20472 |
| Q96PD5 | PGLYRP2 | -0.46547 | 13.17715 | -0.83222 | 0.408859 | 0.957369 | -5.20755 |
| P12004 | PCNA | 0.413821 | 11.20119 | 0.831215 | 0.409419 | 0.957369 | -5.20825 |
| P02647 | APOA1 | 0.599621 | 15.5712 | 0.82959 | 0.41033 | 0.957369 | -5.20938 |
| Q15366 | PCBP2 | 0.802998 | 12.64187 | 0.829579 | 0.410336 | 0.957369 | -5.20939 |
| Q9H299 | SH3BGRL3 | 0.581731 | 8.301393 | 0.829731 | 0.410938 | 0.957369 | -5.20869 |
| Q12906-7 | #N/D | -0.54138 | 10.81171 | -0.82601 | 0.412339 | 0.957369 | -5.21187 |
| P37108 | SRP14 | 0.786263 | 8.187199 | 0.828789 | 0.413105 | 0.957369 | -5.07599 |
| P27635 | RPL10 | -0.46944 | 8.546315 | -0.82419 | 0.41343 | 0.957369 | -5.17009 |
| P0DOX8 | #N/D | -1.00648 | 13.34101 | -0.82413 | 0.413463 | 0.957369 | -5.17013 |
| P14618-3 | #N/D | 0.538276 | 9.955675 | 0.812979 | 0.419773 | 0.957369 | -5.22078 |
| P14868-2 | #N/D | 0.505749 | 8.099459 | 0.81191 | 0.420988 | 0.957369 | -5.17784 |
| P05388 | RPLP0 | 0.477481 | 11.63871 | 0.81001 | 0.421399 | 0.957369 | -5.22286 |
| P11586 | MTHFD1 | -1.57163 | 11.82919 | -0.80768 | 0.422782 | 0.957369 | -5.2244 |
| Q16798 | ME3 | 0.472726 | 8.140939 | 0.807705 | 0.423055 | 0.957369 | -5.18092 |
| P35858 | IGFALS | 0.554922 | 11.34869 | 0.805458 | 0.423998 | 0.957369 | -5.22595 |
| Q16352 | INA | 0.353926 | 13.06932 | 0.803606 | 0.425058 | 0.957369 | -5.2272 |
| Q7KZF4 | SND1 | -0.55682 | 9.615375 | -0.80333 | 0.425415 | 0.957369 | -5.22722 |
| A0A0A0MT | #N/D | -1.01043 | 9.911469 | -0.80349 | 0.425875 | 0.957369 | -5.1459 |

| | | | | | | | |
|---|---|---|---|---|---|---|---|
| Q99447 | PCYT2 | -0.72734 | 9.931576 | -0.797 | 0.428855 | 0.957369 | -5.23165 |
| P62263 | RPS14 | 0.625792 | 12.25025 | 0.796743 | 0.429001 | 0.957369 | -5.23182 |
| O14594 | NCAN | -0.45483 | 9.186149 | -0.7968 | 0.429303 | 0.957369 | -5.23148 |
| P05387 | RPLP2 | 0.486336 | 9.837368 | 0.796205 | 0.429373 | 0.957369 | -5.23212 |
| P14618-2 | #N/D | 0.655849 | 11.99726 | 0.793495 | 0.430874 | 0.957369 | -5.23399 |
| P60028 | #N/D | 0.576441 | 9.154595 | 0.791815 | 0.432037 | 0.957369 | -5.19151 |
| Q9UBC3 | DNMT3B | 0.733945 | 11.72397 | 0.790653 | 0.432517 | 0.957369 | -5.23588 |
| P36578 | RPL4 | -0.31406 | 12.0117 | -0.79021 | 0.432771 | 0.957369 | -5.23617 |
| O75368 | SH3BGRL | -0.73711 | 9.591212 | -0.78568 | 0.435528 | 0.957369 | -5.23906 |
| Q96FW1 | OTUB1 | 0.60399 | 7.493366 | 0.784688 | 0.437449 | 0.957369 | -5.06077 |
| P33993 | MCM7 | -0.50472 | 8.945521 | -0.78204 | 0.437582 | 0.957369 | -5.24152 |
| P68371 | TUBB4B | -0.78885 | 11.99643 | -0.78142 | 0.437882 | 0.957369 | -5.24198 |
| P00747 | PLG | 0.489252 | 12.64273 | 0.7804 | 0.438477 | 0.957369 | -5.24265 |
| P04433 | #N/D | 0.683757 | 12.95042 | 0.779393 | 0.439064 | 0.957369 | -5.24331 |
| P18621 | RPL17 | 0.804147 | 11.78909 | 0.778607 | 0.439523 | 0.957369 | -5.24382 |
| P04259 | KRT6B | 0.592886 | 11.57937 | 0.778125 | 0.439866 | 0.957369 | -5.24409 |
| Q15233 | NONO | 0.512262 | 9.777373 | 0.775149 | 0.441547 | 0.957369 | -5.24608 |
| P50454 | SERPINH1 | -0.48986 | 9.103501 | -0.7735 | 0.442573 | 0.957369 | -5.2471 |
| Q93050 | ATP6V0A1 | -0.37954 | 10.19313 | -0.77287 | 0.442881 | 0.957369 | -5.24756 |
| P19652 | ORM2 | -0.44292 | 9.423031 | -0.77297 | 0.443152 | 0.957369 | -5.20354 |
| P23471 | PTPRZ1 | 1.053949 | 10.95353 | 0.772483 | 0.443233 | 0.957369 | -5.24771 |
| P02671 | FGA | -0.37859 | 12.54848 | -0.77188 | 0.443465 | 0.957369 | -5.24821 |
| O75781 | PALM | 0.442339 | 8.620566 | 0.77073 | 0.444263 | 0.957369 | -5.24885 |
| P50453 | SERPINB9 | -0.75697 | 9.530376 | -0.76795 | 0.446169 | 0.957369 | -5.16874 |
| Q92954 | PRG4 | 0.659145 | 10.51058 | 0.766897 | 0.446398 | 0.957369 | -5.25143 |
| Q9Y2J8 | PADI2 | -0.68595 | 10.26827 | -0.76544 | 0.447255 | 0.957369 | -5.25237 |
| P09972 | ALDOC | 0.402502 | 13.29965 | 0.764346 | 0.447904 | 0.957369 | -5.25308 |
| P26583 | HMGB2 | -0.56547 | 9.427067 | -0.76434 | 0.44791 | 0.957369 | -5.25308 |
| P12277 | CKB | 0.53713 | 13.54112 | 0.763768 | 0.448245 | 0.957369 | -5.25345 |
| P46781 | RPS9 | 0.46079 | 10.5113 | 0.760506 | 0.450176 | 0.957369 | -5.25554 |
| Q9UJU6 | DBNL | 0.53881 | 10.24301 | 0.760605 | 0.450177 | 0.957369 | -5.25542 |

| | | | | | | | |
|---|---|---|---|---|---|---|---|
| O76054 | SEC14L2 | -0.53402 | 8.396433 | -0.76051 | 0.451061 | 0.957369 | -5.21092 |
| Q05193 | DNM1 | -0.4159 | 10.1149 | -0.75766 | 0.451867 | 0.957369 | -5.25736 |
| O95782 | AP2A1 | 0.568605 | 8.500907 | 0.756577 | 0.45339 | 0.957369 | -5.17533 |
| P52306-6 | #N/D | -0.5703 | 11.71115 | -0.75507 | 0.453404 | 0.957369 | -5.259 |
| Q04917 | YWHAH | -0.55334 | 11.45903 | -0.75487 | 0.453524 | 0.957369 | -5.25913 |
| P13010 | XRCC5 | -0.40701 | 12.2267 | -0.75474 | 0.453602 | 0.957369 | -5.25922 |
| P17655 | CAPN2 | 0.550258 | 9.117694 | 0.755213 | 0.453857 | 0.957369 | -5.21454 |
| Q9NR46 | SH3GLB2 | -0.5063 | 8.407993 | -0.74591 | 0.459053 | 0.957369 | -5.2206 |
| P23246 | SFPQ | 0.446037 | 12.17656 | 0.742523 | 0.460907 | 0.957369 | -5.26691 |
| Q86V81 | ALYREF | -0.6783 | 8.628483 | -0.74281 | 0.461044 | 0.957369 | -5.26645 |
| Q13153 | PAK1 | 0.699799 | 7.407468 | 0.744059 | 0.46237 | 0.957369 | -5.04621 |
| P46783 | RPS10 | -0.61284 | 10.88473 | -0.73595 | 0.464868 | 0.957369 | -5.271 |
| P11142 | HSPA8 | -0.37236 | 13.19735 | -0.73297 | 0.466663 | 0.957369 | -5.27284 |
| Q9Y4L1 | HYOU1 | -0.43792 | 8.672707 | -0.73195 | 0.467342 | 0.957369 | -5.27343 |
| Q7Z3B1 | NEGR1 | 0.56957 | 7.928181 | 0.73233 | 0.467995 | 0.957369 | -5.22822 |
| O75363-2 | #N/D | 0.567487 | 8.935009 | 0.730699 | 0.468098 | 0.957369 | -5.22997 |
| P04179 | SOD2 | 0.449062 | 10.77234 | 0.728071 | 0.469636 | 0.957369 | -5.27586 |
| P09429 | HMGB1 | -0.35347 | 11.33429 | -0.72736 | 0.470066 | 0.957369 | -5.27629 |
| P25789 | PSMA4 | 0.462952 | 7.883878 | 0.727815 | 0.470232 | 0.957369 | -5.23137 |
| P36957 | DLST | -0.60718 | 8.614293 | -0.72238 | 0.473845 | 0.957369 | -5.17288 |
| P34897 | SHMT2 | -0.48423 | 10.26867 | -0.72111 | 0.473876 | 0.957369 | -5.28011 |
| P61970 | NUTF2 | 0.533331 | 8.594163 | 0.720736 | 0.474157 | 0.957369 | -5.28028 |
| P06744 | GPI | 0.424996 | 11.87388 | 0.719638 | 0.474772 | 0.957369 | -5.281 |
| P47914 | RPL29 | -0.59871 | 8.941895 | -0.7197 | 0.474967 | 0.957369 | -5.19788 |
| P08603 | CFH | -0.41051 | 14.2247 | -0.71846 | 0.475492 | 0.957369 | -5.28171 |
| Q9P2D7-8 | #N/D | -0.61731 | 8.669631 | -0.71819 | 0.475659 | 0.957369 | -5.28188 |
| P05787 | KRT8 | 0.378025 | 11.88015 | 0.713882 | 0.478297 | 0.957369 | -5.28447 |
| P35268 | RPL22 | -0.40486 | 11.96432 | -0.71287 | 0.478916 | 0.957369 | -5.28508 |
| P07225 | PROS1 | 0.3066 | 13.16055 | 0.711516 | 0.479749 | 0.957369 | -5.2859 |
| Q15084 | PDIA6 | -0.42047 | 12.47948 | -0.71109 | 0.480009 | 0.957369 | -5.28615 |
| P02649 | APOE | 0.500097 | 16.1627 | 0.709877 | 0.480757 | 0.957369 | -5.28688 |

| | | | | | | | |
|---|---|---|---|---|---|---|---|
| P01764 | #N/D | -0.53447 | 9.51328 | -0.70882 | 0.481765 | 0.957369 | -5.2427 |
| O75891 | ALDH1L1 | -0.53143 | 12.21429 | -0.70094 | 0.486275 | 0.957369 | -5.29219 |
| Q9P2U7 | SLC17A7 | 0.651668 | 10.35073 | 0.697774 | 0.488345 | 0.957369 | -5.29396 |
| O95394 | PGM3 | -0.52227 | 8.458672 | -0.6968 | 0.488896 | 0.957369 | -5.29458 |
| P30041 | PRDX6 | -0.65777 | 12.92684 | -0.69477 | 0.490102 | 0.957369 | -5.29582 |
| P31948 | STIP1 | 0.326864 | 10.97486 | 0.694626 | 0.490192 | 0.957369 | -5.2959 |
| P61978 | HNRNPK | -0.54296 | 11.47478 | -0.69219 | 0.491709 | 0.957369 | -5.29733 |
| P07108 | DBI | 0.45739 | 7.299068 | 0.693379 | 0.491765 | 0.957369 | -5.21245 |
| P04114 | APOB | 0.591647 | 15.2199 | 0.691235 | 0.492303 | 0.957369 | -5.29788 |
| P29762 | CRABP1 | 0.715281 | 11.46855 | 0.690591 | 0.492705 | 0.957369 | -5.29826 |
| P13489 | RNH1 | -0.60556 | 9.869775 | -0.6882 | 0.494196 | 0.957369 | -5.29965 |
| P09936 | UCHL1 | 0.401482 | 12.48925 | 0.687533 | 0.494614 | 0.957369 | -5.30003 |
| P48637 | GSS | 0.651574 | 8.001266 | 0.688821 | 0.494971 | 0.957369 | -5.15991 |
| P00505 | GOT2 | 0.36358 | 12.49377 | 0.68437 | 0.496594 | 0.957369 | -5.30186 |
| P40926 | MDH2 | -0.34416 | 13.17949 | -0.6827 | 0.497641 | 0.957369 | -5.30283 |
| O00499 | BIN1 | 0.556681 | 8.599564 | 0.677936 | 0.501675 | 0.957369 | -5.10416 |
| P07451 | CA3 | -0.58919 | 9.536455 | -0.67572 | 0.502082 | 0.957369 | -5.30678 |
| A2NJV5 | #N/D | 0.939899 | 8.920818 | 0.675293 | 0.502703 | 0.957369 | -5.26183 |
| P55056 | APOC4 | 0.561573 | 9.255911 | 0.675987 | 0.502718 | 0.957369 | -5.19871 |
| P56747 | CLDN6 | 0.477999 | 8.759588 | 0.67451 | 0.503333 | 0.957369 | -5.26215 |
| P78347 | GTF2I | -0.41107 | 9.347322 | -0.67364 | 0.50339 | 0.957369 | -5.30796 |
| P69891 | HBG1 | -0.53701 | 10.30411 | -0.67335 | 0.503575 | 0.957369 | -5.30813 |
| P35908 | KRT2 | -0.72213 | 14.98502 | -0.6718 | 0.504503 | 0.957369 | -5.30905 |
| P01859 | #N/D | -0.53609 | 13.5427 | -0.67121 | 0.504878 | 0.957369 | -5.30939 |
| P0DOX5 | #N/D | 0.356948 | 14.50645 | 0.669936 | 0.505681 | 0.957369 | -5.31011 |
| E9PAV3 | #N/D | 0.574099 | 12.14083 | 0.669721 | 0.505868 | 0.957369 | -5.31018 |
| P30405 | PPIF | -0.54982 | 7.967455 | -0.6701 | 0.507143 | 0.957369 | -5.0522 |
| P55209 | NAP1L1 | 0.634287 | 12.57393 | 0.662483 | 0.510458 | 0.957369 | -5.31425 |
| P26641 | EEF1G | -0.37173 | 10.13888 | -0.65768 | 0.513514 | 0.957369 | -5.31692 |
| P49368 | CCT3 | -0.68526 | 10.62952 | -0.65694 | 0.51394 | 0.957369 | -5.31738 |
| P22061 | PCMT1 | -0.37484 | 10.41461 | -0.65235 | 0.516872 | 0.957369 | -5.31992 |

| | | | | | | | |
|---|---|---|---|---|---|---|---|
| Q15121 | PEA15 | 0.315755 | 10.46576 | 0.64985 | 0.518474 | 0.957369 | -5.32129 |
| Q01105 | SET | -0.37041 | 12.74759 | -0.64856 | 0.519304 | 0.957369 | -5.322 |
| P08237-3 | #N/D | -0.35141 | 9.903447 | -0.64838 | 0.519567 | 0.957369 | -5.27677 |
| P30048 | PRDX3 | -0.51367 | 9.178145 | -0.64798 | 0.520659 | 0.957369 | -5.27626 |
| P00915 | CA1 | -0.44154 | 10.62668 | -0.64626 | 0.520777 | 0.957369 | -5.32325 |
| P55084 | HADHB | 0.411965 | 8.238028 | 0.646768 | 0.520968 | 0.957369 | -5.27732 |
| P14415 | ATP1B2 | 0.422642 | 8.426782 | 0.645969 | 0.521229 | 0.957369 | -5.32318 |
| P06703 | S100A6 | -0.73409 | 9.228704 | -0.63904 | 0.525428 | 0.957369 | -5.32717 |
| Q9NSD9 | FARSB | -0.4322 | 9.390867 | -0.63922 | 0.525691 | 0.957369 | -5.32674 |
| P02749 | APOH | 0.329961 | 12.64195 | 0.637454 | 0.526454 | 0.957369 | -5.32803 |
| P62273 | RPS29 | -0.46349 | 8.012306 | -0.6376 | 0.526803 | 0.957369 | -5.28226 |
| O94760 | DDAH1 | 0.42675 | 9.840467 | 0.636524 | 0.527104 | 0.957369 | -5.32848 |
| P25311 | AZGP1 | -0.32703 | 12.1082 | -0.63374 | 0.528861 | 0.957369 | -5.33002 |
| P20742 | #N/D | -0.44768 | 13.66452 | -0.63276 | 0.529496 | 0.957369 | -5.33055 |
| P13798 | APEH | 0.451487 | 11.77021 | 0.63226 | 0.529817 | 0.957369 | -5.33081 |
| P12270 | TPR | -0.63136 | 9.31589 | -0.63119 | 0.530714 | 0.957369 | -5.22268 |
| P02750 | LRG1 | 0.527495 | 10.4896 | 0.628204 | 0.532451 | 0.957369 | -5.33297 |
| P14866 | HNRNPL | 0.38662 | 12.72325 | 0.627354 | 0.533004 | 0.957369 | -5.33342 |
| O94811 | TPPP | -0.29845 | 11.43773 | -0.6252 | 0.534405 | 0.957369 | -5.33456 |
| P04080 | CSTB | 0.504375 | 11.10002 | 0.624447 | 0.534897 | 0.957369 | -5.33496 |
| P24539 | ATP5PB | 0.579074 | 9.276826 | 0.623077 | 0.536219 | 0.957369 | -5.28985 |
| P62888 | RPL30 | 0.427943 | 9.168351 | 0.621352 | 0.536962 | 0.957369 | -5.33654 |
| P61106 | RAB14 | -0.37305 | 9.911827 | -0.61928 | 0.538319 | 0.957369 | -5.33763 |
| P50990 | CCT8 | -0.29674 | 13.71428 | -0.61919 | 0.538331 | 0.957369 | -5.33772 |
| Q9HC38 | GLOD4 | -0.45509 | 9.301142 | -0.61885 | 0.538548 | 0.957369 | -5.33789 |
| P21579 | SYT1 | -0.43048 | 9.733547 | -0.61806 | 0.539066 | 0.957369 | -5.33831 |
| Q9Y696 | CLIC4 | 0.513392 | 8.300412 | 0.617374 | 0.540005 | 0.957369 | -5.33824 |
| Q86YZ3 | HRNR | -0.59983 | 10.14946 | -0.61591 | 0.540725 | 0.957369 | -5.33921 |
| Q13449 | LSAMP | -0.34791 | 9.843599 | -0.61366 | 0.541953 | 0.957369 | -5.3406 |
| O43776 | NARS1 | -0.37274 | 10.11421 | -0.61043 | 0.544075 | 0.957369 | -5.34227 |
| P07954 | FH | -0.49284 | 11.84092 | -0.61027 | 0.544226 | 0.957369 | -5.34231 |

| | | | | | | | |
|---|---|---|---|---|---|---|---|
| P02790 | HPX | 0.483526 | 14.96138 | 0.609298 | 0.544817 | 0.957369 | -5.34285 |
| Q16623-3 | #N/D | 0.310209 | 9.890803 | 0.605899 | 0.547055 | 0.957369 | -5.3446 |
| Q9NPH9 | IL26 | 0.584983 | 9.277746 | 0.606201 | 0.547101 | 0.957369 | -5.34423 |
| P08519 | #N/D | 0.502374 | 15.54961 | 0.602935 | 0.549012 | 0.957369 | -5.34611 |
| Q15392 | DHCR24 | -0.52907 | 8.63106 | -0.6008 | 0.550663 | 0.957369 | -5.30129 |
| P01042-2 | #N/D | 0.613827 | 11.83736 | 0.599736 | 0.551218 | 0.957369 | -5.30195 |
| P03952 | KLKB1 | 0.409676 | 13.33419 | 0.59937 | 0.551369 | 0.957369 | -5.34792 |
| P02689 | PMP2 | 0.441006 | 10.41906 | 0.598723 | 0.551798 | 0.957369 | -5.34825 |
| Q8IXJ6 | SIRT2 | 0.622593 | 11.31293 | 0.597648 | 0.552648 | 0.957369 | -5.30295 |
| Q8NC51 | SERBP1 | 0.70715 | 11.27405 | 0.596064 | 0.55356 | 0.957369 | -5.34959 |
| Q06033 | ITIH3 | -0.70928 | 13.55378 | -0.59386 | 0.555025 | 0.957369 | -5.3507 |
| Q9Y6I3 | EPN1 | -0.3916 | 8.193578 | -0.5927 | 0.556456 | 0.957369 | -5.26507 |
| P62318 | SNRPD3 | -0.36897 | 9.893724 | -0.58991 | 0.557651 | 0.957369 | -5.35268 |
| Q9Y5K8 | ATP6V1D | 0.456315 | 7.792041 | 0.588943 | 0.559098 | 0.957369 | -5.26676 |
| Q99798 | ACO2 | 0.314961 | 13.35696 | 0.586392 | 0.559993 | 0.957369 | -5.35442 |
| P00568 | AK1 | 0.390852 | 11.14475 | 0.585726 | 0.560438 | 0.957369 | -5.35475 |
| P0DP25 | CALM1 | 0.795226 | 11.85239 | 0.583036 | 0.562235 | 0.957369 | -5.35608 |
| O75947 | ATP5PD | -0.46234 | 8.722357 | -0.58092 | 0.563734 | 0.957369 | -5.35705 |
| P78559 | MAP1A | 0.406814 | 10.07764 | 0.579458 | 0.564762 | 0.957369 | -5.31182 |
| P60842 | EIF4A1 | 0.527584 | 11.09096 | 0.576314 | 0.566736 | 0.957369 | -5.35938 |
| A0A075B6. | #N/D | -0.55479 | 10.90519 | -0.57428 | 0.568233 | 0.957369 | -5.36025 |
| Q13561 | DCTN2 | 0.359495 | 8.671465 | 0.573585 | 0.568702 | 0.957369 | -5.36059 |
| P00739 | HPR | -0.43539 | 13.20664 | -0.57316 | 0.568854 | 0.957369 | -5.36091 |
| P12036 | NEFH | -0.21349 | 12.50765 | -0.56995 | 0.571016 | 0.957369 | -5.36246 |
| P31946 | YWHAB | 0.37064 | 9.674481 | 0.56947 | 0.57147 | 0.957369 | -5.36258 |
| P11217 | PYGM | -0.30265 | 10.22972 | -0.56788 | 0.572411 | 0.957369 | -5.36346 |
| Q9NQC3 | RTN4 | 0.335201 | 10.95603 | 0.567063 | 0.572962 | 0.957369 | -5.36385 |
| Q08380 | LGALS3BP | -0.34198 | 14.57471 | -0.56576 | 0.573838 | 0.957369 | -5.36447 |
| P52292 | KPNA2 | -0.36542 | 7.997937 | -0.56564 | 0.574421 | 0.957369 | -5.3641 |
| P02748 | C9 | -0.45797 | 13.54131 | -0.56147 | 0.576741 | 0.959397 | -5.36652 |
| P13861 | PRKAR2A | 0.446141 | 8.585476 | 0.55726 | 0.579768 | 0.95942 | -5.36836 |

| | | | | | | | |
|---|---|---|---|---|---|---|---|
| P00751 | CFB | -0.33106 | 13.86732 | -0.55573 | 0.580631 | 0.95942 | -5.36923 |
| A0A0C4DH | #N/D | 0.306284 | 12.25946 | 0.55509 | 0.581067 | 0.95942 | -5.36953 |
| P55786 | NPEPPS | -0.3412 | 9.711469 | -0.55431 | 0.5816 | 0.95942 | -5.3699 |
| A0MZ66 | SHTN1 | 0.468239 | 8.930068 | 0.552428 | 0.583197 | 0.95942 | -5.32435 |
| P43652 | AFM | -0.27116 | 12.25694 | -0.5517 | 0.583372 | 0.95942 | -5.37112 |
| P28161 | GSTM2 | -0.3871 | 8.502551 | -0.55055 | 0.584703 | 0.959795 | -5.28475 |
| P0DJI8 | SAA1 | -0.42815 | 12.87564 | -0.54795 | 0.585927 | 0.959994 | -5.37287 |
| P13716-2 | #N/D | 0.436548 | 8.80277 | 0.545045 | 0.588079 | 0.961708 | -5.32783 |
| O60282 | KIF5C | -0.44343 | 9.337804 | -0.54199 | 0.59012 | 0.963236 | -5.32924 |
| Q96F07 | CYFIP2 | 0.637482 | 9.351898 | 0.537106 | 0.593511 | 0.965845 | -5.3314 |
| P16949 | STMN1 | 0.392544 | 11.41839 | 0.536239 | 0.593939 | 0.965845 | -5.37824 |
| P30626 | SRI | 0.53092 | 7.800031 | 0.535032 | 0.595794 | 0.967053 | -5.11099 |
| P62851 | RPS25 | 0.381967 | 10.64803 | 0.528365 | 0.599356 | 0.968525 | -5.38178 |
| Q9UQ80 | PA2G4 | 0.356476 | 9.950951 | 0.528293 | 0.599405 | 0.968525 | -5.38182 |
| Q99623 | PHB2 | -0.49191 | 10.71072 | -0.52737 | 0.60004 | 0.968525 | -5.38223 |
| P01615 | #N/D | 0.427137 | 11.88343 | 0.523947 | 0.602756 | 0.969458 | -5.33704 |
| Q92752 | TNR | -0.24329 | 11.57348 | -0.52206 | 0.60371 | 0.969458 | -5.38459 |
| P09471 | GNAO1 | 0.297779 | 11.83785 | 0.521696 | 0.603961 | 0.969458 | -5.38475 |
| P16112 | #N/D | -0.32191 | 8.198785 | -0.51902 | 0.606109 | 0.970026 | -5.33921 |
| P00558 | PGK1 | -0.22478 | 12.86953 | -0.51796 | 0.606545 | 0.970026 | -5.38639 |
| Q71U36 | TUBA1A | -0.49119 | 13.10907 | -0.51765 | 0.608449 | 0.971285 | -5.29815 |
| P01591 | JCHAIN | 0.29943 | 14.17635 | 0.512438 | 0.610381 | 0.971776 | -5.3888 |
| P06730-2 | #N/D | -0.44041 | 8.849688 | -0.51055 | 0.612033 | 0.971776 | -5.3428 |
| Q9BVA1 | TUBB2B | -0.47412 | 10.28254 | -0.50779 | 0.613614 | 0.971776 | -5.39081 |
| P01701 | #N/D | 0.419477 | 10.54093 | 0.50582 | 0.615145 | 0.971776 | -5.39153 |
| P46821 | MAP1B | 0.300155 | 12.35155 | 0.504061 | 0.616217 | 0.971776 | -5.39241 |
| O14967 | CLGN | 0.344462 | 11.39778 | 0.503623 | 0.61681 | 0.971776 | -5.39236 |
| P49913 | CAMP | -0.34283 | 8.94521 | -0.50228 | 0.617954 | 0.971776 | -5.34616 |
| P04075 | ALDOA | 0.238998 | 13.09657 | 0.500227 | 0.618896 | 0.971776 | -5.39404 |
| P05141 | SLC25A5 | 0.404166 | 10.10956 | 0.499406 | 0.619507 | 0.971776 | -5.39436 |
| P61764 | STXBP1 | 0.333278 | 13.40973 | 0.498755 | 0.619926 | 0.971776 | -5.39466 |

| | | | | | | | |
|---|---|---|---|---|---|---|---|
| P02686 | MBP | -0.20523 | 12.52744 | -0.49524 | 0.622386 | 0.973194 | -5.39614 |
| A0A075B6I | #N/D | 0.31987 | 9.618077 | 0.494272 | 0.623068 | 0.973194 | -5.39655 |
| P26038 | MSN | 0.424016 | 8.378736 | 0.492084 | 0.624884 | 0.973477 | -5.28528 |
| Q8WVE0 | EEF1AKMT1 | -0.48707 | 12.59871 | -0.49369 | 0.62594 | 0.973477 | -5.34816 |
| P20916 | MAG | 0.356163 | 9.172305 | 0.489569 | 0.626606 | 0.973477 | -5.3516 |
| O14576 | DYNC1I1 | 0.379247 | 8.850936 | 0.484616 | 0.629894 | 0.975316 | -5.40052 |
| P22234 | PAICS | -0.28605 | 10.08226 | -0.47888 | 0.633908 | 0.975316 | -5.40289 |
| P07237 | P4HB | -0.37728 | 10.09498 | -0.47695 | 0.635378 | 0.975316 | -5.40359 |
| P25788 | PSMA3 | -0.36027 | 9.356898 | -0.47589 | 0.63621 | 0.975316 | -5.31622 |
| P46779 | RPL28 | 0.366167 | 9.65877 | 0.474821 | 0.636815 | 0.975316 | -5.40451 |
| Q9H4G0 | EPB41L1 | 0.308976 | 10.09245 | 0.470712 | 0.639694 | 0.975316 | -5.40618 |
| P02751-8 | #N/D | 0.230222 | 15.47018 | 0.470239 | 0.64003 | 0.975316 | -5.40637 |
| P31146 | CORO1A | 0.384126 | 9.870592 | 0.470238 | 0.640136 | 0.975316 | -5.40629 |
| P09622 | DLD | -0.32506 | 8.688977 | -0.4704 | 0.640368 | 0.975316 | -5.35907 |
| P22792 | CPN2 | -0.38053 | 10.29617 | -0.46612 | 0.642957 | 0.975316 | -5.408 |
| P06312 | #N/D | -0.34176 | 13.06851 | -0.46554 | 0.643372 | 0.975316 | -5.40823 |
| P05783 | KRT18 | 0.416698 | 11.19722 | 0.465555 | 0.643427 | 0.975316 | -5.40817 |
| P38606 | ATP6V1A | 0.328489 | 12.55831 | 0.465449 | 0.643434 | 0.975316 | -5.40827 |
| P52597 | HNRNPF | -0.25171 | 9.211955 | -0.46542 | 0.643485 | 0.975316 | -5.40825 |
| P05155 | SERPING1 | -0.30019 | 12.77219 | -0.46207 | 0.645839 | 0.975668 | -5.4096 |
| P16403 | H1-2 | 0.198055 | 11.57771 | 0.461504 | 0.646244 | 0.975668 | -5.40982 |
| Q99962 | SH3GL2 | 0.232873 | 12.54019 | 0.457086 | 0.649396 | 0.975668 | -5.41154 |
| Q01484 | ANK2 | -0.36556 | 9.391498 | -0.45567 | 0.650439 | 0.975668 | -5.41206 |
| A0A0B4J1Y | #N/D | -0.33958 | 9.589963 | -0.45594 | 0.650474 | 0.975668 | -5.36479 |
| P13647 | KRT5 | 0.329682 | 13.1342 | 0.454824 | 0.651013 | 0.975668 | -5.41241 |
| P01817 | #N/D | -0.27925 | 11.01698 | -0.4518 | 0.653178 | 0.975668 | -5.41358 |
| Q8N573 | OXR1 | 0.371932 | 8.278275 | 0.450916 | 0.654652 | 0.975668 | -5.30048 |
| P62258 | YWHAE | -0.19592 | 12.79759 | -0.44968 | 0.654698 | 0.975668 | -5.41439 |
| P62266 | RPS23 | 0.334899 | 9.096047 | 0.449593 | 0.654931 | 0.975668 | -5.36724 |
| P26599 | PTBP1 | 0.228028 | 10.40181 | 0.446811 | 0.656753 | 0.97671 | -5.41548 |
| P78417 | GSTO1 | 0.342947 | 11.81208 | 0.443171 | 0.659368 | 0.977746 | -5.41685 |

| | | | | | | | |
|---|---|---|---|---|---|---|---|
| Q9UN36 | NDRG2 | 0.322698 | 11.61186 | 0.439769 | 0.661815 | 0.977746 | -5.41812 |
| P46459 | NSF | -0.31867 | 12.65607 | -0.43731 | 0.663585 | 0.977746 | -5.41904 |
| Q9Y617 | PSAT1 | 0.277812 | 11.24982 | 0.433746 | 0.666157 | 0.977746 | -5.42035 |
| P08779 | KRT16 | 0.429341 | 8.523877 | 0.434789 | 0.66643 | 0.977746 | -5.30611 |
| P20339 | RAB5A | 0.252864 | 8.711876 | 0.432778 | 0.666856 | 0.977746 | -5.42071 |
| Q16555 | DPYSL2 | 0.2124 | 14.86964 | 0.432743 | 0.666881 | 0.977746 | -5.42072 |
| P68402 | PAFAH1B2 | -0.45114 | 6.807079 | -0.43653 | 0.66753 | 0.977746 | -5.14399 |
| P11216 | PYGB | -0.22872 | 10.47934 | -0.42882 | 0.669716 | 0.977746 | -5.42216 |
| P78371 | CCT2 | 0.323052 | 10.10086 | 0.428156 | 0.670197 | 0.977746 | -5.4224 |
| Q5IFJ7 | #N/D | 0.522434 | 10.61788 | 0.428013 | 0.670331 | 0.977746 | -5.42243 |
| P63104 | YWHAZ | -0.18133 | 13.41142 | -0.42504 | 0.672455 | 0.977746 | -5.42352 |
| Q96GW7 | BCAN | -0.23405 | 8.242077 | -0.42448 | 0.673059 | 0.977746 | -5.33507 |
| A0A0A0MS | #N/D | -0.29008 | 10.69979 | -0.42288 | 0.67402 | 0.977746 | -5.4243 |
| P28074 | PSMB5 | 0.269157 | 8.249193 | 0.420415 | 0.676165 | 0.977746 | -5.42492 |
| P02763 | ORM1 | 0.309083 | 11.84132 | 0.419038 | 0.676808 | 0.977746 | -5.42567 |
| Q5TFQ8 | SIRPB1 | 0.336497 | 8.396877 | 0.418352 | 0.67796 | 0.977746 | -5.27817 |
| Q15907 | RAB11B | 0.266117 | 9.911453 | 0.416921 | 0.678377 | 0.977746 | -5.4264 |
| Q12905 | ILF2 | -0.3146 | 9.328399 | -0.41399 | 0.680539 | 0.977746 | -5.38011 |
| Q16658 | FSCN1 | 0.250254 | 13.04135 | 0.413139 | 0.681099 | 0.977746 | -5.42776 |
| P17987 | TCP1 | 0.346693 | 10.0471 | 0.412028 | 0.681909 | 0.977746 | -5.42814 |
| P48740 | MASP1 | 0.288452 | 8.81856 | 0.411744 | 0.682175 | 0.977746 | -5.4282 |
| P49321 | NASP | 0.232027 | 13.01531 | 0.406644 | 0.685836 | 0.97947 | -5.43001 |
| Q9UNQ0 | ABCG2 | 0.483657 | 11.29824 | 0.405028 | 0.687138 | 0.97947 | -5.38312 |
| P02760 | AMBP | 0.190318 | 13.73303 | 0.404369 | 0.687498 | 0.97947 | -5.4308 |
| Q13885 | TUBB2A | -0.29161 | 12.3616 | -0.40307 | 0.688451 | 0.97947 | -5.43124 |
| P49773 | HINT1 | -0.2704 | 8.488339 | -0.40175 | 0.6895 | 0.97947 | -5.43163 |
| P05546 | SERPIND1 | 0.25936 | 11.79322 | 0.399409 | 0.691127 | 0.97947 | -5.43249 |
| P20042 | EIF2S2 | 0.38462 | 8.98351 | 0.399268 | 0.691258 | 0.97947 | -5.43252 |
| P08708 | RPS17 | -0.23616 | 7.699005 | -0.39737 | 0.69288 | 0.9798 | -5.433 |
| P09871 | C1S | 0.21975 | 13.55189 | 0.395839 | 0.693744 | 0.9798 | -5.43369 |
| P35520 | CBS | 0.260247 | 8.684517 | 0.39133 | 0.697202 | 0.979886 | -5.4351 |

| | | | | | | | |
|---|---|---|---|---|---|---|---|
| P01619 | #N/D | 0.232706 | 14.00764 | 0.390399 | 0.697855 | 0.979886 | -5.43543 |
| A0A075B6I | #N/D | 0.390364 | 7.939523 | 0.389716 | 0.69908 | 0.979886 | -5.19591 |
| P21281 | ATP6V1B2 | -0.26845 | 13.63954 | -0.38793 | 0.699555 | 0.979886 | -5.43632 |
| Q16653-3 | #N/D | 0.267486 | 9.748252 | 0.386956 | 0.700271 | 0.979886 | -5.43664 |
| O14791 | APOL1 | 0.257779 | 13.07569 | 0.386561 | 0.700562 | 0.979886 | -5.43677 |
| P35527 | KRT9 | 0.256836 | 15.65986 | 0.383581 | 0.702757 | 0.98087 | -5.43775 |
| O76070 | SNCG | 0.202599 | 10.89826 | 0.382306 | 0.703697 | 0.98087 | -5.43816 |
| P07910 | HNRNPC | -0.16937 | 10.97799 | -0.38039 | 0.705112 | 0.98087 | -5.43878 |
| P04264 | KRT1 | -0.24291 | 16.88435 | -0.37859 | 0.706439 | 0.98087 | -5.43936 |
| P02533 | KRT14 | -0.29685 | 11.09842 | -0.37416 | 0.709717 | 0.98087 | -5.44078 |
| P22087 | FBL | -0.22565 | 8.6313 | -0.37286 | 0.710844 | 0.98087 | -5.39349 |
| P48506 | GCLC | 0.267456 | 8.866158 | 0.370413 | 0.712487 | 0.98087 | -5.44196 |
| P05023 | ATP1A1 | 0.252103 | 13.61348 | 0.37008 | 0.712734 | 0.98087 | -5.44207 |
| O95197 | RTN3 | 0.321606 | 8.031271 | 0.370726 | 0.712878 | 0.98087 | -5.35223 |
| P24752 | ACAT1 | -0.25716 | 10.23426 | -0.36966 | 0.713043 | 0.98087 | -5.4422 |
| P18124 | RPL7 | -0.19857 | 10.99238 | -0.36882 | 0.713667 | 0.98087 | -5.44246 |
| A0A087WS | #N/D | -0.33067 | 13.2827 | -0.36427 | 0.717043 | 0.983955 | -5.44388 |
| P52758 | RIDA | -0.22981 | 8.569229 | -0.35862 | 0.721317 | 0.988261 | -5.44556 |
| P35579 | MYH9 | 0.274326 | 12.21283 | 0.355271 | 0.723735 | 0.989202 | -5.44663 |
| P35611 | ADD1 | 0.353774 | 8.364787 | 0.354857 | 0.724744 | 0.989202 | -5.23026 |
| Q02543 | RPL18A | 0.346132 | 12.91705 | 0.346295 | 0.730433 | 0.989202 | -5.4493 |
| O94919 | ENDOD1 | 0.228125 | 9.449366 | 0.344975 | 0.731468 | 0.989202 | -5.44966 |
| P02766 | TTR | -0.30373 | 13.95342 | -0.34372 | 0.73236 | 0.989202 | -5.45006 |
| Q13247 | SRSF6 | -0.23124 | 8.67543 | -0.34368 | 0.732439 | 0.989202 | -5.45004 |
| A0A0B4J1\ | #N/D | 0.337703 | 8.761429 | 0.343088 | 0.73331 | 0.989202 | -5.40218 |
| P34932 | HSPA4 | -0.18521 | 12.19409 | -0.34242 | 0.733333 | 0.989202 | -5.45044 |
| P07437 | TUBB | -0.27056 | 12.82403 | -0.3414 | 0.734091 | 0.989202 | -5.45073 |
| P05160 | F13B | 0.280879 | 11.66103 | 0.340852 | 0.734552 | 0.989202 | -5.45086 |
| Q8WUM4 | PDCD6IP | 0.471938 | 7.545402 | 0.34285 | 0.73477 | 0.989202 | -4.98993 |
| P00403 | COX2 | -0.20481 | 8.386699 | -0.33975 | 0.735648 | 0.989202 | -5.40323 |
| P62249 | RPS16 | 0.158402 | 10.28851 | 0.333238 | 0.740212 | 0.990326 | -5.45307 |

| | | | | | | | |
|---|---|---|---|---|---|---|---|
| P07477 | PRSS1 | 0.485302 | 13.06236 | 0.332843 | 0.740532 | 0.990326 | -5.45317 |
| P02538 | KRT6A | -0.34931 | 11.77848 | -0.33264 | 0.74073 | 0.990326 | -5.45319 |
| P05156 | CFI | 0.22688 | 13.67799 | 0.33214 | 0.741037 | 0.990326 | -5.45338 |
| Q969P0 | IGSF8 | -0.19571 | 10.87703 | -0.32532 | 0.746167 | 0.993607 | -5.45529 |
| P27695 | APEX1 | -0.34444 | 9.067064 | -0.32373 | 0.74777 | 0.993607 | -5.4076 |
| O43390 | HNRNPR | 0.179531 | 8.163912 | 0.320575 | 0.749857 | 0.993607 | -5.45652 |
| P09104 | ENO2 | 0.17225 | 11.61892 | 0.317707 | 0.751901 | 0.993607 | -5.45737 |
| P10643 | C7 | -0.15584 | 12.83675 | -0.31584 | 0.753307 | 0.993607 | -5.45787 |
| P02654 | APOC1 | 0.203391 | 11.84732 | 0.315425 | 0.753645 | 0.993607 | -5.45797 |
| P10909 | CLU | 0.124425 | 13.71219 | 0.314978 | 0.753961 | 0.993607 | -5.4581 |
| P62917 | RPL8 | -0.33584 | 11.02457 | -0.31365 | 0.754983 | 0.993607 | -5.45844 |
| P27169 | PON1 | -0.28411 | 14.83311 | -0.31349 | 0.755106 | 0.993607 | -5.45848 |
| P00441 | SOD1 | -0.21156 | 11.21279 | -0.31208 | 0.756169 | 0.993607 | -5.45886 |
| P62913 | RPL11 | -0.21745 | 12.51404 | -0.31154 | 0.756557 | 0.993607 | -5.45901 |
| P53634 | CTSC | -0.22823 | 8.483897 | -0.31018 | 0.757758 | 0.993607 | -5.41133 |
| A0A0C4DH | #N/D | 0.24815 | 11.05438 | 0.307056 | 0.759952 | 0.993607 | -5.46019 |
| Q16775 | HAGH | 0.264166 | 10.09877 | 0.305865 | 0.760854 | 0.993607 | -5.4605 |
| P0C0L4 | C4A | -0.31388 | 11.8147 | -0.30353 | 0.762814 | 0.993607 | -5.41301 |
| P62424 | RPL7A | -0.12329 | 11.51419 | -0.30314 | 0.762922 | 0.993607 | -5.46121 |
| A0A0C4DH | #N/D | 0.259003 | 9.100349 | 0.302519 | 0.76341 | 0.993607 | -5.41337 |
| P07360 | C8G | 0.268016 | 8.367863 | 0.297056 | 0.767984 | 0.993607 | -5.31265 |
| O00187 | MASP2 | -0.25607 | 13.7813 | -0.29497 | 0.769139 | 0.993607 | -5.46327 |
| Q01813 | PFKP | -0.16094 | 10.6556 | -0.29397 | 0.769876 | 0.993607 | -5.46353 |
| P35613-2 | #N/D | 0.155723 | 10.74277 | 0.293497 | 0.770239 | 0.993607 | -5.46365 |
| P53396 | ACLY | 0.22936 | 8.061162 | 0.292689 | 0.770938 | 0.993607 | -5.46381 |
| P41250 | GARS1 | -0.22589 | 9.552422 | -0.29187 | 0.771659 | 0.993607 | -5.41591 |
| P13591 | NCAM1 | 0.195161 | 12.6968 | 0.29094 | 0.772183 | 0.993607 | -5.46429 |
| P30101 | PDIA3 | -0.21245 | 12.87952 | -0.28775 | 0.774608 | 0.993607 | -5.46507 |
| Q01518 | CAP1 | 0.204015 | 11.32087 | 0.286711 | 0.775403 | 0.993607 | -5.46533 |
| Q12765 | SCRN1 | -0.12038 | 11.08891 | -0.28631 | 0.77571 | 0.993607 | -5.46542 |
| P10606 | COX5B | -0.23951 | 8.344202 | -0.282 | 0.779121 | 0.993607 | -5.41831 |

| | | | | | | | |
|---|---|---|---|---|---|---|---|
| P04003 | C4BPA | -0.15611 | 14.39715 | -0.28152 | 0.779358 | 0.993607 | -5.46658 |
| Q07955 | SRSF1 | -0.17067 | 11.42114 | -0.27849 | 0.781674 | 0.993607 | -5.4673 |
| Q12931 | TRAP1 | 0.24456 | 11.45079 | 0.27669 | 0.783047 | 0.993607 | -5.46773 |
| P19827 | ITIH1 | 0.117966 | 15.08481 | 0.276386 | 0.78328 | 0.993607 | -5.4678 |
| P13671 | C6 | 0.128923 | 13.17346 | 0.273956 | 0.785137 | 0.993607 | -5.46837 |
| Q15149 | PLEC | -0.12085 | 12.56068 | -0.27368 | 0.78535 | 0.993607 | -5.46843 |
| P48426 | PIP4K2A | -0.15531 | 8.06123 | -0.27278 | 0.786444 | 0.993607 | -5.37814 |
| P50213 | IDH3A | 0.188269 | 10.5341 | 0.27177 | 0.786809 | 0.993607 | -5.46887 |
| Q15102 | PAFAH1B3 | -0.22309 | 9.201983 | -0.27135 | 0.787164 | 0.993607 | -5.46895 |
| P31327 | CPS1 | 0.210763 | 9.318571 | 0.271116 | 0.787309 | 0.993607 | -5.46902 |
| Q7L0J3 | SV2A | -0.24966 | 9.396699 | -0.26875 | 0.789488 | 0.993607 | -5.42122 |
| P14136 | GFAP | -0.13127 | 13.25535 | -0.26743 | 0.790129 | 0.993607 | -5.46987 |
| Q9Y2T3 | GDA | 0.234143 | 10.08686 | 0.267235 | 0.790317 | 0.993607 | -5.35417 |
| Q9H9Z2 | LIN28A | -0.21711 | 9.50441 | -0.26549 | 0.791632 | 0.993823 | -5.4703 |
| P27348 | YWHAQ | -0.20535 | 11.63346 | -0.2632 | 0.793389 | 0.994012 | -5.47081 |
| P14324 | FDPS | 0.155267 | 8.575581 | 0.262242 | 0.794295 | 0.994012 | -5.42276 |
| P01700 | #N/D | 0.280915 | 10.59788 | 0.258661 | 0.796873 | 0.994012 | -5.42362 |
| Q5T7N2 | L1TD1 | -0.15317 | 8.91276 | -0.25797 | 0.797482 | 0.994012 | -5.42373 |
| P29218 | IMPA1 | -0.15414 | 11.50747 | -0.25783 | 0.797495 | 0.994012 | -5.47201 |
| P32119 | PRDX2 | 0.141289 | 13.16617 | 0.254682 | 0.799912 | 0.995598 | -5.4727 |
| P01024 | C3 | 0.153657 | 15.31739 | 0.250015 | 0.8035 | 0.998604 | -5.4737 |
| P49327 | FASN | -0.13388 | 12.45025 | -0.24856 | 0.804622 | 0.998604 | -5.47401 |
| P32322 | PYCR1 | 0.304026 | 8.954011 | 0.245581 | 0.806948 | 0.999854 | -5.47462 |
| Q14240 | EIF4A2 | -0.14568 | 9.225594 | -0.24268 | 0.809151 | 0.999854 | -5.47524 |
| P27918 | CFP | 0.16021 | 11.216 | 0.239141 | 0.811879 | 0.999854 | -5.47596 |
| P35080 | PFN2 | 0.175477 | 10.95784 | 0.237466 | 0.813187 | 0.999854 | -5.47629 |
| P51884 | LUM | -0.15992 | 8.535854 | -0.23482 | 0.815383 | 0.999854 | -5.42846 |
| Q9UI12 | ATP6V1H | -0.21412 | 8.594544 | -0.23082 | 0.818337 | 0.999854 | -5.47761 |
| O75475 | PSIP1 | -0.17692 | 10.76389 | -0.22855 | 0.82006 | 0.999854 | -5.47807 |
| P08185 | SERPINA6 | -0.1832 | 9.106814 | -0.22832 | 0.820611 | 0.999854 | -5.32701 |
| P80404 | ABAT | -0.10754 | 7.872349 | -0.22723 | 0.821362 | 0.999854 | -5.42987 |

| | | | | | | | |
|---|---|---|---|---|---|---|---|
| Q13509 | TUBB3 | -0.12204 | 9.79097 | -0.22474 | 0.823021 | 0.999854 | -5.47879 |
| P50914 | RPL14 | 0.153765 | 9.06483 | 0.215044 | 0.830584 | 0.999854 | -5.48059 |
| P00450 | CP | -0.12322 | 14.6821 | -0.21388 | 0.831426 | 0.999854 | -5.48083 |
| Q8B6J5 | #N/D | 0.217557 | 11.86572 | 0.212331 | 0.832799 | 0.999854 | -5.48104 |
| P02753 | RBP4 | -0.20451 | 9.614475 | -0.20861 | 0.835575 | 0.999854 | -5.43335 |
| P52209 | PGD | 0.091299 | 9.886713 | 0.208366 | 0.835707 | 0.999854 | -5.48182 |
| P23396 | RPS3 | 0.142041 | 12.74094 | 0.20699 | 0.836776 | 0.999854 | -5.48206 |
| Q9NUQ9 | CYRIB | -0.17634 | 9.681427 | -0.20685 | 0.836932 | 0.999854 | -5.43366 |
| P02746 | C1QB | -0.14958 | 14.57706 | -0.20576 | 0.837729 | 0.999854 | -5.48228 |
| P00338 | LDHA | -0.10761 | 12.44144 | -0.20542 | 0.837994 | 0.999854 | -5.48234 |
| Q9UHY7 | ENOPH1 | 0.123951 | 8.101761 | 0.205132 | 0.838386 | 0.999854 | -5.43391 |
| P61266-2 | #N/D | -0.09491 | 9.516859 | -0.20387 | 0.839199 | 0.999854 | -5.48261 |
| Q16181 | SEPTIN7 | -0.26324 | 11.66741 | -0.20314 | 0.839768 | 0.999854 | -5.48273 |
| P10636-5 | #N/D | -0.08995 | 11.53737 | -0.20106 | 0.841391 | 0.999854 | -5.48309 |
| P62987 | UBA52 | -0.07739 | 11.77756 | -0.2003 | 0.84198 | 0.999854 | -5.48322 |
| P20671 | H2AC7 | -0.14677 | 8.62645 | -0.20025 | 0.84222 | 0.999854 | -5.43472 |
| Q02252 | ALDH6A1 | -0.2691 | 13.39322 | -0.19894 | 0.84304 | 0.999854 | -5.48345 |
| P36955 | SERPINF1 | 0.116975 | 12.75803 | 0.19706 | 0.844501 | 0.999854 | -5.48377 |
| P46776 | RPL27A | 0.161611 | 10.85159 | 0.196143 | 0.845229 | 0.999854 | -5.48392 |
| Q13151 | HNRNPA0 | -0.14208 | 9.990869 | -0.19383 | 0.847054 | 0.999854 | -5.43584 |
| Q9UPY8 | MAPRE3 | 0.090426 | 11.03301 | 0.192086 | 0.848377 | 0.999854 | -5.48459 |
| P21333 | FLNA | -0.11157 | 13.07842 | -0.18816 | 0.851437 | 0.999854 | -5.48523 |
| P60866 | RPS20 | -0.13351 | 8.421034 | -0.18184 | 0.856441 | 0.999854 | -5.43771 |
| O75390 | CS | 0.152879 | 11.12214 | 0.180326 | 0.857555 | 0.999854 | -5.48645 |
| Q9HB71 | CACYBP | 0.151423 | 8.507432 | 0.180048 | 0.857916 | 0.999854 | -5.48644 |
| P02792 | FTL | 0.129399 | 9.643032 | 0.176798 | 0.860349 | 0.999854 | -5.43847 |
| P01876 | #N/D | 0.135153 | 15.78121 | 0.176249 | 0.860741 | 0.999854 | -5.48707 |
| P07996 | THBS1 | -0.08044 | 13.73619 | -0.17253 | 0.863646 | 0.999854 | -5.48762 |
| P05026 | ATP1B1 | 0.127517 | 9.719583 | 0.171632 | 0.864414 | 0.999854 | -5.43922 |
| P49419 | ALDH7A1 | 0.119351 | 13.146 | 0.170504 | 0.865235 | 0.999854 | -5.48792 |
| O14980 | XPO1 | -0.11889 | 7.346187 | -0.17069 | 0.865384 | 0.999854 | -5.39675 |

| | | | | | | | |
|---|---|---|---|---|---|---|---|
| P13639 | EEF2 | 0.093391 | 12.11228 | 0.168473 | 0.866824 | 0.999854 | -5.48821 |
| P01706 | #N/D | -0.09931 | 8.121541 | -0.16671 | 0.868321 | 0.999854 | -5.43989 |
| O14531 | DPYSL4 | 0.109454 | 8.30417 | 0.166524 | 0.868536 | 0.999854 | -5.4399 |
| P50993 | ATP1A2 | -0.14239 | 10.45463 | -0.16624 | 0.868588 | 0.999854 | -5.48853 |
| P04275 | VWF | -0.12402 | 14.63316 | -0.16575 | 0.868953 | 0.999854 | -5.4886 |
| P42766 | RPL35 | -0.09657 | 9.474201 | -0.16463 | 0.869846 | 0.999854 | -5.48875 |
| P20700 | LMNB1 | 0.125497 | 10.71505 | 0.16354 | 0.870688 | 0.999854 | -5.48891 |
| P01599 | #N/D | -0.14365 | 9.808588 | -0.16246 | 0.871555 | 0.999854 | -5.44051 |
| P02679-2 | #N/D | -0.07554 | 11.17822 | -0.1608 | 0.872839 | 0.999854 | -5.48929 |
| Q9BY11 | PACSIN1 | -0.14476 | 11.03934 | -0.16079 | 0.872844 | 0.999854 | -5.48929 |
| P20073 | ANXA7 | -0.08933 | 8.552518 | -0.1607 | 0.872922 | 0.999854 | -5.4893 |
| P11169 | SLC2A3 | 0.130518 | 9.491977 | 0.158761 | 0.874456 | 0.999854 | -5.48956 |
| P01742 | #N/D | -0.11035 | 13.16148 | -0.15881 | 0.874559 | 0.999854 | -5.44095 |
| P48643 | CCT5 | -0.10643 | 9.491954 | -0.1586 | 0.87456 | 0.999854 | -5.48959 |
| Q04837 | SSBP1 | -0.13534 | 9.844274 | -0.15845 | 0.874685 | 0.999854 | -5.4896 |
| P61604 | HSPE1 | 0.111926 | 11.60591 | 0.157924 | 0.875091 | 0.999854 | -5.48968 |
| P62829 | RPL23 | -0.18184 | 7.660806 | -0.15803 | 0.875261 | 0.999854 | -5.24351 |
| O95989 | NUDT3 | 0.119208 | 9.707781 | 0.155919 | 0.876663 | 0.999854 | -5.48995 |
| Q04637 | EIF4G1 | 0.116591 | 9.520493 | 0.155902 | 0.876709 | 0.999854 | -5.48994 |
| Q99729-3 | #N/D | -0.16687 | 12.65612 | -0.15503 | 0.877363 | 0.999854 | -5.49006 |
| Q96AX9-5 | #N/D | -0.13555 | 9.366756 | -0.15241 | 0.87943 | 0.999854 | -5.4904 |
| Q04695 | KRT17 | 0.10516 | 10.32094 | 0.149701 | 0.881575 | 0.999854 | -5.49075 |
| Q9UKX2 | MYH2 | 0.140296 | 9.36748 | 0.147969 | 0.882957 | 0.999854 | -5.49096 |
| O43491-4 | #N/D | 0.099803 | 9.677072 | 0.14789 | 0.882967 | 0.999854 | -5.49098 |
| P07197 | NEFM | -0.06218 | 12.44543 | -0.14648 | 0.884071 | 0.999854 | -5.49116 |
| O43866 | CD5L | -0.07123 | 13.65287 | -0.14485 | 0.885357 | 0.999854 | -5.49136 |
| P27361 | MAPK3 | 0.090174 | 9.665968 | 0.144329 | 0.885764 | 0.999854 | -5.49143 |
| Q92597 | NDRG1 | -0.09922 | 10.27498 | -0.14004 | 0.889152 | 0.999854 | -5.44334 |
| P46782 | RPS5 | -0.08676 | 8.807193 | -0.13981 | 0.88938 | 0.999854 | -5.49196 |
| P00734 | F2 | 0.08579 | 14.7427 | 0.138671 | 0.890213 | 0.999854 | -5.49211 |
| P05455 | SSB | 0.111866 | 10.60567 | 0.135326 | 0.892853 | 0.999854 | -5.4925 |

| | | | | | | | |
|---|---|---|---|---|---|---|---|
| Q12860 | CNTN1 | -0.08902 | 11.73525 | -0.13512 | 0.89301 | 0.999854 | -5.49252 |
| P09960 | LTA4H | 0.083455 | 8.842087 | 0.134237 | 0.893772 | 0.999854 | -5.44399 |
| Q15435 | PPP1R7 | -0.11468 | 8.920446 | -0.13358 | 0.894274 | 0.999854 | -5.44406 |
| P13637 | ATP1A3 | 0.103915 | 12.48297 | 0.133475 | 0.894301 | 0.999854 | -5.49271 |
| Q03591 | CFHR1 | -0.0872 | 9.2033 | -0.13301 | 0.89473 | 0.999854 | -5.44413 |
| P13667 | PDIA4 | -0.08943 | 11.88336 | -0.12988 | 0.897135 | 0.999854 | -5.49311 |
| Q5JNZ5 | #N/D | 0.101131 | 10.43992 | 0.129861 | 0.897147 | 0.999854 | -5.49312 |
| P39019 | RPS19 | -0.12554 | 12.71185 | -0.12916 | 0.897702 | 0.999854 | -5.49319 |
| P68133 | ACTA1 | 0.073792 | 8.536068 | 0.128635 | 0.898255 | 0.999854 | -5.40196 |
| P07357 | C8A | -0.07228 | 14.30403 | -0.12786 | 0.89872 | 0.999854 | -5.49333 |
| P60709 | ACTB | 0.074607 | 12.29443 | 0.124767 | 0.90116 | 0.999876 | -5.49367 |
| P06753-2 | #N/D | 0.103926 | 11.55725 | 0.124361 | 0.901524 | 0.999876 | -5.4937 |
| P04216 | THY1 | -0.11335 | 9.984486 | -0.11862 | 0.906007 | 0.999876 | -5.4943 |
| P12532 | CKMT1B | -0.07437 | 9.736519 | -0.11017 | 0.912674 | 0.999876 | -5.49513 |
| O00154 | ACOT7 | -0.08598 | 10.65194 | -0.10327 | 0.918121 | 0.999876 | -5.49575 |
| Q96AE4 | FUBP1 | 0.07831 | 9.61624 | 0.101326 | 0.919658 | 0.999876 | -5.49592 |
| P50991 | CCT4 | -0.05159 | 13.00041 | -0.10024 | 0.920516 | 0.999876 | -5.49602 |
| P07196 | NEFL | 0.038281 | 12.62642 | 0.099605 | 0.921017 | 0.999876 | -5.49607 |
| O95865 | DDAH2 | -0.09008 | 8.695045 | -0.0982 | 0.922179 | 0.999876 | -5.44749 |
| Q02218 | OGDH | 0.061393 | 8.055839 | 0.094237 | 0.925345 | 0.999876 | -5.4478 |
| P13645 | KRT10 | 0.054788 | 15.33348 | 0.092621 | 0.926539 | 0.999876 | -5.49664 |
| A0A075B6I | #N/D | 0.089446 | 9.154402 | 0.089503 | 0.929076 | 0.999876 | -5.44817 |
| P30050 | RPL12 | -0.07439 | 10.32925 | -0.08678 | 0.931158 | 0.999876 | -5.49709 |
| Q15717 | ELAVL1 | 0.071037 | 11.8003 | 0.086845 | 0.93116 | 0.999876 | -5.44837 |
| P53004 | BLVRA | 0.066318 | 7.896539 | 0.086131 | 0.931717 | 0.999876 | -5.44842 |
| P0C0S5 | H2AZ1 | -0.06459 | 10.02911 | -0.0853 | 0.932331 | 0.999876 | -5.4972 |
| Q9Y2W1 | THRAP3 | -0.05317 | 8.282812 | -0.08384 | 0.933507 | 0.999876 | -5.4973 |
| P46108 | CRK | 0.054532 | 8.591725 | 0.082353 | 0.934686 | 0.999876 | -5.44869 |
| P02042 | HBD | 0.048887 | 10.61689 | 0.082136 | 0.934835 | 0.999876 | -5.49742 |
| P63162 | SNRPN | 0.131004 | 6.935315 | 0.082701 | 0.934936 | 0.999876 | -5.14946 |
| P62280 | RPS11 | -0.04945 | 11.12395 | -0.07988 | 0.93662 | 0.999876 | -5.49758 |

| | | | | | | | |
|---|---|---|---|---|---|---|---|
| P02741 | CRP | 0.096928 | 11.11435 | 0.079186 | 0.937176 | 0.999876 | -5.49762 |
| P04217 | A1BG | -0.04475 | 14.14437 | -0.07899 | 0.937326 | 0.999876 | -5.49764 |
| Q9H3S7 | PTPN23 | 0.042528 | 9.197781 | 0.07548 | 0.940104 | 0.999876 | -5.49787 |
| Q9BWD1-2 | #N/D | -0.05003 | 7.847079 | -0.07364 | 0.941648 | 0.999876 | -5.44925 |
| P18206 | VCL | -0.02874 | 12.66382 | -0.07309 | 0.942 | 0.999876 | -5.49802 |
| P54652 | HSPA2 | 0.041443 | 13.29815 | 0.072045 | 0.942826 | 0.999876 | -5.49808 |
| P49591 | SARS1 | -0.0439 | 8.044302 | -0.07151 | 0.943309 | 0.999876 | -5.49811 |
| P01011 | SERPINA3 | 0.028869 | 14.45594 | 0.070866 | 0.94376 | 0.999876 | -5.49816 |
| P09211 | GSTP1 | -0.04791 | 13.54278 | -0.07074 | 0.943913 | 0.999876 | -5.44943 |
| P02511 | CRYAB | 0.033087 | 10.8827 | 0.069525 | 0.944822 | 0.999876 | -5.49824 |
| Q92945 | KHSRP | 0.049966 | 11.19284 | 0.069468 | 0.944867 | 0.999876 | -5.49824 |
| P23526 | AHCY | 0.035301 | 12.4871 | 0.069352 | 0.944959 | 0.999876 | -5.49825 |
| P62861 | #N/D | 0.050152 | 9.145956 | 0.068854 | 0.945373 | 0.999876 | -5.44954 |
| P36542 | ATP5F1C | 0.059217 | 8.889386 | 0.065153 | 0.948291 | 0.999876 | -5.49849 |
| P02675 | FGB | 0.03256 | 14.18233 | 0.064829 | 0.948544 | 0.999876 | -5.4985 |
| P05109 | S100A8 | -0.05212 | 9.04775 | -0.06342 | 0.949672 | 0.999876 | -5.49858 |
| A0A0G2JS( | #N/D | -0.07218 | 9.269116 | -0.0634 | 0.949724 | 0.999876 | -5.40711 |
| P01009 | SERPINA1 | 0.031242 | 14.60704 | 0.061549 | 0.951143 | 0.999876 | -5.49868 |
| P14625 | HSP90B1 | 0.060982 | 13.43176 | 0.060575 | 0.951915 | 0.999876 | -5.49873 |
| P61981 | YWHAG | -0.05572 | 13.78262 | -0.06052 | 0.951958 | 0.999876 | -5.49873 |
| Q14697-2 | #N/D | 0.040381 | 9.735768 | 0.058693 | 0.953419 | 0.999876 | -5.45009 |
| P28482 | MAPK1 | 0.027947 | 10.57932 | 0.058527 | 0.953543 | 0.999876 | -5.49884 |
| P31943 | HNRNPH1 | -0.03648 | 12.25814 | -0.05845 | 0.9536 | 0.999876 | -5.49884 |
| P07339 | CTSD | -0.0438 | 12.19536 | -0.05641 | 0.955216 | 0.999876 | -5.49894 |
| P52565 | ARHGDIA | -0.04961 | 10.94718 | -0.05538 | 0.95604 | 0.999876 | -5.49899 |
| Q9UNZ2 | NSFL1C | -0.04337 | 8.71179 | -0.05474 | 0.956558 | 0.999876 | -5.40754 |
| P61313 | RPL15 | -0.02278 | 10.74977 | -0.05145 | 0.959151 | 0.999876 | -5.49917 |
| Q9H115 | NAPB | 0.032379 | 9.074792 | 0.051093 | 0.959462 | 0.999876 | -5.45043 |
| Q15848 | ADIPOQ | 0.035205 | 10.82374 | 0.050633 | 0.9598 | 0.999876 | -5.4992 |
| A0A0B4J1Y | #N/D | -0.03481 | 11.1449 | -0.04814 | 0.961782 | 0.999876 | -5.49931 |
| Q14624-3 | #N/D | -0.04398 | 8.418841 | -0.04664 | 0.963074 | 0.999876 | -5.34693 |

| | | | | | | | |
|---|---|---|---|---|---|---|---|
| P15121 | AKR1B1 | -0.02858 | 9.04325 | -0.04271 | 0.966116 | 0.999876 | -5.45076 |
| P08670 | VIM | 0.016779 | 13.17483 | 0.04222 | 0.966475 | 0.999876 | -5.49953 |
| Q8IV08 | PLD3 | -0.02841 | 9.113681 | -0.04094 | 0.967492 | 0.999876 | -5.49958 |
| P02787 | TF | -0.0226 | 14.24336 | -0.04065 | 0.967718 | 0.999876 | -5.49959 |
| P63010 | AP2B1 | -0.02836 | 10.0117 | -0.0381 | 0.969741 | 0.999876 | -5.49968 |
| O60506 | SYNCRIP | -0.02751 | 8.844164 | -0.03755 | 0.970198 | 0.999876 | -5.45093 |
| P11137 | MAP2 | -0.02036 | 10.40085 | -0.03481 | 0.972353 | 0.999876 | -5.49978 |
| P32004 | L1CAM | 0.023184 | 9.227821 | 0.034473 | 0.972639 | 0.999876 | -5.45103 |
| Q99832 | CCT7 | 0.013753 | 11.03507 | 0.026024 | 0.979331 | 0.999876 | -5.50001 |
| Q96FC7 | PHYHIPL | -0.02601 | 10.07371 | -0.02583 | 0.979489 | 0.999876 | -5.50001 |
| Q9H0U4 | RAB1B | 0.014265 | 8.154821 | 0.025155 | 0.980042 | 0.999876 | -5.45126 |
| Q9Y266 | NUDC | -0.01889 | 9.322208 | -0.0232 | 0.981581 | 0.999876 | -5.4513 |
| P08697 | SERPINF2 | 0.010555 | 12.89207 | 0.023159 | 0.981606 | 0.999876 | -5.50007 |
| P49418 | AMPH | 0.018199 | 10.79496 | 0.022569 | 0.982075 | 0.999876 | -5.50008 |
| P02765 | AHSG | 0.011712 | 11.76711 | 0.018574 | 0.985248 | 0.999876 | -5.50015 |
| P62826 | RAN | 0.008053 | 12.49633 | 0.01684 | 0.986624 | 0.999876 | -5.50017 |
| O15540 | FABP7 | 0.008149 | 10.93642 | 0.016708 | 0.98673 | 0.999876 | -5.50017 |
| P0DJI9 | SAA2 | -0.01451 | 8.735337 | -0.0165 | 0.986901 | 0.999876 | -5.45141 |
| O94819 | KBTBD11 | -0.00927 | 9.342712 | -0.01443 | 0.98854 | 0.999876 | -5.50021 |
| P31025 | LCN1 | 0.00995 | 14.44478 | 0.013116 | 0.989582 | 0.999876 | -5.50022 |
| Q14194 | CRMP1 | -0.00604 | 11.68612 | -0.01184 | 0.990594 | 0.999876 | -5.50023 |
| P14649 | MYL6B | 0.006264 | 11.20495 | 0.011605 | 0.990782 | 0.999876 | -5.50024 |
| P35232 | PHB | 0.005049 | 11.88214 | 0.010121 | 0.991961 | 0.999876 | -5.50025 |
| P28070 | PSMB4 | -0.00639 | 9.16426 | -0.00935 | 0.992578 | 0.999876 | -5.50026 |
| Q13363 | CTBP1 | -0.00704 | 9.960849 | -0.00921 | 0.992684 | 0.999876 | -5.50026 |
| Q5HYA8 | TMEM67 | 0.007344 | 9.59421 | 0.009059 | 0.992805 | 0.999876 | -5.50026 |
| P36871 | PGM1 | -0.00407 | 11.50669 | -0.00716 | 0.994312 | 0.999876 | -5.50027 |
| P61254 | RPL26 | -0.00413 | 10.98212 | -0.00656 | 0.99479 | 0.999876 | -5.50028 |
| Q6TUY0 | #N/D | -0.00725 | 12.02649 | -0.00481 | 0.996179 | 0.999876 | -5.50028 |
| P01008 | SERPINC1 | 0.002458 | 14.22003 | 0.004759 | 0.99622 | 0.999876 | -5.50028 |
| Q9Y3E1 | HDGFL3 | -0.00435 | 8.28437 | -0.0045 | 0.996429 | 0.999876 | -5.50029 |

| | | | | | | | |
|---|---|---|---|---|---|---|---|
| P01611 | #N/D | 0.00302 | 10.90728 | 0.003517 | 0.997209 | 0.999876 | -5.45152 |
| Q86VP6 | CAND1 | 0.001934 | 9.469724 | 0.002554 | 0.997971 | 0.999876 | -5.50029 |
| P49411 | TUFM | -0.00084 | 11.6554 | -0.00101 | 0.999196 | 0.999876 | -5.50029 |
| P60880 | SNAP25 | 0.000293 | 10.94386 | 0.000436 | 0.999654 | 0.999876 | -5.50029 |
| P10809 | HSPD1 | -6.9E-05 | 14.28311 | -0.00016 | 0.999876 | 0.999876 | -5.50029 |
| Q99714 | HSD17B10 | #N/D | 7.015178 | #N/D | #N/D | #N/D | #N/D |

**Supplemental Table 3.** Differentially expressed proteins in astrocytic extracellular vesicles (aEVs) isolated from patients following CAND treatment compared to the pre-CAND condition and the complete list of all proteins identified in the proteomic analysis.

Each row corresponds to a protein identified by its UniProt accession number, gene symbol, and protein symbol. Expression changes are reported as $log_2$ fold change (logFC); positive values indicate upregulation after CAND treatment, whereas negative values indicate downregulation. AveExpr represents the average expression across all samples. The t-statistic and associated P value were derived from moderated t-tests. The B statistic reflects the log-odds that a given protein is differentially expressed.

#### Supplemental Table 3. Differentially Expressed Proteins in aEVs

| UniProt | Gene Symbol | Protein name | logFC | AveExpr | t | P.Value | B |
|---|---|---|---|---|---|---|---|
| P46778 | RPL21 | RL21 | 3.844040513 | 9.748979406 | 2.132803542 | 0.037820006 | -3.740386164 |
| P28070 | PSMB4 | PSB4 | 2.365177945 | 9.164259623 | 3.456944568 | 0.00107959 | -1.876924058 |
| P62899 | RPL31 | RPL31 | 2.310880994 | 12.22518748 | 2.198721005 | 0.03209017 | -3.657572305 |
| P80108 | GPLD1 | PHLD | 2.088219059 | 9.22442799 | 2.722394401 | 0.008779955 | -3.151381253 |
| A0A075B6J9 | IGLV2-18 | LV218 | 2.059471338 | 10.90519084 | 2.131824247 | 0.037727682 | -3.739675371 |
| Q9NSD9 | FARSB | SYFB | 2.058546445 | 9.390867496 | 2.723164644 | 0.008965342 | -3.122403466 |
| P12277 | CKB | KCRB | 1.725714468 | 13.54112066 | 2.453866194 | 0.017303457 | -3.337852224 |
| Q13449 | LSAMP | LSAMP | 1.65404696 | 9.843598817 | 2.917492907 | 0.005088103 | -2.694846956 |
| P28074 | PSMB5 | PSB5 | 1.578010565 | 8.249192656 | 2.041056117 | 0.047081941 | -3.934705206 |
| P80748 | IGLV3-21 | LV321 | 1.565751746 | 13.59399095 | 2.196031697 | 0.032292556 | -3.660800338 |
| P04040 | CAT | CATA | -1.270294159 | 8.854697609 | -2.605122561 | 0.01237322 | -3.274646142 |
| O43301 | HSPA12A | HS12A | -1.277785478 | 10.41348104 | -2.176031326 | 0.033996128 | -3.686670832 |
| Q9UHY7 | ENOPH1 | ENOPH | -1.46321664 | 8.101761466 | -2.297263674 | 0.026264973 | -3.627364289 |
| P25788 | PSMA3 | PSA3 | -1.544552579 | 9.35689776 | -2.150620119 | 0.036324579 | -3.7498115 |
| P31939 | ATIC | PUR9 | -1.631499286 | 12.54945522 | -2.241945829 | 0.028987135 | -3.605271868 |
| P06310 | GKV2-30 | KV230 | -1.636827524 | 13.49282589 | -2.434638152 | 0.018153959 | -3.362847707 |
| P01876 | IGHA1 | IGHA1 | -1.705896388 | 15.78121441 | -2.224617786 | 0.030197738 | -3.626332071 |
| O60282 | KIF5C | KIF5C | -1.821629381 | 9.337803981 | -2.226506162 | 0.030301965 | -3.661959166 |
| Q5JNZ5 | RPS26P11 | RS26L | -2.477657897 | 10.43992318 | -3.181518136 | 0.002401206 | -2.297821777 |
| A5YM72 | CARNS1 | CRNS1 | -2.59618676 | 9.654884741 | -2.97793846 | 0.004561601 | -2.724070103 |
| Q06033 | ITIH3 | ITIH3 | -2.763625414 | 13.55378255 | -2.313896049 | 0.024407399 | -3.516499021 |
| P61956 | SUMO2 | SUMO2 | -3.676756279 | 9.670243454 | -4.212831665 | 9.55098E-05 | -0.606077264 |

#### Supplemental Table 3. All Quantified proteins

| UniProt | Gene Symbol | logFC | AveExpr | t | P.Value | adj.P.Val | B |
|---|---|---|---|---|---|---|---|
| P61956 | SUMO2 | -3.67676 | 9.670243 | -4.21283 | 9.55E-05 | 0.082903 | -0.60608 |
| P28070 | PSMB4 | 2.365178 | 9.16426 | 3.456945 | 0.00108 | 0.468542 | -1.87692 |
| Q5JNZ5 | #N/D | -2.47766 | 10.43992 | -3.18152 | 0.002401 | 0.694749 | -2.29782 |
| A5YM72 | CARNS1 | -2.59619 | 9.654885 | -2.97794 | 0.004562 | 0.883295 | -2.72407 |
| Q13449 | LSAMP | 1.654047 | 9.843599 | 2.917493 | 0.005088 | 0.883295 | -2.69485 |
| P80108 | GPLD1 | 2.088219 | 9.224428 | 2.722394 | 0.00878 | 0.994456 | -3.15138 |
| Q9NSD9 | FARSB | 2.058546 | 9.390867 | 2.723165 | 0.008965 | 0.994456 | -3.1224 |
| P04040 | CAT | -1.27029 | 8.854698 | -2.60512 | 0.012373 | 0.994456 | -3.27465 |
| P12277 | CKB | 1.725714 | 13.54112 | 2.453866 | 0.017303 | 0.994456 | -3.33785 |
| P06310 | #N/D | -1.63683 | 13.49283 | -2.43464 | 0.018154 | 0.994456 | -3.36285 |
| Q06033 | ITIH3 | -2.76363 | 13.55378 | -2.3139 | 0.024407 | 0.994456 | -3.5165 |
| Q9UHY7 | ENOPH1 | -1.46322 | 8.101761 | -2.29726 | 0.026265 | 0.994456 | -3.62736 |
| P31939 | ATIC | -1.6315 | 12.54946 | -2.24195 | 0.028987 | 0.994456 | -3.60527 |
| P01876 | #N/D | -1.7059 | 15.78121 | -2.22462 | 0.030198 | 0.994456 | -3.62633 |
| O60282 | KIF5C | -1.82163 | 9.337804 | -2.22651 | 0.030302 | 0.994456 | -3.66196 |
| P62899 | RPL31 | 2.310881 | 12.22519 | 2.198721 | 0.03209 | 0.994456 | -3.65757 |
| P80748 | #N/D | 1.565752 | 13.59399 | 2.196032 | 0.032293 | 0.994456 | -3.6608 |
| O43301 | HSPA12A | -1.27779 | 10.41348 | -2.17603 | 0.033996 | 0.994456 | -3.68667 |
| P25788 | PSMA3 | -1.54455 | 9.356898 | -2.15062 | 0.036325 | 0.994456 | -3.74981 |
| A0A075B6. | #N/D | 2.059471 | 10.90519 | 2.131824 | 0.037728 | 0.994456 | -3.73968 |
| P46778 | RPL21 | 3.844041 | 9.748979 | 2.132804 | 0.03782 | 0.994456 | -3.74039 |
| P28074 | PSMB5 | 1.578011 | 8.249193 | 2.041056 | 0.047082 | 0.994456 | -3.93471 |
| O00264 | PGRMC1 | -1.47575 | 9.27944 | -1.99166 | 0.05184 | 0.994456 | -3.90036 |
| P06331 | #N/D | 1.365481 | 11.07987 | 1.974128 | 0.05336 | 0.994456 | -3.91636 |
| P27361 | MAPK3 | 1.212404 | 9.665968 | 1.940535 | 0.057409 | 0.994456 | -3.95314 |
| P21579 | SYT1 | -1.34828 | 9.733547 | -1.93579 | 0.058002 | 0.994456 | -3.95829 |
| P36578 | RPL4 | 0.767935 | 12.0117 | 1.932252 | 0.058447 | 0.994456 | -3.96213 |
| A0A075B6I | #N/D | 1.245712 | 9.618077 | 1.924909 | 0.05938 | 0.994456 | -3.97007 |

| | | | | | | | |
|---|---|---|---|---|---|---|---|
| Q16851 | UGP2 | 0.759099 | 12.72477 | 1.924169 | 0.059475 | 0.994456 | -3.97087 |
| P12036 | NEFH | 0.715422 | 12.50765 | 1.90995 | 0.061321 | 0.994456 | -3.98617 |
| P27797 | CALR | 1.028188 | 10.33747 | 1.904096 | 0.062095 | 0.994456 | -3.99244 |
| O75781 | PALM | 1.148433 | 8.620566 | 1.886583 | 0.064657 | 0.994456 | -4.02768 |
| P15121 | AKR1B1 | 1.312317 | 9.04325 | 1.860965 | 0.06923 | 0.994456 | -4.07247 |
| Q5IFJ7 | #N/D | -2.24747 | 10.61788 | -1.84128 | 0.071034 | 0.994456 | -4.05917 |
| P13671 | C6 | 0.857003 | 13.17346 | 1.8211 | 0.073992 | 0.994456 | -4.07965 |
| P12532 | CKMT1B | -1.22793 | 9.736519 | -1.81901 | 0.074315 | 0.994456 | -4.08181 |
| P12004 | PCNA | 0.902599 | 11.20119 | 1.81299 | 0.075251 | 0.994456 | -4.088 |
| P00739 | HPR | -1.34702 | 13.20664 | -1.77326 | 0.08168 | 0.994456 | -4.12843 |
| P01764 | #N/D | -1.33684 | 9.51328 | -1.77295 | 0.082401 | 0.994456 | -4.14085 |
| P38117 | ETFB | 1.629699 | 8.695554 | 1.747675 | 0.08686 | 0.994456 | -4.1817 |
| P68133 | ACTA1 | -1.00162 | 8.536068 | -1.74603 | 0.088045 | 0.994456 | -4.17902 |
| O00499 | BIN1 | -1.3874 | 8.599564 | -1.73132 | 0.091023 | 0.994456 | -4.22556 |
| Q96KP4 | CNDP2 | -0.85006 | 11.48894 | -1.70552 | 0.093698 | 0.994456 | -4.19562 |
| P01009 | SERPINA1 | 0.864694 | 14.60704 | 1.703522 | 0.094073 | 0.994456 | -4.19757 |
| P09012 | SNRPA | -1.64154 | 9.436966 | -1.70675 | 0.094852 | 0.994456 | -4.21269 |
| P60201 | PLP1 | 0.942654 | 12.86375 | 1.697249 | 0.09526 | 0.994456 | -4.20367 |
| Q9HB71 | CACYBP | -1.94835 | 8.507432 | -1.69185 | 0.097525 | 0.994456 | -4.26744 |
| Q9NPH9 | IL26 | 1.615563 | 9.277746 | 1.674161 | 0.100293 | 0.994456 | -4.2273 |
| O43143 | DHX15 | 1.693122 | 7.908015 | 1.670393 | 0.10257 | 0.994456 | -4.29697 |
| P02774-3 | #N/D | 1.305776 | 14.29976 | 1.65965 | 0.102634 | 0.994456 | -4.23984 |
| P30044 | PRDX5 | -1.95246 | 11.42963 | -1.65694 | 0.103184 | 0.994456 | -4.24242 |
| P13716-2 | #N/D | -1.24856 | 8.80277 | -1.65344 | 0.104336 | 0.994456 | -4.24672 |
| P49189 | ALDH9A1 | 1.434625 | 8.956597 | 1.641819 | 0.106615 | 0.994456 | -4.26106 |
| P00450 | CP | 0.935479 | 14.6821 | 1.623779 | 0.110098 | 0.994456 | -4.27369 |
| Q9Y2J2 | EPB41L3 | 1.210174 | 8.157606 | 1.612124 | 0.114182 | 0.994456 | -4.29373 |
| Q8IV08 | PLD3 | 1.108161 | 9.113681 | 1.59693 | 0.116073 | 0.994456 | -4.29878 |
| P02788 | LTF | -1.61618 | 9.520811 | -1.58808 | 0.118171 | 0.994456 | -4.3071 |
| Q9Y6I3 | EPN1 | 1.108256 | 8.193578 | 1.552947 | 0.127696 | 0.994456 | -4.3475 |
| Q9UNQ0 | ABCG2 | 1.739354 | 11.29824 | 1.54494 | 0.128494 | 0.994456 | -4.34643 |

| | | | | | | | |
|---|---|---|---|---|---|---|---|
| P62873 | GNB1 | -0.95438 | 12.21737 | -1.5425 | 0.128644 | 0.994456 | -4.34799 |
| P26641 | EEF1G | -0.92299 | 10.13888 | -1.53958 | 0.129457 | 0.994456 | -4.34982 |
| P14618-3 | #N/D | 1.01485 | 9.955675 | 1.532767 | 0.131127 | 0.994456 | -4.3568 |
| P05156 | CFI | -1.04607 | 13.67799 | -1.53138 | 0.131364 | 0.994456 | -4.35789 |
| P56747 | CLDN6 | -1.02399 | 8.759588 | -1.53262 | 0.132158 | 0.994456 | -4.35826 |
| Q92823 | NRCAM | 0.777189 | 13.18446 | 1.52621 | 0.132645 | 0.994456 | -4.36247 |
| P84103 | SRSF3 | 0.96971 | 7.821272 | 1.528823 | 0.1334 | 0.994456 | -4.36131 |
| Q16695 | H3-4 | 1.003078 | 10.26812 | 1.521723 | 0.133765 | 0.994456 | -4.36644 |
| P52565 | ARHGDIA | -1.3611 | 10.94718 | -1.51934 | 0.134468 | 0.994456 | -4.36867 |
| P55290 | CDH13 | -1.23183 | 9.449083 | -1.50597 | 0.138316 | 0.994456 | -4.3786 |
| P05023 | ATP1A1 | 1.020612 | 13.61348 | 1.498227 | 0.13975 | 0.994456 | -4.38704 |
| Q15717 | ELAVL1 | -1.21981 | 11.8003 | -1.49125 | 0.14252 | 0.994456 | -4.39119 |
| Q9UKX2 | MYH2 | -1.49564 | 9.36748 | -1.48724 | 0.14318 | 0.994456 | -4.39405 |
| Q92686 | NRGN | 1.220737 | 9.047773 | 1.482081 | 0.144306 | 0.994456 | -4.40137 |
| P62913 | RPL11 | -1.02376 | 12.51404 | -1.46675 | 0.148098 | 0.994456 | -4.41419 |
| P83731 | RPL24 | -1.15309 | 8.556325 | -1.47154 | 0.148527 | 0.994456 | -4.40654 |
| P11169 | SLC2A3 | -1.20338 | 9.491977 | -1.46378 | 0.149114 | 0.994456 | -4.41692 |
| Q6UWR7 | ENPP6 | -0.79364 | 11.19948 | -1.46194 | 0.149406 | 0.994456 | -4.41829 |
| O14531 | DPYSL4 | 1.094675 | 8.30417 | 1.462778 | 0.150899 | 0.994456 | -4.41347 |
| P52292 | KPNA2 | 1.133609 | 7.997937 | 1.453062 | 0.153082 | 0.994456 | -4.41991 |
| Q08209-3 | #N/D | 0.855226 | 10.11222 | 1.448749 | 0.153146 | 0.994456 | -4.42957 |
| P09936 | UCHL1 | 0.845633 | 12.48925 | 1.448138 | 0.153213 | 0.994456 | -4.42999 |
| O15540 | FABP7 | -0.70625 | 10.93642 | -1.44794 | 0.153269 | 0.994456 | -4.43016 |
| P01860 | #N/D | -1.16638 | 14.17562 | -1.43481 | 0.156962 | 0.994456 | -4.44121 |
| O75947 | ATP5PD | -1.13952 | 8.722357 | -1.43181 | 0.158024 | 0.994456 | -4.44388 |
| P17987 | TCP1 | 1.20133 | 10.0471 | 1.427724 | 0.158983 | 0.994456 | -4.44713 |
| Q9H4G4 | GLIPR2 | 0.983488 | 9.978347 | 1.422843 | 0.16049 | 0.994456 | -4.45127 |
| P51674 | GPM6A | 1.118414 | 9.977344 | 1.412187 | 0.163486 | 0.994456 | -4.46002 |
| P46777 | RPL5 | -1.27015 | 8.455774 | -1.41589 | 0.163631 | 0.994456 | -4.44641 |
| A0MZ66 | SHTN1 | -1.25752 | 8.930068 | -1.40749 | 0.16567 | 0.994456 | -4.45422 |
| P63104 | YWHAZ | -0.59892 | 13.41142 | -1.40389 | 0.165932 | 0.994456 | -4.46685 |

| | | | | | | | |
|---|---|---|---|---|---|---|---|
| P02751-8 | #N/D | 0.686577 | 15.47018 | 1.402362 | 0.166385 | 0.994456 | -4.4681 |
| Q02218 | OGDH | -0.91622 | 8.055839 | -1.40638 | 0.166571 | 0.994456 | -4.45935 |
| Q9NQC3 | RTN4 | 0.823868 | 10.95603 | 1.393743 | 0.16896 | 0.994456 | -4.47516 |
| P49588 | AARS1 | 1.908356 | 10.01352 | 1.393006 | 0.16973 | 0.994456 | -4.46916 |
| P62701 | RPS4X | 0.613542 | 10.53558 | 1.38685 | 0.171041 | 0.994456 | -4.48077 |
| P02766 | TTR | -1.22108 | 13.95342 | -1.38184 | 0.172565 | 0.994456 | -4.48483 |
| P69891 | HBG1 | 1.101051 | 10.30411 | 1.380591 | 0.17305 | 0.994456 | -4.4859 |
| Q05193 | DNM1 | 0.757596 | 10.1149 | 1.380137 | 0.173087 | 0.994456 | -4.48621 |
| O75475 | PSIP1 | -1.05881 | 10.76389 | -1.36782 | 0.176889 | 0.994456 | -4.49613 |
| Q9BW30 | TPPP3 | -1.19872 | 7.409834 | -1.37562 | 0.177827 | 0.994456 | -4.49698 |
| P15880 | RPS2 | -1.24792 | 12.70417 | -1.36452 | 0.177921 | 0.994456 | -4.49878 |
| Q99426 | TBCB | 1.04535 | 8.295429 | 1.369744 | 0.177975 | 0.994456 | -4.47774 |
| P61353 | RPL27 | -0.73016 | 9.636353 | -1.36145 | 0.178882 | 0.994456 | -4.50123 |
| Q13561 | DCTN2 | -0.95374 | 8.671465 | -1.36107 | 0.179314 | 0.994456 | -4.48799 |
| P36542 | ATP5F1C | 1.311159 | 8.889386 | 1.360079 | 0.179413 | 0.994456 | -4.49414 |
| P35232 | PHB | -0.67583 | 11.88214 | -1.35469 | 0.181013 | 0.994456 | -4.50661 |
| P02745 | C1QA | -1.28678 | 10.24676 | -1.35466 | 0.181449 | 0.994456 | -4.49841 |
| A0A0B4J1Y | #N/D | -0.97697 | 11.1449 | -1.35093 | 0.182207 | 0.994456 | -4.5096 |
| P49321 | NASP | 0.768663 | 13.01531 | 1.347138 | 0.183419 | 0.994456 | -4.51261 |
| P14136 | GFAP | 0.661101 | 13.25535 | 1.346886 | 0.1835 | 0.994456 | -4.5128 |
| Q12931 | TRAP1 | 1.187446 | 11.45079 | 1.343453 | 0.184601 | 0.994456 | -4.51552 |
| Q13363 | CTBP1 | 1.025757 | 9.960849 | 1.342025 | 0.185061 | 0.994456 | -4.51664 |
| P62249 | RPS16 | 0.637334 | 10.28851 | 1.340791 | 0.185459 | 0.994456 | -4.51762 |
| P49773 | HINT1 | 0.956297 | 8.488339 | 1.339565 | 0.186166 | 0.994456 | -4.50971 |
| P04264 | KRT1 | -0.85857 | 16.88435 | -1.33814 | 0.186316 | 0.994456 | -4.5197 |
| O76021 | RSL1D1 | 1.024103 | 9.23965 | 1.338344 | 0.186561 | 0.994456 | -4.51967 |
| O14967 | CLGN | -0.91567 | 11.39778 | -1.33877 | 0.186898 | 0.994456 | -4.51954 |
| P62263 | RPS14 | 1.04755 | 12.25025 | 1.333716 | 0.187753 | 0.994456 | -4.52318 |
| P10515 | DLAT | -0.91266 | 8.7176 | -1.3334 | 0.18806 | 0.994456 | -4.51426 |
| P13667 | PDIA4 | 0.917554 | 11.88336 | 1.33259 | 0.18812 | 0.994456 | -4.52406 |
| P40227 | CCT6A | 0.716925 | 12.26876 | 1.331758 | 0.188392 | 0.994456 | -4.52471 |

| | | | | | | | |
|---|---|---|---|---|---|---|---|
| A0A0A0MT | #N/D | -1.67878 | 9.911469 | -1.33497 | 0.188541 | 0.994456 | -4.50713 |
| Q06830 | PRDX1 | -0.69512 | 12.88336 | -1.31597 | 0.193601 | 0.994456 | -4.537 |
| P01817 | #N/D | 0.812291 | 11.01698 | 1.314196 | 0.194194 | 0.994456 | -4.53838 |
| P35611 | ADD1 | 1.436205 | 8.364787 | 1.320335 | 0.194962 | 0.994456 | -4.50502 |
| Q5HYA8 | TMEM67 | 1.063361 | 9.59421 | 1.311641 | 0.19505 | 0.994456 | -4.54035 |
| P30405 | PPIF | -1.02617 | 7.967455 | -1.31829 | 0.195871 | 0.994456 | -4.50662 |
| P00918 | CA2 | -0.8437 | 9.133649 | -1.30759 | 0.196614 | 0.994456 | -4.53333 |
| P48506 | GCLC | 0.942678 | 8.866158 | 1.305563 | 0.197096 | 0.994456 | -4.54503 |
| Q12860 | CNTN1 | 0.859114 | 11.73525 | 1.303933 | 0.197647 | 0.994456 | -4.54629 |
| P00740 | F9 | 0.753819 | 11.14519 | 1.298998 | 0.199324 | 0.994456 | -4.55007 |
| Q01484 | ANK2 | -1.04193 | 9.391498 | -1.29876 | 0.199504 | 0.994456 | -4.55028 |
| P08133 | ANXA6 | -0.5642 | 11.75849 | -1.29754 | 0.199819 | 0.994456 | -4.55118 |
| A0A0B4J1V | #N/D | -0.92156 | 12.65679 | -1.2944 | 0.200894 | 0.994456 | -4.55358 |
| A0A0B4J1Y | #N/D | -0.96286 | 9.589963 | -1.2928 | 0.202217 | 0.994456 | -4.54427 |
| P62826 | RAN | 0.616687 | 12.49633 | 1.289687 | 0.202516 | 0.994456 | -4.55717 |
| O95394 | PGM3 | -1.02243 | 8.458672 | -1.28608 | 0.203862 | 0.994456 | -4.54894 |
| A0A0B4J1U | #N/D | -0.9439 | 9.305291 | -1.28407 | 0.205358 | 0.994456 | -4.54249 |
| Q969P0 | IGSF8 | 0.76593 | 10.87703 | 1.27315 | 0.20828 | 0.994456 | -4.56967 |
| Q7L099-4 | #N/D | -0.78025 | 10.43573 | -1.27103 | 0.209026 | 0.994456 | -4.57125 |
| P55056 | APOC4 | -1.10466 | 9.255911 | -1.27146 | 0.210494 | 0.994456 | -4.54162 |
| P01591 | JCHAIN | 0.739126 | 14.17635 | 1.264924 | 0.211192 | 0.994456 | -4.57583 |
| P06748 | NPM1 | 1.722876 | 9.025254 | 1.263574 | 0.21345 | 0.994456 | -4.53438 |
| P05455 | SSB | -1.03876 | 10.60567 | -1.25661 | 0.214262 | 0.994456 | -4.58203 |
| P30626 | SRI | 1.121183 | 7.800031 | 1.26323 | 0.214322 | 0.994456 | -4.54077 |
| P50990 | CCT8 | -0.59851 | 13.71428 | -1.24887 | 0.216964 | 0.994456 | -4.58775 |
| P32004 | L1CAM | -0.79286 | 9.227821 | -1.25042 | 0.217034 | 0.994456 | -4.58669 |
| P04843 | RPN1 | -0.77078 | 8.447986 | -1.2483 | 0.217467 | 0.994456 | -4.56657 |
| P01859 | #N/D | -0.99392 | 13.5427 | -1.24441 | 0.218586 | 0.994456 | -4.59103 |
| Q8IXJ6 | SIRT2 | -1.22149 | 11.31293 | -1.24368 | 0.21915 | 0.994456 | -4.5916 |
| P07954 | FH | 1.00189 | 11.84092 | 1.240615 | 0.220071 | 0.994456 | -4.59383 |
| P07196 | NEFL | 0.474549 | 12.62642 | 1.234751 | 0.222133 | 0.994456 | -4.59811 |

| | | | | | | | |
|---|---|---|---|---|---|---|---|
| O00425 | IGF2BP3 | -1.00162 | 8.291206 | -1.22669 | 0.226704 | 0.994456 | -4.56931 |
| A0A0C4DH | #N/D | -1.02418 | 14.43238 | -1.21913 | 0.228153 | 0.994456 | -4.60947 |
| P02511 | CRYAB | -0.57452 | 10.8827 | -1.20723 | 0.232473 | 0.994456 | -4.61801 |
| P14649 | MYL6B | 0.651201 | 11.20495 | 1.20639 | 0.232793 | 0.994456 | -4.6186 |
| Q99798 | ACO2 | 0.647103 | 13.35696 | 1.204773 | 0.233411 | 0.994456 | -4.61976 |
| P14618-2 | #N/D | -0.99454 | 11.99726 | -1.20327 | 0.233988 | 0.994456 | -4.62083 |
| P02792 | FTL | -0.82963 | 9.643032 | -1.20229 | 0.234656 | 0.994456 | -4.62154 |
| P21283 | ATP6V1C1 | -1.06892 | 9.48756 | -1.19806 | 0.236182 | 0.994456 | -4.62454 |
| P06703 | S100A6 | 1.371616 | 9.228704 | 1.194031 | 0.237552 | 0.994456 | -4.6274 |
| P36955 | SERPINF1 | 0.703724 | 12.75803 | 1.185519 | 0.240871 | 0.994456 | -4.6334 |
| Q15485 | FCN2 | -1.1727 | 12.63067 | -1.1802 | 0.242961 | 0.994456 | -4.63713 |
| P04350 | TUBB4A | 0.806209 | 11.70512 | 1.17591 | 0.244658 | 0.994456 | -4.64013 |
| P0DP09 | #N/D | -0.912 | 13.65182 | -1.16707 | 0.248566 | 0.994456 | -4.63112 |
| Q16798 | ME3 | 0.68129 | 8.140939 | 1.16406 | 0.249879 | 0.994456 | -4.6331 |
| Q86YZ3 | HRNR | 1.264354 | 10.14946 | 1.161191 | 0.251033 | 0.994456 | -4.623 |
| Q92804 | TAF15 | 1.013076 | 9.172634 | 1.160452 | 0.251555 | 0.994456 | -4.60865 |
| P01703 | #N/D | 1.020749 | 13.56328 | 1.156521 | 0.25243 | 0.994456 | -4.65356 |
| P04196 | HRG | -1.19813 | 14.14042 | -1.15048 | 0.254886 | 0.994456 | -4.6577 |
| P30050 | RPL12 | 0.986129 | 10.32925 | 1.150465 | 0.254893 | 0.994456 | -4.65771 |
| P05109 | S100A8 | -0.94525 | 9.04775 | -1.15019 | 0.255286 | 0.994456 | -4.65787 |
| Q14152 | EIF3A | 0.937669 | 8.131085 | 1.151538 | 0.256448 | 0.994456 | -4.62893 |
| P20916 | MAG | -0.82999 | 9.172305 | -1.14088 | 0.259419 | 0.994456 | -4.64818 |
| P35858 | IGFALS | 0.781273 | 11.34869 | 1.134002 | 0.261676 | 0.994456 | -4.6689 |
| P63010 | AP2B1 | 0.841806 | 10.0117 | 1.131213 | 0.262837 | 0.994456 | -4.67078 |
| P14625 | HSP90B1 | 1.137789 | 13.43176 | 1.130202 | 0.263259 | 0.994456 | -4.67146 |
| Q15334 | LLGL1 | -0.78906 | 8.598936 | -1.12892 | 0.263797 | 0.994456 | -4.67232 |
| Q9UJU6 | DBNL | -0.79391 | 10.24301 | -1.12071 | 0.267333 | 0.994456 | -4.6778 |
| Q15365 | PCBP1 | 0.859725 | 11.13094 | 1.118223 | 0.268476 | 0.994456 | -4.67944 |
| Q99878 | H2AC14 | 1.021979 | 8.686371 | 1.114624 | 0.27075 | 0.994456 | -4.64412 |
| P07108 | DBI | 0.735512 | 7.299068 | 1.114997 | 0.270986 | 0.994456 | -4.65126 |
| Q15435 | PPP1R7 | -0.95415 | 8.920446 | -1.1114 | 0.271774 | 0.994456 | -4.66696 |

| | | | | | | | |
|---|---|---|---|---|---|---|---|
| P07360 | C8G | 1.200658 | 8.367863 | 1.113388 | 0.272286 | 0.994456 | -4.60754 |
| P68871 | HBB | -0.91697 | 14.57697 | -1.10878 | 0.272313 | 0.994456 | -4.68574 |
| P16949 | STMN1 | 0.810938 | 11.41839 | 1.107793 | 0.272736 | 0.994456 | -4.68639 |
| Q9Y2J8 | PADI2 | -0.99263 | 10.26827 | -1.10766 | 0.272791 | 0.994456 | -4.68647 |
| P35268 | RPL22 | 0.629059 | 11.96432 | 1.107643 | 0.272801 | 0.994456 | -4.68649 |
| Q14624-3 | #N/D | 1.944328 | 8.418841 | 1.113585 | 0.273404 | 0.994456 | -4.5837 |
| P49913 | CAMP | 0.797441 | 8.94521 | 1.108377 | 0.273665 | 0.994456 | -4.65525 |
| Q16555 | DPYSL2 | -0.54152 | 14.86964 | -1.10329 | 0.274668 | 0.994456 | -4.68935 |
| Q16653-3 | #N/D | 0.761685 | 9.748252 | 1.101882 | 0.275276 | 0.994456 | -4.69028 |
| P61088 | UBE2N | -0.82113 | 11.45261 | -1.09779 | 0.277219 | 0.994456 | -4.69292 |
| P27824 | CANX | 0.851152 | 11.05687 | 1.089496 | 0.28065 | 0.994456 | -4.69837 |
| P09429 | HMGB1 | 0.523005 | 11.33429 | 1.076226 | 0.28649 | 0.994456 | -4.70694 |
| P53634 | CTSC | 0.857803 | 8.483897 | 1.070831 | 0.289555 | 0.994456 | -4.66926 |
| P36871 | PGM1 | 0.605865 | 11.50669 | 1.066461 | 0.29084 | 0.994456 | -4.71319 |
| P54652 | HSPA2 | 0.609361 | 13.29815 | 1.059319 | 0.29405 | 0.994456 | -4.71772 |
| P13645 | KRT10 | -0.6259 | 15.33348 | -1.05811 | 0.294596 | 0.994456 | -4.71849 |
| P60880 | SNAP25 | 0.710892 | 10.94386 | 1.056966 | 0.295113 | 0.994456 | -4.71921 |
| P38646 | HSPA9 | 0.501028 | 14.21459 | 1.055114 | 0.295952 | 0.994456 | -4.72038 |
| P00441 | SOD1 | 0.714377 | 11.21279 | 1.053823 | 0.296621 | 0.994456 | -4.72117 |
| Q04837 | SSBP1 | 0.953766 | 9.844274 | 1.0528 | 0.297085 | 0.994456 | -4.70306 |
| Q14103 | HNRNPD | -1.08834 | 10.83965 | -1.05108 | 0.297782 | 0.994456 | -4.72292 |
| P07741 | APRT | -1.12539 | 8.592073 | -1.04646 | 0.300546 | 0.994456 | -4.72562 |
| P34932 | HSPA4 | -0.56221 | 12.19409 | -1.03942 | 0.303125 | 0.994456 | -4.73021 |
| Q92597 | NDRG1 | -0.69109 | 10.27498 | -1.03461 | 0.305515 | 0.994456 | -4.73314 |
| P35542 | SAA4 | -0.52953 | 12.48858 | -1.02844 | 0.308209 | 0.994456 | -4.73699 |
| P10768 | ESD | 0.742035 | 10.75161 | 1.023771 | 0.310392 | 0.994456 | -4.73986 |
| P26639 | TARS1 | 0.611175 | 9.377104 | 1.0227 | 0.310975 | 0.994456 | -4.74049 |
| P02768 | ALB | 0.879143 | 17.61783 | 1.016537 | 0.313792 | 0.994456 | -4.74428 |
| Q14520 | HABP2 | -0.92237 | 9.292678 | -1.01511 | 0.314717 | 0.994456 | -4.7252 |
| P49207 | RPL34 | -0.84633 | 9.206154 | -1.01253 | 0.315765 | 0.994456 | -4.72674 |
| Q92945 | KHSRP | 0.722857 | 11.19284 | 1.004983 | 0.319273 | 0.994456 | -4.75127 |

| | | | | | | | |
|---|---|---|---|---|---|---|---|
| P02743 | APCS | -1.41173 | 14.22881 | -1.00459 | 0.31946 | 0.994456 | -4.75151 |
| P01706 | #N/D | -0.59856 | 8.121541 | -1.0048 | 0.320202 | 0.994456 | -4.73094 |
| P26599 | PTBP1 | -0.50961 | 10.40181 | -0.99856 | 0.322347 | 0.994456 | -4.75513 |
| P00747 | PLG | 0.625441 | 12.64273 | 0.997633 | 0.322794 | 0.994456 | -4.75568 |
| P62424 | RPL7A | -0.40437 | 11.51419 | -0.99419 | 0.32445 | 0.994456 | -4.75774 |
| A0A075B6I | #N/D | -1.09176 | 11.3767 | -0.99393 | 0.324825 | 0.994456 | -4.73733 |
| Q01082 | SPTBN1 | 0.39023 | 11.78071 | 0.99328 | 0.324891 | 0.994456 | -4.75828 |
| P36354 | #N/D | 1.061388 | 8.62113 | 0.993761 | 0.325846 | 0.994456 | -4.68249 |
| P25789 | PSMA4 | -0.66442 | 7.883878 | -0.99094 | 0.326642 | 0.994456 | -4.72216 |
| P0DOX2 | #N/D | 0.396073 | 13.86052 | 0.988108 | 0.327395 | 0.994456 | -4.76135 |
| Q14974 | KPNB1 | 1.10434 | 12.05301 | 0.98195 | 0.330392 | 0.994456 | -4.76499 |
| P51659 | HSD17B4 | -0.49761 | 12.08701 | -0.98136 | 0.330682 | 0.994456 | -4.76534 |
| P20742 | #N/D | 0.693207 | 13.66452 | 0.979786 | 0.33145 | 0.994456 | -4.76626 |
| P78559 | MAP1A | 0.686612 | 10.07764 | 0.977998 | 0.332568 | 0.994456 | -4.7463 |
| P08237-3 | #N/D | 0.49828 | 9.903447 | 0.975136 | 0.333972 | 0.994456 | -4.76889 |
| P19338 | NCL | 0.712919 | 14.24 | 0.974477 | 0.334054 | 0.994456 | -4.76937 |
| Q12765 | SCRN1 | -0.40944 | 11.08891 | -0.97382 | 0.334376 | 0.994456 | -4.76975 |
| P06454 | PTMA | -0.72602 | 9.310123 | -0.97301 | 0.334777 | 0.994456 | -4.77023 |
| Q9BVA1 | TUBB2B | -0.90829 | 10.28254 | -0.9728 | 0.334879 | 0.994456 | -4.77035 |
| P00505 | GOT2 | 0.515523 | 12.49377 | 0.970375 | 0.336076 | 0.994456 | -4.77177 |
| P08708 | RPS17 | -0.66383 | 7.699005 | -0.96733 | 0.338286 | 0.994456 | -4.72485 |
| P05387 | RPLP2 | -0.59003 | 9.837368 | -0.96597 | 0.338333 | 0.994456 | -4.77429 |
| P68104 | EEF1A1 | 0.678085 | 9.086982 | 0.965702 | 0.338466 | 0.994456 | -4.77445 |
| O60506 | SYNCRIP | 0.667147 | 8.844164 | 0.96589 | 0.338803 | 0.994456 | -4.77416 |
| P07197 | NEFM | -0.40921 | 12.44543 | -0.96399 | 0.339238 | 0.994456 | -4.77547 |
| P01742 | #N/D | 0.670416 | 13.16148 | 0.964835 | 0.339971 | 0.994456 | -4.75325 |
| P63000 | RAC1 | -0.65153 | 11.00234 | -0.95887 | 0.341788 | 0.994456 | -4.77843 |
| P60981 | DSTN | 0.677703 | 11.21887 | 0.954161 | 0.344222 | 0.994456 | -4.7811 |
| Q9BWD1-2 | #N/D | 0.791053 | 7.847079 | 0.956761 | 0.34424 | 0.994456 | -4.69946 |
| P06681 | C2 | -0.6121 | 10.88017 | -0.95408 | 0.344422 | 0.994456 | -4.7595 |
| P50993 | ATP1A2 | 0.816057 | 10.45463 | 0.95269 | 0.34496 | 0.994456 | -4.78194 |

| | | | | | | | |
|---|---|---|---|---|---|---|---|
| P51149 | RAB7A | 0.470455 | 8.780581 | 0.950953 | 0.345993 | 0.994456 | -4.78287 |
| P01019 | AGT | -0.77819 | 13.11805 | -0.94979 | 0.346341 | 0.994456 | -4.78363 |
| P31948 | STIP1 | 0.446343 | 10.97486 | 0.948532 | 0.346977 | 0.994456 | -4.78435 |
| Q7L0J3 | SV2A | -0.99729 | 9.396699 | -0.94291 | 0.35133 | 0.994456 | -4.7236 |
| P61626 | LYZ | -0.74405 | 13.1144 | -0.93794 | 0.352348 | 0.994456 | -4.79035 |
| P07339 | CTSD | 0.727966 | 12.19536 | 0.937544 | 0.352547 | 0.994456 | -4.79057 |
| P49753 | ACOT2 | 0.640336 | 10.06568 | 0.932575 | 0.355085 | 0.994456 | -4.79337 |
| P25311 | AZGP1 | -0.48079 | 12.1082 | -0.93171 | 0.355529 | 0.994456 | -4.79385 |
| Q9Y4L1 | HYOU1 | 0.55683 | 8.672707 | 0.930689 | 0.356125 | 0.994456 | -4.79439 |
| P63241 | EIF5A | -0.6837 | 8.213411 | -0.92872 | 0.35764 | 0.994456 | -4.75472 |
| P52272 | HNRNPM | 0.524499 | 12.38158 | 0.925067 | 0.358942 | 0.994456 | -4.79756 |
| P05783 | KRT18 | -0.82624 | 11.19722 | -0.92312 | 0.360097 | 0.994456 | -4.79857 |
| P14415 | ATP1B2 | -0.72746 | 8.426782 | -0.9207 | 0.36159 | 0.994456 | -4.73447 |
| P29966 | MARCKS | -0.73565 | 12.49708 | -0.9175 | 0.362854 | 0.994456 | -4.80175 |
| P10636-5 | #N/D | 0.40882 | 11.53737 | 0.91383 | 0.364765 | 0.994456 | -4.80377 |
| P02538 | KRT6A | -1.01743 | 11.77848 | -0.91349 | 0.365169 | 0.994456 | -4.78119 |
| P01594 | #N/D | -0.66726 | 13.40637 | -0.91234 | 0.365933 | 0.994456 | -4.78172 |
| O14594 | NCAN | 0.581536 | 9.186149 | 0.911221 | 0.366515 | 0.994456 | -4.7636 |
| P23142 | FBLN1 | -0.54193 | 9.472544 | -0.91062 | 0.366746 | 0.994456 | -4.78266 |
| Q9NUQ9 | CYRIB | 0.725663 | 9.681427 | 0.902812 | 0.370756 | 0.994456 | -4.80969 |
| Q13885 | TUBB2A | 0.652727 | 12.3616 | 0.902204 | 0.370853 | 0.994456 | -4.81013 |
| P13611 | VCAN | 0.340509 | 12.41909 | 0.901098 | 0.371436 | 0.994456 | -4.81073 |
| P10809 | HSPD1 | 0.400313 | 14.28311 | 0.901029 | 0.371472 | 0.994456 | -4.81077 |
| O75891 | ALDH1L1 | 0.682675 | 12.21429 | 0.900418 | 0.371794 | 0.994456 | -4.8111 |
| P22314 | UBA1 | 0.598242 | 12.03395 | 0.898751 | 0.372674 | 0.994456 | -4.812 |
| P42704 | LRPPRC | 0.827259 | 13.36178 | 0.897416 | 0.37338 | 0.994456 | -4.81272 |
| P50502 | ST13 | -0.73446 | 12.29466 | -0.89359 | 0.375628 | 0.994456 | -4.81468 |
| A0A0J9YXX | #N/D | 0.688201 | 10.65969 | 0.891997 | 0.376251 | 0.994456 | -4.81564 |
| P0DP25 | CALM1 | -1.21657 | 11.85239 | -0.89195 | 0.376276 | 0.994456 | -4.81567 |
| P04003 | C4BPA | 0.491222 | 14.39715 | 0.885823 | 0.379541 | 0.994456 | -4.81895 |
| Q86V81 | ALYREF | 0.858043 | 8.628483 | 0.885916 | 0.379869 | 0.994456 | -4.79534 |

| | | | | | | | |
|---|---|---|---|---|---|---|---|
| P20700 | LMNB1 | -0.67892 | 10.71505 | -0.88473 | 0.380124 | 0.994456 | -4.81953 |
| P23471 | PTPRZ1 | 1.197283 | 10.95353 | 0.877539 | 0.384124 | 0.994456 | -4.82328 |
| P28838 | LAP3 | 0.518444 | 11.12429 | 0.876481 | 0.384553 | 0.994456 | -4.82391 |
| P02533 | KRT14 | -0.69426 | 11.09842 | -0.87507 | 0.385315 | 0.994456 | -4.82465 |
| P55786 | NPEPPS | 0.538442 | 9.711469 | 0.874733 | 0.385495 | 0.994456 | -4.82483 |
| P61978 | HNRNPK | 0.686136 | 11.47478 | 0.874721 | 0.385501 | 0.994456 | -4.82484 |
| P12270 | TPR | 0.754536 | 9.31589 | 0.871027 | 0.387788 | 0.994456 | -4.82663 |
| P62241 | RPS8 | 0.505734 | 9.619243 | 0.86903 | 0.388579 | 0.994456 | -4.82783 |
| Q9H9Z2 | LIN28A | 0.708181 | 9.50441 | 0.866023 | 0.39028 | 0.994456 | -4.82936 |
| P46821 | MAP1B | 0.511925 | 12.35155 | 0.859694 | 0.393662 | 0.994456 | -4.83269 |
| Q01518 | CAP1 | -0.61071 | 11.32087 | -0.85826 | 0.394448 | 0.994456 | -4.83344 |
| P17600 | SYN1 | 0.656192 | 12.09097 | 0.85698 | 0.395147 | 0.994456 | -4.8341 |
| P02671 | FGA | 0.418613 | 12.54848 | 0.853474 | 0.397071 | 0.994456 | -4.83591 |
| Q99832 | CCT7 | 0.448132 | 11.03507 | 0.847973 | 0.400101 | 0.994456 | -4.83873 |
| P08519 | #N/D | 0.705581 | 15.54961 | 0.846818 | 0.400739 | 0.994456 | -4.83932 |
| Q15366 | PCBP2 | 0.818939 | 12.64187 | 0.846047 | 0.401165 | 0.994456 | -4.83971 |
| P19652 | ORM2 | -0.51038 | 9.423031 | -0.84498 | 0.402113 | 0.994456 | -4.79545 |
| P60866 | RPS20 | 0.620034 | 8.421034 | 0.84445 | 0.402408 | 0.994456 | -4.81594 |
| P01031 | C5 | 0.324944 | 12.98874 | 0.84368 | 0.402476 | 0.994456 | -4.84092 |
| P05154 | SERPINA5 | -0.4879 | 11.45435 | -0.84325 | 0.402715 | 0.994456 | -4.84114 |
| P16152 | CBR1 | -0.47947 | 14.15987 | -0.84107 | 0.403921 | 0.994456 | -4.84225 |
| O75368 | SH3BGRL | -0.78728 | 9.591212 | -0.83915 | 0.405124 | 0.994456 | -4.84315 |
| Q15149 | PLEC | -0.37029 | 12.56068 | -0.83855 | 0.405326 | 0.994456 | -4.84353 |
| P48147 | PREP | -0.65952 | 7.680943 | -0.83844 | 0.406484 | 0.994456 | -4.77099 |
| A0A075B6I | #N/D | 0.81057 | 8.760973 | 0.833395 | 0.408553 | 0.994456 | -4.84594 |
| Q15424 | SAFB | -0.49965 | 10.29881 | -0.83132 | 0.409429 | 0.994456 | -4.84714 |
| P35580 | MYH10 | 0.615592 | 9.704816 | 0.830826 | 0.409637 | 0.994456 | -4.84742 |
| P62851 | RPS25 | 0.599909 | 10.64803 | 0.82984 | 0.41019 | 0.994456 | -4.84791 |
| P02741 | CRP | -1.07718 | 11.11435 | -0.82967 | 0.410348 | 0.994456 | -4.82319 |
| P35579 | MYH9 | -0.64023 | 12.21283 | -0.82914 | 0.410584 | 0.994456 | -4.84826 |
| Q04695 | KRT17 | -0.61801 | 10.32094 | -0.82945 | 0.410612 | 0.994456 | -4.82323 |

| | | | | | | | |
|---|---|---|---|---|---|---|---|
| P11940 | PABPC1 | 0.51734 | 9.661148 | 0.827102 | 0.411859 | 0.994456 | -4.84921 |
| Q9BY11 | PACSIN1 | -0.73955 | 11.03934 | -0.82144 | 0.414916 | 0.994456 | -4.8521 |
| A0A0B4J1V | #N/D | 0.922633 | 8.761429 | 0.823282 | 0.41518 | 0.994456 | -4.77732 |
| P06396 | GSN | -0.5161 | 14.7431 | -0.81995 | 0.415757 | 0.994456 | -4.85284 |
| Q6YN16 | HSDL2 | 0.738145 | 9.242755 | 0.819124 | 0.416496 | 0.994456 | -4.8531 |
| P09622 | DLD | 0.565393 | 8.688977 | 0.818207 | 0.417607 | 0.994456 | -4.82822 |
| P21796 | VDAC1 | 0.715339 | 8.076572 | 0.818498 | 0.417648 | 0.994456 | -4.8072 |
| Q08380 | LGALS3BP | -0.49236 | 14.57471 | -0.81455 | 0.418818 | 0.994456 | -4.85551 |
| Q9H3S7 | PTPN23 | 0.457556 | 9.197781 | 0.812092 | 0.420214 | 0.994456 | -4.85671 |
| P13637 | ATP1A3 | 0.631469 | 12.48297 | 0.811101 | 0.420778 | 0.994456 | -4.8572 |
| P30153 | PPP2R1A | 0.635318 | 10.68953 | 0.810158 | 0.421315 | 0.994456 | -4.85766 |
| Q9NZL9 | MAT2B | 0.685671 | 9.370227 | 0.809715 | 0.421631 | 0.994456 | -4.85784 |
| P40926 | MDH2 | -0.40013 | 13.17949 | -0.79372 | 0.430745 | 0.994456 | -4.86563 |
| P61970 | NUTF2 | -0.6228 | 8.594163 | -0.79351 | 0.430927 | 0.994456 | -4.84009 |
| Q9H4G0 | EPB41L1 | -0.52068 | 10.09245 | -0.79323 | 0.431025 | 0.994456 | -4.86586 |
| P62266 | RPS23 | 0.557414 | 9.096047 | 0.793707 | 0.431086 | 0.994456 | -4.86545 |
| P35527 | KRT9 | -0.53039 | 15.65986 | -0.79214 | 0.431659 | 0.994456 | -4.86639 |
| O43426 | SYNJ1 | -1.65742 | 13.74991 | -0.79205 | 0.431764 | 0.994456 | -4.8664 |
| Q9NRX4 | PHPT1 | -1.00459 | 8.126823 | -0.79443 | 0.43209 | 0.994456 | -4.683 |
| P01611 | #N/D | -0.67711 | 10.90728 | -0.78841 | 0.434463 | 0.994456 | -4.8421 |
| P20671 | H2AC7 | -0.65685 | 8.62645 | -0.78714 | 0.435476 | 0.994456 | -4.79233 |
| P0DMV9 | HSPA1A | 0.344323 | 12.81602 | 0.782996 | 0.436963 | 0.994456 | -4.87074 |
| P07357 | C8A | -0.4426 | 14.30403 | -0.78295 | 0.43699 | 0.994456 | -4.87077 |
| P0DJI9 | SAA2 | -0.68648 | 8.735337 | -0.78106 | 0.438649 | 0.994456 | -4.84547 |
| A0A075B6I | #N/D | 0.433841 | 14.94383 | 0.77989 | 0.438774 | 0.994456 | -4.87221 |
| P07358 | C8B | 0.513778 | 13.46551 | 0.774282 | 0.442055 | 0.994456 | -4.87485 |
| Q9Y2W1 | THRAP3 | 0.520282 | 8.282812 | 0.773514 | 0.44276 | 0.994456 | -4.84901 |
| P80723 | BASP1 | -0.68937 | 10.51359 | -0.77223 | 0.443262 | 0.994456 | -4.87581 |
| P62081 | RPS7 | 0.474876 | 10.42682 | 0.771061 | 0.443946 | 0.994456 | -4.87636 |
| Q15121 | PEA15 | 0.37368 | 10.46576 | 0.769065 | 0.44512 | 0.994456 | -4.87729 |
| Q16623-3 | #N/D | 0.393189 | 9.890803 | 0.767975 | 0.445762 | 0.994456 | -4.87779 |

| | | | | | | | |
|---|---|---|---|---|---|---|---|
| Q04637 | EIF4G1 | 0.574208 | 9.520493 | 0.767816 | 0.446042 | 0.994456 | -4.87776 |
| P13647 | KRT5 | -0.55556 | 13.1342 | -0.76644 | 0.446668 | 0.994456 | -4.87851 |
| O76070 | SNCG | -0.40614 | 10.89826 | -0.76638 | 0.4467 | 0.994456 | -4.87853 |
| P00367 | GLUD1 | 0.402803 | 13.41429 | 0.761001 | 0.449883 | 0.994456 | -4.88102 |
| O76054 | SEC14L2 | 0.609493 | 8.396433 | 0.762362 | 0.449966 | 0.994456 | -4.80216 |
| P07996 | THBS1 | 0.35294 | 13.73619 | 0.757 | 0.452257 | 0.994456 | -4.88286 |
| P14550 | AKR1A1 | 0.527359 | 7.585267 | 0.756252 | 0.453491 | 0.994456 | -4.85633 |
| P07900-2 | #N/D | 0.535889 | 14.11273 | 0.754441 | 0.453779 | 0.994456 | -4.88403 |
| P50453 | SERPINB9 | 0.664729 | 9.530376 | 0.75397 | 0.454445 | 0.994456 | -4.88402 |
| P07237 | P4HB | 0.592826 | 10.09498 | 0.749438 | 0.456944 | 0.994456 | -4.8862 |
| P42766 | RPL35 | -0.43943 | 9.474201 | -0.74915 | 0.456993 | 0.994456 | -4.88641 |
| P0C0L4 | C4A | -0.76811 | 11.8147 | -0.74279 | 0.461276 | 0.994456 | -4.8623 |
| P08670 | VIM | -0.29473 | 13.17483 | -0.74161 | 0.461458 | 0.994456 | -4.88985 |
| P60953 | CDC42 | -0.62303 | 9.514288 | -0.74028 | 0.462499 | 0.994456 | -4.86353 |
| A0A0C4DH | #N/D | 0.407638 | 12.25946 | 0.738775 | 0.463161 | 0.994456 | -4.89112 |
| P02655 | APOC2 | 0.531438 | 11.22537 | 0.735773 | 0.465088 | 0.994456 | -4.89239 |
| P05388 | RPLP0 | -0.43347 | 11.63871 | -0.73535 | 0.465228 | 0.994456 | -4.89265 |
| Q9HC38 | GLOD4 | 0.539869 | 9.301142 | 0.734148 | 0.465954 | 0.994456 | -4.89318 |
| Q93050 | ATP6V0A1 | 0.35994 | 10.19313 | 0.73296 | 0.466672 | 0.994456 | -4.89371 |
| P07437 | TUBB | -0.58042 | 12.82403 | -0.7324 | 0.467009 | 0.994456 | -4.89396 |
| P11166 | SLC2A1 | 0.57187 | 9.8587 | 0.731668 | 0.467454 | 0.994456 | -4.89428 |
| P48426 | PIP4K2A | 0.510741 | 8.06123 | 0.732439 | 0.468234 | 0.994456 | -4.77031 |
| Q04917 | YWHAH | 0.530324 | 11.45903 | 0.72347 | 0.472434 | 0.994456 | -4.8979 |
| P01780 | #N/D | -0.60388 | 11.11587 | -0.71542 | 0.477356 | 0.994456 | -4.90141 |
| P09211 | GSTP1 | 0.484284 | 13.54278 | 0.715145 | 0.47818 | 0.994456 | -4.87385 |
| P11586 | MTHFD1 | 1.388621 | 11.82919 | 0.713631 | 0.478498 | 0.994456 | -4.90216 |
| P07225 | PROS1 | 0.306089 | 13.16055 | 0.710329 | 0.480479 | 0.994456 | -4.90361 |
| Q02543 | RPL18A | -0.70802 | 12.91705 | -0.70835 | 0.481699 | 0.994456 | -4.90447 |
| Q9P2D7-8 | #N/D | 0.605659 | 8.669631 | 0.704629 | 0.483992 | 0.994456 | -4.90606 |
| O14818 | PSMA7 | -0.42443 | 8.736898 | -0.70058 | 0.486553 | 0.994456 | -4.90776 |
| Q16799 | RTN1 | -0.49229 | 10.36556 | -0.69941 | 0.487221 | 0.994456 | -4.90828 |

| | | | | | | | |
|---|---|---|---|---|---|---|---|
| P00403 | COX2 | 0.479732 | 8.386699 | 0.698962 | 0.48822 | 0.994456 | -4.82596 |
| P62277 | RPS13 | -0.4219 | 10.17611 | -0.69697 | 0.488733 | 0.994456 | -4.90932 |
| P62805 | H4C9 | 0.540935 | 11.74498 | 0.696051 | 0.489306 | 0.994456 | -4.90971 |
| P01024 | C3 | 0.42771 | 15.31739 | 0.695928 | 0.489383 | 0.994456 | -4.90976 |
| P35080 | PFN2 | 0.514294 | 10.95784 | 0.695973 | 0.489408 | 0.994456 | -4.90971 |
| O00187 | MASP2 | 0.60417 | 13.7813 | 0.695938 | 0.489429 | 0.994456 | -4.90972 |
| P13010 | XRCC5 | 0.3752 | 12.2267 | 0.695746 | 0.489495 | 0.994456 | -4.90983 |
| P0DJI8 | SAA1 | 0.54315 | 12.87564 | 0.695133 | 0.489877 | 0.994456 | -4.91009 |
| P39748 | FEN1 | -0.84728 | 9.947938 | -0.69238 | 0.491591 | 0.994456 | -4.91125 |
| P09471 | GNAO1 | -0.39513 | 11.83785 | -0.69225 | 0.49167 | 0.994456 | -4.91131 |
| P09417 | QDPR | -0.50249 | 11.79198 | -0.68918 | 0.493587 | 0.994456 | -4.9126 |
| P24539 | ATP5PB | 0.723362 | 9.276826 | 0.683614 | 0.497546 | 0.994456 | -4.83152 |
| Q99880 | H2BC13 | 0.497379 | 12.3185 | 0.680936 | 0.498747 | 0.994456 | -4.91602 |
| P69905 | HBA1 | -0.34182 | 14.01226 | -0.67658 | 0.501488 | 0.994456 | -4.91782 |
| P08779 | KRT16 | -1.05856 | 8.523877 | -0.67799 | 0.502312 | 0.994456 | -4.70985 |
| P15169 | CPN1 | 0.555255 | 9.464541 | 0.674325 | 0.503068 | 0.994456 | -4.89053 |
| A0A0G2JS( | #N/D | -0.76307 | 9.269116 | -0.67028 | 0.506076 | 0.994456 | -4.86816 |
| P30040 | ERP29 | -0.72871 | 8.785068 | -0.66992 | 0.506455 | 0.994456 | -4.86821 |
| P78417 | GSTO1 | 0.516329 | 11.81208 | 0.667222 | 0.5074 | 0.994456 | -4.92164 |
| Q00765 | REEP5 | -0.47923 | 8.66685 | -0.66766 | 0.507588 | 0.994456 | -4.89294 |
| P30038 | ALDH4A1 | 0.345084 | 11.58862 | 0.666797 | 0.507669 | 0.994456 | -4.92181 |
| P17844 | DDX5 | 0.302053 | 11.72587 | 0.664956 | 0.508837 | 0.994456 | -4.92256 |
| A0A075B6I | #N/D | 0.768618 | 7.939523 | 0.664537 | 0.510651 | 0.994456 | -4.7478 |
| P20851-2 | #N/D | 0.510378 | 11.52893 | 0.660806 | 0.511475 | 0.994456 | -4.92423 |
| P48735 | IDH2 | 0.436418 | 11.01398 | 0.658868 | 0.512709 | 0.994456 | -4.925 |
| Q14195-2 | #N/D | 0.337585 | 12.44951 | 0.657291 | 0.513715 | 0.994456 | -4.92564 |
| P08603 | CFH | 0.372422 | 14.2247 | 0.651806 | 0.517221 | 0.994456 | -4.92782 |
| P01718 | #N/D | -0.43068 | 12.55539 | -0.65166 | 0.517316 | 0.994456 | -4.92788 |
| Q01813 | PFKP | -0.3562 | 10.6556 | -0.65065 | 0.517963 | 0.994456 | -4.92828 |
| Q92598-4 | #N/D | 0.814063 | 8.188924 | 0.651211 | 0.518689 | 0.994456 | -4.75129 |
| P02749 | APOH | -0.33495 | 12.64195 | -0.64709 | 0.520243 | 0.994456 | -4.92968 |

| | | | | | | | |
|---|---|---|---|---|---|---|---|
| P13797 | PLS3 | -0.41535 | 9.880516 | -0.64704 | 0.520279 | 0.994456 | -4.9297 |
| P07195 | LDHB | 0.290146 | 13.41831 | 0.644216 | 0.522093 | 0.994456 | -4.93081 |
| P21926 | CD9 | 0.507724 | 10.94564 | 0.643783 | 0.522372 | 0.994456 | -4.93098 |
| A0A0C4DH | #N/D | 0.518107 | 11.05438 | 0.641094 | 0.524104 | 0.994456 | -4.93203 |
| P05090 | APOD | 0.360168 | 14.6869 | 0.63962 | 0.525055 | 0.994456 | -4.9326 |
| A0A0C4DH | #N/D | -0.61143 | 10.49513 | -0.63806 | 0.526267 | 0.994456 | -4.90428 |
| P10643 | C7 | -0.31452 | 12.83675 | -0.63743 | 0.526472 | 0.994456 | -4.93345 |
| P09543 | CNP | -0.2669 | 12.18182 | -0.63718 | 0.526631 | 0.994456 | -4.93355 |
| Q15848 | ADIPOQ | 0.43638 | 10.82374 | 0.627609 | 0.532838 | 0.994456 | -4.93723 |
| Q14194 | CRMP1 | 0.3192 | 11.68612 | 0.625523 | 0.534196 | 0.994456 | -4.93803 |
| P01700 | #N/D | 0.640293 | 10.59788 | 0.625331 | 0.534368 | 0.994456 | -4.93807 |
| P09104 | ENO2 | 0.338653 | 11.61892 | 0.624629 | 0.534778 | 0.994456 | -4.93836 |
| P63162 | SNRPN | -1.21891 | 6.935315 | -0.62828 | 0.537142 | 0.994456 | -4.70655 |
| P62942 | FKBP1A | 0.603494 | 10.83055 | 0.61902 | 0.53844 | 0.994456 | -4.94049 |
| P37840 | SNCA | 0.508293 | 9.699817 | 0.618346 | 0.538975 | 0.994456 | -4.94068 |
| P12956 | XRCC6 | 0.444933 | 12.11524 | 0.615558 | 0.540706 | 0.994456 | -4.94179 |
| P20073 | ANXA7 | -0.34215 | 8.552518 | -0.61556 | 0.540753 | 0.994456 | -4.94176 |
| P63244 | RACK1 | -0.40614 | 12.16177 | -0.61304 | 0.542357 | 0.994456 | -4.94273 |
| P04275 | VWF | -0.4585 | 14.63316 | -0.61279 | 0.542522 | 0.994456 | -4.94282 |
| P29762 | CRABP1 | -0.63357 | 11.46855 | -0.6117 | 0.54324 | 0.994456 | -4.94323 |
| P62888 | RPL30 | -0.44363 | 9.168351 | -0.60729 | 0.546188 | 0.994456 | -4.91547 |
| P53004 | BLVRA | 0.467116 | 7.896539 | 0.606669 | 0.546905 | 0.994456 | -4.91551 |
| P61764 | STXBP1 | 0.403359 | 13.40973 | 0.603632 | 0.548552 | 0.994456 | -4.94621 |
| P07451 | CA3 | 0.522853 | 9.536455 | 0.599636 | 0.551238 | 0.994456 | -4.94765 |
| Q9H115 | NAPB | -0.40077 | 9.074792 | -0.59994 | 0.551344 | 0.994456 | -4.89301 |
| Q96AE4 | FUBP1 | 0.459439 | 9.61624 | 0.59447 | 0.554617 | 0.994456 | -4.94955 |
| Q9UMF0 | ICAM5 | 0.387112 | 9.084601 | 0.594095 | 0.554866 | 0.994456 | -4.94969 |
| P02654 | APOC1 | 0.383031 | 11.84732 | 0.594017 | 0.554962 | 0.994456 | -4.94969 |
| O95782 | AP2A1 | -0.44571 | 8.500907 | -0.59305 | 0.556222 | 0.994456 | -4.89512 |
| P0DOX3 | #N/D | 0.257018 | 13.40279 | 0.590108 | 0.557517 | 0.994456 | -4.95113 |
| Q99536 | VAT1 | 0.345843 | 11.20367 | 0.588546 | 0.558557 | 0.994456 | -4.95169 |

| | | | | | | | |
|---|---|---|---|---|---|---|---|
| P40925 | MDH1 | 0.324162 | 13.09244 | 0.587104 | 0.559518 | 0.994456 | -4.9522 |
| P55084 | HADHB | 0.42515 | 8.238028 | 0.586245 | 0.560551 | 0.994456 | -4.86332 |
| P08185 | SERPINA6 | 0.470624 | 9.106814 | 0.586513 | 0.560962 | 0.994456 | -4.86295 |
| P15814 | IGLL1 | -0.84094 | 11.9045 | -0.5823 | 0.562961 | 0.994456 | -4.92399 |
| P45880 | VDAC2 | 0.316157 | 11.73893 | 0.578364 | 0.565362 | 0.994456 | -4.95531 |
| P62273 | RPS29 | -0.50908 | 8.012306 | -0.57537 | 0.567765 | 0.994456 | -4.83925 |
| P14868-2 | #N/D | -0.40745 | 8.099459 | -0.57451 | 0.568395 | 0.994456 | -4.86684 |
| P61106 | RAB14 | -0.34535 | 9.911827 | -0.5733 | 0.568805 | 0.994456 | -4.95706 |
| Q96FC7 | PHYHIPL | -0.57589 | 10.07371 | -0.57179 | 0.569818 | 0.994456 | -4.95758 |
| P00742 | F10 | -0.4791 | 12.22352 | -0.57105 | 0.570316 | 0.994456 | -4.95784 |
| Q7KZF4 | SND1 | -0.41991 | 9.615375 | -0.57116 | 0.570332 | 0.994456 | -4.92777 |
| Q15084 | PDIA6 | -0.33739 | 12.47948 | -0.57058 | 0.570593 | 0.994456 | -4.95803 |
| O43390 | HNRNPR | 0.355046 | 8.163912 | 0.567047 | 0.573199 | 0.994456 | -4.90371 |
| P10620 | MGST1 | 0.435069 | 10.88042 | 0.563344 | 0.575473 | 0.994456 | -4.96053 |
| O43776 | NARS1 | 0.343579 | 10.11421 | 0.562662 | 0.575934 | 0.994456 | -4.96077 |
| P61247 | RPS3A | 0.245039 | 12.29605 | 0.561423 | 0.576772 | 0.994456 | -4.96119 |
| P50213 | IDH3A | -0.38887 | 10.5341 | -0.56134 | 0.576827 | 0.994456 | -4.96122 |
| Q14204 | DYNC1H1 | 0.270659 | 13.22936 | 0.561083 | 0.577003 | 0.994456 | -4.96131 |
| P48539 | PCP4 | 0.305479 | 8.670542 | 0.559251 | 0.578328 | 0.994456 | -4.96188 |
| P05026 | ATP1B1 | 0.390708 | 9.719583 | 0.557776 | 0.579466 | 0.994456 | -4.9623 |
| Q9Y266 | NUDC | -0.42569 | 9.322208 | -0.55453 | 0.581714 | 0.994456 | -4.96337 |
| Q9Y5K8 | ATP6V1D | -0.55434 | 7.792041 | -0.55419 | 0.58243 | 0.994456 | -4.80333 |
| O43175 | PHGDH | 0.247037 | 13.11601 | 0.549168 | 0.585097 | 0.994456 | -4.96534 |
| Q02878 | RPL6 | 0.471388 | 11.19865 | 0.548246 | 0.585725 | 0.994456 | -4.96565 |
| P04114 | APOB | -0.46896 | 15.2199 | -0.5479 | 0.58596 | 0.994456 | -4.96576 |
| O94760 | DDAH1 | 0.367167 | 9.840467 | 0.547653 | 0.58617 | 0.994456 | -4.96582 |
| P02689 | PMP2 | 0.402994 | 10.41906 | 0.547116 | 0.586496 | 0.994456 | -4.96602 |
| P62861 | #N/D | -0.37532 | 9.145956 | -0.54654 | 0.587062 | 0.994456 | -4.96612 |
| P09661 | SNRPA1 | -0.671 | 11.24558 | -0.54579 | 0.587401 | 0.994456 | -4.96647 |
| P19827 | ITIH1 | -0.23134 | 15.08481 | -0.54201 | 0.589984 | 0.994456 | -4.96772 |
| P45974 | USP5 | 0.311796 | 9.969402 | 0.539119 | 0.591964 | 0.994456 | -4.96867 |

| | | | | | | | |
|---|---|---|---|---|---|---|---|
| P08195 | SLC3A2 | -0.4359 | 10.25962 | -0.5385 | 0.592428 | 0.994456 | -4.93835 |
| P13861 | PRKAR2A | 0.491645 | 8.585476 | 0.531824 | 0.597138 | 0.994456 | -4.89932 |
| P61313 | RPL15 | -0.23486 | 10.74977 | -0.53055 | 0.597852 | 0.994456 | -4.97147 |
| P0C0L5 | C4B | 0.306568 | 12.58969 | 0.529464 | 0.598637 | 0.994456 | -4.9718 |
| P61026 | RAB10 | -0.39033 | 8.602059 | -0.52984 | 0.598922 | 0.994456 | -4.94074 |
| P15104 | GLUL | -0.39052 | 13.57473 | -0.52856 | 0.599219 | 0.994456 | -4.97211 |
| P00558 | PGK1 | -0.22731 | 12.86953 | -0.52379 | 0.602514 | 0.994456 | -4.97364 |
| P01701 | #N/D | -0.45579 | 10.54093 | -0.51817 | 0.606561 | 0.994456 | -4.94453 |
| P61020 | RAB5B | 0.289317 | 8.987505 | 0.517491 | 0.607033 | 0.994456 | -4.97555 |
| Q9P258 | RCC2 | 0.281944 | 11.42629 | 0.514165 | 0.609181 | 0.994456 | -4.97669 |
| O95989 | NUDT3 | 0.392324 | 9.707781 | 0.513143 | 0.609891 | 0.994456 | -4.97701 |
| Q92954 | PRG4 | -0.44089 | 10.51058 | -0.51296 | 0.610017 | 0.994456 | -4.97707 |
| O00154 | ACOT7 | 0.426738 | 10.65194 | 0.512572 | 0.610288 | 0.994456 | -4.97719 |
| P04075 | ALDOA | -0.2447 | 13.09657 | -0.51216 | 0.610574 | 0.994456 | -4.97732 |
| P0DOX5 | #N/D | 0.272864 | 14.50645 | 0.512123 | 0.6106 | 0.994456 | -4.97733 |
| P24752 | ACAT1 | -0.35592 | 10.23426 | -0.51164 | 0.610939 | 0.994456 | -4.97748 |
| P02753 | RBP4 | -0.46924 | 9.614475 | -0.50768 | 0.613846 | 0.994456 | -4.97862 |
| P43487 | RANBP1 | -0.35014 | 11.29012 | -0.50617 | 0.61499 | 0.994456 | -4.97904 |
| P60028 | #N/D | 0.367887 | 9.154595 | 0.50534 | 0.615439 | 0.994456 | -4.94837 |
| P01023 | A2M | -0.36404 | 15.60344 | -0.50425 | 0.616083 | 0.994456 | -4.97977 |
| Q15102 | PAFAH1B3 | -0.41348 | 9.201983 | -0.50294 | 0.617077 | 0.994456 | -4.98013 |
| P25705 | ATP5F1A | -0.50182 | 13.38486 | -0.50187 | 0.617748 | 0.994456 | -4.9805 |
| P04433 | #N/D | -0.43748 | 12.95042 | -0.49867 | 0.619984 | 0.994456 | -4.98148 |
| Q00325 | SLC25A3 | -0.439 | 11.25621 | -0.49722 | 0.621002 | 0.994456 | -4.98192 |
| Q8N163 | CCAR2 | -0.37554 | 8.405871 | -0.49597 | 0.621989 | 0.994456 | -4.9511 |
| A0A0C4DH | #N/D | -0.39852 | 9.100349 | -0.49372 | 0.623493 | 0.994456 | -4.98296 |
| P02656 | APOC3 | -0.37646 | 12.92227 | -0.49204 | 0.624634 | 0.994456 | -4.98349 |
| P53680 | AP2S1 | -0.38259 | 7.833872 | -0.49163 | 0.625517 | 0.994456 | -4.88957 |
| P61981 | YWHAG | 0.441958 | 13.78262 | 0.480034 | 0.633092 | 0.994456 | -4.98705 |
| P21333 | FLNA | 0.284505 | 13.07842 | 0.479833 | 0.633235 | 0.994456 | -4.98711 |
| A0A0A0MS | #N/D | -0.32635 | 10.69979 | -0.47575 | 0.636123 | 0.994456 | -4.9883 |

| | | | | | | | |
|---|---|---|---|---|---|---|---|
| P07737 | PFN1 | 0.455938 | 12.48645 | 0.475216 | 0.636686 | 0.994456 | -4.98835 |
| Q9NR46 | SH3GLB2 | 0.30374 | 8.407993 | 0.474631 | 0.637022 | 0.994456 | -4.98857 |
| Q8NC51 | SERBP1 | -0.56279 | 11.27405 | -0.47438 | 0.637094 | 0.994456 | -4.9887 |
| O00429 | DNM1L | 0.403968 | 7.713053 | 0.473075 | 0.638644 | 0.994456 | -4.82116 |
| P06733 | ENO1 | -0.19697 | 12.59836 | -0.47193 | 0.638831 | 0.994456 | -4.98941 |
| P50395 | GDI2 | 0.248952 | 11.20433 | 0.468265 | 0.641432 | 0.994456 | -4.99046 |
| A0A0C4DH | #N/D | -0.42497 | 9.855679 | -0.46735 | 0.642304 | 0.994456 | -4.95906 |
| P30086 | PEBP1 | 0.222434 | 12.34105 | 0.464908 | 0.643819 | 0.994456 | -4.99142 |
| P35520 | CBS | -0.30832 | 8.684517 | -0.46362 | 0.644918 | 0.994456 | -4.99168 |
| Q99747 | NAPG | -0.24959 | 8.259074 | -0.46076 | 0.646804 | 0.994456 | -4.96094 |
| P40429 | RPL13A | -0.2881 | 11.00395 | -0.46061 | 0.646879 | 0.994456 | -4.99263 |
| Q9UDR5 | AASS | 0.587756 | 10.55739 | 0.457989 | 0.648752 | 0.994456 | -4.99337 |
| P19367 | HK1 | 0.218464 | 11.89524 | 0.455404 | 0.650598 | 0.994456 | -4.99409 |
| P62258 | YWHAE | -0.1978 | 12.79759 | -0.45398 | 0.651614 | 0.994456 | -4.99448 |
| P37837 | TALDO1 | 0.235583 | 13.06675 | 0.453544 | 0.651928 | 0.994456 | -4.9946 |
| P09960 | LTA4H | 0.281445 | 8.842087 | 0.452704 | 0.652784 | 0.994456 | -4.96297 |
| P18206 | VCL | -0.17753 | 12.66382 | -0.45151 | 0.653385 | 0.994456 | -4.99517 |
| P04406 | GAPDH | -0.31065 | 14.71391 | -0.4514 | 0.653462 | 0.994456 | -4.9952 |
| P51178-2 | #N/D | -0.39343 | 9.240432 | -0.45105 | 0.653889 | 0.994456 | -4.96345 |
| P26038 | MSN | -0.33629 | 8.378736 | -0.45065 | 0.654251 | 0.994456 | -4.99527 |
| P46108 | CRK | -0.27972 | 8.591725 | -0.44805 | 0.656001 | 0.994456 | -4.99605 |
| P00738 | HP | -0.22762 | 16.23776 | -0.44737 | 0.656354 | 0.994456 | -4.99631 |
| P84077 | ARF1 | 0.325009 | 9.006425 | 0.447072 | 0.656597 | 0.994456 | -4.99637 |
| Q9UQM7 | CAMK2A | -0.35115 | 11.74729 | -0.44571 | 0.657546 | 0.994456 | -4.99676 |
| O14791 | APOL1 | 0.297081 | 13.07569 | 0.445497 | 0.657697 | 0.994456 | -4.99682 |
| P62937 | PPIA | 0.222775 | 11.22551 | 0.444963 | 0.65808 | 0.994456 | -4.99696 |
| P46459 | NSF | 0.323011 | 12.65607 | 0.443268 | 0.659297 | 0.994456 | -4.99742 |
| P61604 | HSPE1 | -0.31228 | 11.60591 | -0.44062 | 0.661203 | 0.994456 | -4.99814 |
| Q99439 | CNN2 | 0.284664 | 9.43566 | 0.439689 | 0.661904 | 0.994456 | -4.99837 |
| P30084 | ECHS1 | -0.20529 | 10.15609 | -0.43747 | 0.663468 | 0.994456 | -4.99898 |
| Q99497 | PARK7 | -0.41936 | 9.50833 | -0.43655 | 0.664267 | 0.994456 | -4.90301 |

| | | | | | | | |
|---|---|---|---|---|---|---|---|
| P62917 | RPL8 | 0.494814 | 11.02457 | 0.435695 | 0.664782 | 0.994456 | -4.96749 |
| P14324 | FDPS | 0.257937 | 8.575581 | 0.435651 | 0.66511 | 0.994456 | -4.96735 |
| P50914 | RPL14 | -0.33018 | 9.06483 | -0.43535 | 0.665131 | 0.994456 | -4.96752 |
| Q15233 | NONO | -0.2868 | 9.777373 | -0.43398 | 0.665987 | 0.994456 | -4.99991 |
| Q13938 | #N/D | -0.24318 | 11.20708 | -0.43303 | 0.666677 | 0.994456 | -5.00017 |
| O75390 | CS | 0.361454 | 11.12214 | 0.426347 | 0.671507 | 0.994456 | -5.00192 |
| P80404 | ABAT | -0.24572 | 7.872349 | -0.42658 | 0.671892 | 0.994456 | -4.87517 |
| P04217 | A1BG | 0.239966 | 14.14437 | 0.42353 | 0.673548 | 0.994456 | -5.00265 |
| O14980 | XPO1 | -0.37912 | 7.346187 | -0.42163 | 0.67571 | 0.994456 | -4.83095 |
| Q9GZV7 | HAPLN2 | -0.24093 | 13.03861 | -0.41482 | 0.679876 | 0.994456 | -5.00489 |
| P67936 | TPM4 | -0.36151 | 9.11576 | -0.41251 | 0.681862 | 0.994456 | -4.97308 |
| P53396 | ACLY | -0.32266 | 8.061162 | -0.41175 | 0.682231 | 0.994456 | -5.0056 |
| P01766 | #N/D | 0.294525 | 9.036312 | 0.408106 | 0.684959 | 0.994456 | -4.97419 |
| P13798 | APEH | -0.28984 | 11.77021 | -0.40589 | 0.686387 | 0.994456 | -5.00713 |
| P61254 | RPL26 | 0.252967 | 10.98212 | 0.401456 | 0.689629 | 0.994456 | -5.00822 |
| O94819 | KBTBD11 | 0.257831 | 9.342712 | 0.40125 | 0.689807 | 0.994456 | -5.00826 |
| P06576 | ATP5F1B | 0.281145 | 11.92408 | 0.401067 | 0.689913 | 0.994456 | -5.00832 |
| Q96FW1 | OTUB1 | 0.377451 | 7.493366 | 0.400389 | 0.691088 | 0.994456 | -4.80184 |
| Q9NRW1 | RAB6B | -0.2719 | 9.368933 | -0.39894 | 0.691525 | 0.994456 | -5.00881 |
| P04004 | VTN | 0.231487 | 14.98777 | 0.398005 | 0.692156 | 0.994456 | -5.00907 |
| Q9UPY8 | MAPRE3 | -0.18723 | 11.03301 | -0.39773 | 0.692358 | 0.994456 | -5.00913 |
| Q8N573 | OXR1 | -0.3892 | 8.278275 | -0.3979 | 0.692968 | 0.994456 | -4.85595 |
| P22061 | PCMT1 | -0.22761 | 10.41461 | -0.39613 | 0.693534 | 0.994456 | -5.00952 |
| Q16143 | SNCB | 0.239553 | 11.41947 | 0.396068 | 0.693576 | 0.994456 | -5.00954 |
| P35613-2 | #N/D | 0.208829 | 10.74277 | 0.39359 | 0.695394 | 0.994456 | -5.01014 |
| P38159 | RBMX | 0.392241 | 13.33236 | 0.392693 | 0.696053 | 0.994456 | -5.01035 |
| P27348 | YWHAQ | -0.30586 | 11.63346 | -0.39204 | 0.696563 | 0.994456 | -5.0105 |
| P01714 | #N/D | -0.34012 | 10.84083 | -0.39103 | 0.697278 | 0.994456 | -5.01075 |
| Q9NRV9 | HEBP1 | 0.366225 | 10.05472 | 0.390536 | 0.697819 | 0.994456 | -4.97829 |
| Q96AX9-5 | #N/D | 0.365206 | 9.366756 | 0.387136 | 0.700166 | 0.994456 | -4.97913 |
| P05155 | SERPING1 | -0.24757 | 12.77219 | -0.38108 | 0.704598 | 0.994456 | -5.0131 |

| | | | | | | | |
|---|---|---|---|---|---|---|---|
| Q07954 | LRP1 | -0.27527 | 9.068573 | -0.37972 | 0.705686 | 0.994456 | -5.01338 |
| A0A0B4J1X | #N/D | -0.47789 | 10.0226 | -0.37863 | 0.706434 | 0.994456 | -5.01366 |
| P39687 | ANP32A | -0.25671 | 9.050789 | -0.37858 | 0.706558 | 0.994456 | -5.01363 |
| P01599 | #N/D | -0.31531 | 9.808588 | -0.37822 | 0.706763 | 0.994456 | -5.01374 |
| P00751 | CFB | 0.224989 | 13.86732 | 0.377671 | 0.707118 | 0.994456 | -5.01389 |
| A0A075B6I | #N/D | 0.396436 | 9.154402 | 0.376333 | 0.708422 | 0.994456 | -4.95367 |
| P02787 | TF | 0.208401 | 14.24336 | 0.374826 | 0.70922 | 0.994456 | -5.01455 |
| P52209 | PGD | 0.164126 | 9.886713 | 0.374575 | 0.709406 | 0.994456 | -5.01461 |
| O14576 | DYNC1I1 | 0.291307 | 8.850936 | 0.372243 | 0.711158 | 0.994456 | -5.01513 |
| P04216 | THY1 | -0.35463 | 9.984486 | -0.37111 | 0.711973 | 0.994456 | -5.0154 |
| Q5IS67 | #N/D | -0.419 | 8.456854 | -0.37068 | 0.712599 | 0.994456 | -4.9167 |
| P07910 | HNRNPC | -0.16465 | 10.97799 | -0.36978 | 0.712956 | 0.994456 | -5.0157 |
| Q03591 | CFHR1 | -0.22815 | 9.2033 | -0.3691 | 0.713627 | 0.994456 | -5.01577 |
| Q12906-7 | #N/D | 0.24167 | 10.81171 | 0.368731 | 0.713733 | 0.994456 | -5.01594 |
| Q9UN36 | NDRG2 | -0.27043 | 11.61186 | -0.36854 | 0.713878 | 0.994456 | -5.01598 |
| Q96F85 | CNRIP1 | 0.212239 | 10.63719 | 0.368375 | 0.713997 | 0.994456 | -5.01602 |
| P29401 | TKT | 0.170682 | 12.31095 | 0.368189 | 0.714135 | 0.994456 | -5.01606 |
| A0A0C4DH | #N/D | 0.276369 | 12.36129 | 0.367374 | 0.714739 | 0.994456 | -5.01624 |
| Q99962 | SH3GL2 | -0.18573 | 12.54019 | -0.36455 | 0.716838 | 0.994456 | -5.01688 |
| Q9UNZ2 | NSFL1C | 0.272089 | 8.71179 | 0.362024 | 0.718847 | 0.994456 | -4.98457 |
| A0A087WS | #N/D | -0.32846 | 13.2827 | -0.36184 | 0.718851 | 0.994456 | -5.01748 |
| P0C0S5 | H2AZ1 | 0.273799 | 10.02911 | 0.361595 | 0.71903 | 0.994456 | -5.01753 |
| P02748 | C9 | -0.29463 | 13.54131 | -0.36122 | 0.719311 | 0.994456 | -5.01761 |
| Q13509 | TUBB3 | -0.20505 | 9.79097 | -0.35602 | 0.723204 | 0.994456 | -4.98588 |
| P36957 | DLST | 0.342558 | 8.614293 | 0.355742 | 0.723718 | 0.994456 | -4.87797 |
| Q14203 | DCTN1 | 0.347689 | 7.611697 | 0.354605 | 0.725416 | 0.994456 | -4.88857 |
| P02042 | HBD | 0.209294 | 10.61689 | 0.351641 | 0.726441 | 0.994456 | -5.01971 |
| P11021 | HSPA5 | 0.177674 | 14.82701 | 0.351419 | 0.726606 | 0.994456 | -5.01975 |
| P00338 | LDHA | -0.18318 | 12.44144 | -0.34966 | 0.727916 | 0.994456 | -5.02013 |
| P02649 | APOE | 0.246271 | 16.1627 | 0.349576 | 0.727982 | 0.994456 | -5.02015 |
| P06744 | GPI | 0.204695 | 11.87388 | 0.346607 | 0.730199 | 0.994456 | -5.02078 |

| | | | | | | | |
|---|---|---|---|---|---|---|---|
| O95445 | APOM | -0.37334 | 11.61754 | -0.34591 | 0.730721 | 0.994456 | -5.02093 |
| P23246 | SFPQ | 0.204982 | 12.17656 | 0.341237 | 0.734216 | 0.994456 | -5.02191 |
| P02675 | FGB | 0.170446 | 14.18233 | 0.339366 | 0.735617 | 0.994456 | -5.0223 |
| P46776 | RPL27A | 0.279119 | 10.85159 | 0.33876 | 0.736094 | 0.994456 | -5.02242 |
| Q9Y696 | CLIC4 | -0.29892 | 8.300412 | -0.3389 | 0.736211 | 0.994456 | -4.98925 |
| P22087 | FBL | -0.19142 | 8.6313 | -0.33548 | 0.738681 | 0.994456 | -5.02304 |
| Q5IS61 | #N/D | -0.23358 | 8.614239 | -0.33531 | 0.73881 | 0.994456 | -4.99 |
| Q9P2U7 | SLC17A7 | 0.310715 | 10.35073 | 0.332699 | 0.740664 | 0.994456 | -5.02365 |
| Q9UI12 | ATP6V1H | 0.30814 | 8.594544 | 0.332177 | 0.741055 | 0.994456 | -5.02376 |
| Q07020 | RPL18 | 0.317129 | 11.58711 | 0.331597 | 0.741444 | 0.994456 | -5.0239 |
| P01011 | SERPINA3 | -0.13327 | 14.45594 | -0.32714 | 0.744795 | 0.994456 | -5.0248 |
| P51649 | ALDH5A1 | 0.150134 | 11.25326 | 0.326791 | 0.745057 | 0.994456 | -5.02487 |
| P49418 | AMPH | 0.262599 | 10.79496 | 0.325661 | 0.745907 | 0.994456 | -5.02509 |
| P03952 | KLKB1 | 0.222154 | 13.33419 | 0.325018 | 0.746391 | 0.994456 | -5.02522 |
| P15313 | ATP6V1B1 | -0.22161 | 8.509029 | -0.3243 | 0.747029 | 0.994456 | -4.99215 |
| P31946 | YWHAB | 0.23576 | 9.674481 | 0.323992 | 0.747234 | 0.994456 | -4.96399 |
| P25398 | RPS12 | -0.19629 | 8.911054 | -0.32354 | 0.74757 | 0.994456 | -5.02549 |
| P49411 | TUFM | -0.26837 | 11.6554 | -0.32236 | 0.748392 | 0.994456 | -5.02575 |
| P60900 | PSMA6 | -0.31738 | 10.92948 | -0.3216 | 0.748963 | 0.994456 | -5.0259 |
| Q7Z3B1 | NEGR1 | -0.30345 | 7.928181 | -0.32056 | 0.750121 | 0.994456 | -4.89444 |
| P32119 | PRDX2 | 0.17657 | 13.16617 | 0.318279 | 0.75147 | 0.994456 | -5.02655 |
| Q9UHG2 | PCSK1N | -0.24385 | 7.955461 | -0.31814 | 0.751974 | 0.994456 | -4.94828 |
| P62750 | RPL23A | 0.195746 | 11.64551 | 0.31757 | 0.752004 | 0.994456 | -5.02669 |
| P13533 | MYH6 | 0.173605 | 10.46117 | 0.317137 | 0.752332 | 0.994456 | -5.02677 |
| P00736 | C1R | 0.246749 | 13.74472 | 0.31511 | 0.753862 | 0.994456 | -5.02717 |
| P05546 | SERPIND1 | 0.202666 | 11.79322 | 0.312102 | 0.756135 | 0.994456 | -5.02774 |
| P00492 | HPRT1 | 0.276996 | 10.76362 | 0.312081 | 0.756172 | 0.994456 | -5.02774 |
| Q02252 | ALDH6A1 | 0.422044 | 13.39322 | 0.312001 | 0.75621 | 0.994456 | -5.02776 |
| P55209 | NAP1L1 | -0.29732 | 12.57393 | -0.31053 | 0.757341 | 0.994456 | -5.02803 |
| P14314 | PRKCSH | 0.24193 | 12.48603 | 0.310303 | 0.757561 | 0.994456 | -5.02806 |
| P22626 | HNRNPA2B1 | 0.143414 | 12.6985 | 0.309766 | 0.757901 | 0.994456 | -5.02819 |

| | | | | | | | |
|---|---|---|---|---|---|---|---|
| Q14894 | CRYM | -0.18889 | 12.35016 | -0.30787 | 0.759337 | 0.994456 | -5.02855 |
| P78347 | GTF2I | -0.19908 | 9.347322 | -0.30759 | 0.75957 | 0.994456 | -4.99526 |
| O94919 | ENDOD1 | -0.21477 | 9.449366 | -0.3062 | 0.760643 | 0.994456 | -4.9955 |
| P62280 | RPS11 | 0.188899 | 11.12395 | 0.305136 | 0.761406 | 0.994456 | -5.02906 |
| A0A0C4DH | #N/D | 0.330937 | 10.33687 | 0.30379 | 0.762446 | 0.994456 | -5.0293 |
| Q6TUY0 | #N/D | -0.45753 | 12.02649 | -0.30371 | 0.762529 | 0.994456 | -5.02931 |
| P35908 | KRT2 | 0.325773 | 14.98502 | 0.303066 | 0.762974 | 0.994456 | -5.02945 |
| P35637 | FUS | -0.20247 | 9.460308 | -0.30241 | 0.763494 | 0.994456 | -5.02956 |
| P50454 | SERPINH1 | -0.20265 | 9.103501 | -0.30168 | 0.764043 | 0.994456 | -4.99631 |
| Q5T7N2 | L1TD1 | -0.17893 | 8.91276 | -0.30135 | 0.764388 | 0.994456 | -4.99634 |
| P61266-2 | #N/D | 0.138062 | 9.516859 | 0.296581 | 0.767895 | 0.994456 | -5.03064 |
| Q9UHD8 | SEPTIN9 | -0.28623 | 7.179257 | -0.29604 | 0.768925 | 0.994456 | -4.75246 |
| P02763 | ORM1 | -0.21741 | 11.84132 | -0.29476 | 0.76928 | 0.994456 | -5.03097 |
| Q99729-3 | #N/D | 0.317033 | 12.65612 | 0.294532 | 0.769452 | 0.994456 | -5.03101 |
| P48643 | CCT5 | 0.197151 | 9.491954 | 0.293786 | 0.770019 | 0.994456 | -5.03115 |
| P35998 | PSMC2 | 0.291971 | 10.28438 | 0.293145 | 0.770569 | 0.994456 | -4.99778 |
| P27918 | CFP | -0.19616 | 11.216 | -0.2928 | 0.770772 | 0.994456 | -5.03132 |
| P52758 | RIDA | 0.198855 | 8.569229 | 0.292568 | 0.771007 | 0.994456 | -4.99788 |
| Q13247 | SRSF6 | -0.20834 | 8.67543 | -0.29193 | 0.771472 | 0.994456 | -4.998 |
| Q9H0U4 | RAB1B | 0.188527 | 8.154821 | 0.291992 | 0.771623 | 0.994456 | -4.93026 |
| A2NJV5 | #N/D | 0.405478 | 8.920818 | 0.291325 | 0.772047 | 0.994456 | -4.99805 |
| Q8WUM4 | PDCD6IP | 0.460295 | 7.545402 | 0.289592 | 0.774677 | 0.994456 | -4.71781 |
| Q99623 | PHB2 | 0.267539 | 10.71072 | 0.286826 | 0.775316 | 0.994456 | -5.03238 |
| O75636 | FCN3 | 0.146756 | 11.6985 | 0.28343 | 0.777903 | 0.994456 | -5.03298 |
| P26583 | HMGB2 | -0.20852 | 9.427067 | -0.28185 | 0.779107 | 0.994456 | -5.03325 |
| P28161 | GSTM2 | -0.19612 | 8.502551 | -0.27892 | 0.781601 | 0.994456 | -4.97151 |
| P29972 | AQP1 | 0.205747 | 8.612235 | 0.277818 | 0.782312 | 0.994456 | -5.0339 |
| Q96F07 | CYFIP2 | 0.328611 | 9.351898 | 0.276869 | 0.782991 | 0.994456 | -5.00048 |
| Q5U7I5 | #N/D | 0.321169 | 12.18705 | 0.27599 | 0.783601 | 0.994456 | -5.03425 |
| P27169 | PON1 | -0.25005 | 14.83311 | -0.27591 | 0.783664 | 0.994456 | -5.03426 |
| O75083 | WDR1 | -0.13594 | 11.68371 | -0.2753 | 0.784109 | 0.994456 | -5.03437 |

| | | | | | | | |
|---|---|---|---|---|---|---|---|
| P10606 | COX5B | -0.25466 | 8.344202 | -0.27542 | 0.784143 | 0.994456 | -4.95526 |
| O43761 | SYNGR3 | 0.260526 | 10.57735 | 0.274374 | 0.784818 | 0.994456 | -5.03453 |
| P20336 | RAB3A | -0.25861 | 9.798318 | -0.27365 | 0.785387 | 0.994456 | -5.00102 |
| Q16658 | FSCN1 | -0.16279 | 13.04135 | -0.26875 | 0.789124 | 0.994456 | -5.03546 |
| P18621 | RPL17 | 0.276212 | 11.78909 | 0.267439 | 0.790124 | 0.994456 | -5.03568 |
| P18077 | RPL35A | 0.274268 | 11.39346 | 0.267303 | 0.790265 | 0.994456 | -5.03569 |
| P01042-2 | #N/D | 0.255143 | 11.83736 | 0.264408 | 0.792483 | 0.994456 | -5.03616 |
| P23515 | OMG | 0.141547 | 10.03831 | 0.262076 | 0.794235 | 0.994456 | -5.03655 |
| P30101 | PDIA3 | 0.193062 | 12.87952 | 0.261496 | 0.794679 | 0.994456 | -5.03664 |
| Q13838 | DDX39B | -0.24536 | 13.4483 | -0.26148 | 0.794695 | 0.994456 | -5.03665 |
| P02750 | LRG1 | 0.218594 | 10.4896 | 0.260328 | 0.795576 | 0.994456 | -5.03683 |
| Q16775 | HAGH | 0.223929 | 10.09877 | 0.259277 | 0.796382 | 0.994456 | -5.037 |
| P48637 | GSS | -0.26112 | 8.001266 | -0.25822 | 0.797584 | 0.994456 | -4.9032 |
| Q9Y2T3 | GDA | -0.19514 | 10.08686 | -0.25718 | 0.798028 | 0.994456 | -5.03732 |
| P23528 | CFL1 | 0.153229 | 12.52033 | 0.256648 | 0.798401 | 0.994456 | -5.03741 |
| P60174 | TPI1 | -0.26331 | 13.33968 | -0.25555 | 0.799245 | 0.994456 | -5.03759 |
| P14174 | MIF | -0.2224 | 11.07338 | -0.25463 | 0.799955 | 0.994456 | -5.03773 |
| Q15392 | DHCR24 | 0.223375 | 8.63106 | 0.253663 | 0.800788 | 0.994456 | -5.00408 |
| P29622 | SERPINA4 | -0.20234 | 8.812058 | -0.25353 | 0.801053 | 0.994456 | -4.97528 |
| Q14697-2 | #N/D | 0.163297 | 9.735768 | 0.251746 | 0.802222 | 0.994456 | -5.03816 |
| P02765 | AHSG | -0.15799 | 11.76711 | -0.25054 | 0.803098 | 0.994456 | -5.03836 |
| P13489 | RNH1 | -0.21914 | 9.869775 | -0.24905 | 0.804244 | 0.994456 | -5.03859 |
| Q96PD5 | PGLYRP2 | -0.13878 | 13.17715 | -0.24813 | 0.804951 | 0.994456 | -5.03873 |
| P09651 | HNRNPA1 | 0.160529 | 12.21855 | 0.245424 | 0.807035 | 0.994456 | -5.03914 |
| P09871 | C1S | -0.13559 | 13.55189 | -0.24424 | 0.807945 | 0.994456 | -5.03932 |
| P01704 | #N/D | 0.151584 | 11.22917 | 0.243994 | 0.808137 | 0.994456 | -5.03936 |
| P09382 | LGALS1 | 0.177319 | 9.996774 | 0.243228 | 0.808727 | 0.994456 | -5.03947 |
| P62753 | RPS6 | -0.18093 | 9.50014 | -0.2431 | 0.808827 | 0.994456 | -5.03949 |
| P37108 | SRP14 | 0.349423 | 8.187199 | 0.243622 | 0.809013 | 0.994456 | -4.7736 |
| Q01105 | SET | 0.137718 | 12.74759 | 0.241131 | 0.810343 | 0.994456 | -5.03978 |
| Q14624 | ITIH4 | -0.23978 | 9.94507 | -0.23946 | 0.811717 | 0.994456 | -4.97728 |

| | | | | | | | |
|---|---|---|---|---|---|---|---|
| P23284 | PPIB | -0.18341 | 11.31656 | -0.23929 | 0.811816 | 0.994456 | -5.04004 |
| P17096 | HMGA1 | -0.23528 | 10.9518 | -0.23819 | 0.81261 | 0.994456 | -5.04021 |
| P52597 | HNRNPF | 0.127393 | 9.211955 | 0.235554 | 0.814663 | 0.994456 | -5.04059 |
| P62318 | SNRPD3 | 0.145833 | 9.893724 | 0.233159 | 0.816497 | 0.994456 | -5.04094 |
| P00734 | F2 | 0.144221 | 14.7427 | 0.23312 | 0.816528 | 0.994456 | -5.04095 |
| P27635 | RPL10 | -0.12417 | 8.546315 | -0.23122 | 0.818011 | 0.994456 | -5.04121 |
| Q9UBC3 | DNMT3B | -0.21431 | 11.72397 | -0.23087 | 0.818265 | 0.994456 | -5.04127 |
| Q13177 | PAK2 | -0.19475 | 8.338763 | -0.23112 | 0.818304 | 0.994456 | -4.93854 |
| P01861 | #N/D | -0.14003 | 9.667126 | -0.23086 | 0.81832 | 0.994456 | -5.00732 |
| Q92777 | SYN2 | -0.22048 | 10.07691 | -0.22957 | 0.819269 | 0.994456 | -5.04145 |
| P34897 | SHMT2 | -0.15388 | 10.26867 | -0.22916 | 0.819586 | 0.994456 | -5.04151 |
| Q9UMS4 | PRPF19 | 0.188691 | 9.289913 | 0.227914 | 0.820552 | 0.994456 | -5.04168 |
| Q16181 | SEPTIN7 | 0.295196 | 11.66741 | 0.227804 | 0.820637 | 0.994456 | -5.0417 |
| Q9Y3E1 | HDGFL3 | 0.219808 | 8.28437 | 0.227098 | 0.821231 | 0.994456 | -5.04178 |
| P39023 | RPL3 | -0.27838 | 11.33349 | -0.22206 | 0.825087 | 0.994456 | -5.04249 |
| Q9Y6R7 | FCGBP | -0.11499 | 13.98588 | -0.22178 | 0.825301 | 0.994456 | -5.04253 |
| O75347 | TBCA | -0.15263 | 9.096358 | -0.22156 | 0.825471 | 0.994456 | -5.04256 |
| P17174 | GOT1 | 0.15021 | 13.06163 | 0.221405 | 0.825591 | 0.994456 | -5.04258 |
| O95197 | RTN3 | 0.207302 | 8.031271 | 0.221238 | 0.826079 | 0.994456 | -4.93967 |
| Q9BPU6 | DPYSL5 | 0.12671 | 10.87508 | 0.218712 | 0.827678 | 0.994456 | -5.04294 |
| P02747 | C1QC | 0.222816 | 14.27094 | 0.217692 | 0.828469 | 0.994456 | -5.04308 |
| P16401 | H1-5 | -0.16682 | 10.04102 | -0.21667 | 0.829261 | 0.994456 | -5.04322 |
| Q13151 | HNRNPA0 | 0.15849 | 9.990869 | 0.216228 | 0.829649 | 0.994456 | -5.00924 |
| P26373 | RPL13 | -0.1257 | 10.14526 | -0.21478 | 0.830723 | 0.994456 | -5.04347 |
| P78371 | CCT2 | 0.161977 | 10.10086 | 0.214676 | 0.830808 | 0.994456 | -5.04348 |
| P11216 | PYGB | 0.114081 | 10.47934 | 0.213888 | 0.831419 | 0.994456 | -5.04358 |
| P00915 | CA1 | -0.14609 | 10.62668 | -0.21382 | 0.83147 | 0.994456 | -5.04359 |
| Q13153 | PAK1 | -0.26379 | 7.407468 | -0.21035 | 0.834752 | 0.994456 | -4.77602 |
| Q12905 | ILF2 | 0.149984 | 9.328399 | 0.209341 | 0.834978 | 0.994456 | -5.04417 |
| P14866 | HNRNPL | -0.12892 | 12.72325 | -0.20919 | 0.835068 | 0.994456 | -5.04419 |
| P13639 | EEF2 | -0.11586 | 12.11228 | -0.20901 | 0.835204 | 0.994456 | -5.04422 |

| | | | | | | | |
|---|---|---|---|---|---|---|---|
| P21281 | ATP6V1B2 | 0.143976 | 13.63954 | 0.208054 | 0.83595 | 0.994456 | -5.04434 |
| P05787 | KRT8 | 0.110082 | 11.88015 | 0.207885 | 0.836081 | 0.994456 | -5.04436 |
| Q5IS74 | #N/D | -0.16281 | 8.979446 | -0.20724 | 0.836596 | 0.994456 | -5.04444 |
| P39019 | RPS19 | 0.201296 | 12.71185 | 0.207087 | 0.836701 | 0.994456 | -5.04446 |
| P07384 | CAPN1 | 0.178954 | 8.309043 | 0.2069 | 0.836875 | 0.994456 | -4.96424 |
| P62987 | UBA52 | -0.07987 | 11.77756 | -0.20671 | 0.836991 | 0.994456 | -5.04451 |
| Q13813 | SPTAN1 | 0.075284 | 11.99901 | 0.205085 | 0.838257 | 0.994456 | -5.04472 |
| P40939 | HADHA | 0.189162 | 9.753015 | 0.204953 | 0.838374 | 0.994456 | -5.04473 |
| P11142 | HSPA8 | -0.10189 | 13.19735 | -0.20057 | 0.841767 | 0.994456 | -5.04528 |
| Q96GW7 | BCAN | -0.09812 | 8.242077 | -0.19897 | 0.843104 | 0.994456 | -5.04545 |
| P29218 | IMPA1 | 0.11732 | 11.50747 | 0.196239 | 0.845141 | 0.994456 | -5.04581 |
| Q15907 | RAB11B | -0.12359 | 9.911453 | -0.19362 | 0.847194 | 0.994456 | -5.04612 |
| P11137 | MAP2 | 0.113154 | 10.40085 | 0.19346 | 0.847306 | 0.994456 | -5.04614 |
| P04259 | KRT6B | -0.15314 | 11.57937 | -0.18949 | 0.850414 | 0.994456 | -5.01242 |
| Q13228 | SELENBP1 | 0.100701 | 10.91175 | 0.189403 | 0.850469 | 0.994456 | -5.04662 |
| P04181 | OAT | 0.151851 | 9.027395 | 0.188916 | 0.850875 | 0.994456 | -4.98334 |
| E9PAV3 | #N/D | -0.16137 | 12.14083 | -0.18825 | 0.851382 | 0.994456 | -5.04675 |
| P46783 | RPS10 | 0.156334 | 10.88473 | 0.187739 | 0.851767 | 0.994456 | -5.04681 |
| Q96C19 | EFHD2 | -0.18167 | 8.816843 | -0.1857 | 0.853511 | 0.994456 | -4.98365 |
| P51148 | RAB5C | 0.127789 | 8.07766 | 0.184308 | 0.854651 | 0.994456 | -4.9436 |
| Q9NR30 | DDX21 | -0.11642 | 8.743727 | -0.18361 | 0.855031 | 0.994456 | -5.04727 |
| P00568 | AK1 | -0.12078 | 11.14475 | -0.18099 | 0.857033 | 0.994456 | -5.04757 |
| P13591 | NCAM1 | 0.121377 | 12.6968 | 0.180945 | 0.857071 | 0.994456 | -5.04758 |
| P15531 | NME1 | -0.12897 | 8.297834 | -0.17895 | 0.858806 | 0.994456 | -4.94412 |
| O43491-4 | #N/D | 0.120535 | 9.677072 | 0.178611 | 0.858895 | 0.994456 | -5.04783 |
| O75874 | IDH1 | -0.16704 | 10.43067 | -0.17822 | 0.859203 | 0.994456 | -5.04788 |
| P51884 | LUM | 0.12006 | 8.535854 | 0.176288 | 0.860834 | 0.994456 | -5.0138 |
| P08697 | SERPINF2 | 0.078859 | 12.89207 | 0.173026 | 0.863261 | 0.994456 | -5.04844 |
| P28482 | MAPK1 | -0.08188 | 10.57932 | -0.17148 | 0.864482 | 0.994456 | -5.0486 |
| P68371 | TUBB4B | 0.172358 | 11.99643 | 0.170735 | 0.865054 | 0.994456 | -5.04868 |
| A0A075B6I | #N/D | -0.15144 | 12.27284 | -0.16958 | 0.865955 | 0.994456 | -5.0488 |

| | | | | | | | |
|---|---|---|---|---|---|---|---|
| P0DOX8 | #N/D | 0.193514 | 13.34101 | 0.168066 | 0.867154 | 0.994456 | -5.04895 |
| P00748 | F12 | 0.081434 | 11.60206 | 0.164322 | 0.870075 | 0.994456 | -5.04934 |
| Q9H299 | SH3BGRL3 | -0.13648 | 8.301393 | -0.1612 | 0.872639 | 0.994456 | -4.9457 |
| P02652 | APOA2 | 0.102635 | 13.89358 | 0.160952 | 0.872716 | 0.994456 | -5.04967 |
| Q15185 | PTGES3 | 0.177646 | 14.08679 | 0.159006 | 0.874253 | 0.994456 | -5.04986 |
| P49591 | SARS1 | -0.11716 | 8.044302 | -0.15802 | 0.875145 | 0.994456 | -4.94596 |
| P43652 | AFM | 0.077405 | 12.25694 | 0.157486 | 0.875434 | 0.994456 | -5.05001 |
| P09493-4 | #N/D | -0.10521 | 12.6635 | -0.15669 | 0.87606 | 0.994456 | -5.05009 |
| P02679-2 | #N/D | -0.07317 | 11.17822 | -0.15575 | 0.876798 | 0.994456 | -5.05018 |
| P05091 | ALDH2 | -0.09233 | 12.45982 | -0.15558 | 0.876927 | 0.994456 | -5.0502 |
| P20339 | RAB5A | 0.088044 | 8.711876 | 0.150689 | 0.880769 | 0.994456 | -5.05066 |
| Q8WVE0 | EEF1AKMT1 | 0.203269 | 12.59871 | 0.151401 | 0.880903 | 0.994456 | -4.8643 |
| P08559-4 | #N/D | 0.100515 | 10.42252 | 0.148776 | 0.882271 | 0.994456 | -5.05083 |
| A0A0C4DH | #N/D | 0.108634 | 9.696796 | 0.148685 | 0.882342 | 0.994456 | -5.05084 |
| P50991 | CCT4 | 0.076088 | 13.00041 | 0.147826 | 0.883017 | 0.994456 | -5.05092 |
| O95336 | PGLS | -0.11702 | 10.2894 | -0.14576 | 0.884639 | 0.994456 | -5.0511 |
| P28072 | PSMB6 | 0.097015 | 9.15565 | 0.145049 | 0.885199 | 0.994456 | -5.05117 |
| P06312 | #N/D | -0.10643 | 13.06851 | -0.14498 | 0.885253 | 0.994456 | -5.05117 |
| Q9UBB6 | NCDN | -0.12568 | 8.646641 | -0.14453 | 0.885627 | 0.994456 | -5.01681 |
| P17655 | CAPN2 | 0.118782 | 9.117694 | 0.143186 | 0.88675 | 0.994456 | -4.94713 |
| P02790 | HPX | -0.11353 | 14.96138 | -0.14307 | 0.886756 | 0.994456 | -5.05134 |
| P16112 | #N/D | -0.09972 | 8.198785 | -0.14121 | 0.888291 | 0.994456 | -4.94728 |
| Q15631 | TSN | 0.118446 | 8.819731 | 0.140957 | 0.888444 | 0.994456 | -5.05152 |
| P23396 | RPS3 | 0.096285 | 12.74094 | 0.140312 | 0.888922 | 0.994456 | -5.05158 |
| O43813 | LANCL1 | 0.100841 | 12.55751 | 0.139086 | 0.889887 | 0.994456 | -5.05169 |
| Q5TFQ8 | SIRPB1 | 0.119414 | 8.396877 | 0.138874 | 0.890257 | 0.994456 | -4.91478 |
| Q96GD0 | PDXP | 0.093254 | 10.49093 | 0.138206 | 0.890579 | 0.994456 | -5.05176 |
| P08758 | ANXA5 | 0.087603 | 12.52081 | 0.137345 | 0.891256 | 0.994456 | -5.05184 |
| Q6PCE3 | PGM2L1 | 0.112335 | 10.75219 | 0.136901 | 0.891605 | 0.994456 | -5.05187 |
| Q08211 | DHX9 | -0.09884 | 13.43428 | -0.13219 | 0.89531 | 0.994456 | -5.05226 |
| P09874 | PARP1 | -0.05911 | 12.84743 | -0.13157 | 0.895798 | 0.994456 | -5.05231 |

| | | | | | | | |
|---|---|---|---|---|---|---|---|
| P55072 | VCP | 0.081421 | 13.88593 | 0.13122 | 0.896077 | 0.994456 | -5.05234 |
| P11279 | LAMP1 | -0.09144 | 9.296891 | -0.12717 | 0.899311 | 0.994456 | -4.98879 |
| A0A0C4DH | #N/D | -0.11202 | 14.37611 | -0.12663 | 0.89969 | 0.994456 | -5.0527 |
| P22792 | CPN2 | 0.101471 | 10.29617 | 0.124295 | 0.901532 | 0.994456 | -5.05288 |
| P01871 | #N/D | 0.082403 | 16.51568 | 0.123472 | 0.90218 | 0.994456 | -5.05295 |
| P02686 | MBP | 0.050572 | 12.52744 | 0.122038 | 0.903311 | 0.994456 | -5.05306 |
| P16403 | H1-2 | -0.05203 | 11.57771 | -0.12123 | 0.903947 | 0.994456 | -5.05312 |
| Q14123 | PDE1C | 0.147183 | 9.472294 | 0.121199 | 0.903988 | 0.994456 | -5.05312 |
| P25786 | PSMA1 | -0.11643 | 8.552348 | -0.12126 | 0.904019 | 0.994456 | -4.97185 |
| O95865 | DDAH2 | -0.11505 | 8.695045 | -0.11899 | 0.905775 | 0.994456 | -4.98935 |
| Q9UQ80 | PA2G4 | 0.079923 | 9.950951 | 0.118444 | 0.906144 | 0.994456 | -5.05332 |
| Q9Y617 | PSAT1 | 0.07455 | 11.24982 | 0.116395 | 0.90776 | 0.994456 | -5.05347 |
| P01619 | #N/D | -0.07333 | 14.00764 | -0.11599 | 0.908113 | 0.994456 | -5.01898 |
| P23526 | AHCY | 0.058768 | 12.4871 | 0.115455 | 0.908502 | 0.994456 | -5.05354 |
| Q15257-2 | #N/D | 0.10784 | 9.172995 | 0.115458 | 0.908541 | 0.994456 | -5.05353 |
| O94811 | TPPP | -0.05506 | 11.43773 | -0.11535 | 0.908587 | 0.994456 | -5.05354 |
| Q92752 | TNR | -0.052 | 11.57348 | -0.11159 | 0.911548 | 0.994456 | -5.0538 |
| P0DOY3 | #N/D | -0.09402 | 15.80861 | -0.10901 | 0.913587 | 0.994456 | -5.05398 |
| A0A075B6I | #N/D | 0.047083 | 13.46962 | 0.108525 | 0.913971 | 0.994456 | -5.05401 |
| P18124 | RPL7 | 0.058406 | 10.99238 | 0.108479 | 0.914008 | 0.994456 | -5.05402 |
| P01008 | SERPINC1 | -0.05561 | 14.22003 | -0.10765 | 0.91466 | 0.994456 | -5.05407 |
| P06730-2 | #N/D | -0.08596 | 8.849688 | -0.1057 | 0.916268 | 0.994456 | -5.05419 |
| P06727 | #N/D | 0.046008 | 14.15249 | 0.1043 | 0.917308 | 0.994456 | -5.05429 |
| P68366 | TUBA4A | -0.06284 | 8.182302 | -0.10272 | 0.918609 | 0.994456 | -4.97299 |
| Q00610 | CLTC | 0.080591 | 13.78944 | 0.10055 | 0.920271 | 0.994456 | -5.05452 |
| P27695 | APEX1 | 0.110661 | 9.067064 | 0.098672 | 0.921875 | 0.994456 | -4.99058 |
| P05141 | SLC25A5 | -0.07853 | 10.10956 | -0.09704 | 0.923051 | 0.994456 | -5.05474 |
| P33993 | MCM7 | 0.059884 | 8.945521 | 0.092787 | 0.926413 | 0.994456 | -5.05498 |
| P49419 | ALDH7A1 | -0.06114 | 13.146 | -0.08735 | 0.930709 | 0.994456 | -5.05529 |
| P22234 | PAICS | -0.05199 | 10.08226 | -0.08703 | 0.93096 | 0.994456 | -5.0553 |
| P48740 | MASP1 | 0.060195 | 8.81856 | 0.085924 | 0.931848 | 0.994456 | -5.05536 |

| | | | | | | | |
|---|---|---|---|---|---|---|---|
| P32322 | PYCR1 | -0.11237 | 8.954011 | -0.08558 | 0.932123 | 0.994456 | -5.02078 |
| Q07955 | SRSF1 | -0.05117 | 11.42114 | -0.0835 | 0.933751 | 0.994456 | -5.05549 |
| P46782 | RPS5 | -0.05778 | 8.807193 | -0.08328 | 0.933967 | 0.994456 | -4.99138 |
| P10720 | PF4V1 | 0.070159 | 11.14838 | 0.082761 | 0.934346 | 0.994456 | -5.05553 |
| Q92522 | H1-10 | 0.042196 | 9.944587 | 0.082677 | 0.934406 | 0.994456 | -5.05553 |
| P02647 | APOA1 | 0.059708 | 15.5712 | 0.082608 | 0.934461 | 0.994456 | -5.05553 |
| P20042 | EIF2S2 | -0.07823 | 8.98351 | -0.08121 | 0.93557 | 0.994456 | -5.0556 |
| P22102 | GART | 0.072254 | 10.28125 | 0.080393 | 0.936225 | 0.994456 | -5.05564 |
| P11766 | ADH5 | -0.06529 | 10.15171 | -0.07456 | 0.940835 | 0.994456 | -5.05592 |
| P02760 | AMBP | -0.03491 | 13.73303 | -0.07418 | 0.941138 | 0.994456 | -5.05594 |
| P01615 | #N/D | 0.059886 | 11.88343 | 0.073459 | 0.94175 | 0.994456 | -5.02134 |
| P48047 | ATP5PO | -0.07609 | 12.76757 | -0.0715 | 0.943258 | 0.994456 | -5.05606 |
| P16070 | CD44 | -0.05971 | 8.029181 | -0.07025 | 0.9443 | 0.994456 | -4.97451 |
| P31025 | LCN1 | 0.052509 | 14.44478 | 0.06922 | 0.945064 | 0.994456 | -5.05616 |
| P62829 | RPL23 | -0.07524 | 7.660806 | -0.06935 | 0.945069 | 0.994456 | -4.91832 |
| P49006 | MARCKSL1 | 0.069869 | 8.963973 | 0.069113 | 0.945158 | 0.994456 | -5.05616 |
| P60709 | ACTB | -0.04094 | 12.29443 | -0.06847 | 0.945657 | 0.994456 | -5.05619 |
| Q13491-4 | #N/D | -0.04607 | 9.101016 | -0.06779 | 0.946208 | 0.994456 | -5.02158 |
| P67775 | PPP2CA | 0.054164 | 8.996608 | 0.066857 | 0.946977 | 0.994456 | -5.02161 |
| Q08722 | CD47 | 0.040815 | 9.746025 | 0.066398 | 0.9473 | 0.994456 | -5.05628 |
| P31943 | HNRNPH1 | 0.040497 | 12.25814 | 0.06489 | 0.948495 | 0.994456 | -5.05634 |
| P08865 | RPSA | -0.03961 | 13.49287 | -0.06432 | 0.948949 | 0.994456 | -5.05636 |
| P11217 | PYGM | 0.033612 | 10.22972 | 0.063069 | 0.949939 | 0.994456 | -5.05641 |
| Q99447 | PCYT2 | 0.057462 | 9.931576 | 0.062965 | 0.950021 | 0.994456 | -5.05641 |
| O00410 | IPO5 | -0.0505 | 9.76488 | -0.06257 | 0.950333 | 0.994456 | -5.05643 |
| Q16352 | INA | 0.027104 | 13.06932 | 0.061541 | 0.95115 | 0.994456 | -5.05647 |
| P19823 | ITIH2 | 0.035923 | 14.69691 | 0.06106 | 0.951531 | 0.994456 | -5.05648 |
| P18669 | PGAM1 | -0.03301 | 13.03714 | -0.06018 | 0.952228 | 0.994456 | -5.05652 |
| P12268 | IMPDH2 | 0.045077 | 9.058515 | 0.05676 | 0.95497 | 0.994456 | -5.02198 |
| Q14240 | EIF4A2 | -0.03367 | 9.225594 | -0.05608 | 0.955477 | 0.994456 | -5.05666 |
| P04080 | CSTB | 0.045273 | 11.10002 | 0.056051 | 0.955503 | 0.994456 | -5.05666 |

| | | | | | | | |
|---|---|---|---|---|---|---|---|
| P08238 | HSP90AB1 | 0.031235 | 13.56843 | 0.0509 | 0.959588 | 0.995545 | -5.05683 |
| P30041 | PRDX6 | 0.047742 | 12.92684 | 0.050428 | 0.959963 | 0.995545 | -5.05685 |
| P60842 | EIF4A1 | -0.04613 | 11.09096 | -0.05039 | 0.95999 | 0.995545 | -5.05685 |
| O43866 | CD5L | 0.023673 | 13.65287 | 0.048143 | 0.961775 | 0.995854 | -5.05692 |
| O94856 | NFASC | -0.02897 | 13.27301 | -0.04652 | 0.96306 | 0.995854 | -5.05696 |
| P46781 | RPS9 | -0.02768 | 10.5113 | -0.04568 | 0.96373 | 0.995854 | -5.05699 |
| P12814 | ACTN1 | -0.04774 | 7.963896 | -0.04352 | 0.965518 | 0.996516 | -4.83249 |
| Q8B6J5 | #N/D | -0.03857 | 11.86572 | -0.03765 | 0.970133 | 0.997528 | -5.05719 |
| Q00839 | HNRNPU | -0.01886 | 12.5168 | -0.03586 | 0.971519 | 0.997528 | -5.05723 |
| P07477 | PRSS1 | -0.05001 | 13.06236 | -0.0343 | 0.972767 | 0.997528 | -5.05727 |
| P04179 | SOD2 | 0.020519 | 10.77234 | 0.033267 | 0.973581 | 0.997528 | -5.05729 |
| O60641 | SNAP91 | -0.01799 | 12.93002 | -0.03231 | 0.974344 | 0.997528 | -5.05731 |
| P49368 | CCT3 | 0.032329 | 10.62952 | 0.030993 | 0.975386 | 0.997528 | -5.05733 |
| Q71U36 | TUBA1A | 0.03106 | 13.10907 | 0.030305 | 0.976022 | 0.997528 | -4.95223 |
| P47914 | RPL29 | -0.01965 | 8.941895 | -0.02641 | 0.979029 | 0.997528 | -5.05742 |
| P62269 | RPS18 | 0.011622 | 10.80738 | 0.02517 | 0.980011 | 0.997528 | -5.05744 |
| P49327 | FASN | -0.01249 | 12.45025 | -0.02318 | 0.981589 | 0.997528 | -5.05746 |
| P30048 | PRDX3 | -0.0182 | 9.178145 | -0.02178 | 0.982729 | 0.997528 | -4.99318 |
| P31146 | CORO1A | 0.017748 | 9.870592 | 0.021726 | 0.982749 | 0.997528 | -5.05748 |
| P41250 | GARS1 | 0.01923 | 9.552422 | 0.018258 | 0.98551 | 0.997528 | -4.86926 |
| O75340 | PDCD6 | -0.02053 | 7.338176 | -0.01629 | 0.987116 | 0.997528 | -4.7654 |
| P05160 | F13B | -0.01285 | 11.66103 | -0.0156 | 0.987615 | 0.997528 | -5.05755 |
| P09972 | ALDOC | -0.00723 | 13.29965 | -0.01374 | 0.989088 | 0.997528 | -5.05757 |
| P52306-6 | #N/D | 0.009428 | 11.71115 | 0.012483 | 0.990085 | 0.997528 | -5.05758 |
| P31327 | CPS1 | 0.009143 | 9.318571 | 0.011761 | 0.990658 | 0.997528 | -5.05759 |
| P06753-2 | #N/D | -0.00834 | 11.55725 | -0.00998 | 0.992076 | 0.997528 | -5.0576 |
| P38606 | ATP6V1A | -0.00603 | 12.55831 | -0.00855 | 0.993211 | 0.997528 | -5.05761 |
| P46779 | RPL28 | -0.00568 | 9.65877 | -0.00736 | 0.994154 | 0.997528 | -5.05761 |
| O75363-2 | #N/D | -0.00534 | 8.935009 | -0.0073 | 0.994205 | 0.997528 | -5.05761 |
| P99999 | CYCS | 0.006016 | 7.341751 | 0.00639 | 0.994943 | 0.997528 | -4.83285 |
| P02746 | C1QB | 0.003442 | 14.57706 | 0.004735 | 0.996239 | 0.997528 | -5.05762 |

| | | | | | | | |
|---|---|---|---|---|---|---|---|
| P10909 | CLU | -0.00149 | 13.71219 | -0.00376 | 0.997011 | 0.997528 | -5.05763 |
| Q86VP6 | CAND1 | -0.00246 | 9.469724 | -0.00324 | 0.997423 | 0.997528 | -5.05763 |
| Q16629 | SRSF7 | -0.00256 | 9.577583 | -0.00311 | 0.997528 | 0.997528 | -5.05763 |
| P43243 | MATR3 | #N/D | 7.788896 | #N/D | #N/D | #N/D | #N/D |
| P68402 | PAFAH1B2 | #N/D | 6.807079 | #N/D | #N/D | #N/D | #N/D |
| Q99714 | HSD17B10 | #N/D | 7.015178 | #N/D | #N/D | #N/D | #N/D |

**Supplemental Table 4.** Differentially expressed proteins in oligodendrocytic extracellular vesicles (oEVs) isolated from patients following CAND treatment compared to the pre-CAND condition and the complete list of all proteins identified in the proteomic analysis. Each row corresponds to a protein identified by its UniProt accession number, gene symbol, and protein symbol. Expression changes are reported as $\log_2$ fold change (logFC); positive values indicate upregulation after CAND treatment, whereas negative values indicate downregulation. AveExpr represents the average expression across all samples. The t-statistic and associated P-value were derived from moderated t-tests. The B statistic reflects the log-odds that a given protein is differentially expressed.

#### Supplemental Table 4. Differentially Expressed Proteins in oEVs

| UniProt | Gene Symbol | Protein name | logFC | AveExpr | t | P.Value | B |
|---|---|---|---|---|---|---|---|
| P55209 | NAP1L1 | NP1L1 | 3.8688937 | 12.5739297 | 3.809774231 | 0.000355882 | 0.111641967 |
| P30626 | SRI | SORCN | 3.669829544 | 7.800030592 | 3.698254298 | 0.000693632 | -0.471135805 |
| Q99426 | TBCB | TBCB | 3.62432652 | 8.29542931 | 3.141189035 | 0.003063907 | -1.772990007 |
| P02656 | APOC3 | APOC3 | 3.173079509 | 12.92226708 | 4.14725299 | 0.000116525 | 1.102525458 |
| O75947 | ATP5PD | ATP5H | 2.985634964 | 8.722356721 | 3.24885603 | 0.002006479 | -1.387108813 |
| P02538 | KRT6A | K2C6A | 2.931508061 | 11.77847909 | 2.496950751 | 0.015705425 | -3.149160209 |
| A0A075B6J9 | IGLV2-18 | LV218 | 2.899647627 | 10.90519084 | 3.00151743 | 0.004107569 | -2.056730556 |
| Q06033 | ITIH3 | ITIH3 | 2.879375589 | 13.55378255 | 2.410810005 | 0.01926 | -3.389015287 |
| Q96FC7 | PHYHIPL | PHIPL | 2.809929273 | 10.07370766 | 2.63038393 | 0.011069482 | -2.884777878 |
| P35908 | KRT2 | K22E | 2.668527719 | 14.98502041 | 2.482530345 | 0.016102122 | -3.237710504 |
| P68871 | HBB | HBB | 2.559711133 | 14.57696991 | 3.095159971 | 0.00308217 | -1.810708716 |
| P12268 | IMPDH2 | IMPDH2 | 2.521064197 | 9.058515382 | 2.458895343 | 0.017560891 | -3.145814793 |
| Q15365 | PCBP1 | PCBP1 | 2.458727965 | 11.13094115 | 3.198004971 | 0.002327294 | -1.560675199 |
| Q15717 | ELAVL1 | ELAV1 | 2.379461088 | 11.80029837 | 2.253267423 | 0.028906297 | -3.542079328 |
| P31327 | CPS1 | CPSM | 2.342949108 | 9.318571371 | 3.013865753 | 0.003884906 | -2.013081884 |
| P49368 | CCT3 | TCPG | 2.318893817 | 10.62952483 | 2.223062173 | 0.030308572 | -3.768099501 |
| P32004 | L1CAM | L1CAM | 2.287075916 | 9.227820877 | 2.986835822 | 0.00437815 | -2.036828219 |
| P29762 | CRABP1 | RABP1 | 2.257920295 | 11.46854807 | 2.179979681 | 0.033524085 | -3.851524344 |
| P02747 | C1QC | C1QC | 2.205540798 | 14.27094211 | 2.15481999 | 0.035537431 | -3.899614791 |
| P38159 | RBMX | RBMX | 2.155869619 | 13.3323563 | 2.158354082 | 0.035248373 | -3.892887832 |
| Q9UJU6 | DBNL | DBNL | 2.152095971 | 10.24300741 | 3.03798059 | 0.003653927 | -1.957926732 |
| P68366 | DHX9 | DHX9 | 2.121781898 | 13.43428184 | 2.837661245 | 0.006337815 | -2.438583054 |
| P02533 | KRT14 | K1C14 | 2.020688806 | 11.09841923 | 2.546940596 | 0.013672955 | -3.098857608 |
| Q15102 | PAFAH1B3 | PA1B3 | 2.000511224 | 9.201982796 | 2.107297899 | 0.039802856 | -3.89506645 |
| Q7KZF4 | SND1 | SND1 | 1.976168834 | 9.615375041 | 2.550055512 | 0.013731683 | -3.03671501 |
| P04040 | CAT | CATA | 1.91541365 | 8.854697609 | 2.484370894 | 0.01672972 | -3.053635752 |
| P42704 | LRPPRC | LPPRC | 1.888030424 | 13.36178234 | 2.048147637 | 0.045295142 | -4.098285611 |
| P46776 | RPL27A | RL27A | 1.857863643 | 10.85159021 | 2.125886999 | 0.038066267 | -3.914785427 |

| | | | | | | | |
|---|---|---|---|---|---|---|---|
| P06576 | ATP5F1B | ATPB | 1.854788873 | 11.92407979 | 2.645942144 | 0.010582118 | -2.880076461 |
| Q16695 | H3-4 | H31T | 1.81043869 | 10.26811768 | 2.746531438 | 0.008108009 | -2.651308881 |
| P61764 | STXBP1 | STXB1 | 1.803525023 | 13.40972901 | 2.698994613 | 0.009202225 | -2.760221878 |
| P07339 | CTSD | CATD | 1.782307621 | 12.19535528 | 2.295426069 | 0.025517097 | -3.624950835 |
| P62937 | PPIA | PPIA | 1.76788217 | 11.22550632 | 3.531105338 | 0.000841708 | -0.665322681 |
| P25398 | RPS12 | RS12 | 1.7610693 | 8.911054274 | 2.596323425 | 0.012198737 | -2.937278404 |
| Q96GD0 | PDXP | PLPP | 1.73643778 | 10.49092554 | 2.573468953 | 0.012772982 | -3.040864324 |
| Q9H4G0 | EPB41L1 | E41L1 | 1.734992736 | 10.09244966 | 2.643191359 | 0.010658576 | -2.886241669 |
| O43813 | LANCL1 | LANCL1 | 1.723177964 | 12.55751351 | 2.376701342 | 0.020948539 | -3.459730484 |
| P15313 | ATP6V1B1 | VATB1 | 1.70254785 | 8.509028518 | 2.363605943 | 0.02191433 | -3.425025017 |
| P05141 | SLC25A5 | ADT2 | 1.639366786 | 10.10956464 | 2.025674999 | 0.047713605 | -4.13971633 |
| P78559 | MAP1A | MAP1A | 1.636586279 | 10.07764273 | 2.331123812 | 0.023626627 | -3.519846831 |
| P60880 | SNAP25 | SNP25 | 1.61582302 | 10.94386326 | 2.402433579 | 0.019662954 | -3.406456283 |
| P06312 | IGKV4-1 | KV401 | 1.603210763 | 13.0685114 | 2.183828584 | 0.03322509 | -3.8441265 |
| Q92597 | NDRG1 | NDRG1 | 1.583521865 | 10.274978 | 2.235065587 | 0.029617204 | -3.708051315 |
| P19823 | ITIH2 | ITIH2 | 1.556135041 | 14.69691094 | 2.645077845 | 0.010606087 | -2.882014102 |
| P68366 | TUBA4A | TBA4A | 1.516229248 | 8.182302444 | 2.369716787 | 0.021833795 | -3.376767309 |
| P05154 | SERPINA5 | IPSP | 1.510140379 | 11.45434701 | 2.609988717 | 0.011622076 | -2.960270486 |
| P19827 | ITIH1 | ITIH1 | 1.483379929 | 15.08481182 | 3.475464105 | 0.000998396 | -0.816835482 |
| P40925 | MDH1 | MDHC | 1.38752364 | 13.09243818 | 2.513002045 | 0.014908074 | -3.172367809 |
| P61106 | RAB14 | RAB14 | 1.373091293 | 9.911827099 | 2.14903391 | 0.03609689 | -3.871431123 |
| P04264 | KRT1 | K2C1 | 1.330825833 | 16.88434956 | 2.074189931 | 0.042720136 | -4.05057 |
| Q01105 | SET | SET | 1.236927736 | 12.74758991 | 2.165742079 | 0.03465075 | -3.87879543 |
| P62269 | RPS18 | RS18 | 1.230038755 | 10.80738206 | 2.511644997 | 0.015015784 | -3.142724665 |
| P23515 | OMG | OMGP | 1.217952741 | 10.03831417 | 2.255059358 | 0.0280997 | -3.705267842 |
| P04217 | A1BG | A1BG | 1.204039777 | 14.14436628 | 2.125082574 | 0.038053069 | -3.955852506 |
| P07996 | THBS1 | TSP1 | 1.170317842 | 13.73618878 | 2.510145196 | 0.015016517 | -3.178520515 |
| P0DOX2 | HLA-E | IGA2 | -1.007579279 | 13.8605196 | -2.513668686 | 0.014882871 | -3.170931295 |
| P07196 | NEFL | NFL | -1.099586581 | 12.62641688 | -2.861066373 | 0.005944768 | -2.383127406 |
| P45880 | VDAC2 | VDAC2 | -1.160768006 | 11.73892765 | -2.123455519 | 0.038195092 | -3.958910577 |
| P49321 | NASP | NASP | -1.204298376 | 13.01530712 | -2.11062108 | 0.039331705 | -3.982963983 |

| | | | | | | | |
|---|---|---|---|---|---|---|---|
| P62241 | RPS8 | RS8 | -1.29755053 | 9.619243098 | -2.229650451 | 0.029841619 | -3.755222885 |
| O75636 | FCN3 | FCN3 | -1.343234811 | 11.69849824 | -2.594189864 | 0.012107854 | -2.99524361 |
| P02765 | AHSG | FETUA | -1.37747089 | 11.76711324 | -2.18443578 | 0.033178135 | -3.842958439 |
| P24752 | ACAT1 | THIL | -1.472477426 | 10.23425967 | -2.116681257 | 0.038791393 | -3.971621712 |
| P09104 | ENO2 | ENOG | -1.483498961 | 11.61891811 | -2.736236982 | 0.00833429 | -2.675015485 |
| P27797 | CALR | CALR | -1.542426162 | 10.33747203 | -2.856409589 | 0.006021125 | -2.394187512 |
| Q96AE4 | FUBP1 | FUBP1 | -1.554940837 | 9.616239638 | -2.011944201 | 0.049097495 | -4.163765145 |
| P20851-2 | C4BPB | C4BPB | -1.594437683 | 11.52892585 | -2.064379272 | 0.043674875 | -4.068605515 |
| P15104 | GLUL | GLNA | -1.643650255 | 13.5747292 | -2.224628619 | 0.030196968 | -3.765040774 |
| Q12906-7 | ILF3 | ILF3 | -1.680362952 | 10.81171098 | -2.563834148 | 0.013093424 | -3.061980701 |
| P16112 | ACAN | PGCA | -1.777776686 | 8.198785371 | -2.517510235 | 0.01518009 | -3.077648841 |
| P07900-2 | HSP90AA1 | HS90A | -1.837517824 | 14.11273495 | -2.58691642 | 0.01233765 | -3.011289515 |
| Q9H9Z2 | LIN28A | LN28A | -1.893191819 | 9.50440994 | -2.182748534 | 0.033388215 | -3.807585008 |
| Q04637 | EIF4G1 | IF4G1 | -1.973513859 | 9.520493328 | -2.360330131 | 0.022013989 | -3.429966046 |
| P01817 | IGHV2-5 | HV205 | -1.991188261 | 11.01697612 | -3.221519273 | 0.002136217 | -1.488884128 |
| P0DOY3 | GLC3 | IGLC3 | -2.013239169 | 15.80860887 | -2.334228709 | 0.023235648 | -3.546651688 |
| P49913 | CAMP | CAMP | -2.023139583 | 8.945209748 | -2.435260773 | 0.018962643 | -3.225013652 |
| O95865 | DDAH2 | DDAH2 | -2.081381412 | 8.69504494 | -2.152610526 | 0.036361359 | -3.836972592 |
| Q14520 | HABP2 | HABP2 | -2.166936992 | 9.292677581 | -2.529477373 | 0.01446811 | -3.14307117 |
| P49189 | ALDH9A1 | AL9A1 | -2.169813503 | 8.956597096 | -2.633817617 | 0.011072734 | -2.914764511 |
| Q9NSD9 | FARSB | SYFB | -2.235910687 | 9.390867496 | -2.415027341 | 0.019558196 | -3.232241857 |
| P03952 | KLKB1 | KLKB1 | -2.27976213 | 13.33419021 | -3.335368102 | 0.001524802 | -1.191638918 |
| A0A075B6I4 | IGLV10-54 | LVX54 | -2.345882419 | 9.154401513 | -2.226925737 | 0.030952888 | -3.701673514 |
| P55290 | CDH13 | CAD13 | -2.362565418 | 9.449083416 | -2.740131535 | 0.008471848 | -2.626257568 |
| P01042-2 | KNG1 | KNG1 | -2.600531393 | 11.83735509 | -2.540834815 | 0.013998724 | -3.08230816 |
| P49006 | MARCKSL1 | MRP | -2.627628371 | 8.963972567 | -2.250973949 | 0.028525919 | -3.6253474 |
| P0DP09 | IGKV1D-13 | KV113 | -2.678092311 | 13.65181592 | -3.63499061 | 0.000642709 | -0.416948309 |
| Q5U7I5 | TTR | TTHY | -2.771899259 | 12.18705187 | -2.381975639 | 0.020744516 | -3.450379517 |
| P61981 | YWHAG | 1433G | -2.852527559 | 13.78261501 | -3.098281952 | 0.003054682 | -1.802862823 |
| Q9Y696 | CLIC4 | CLIC4 | -2.934780384 | 8.300412466 | -2.733700842 | 0.008835106 | -2.601383064 |
| P08185 | SERPINA6 | CBG | -3.017978393 | 9.106813744 | -2.56934623 | 0.014191214 | -2.920186341 |

| | | | | | | | |
|---|---|---|---|---|---|---|---|
| A2NJV5 | IGKV2-29 | KV229 | -3.86797106 | 8.920817656 | -2.152630744 | 0.036359682 | -3.723827721 |
| P14550 | AKR1A1 | AK1A1 | -5.241864405 | 7.585266854 | -5.822662849 | 5.96435E-07 | 5.464683385 |

#### Supplemental Table 4. All Quantified proteins

| UniProt | Gene Symbol | logFC | AveExpr | t | P.Value | adj.P.Val | B |
|---|---|---|---|---|---|---|---|
| P14550 | AKR1A1 | -5.24186 | 7.585267 | -5.82266 | 5.96E-07 | 0.000508 | 5.464683 |
| P02656 | APOC3 | 3.17308 | 12.92227 | 4.147253 | 0.000117 | 0.04964 | 1.102525 |
| P55209 | NAP1L1 | 3.868894 | 12.57393 | 3.809774 | 0.000356 | 0.10107 | 0.111642 |
| P0DP09 | #N/D | -2.67809 | 13.65182 | -3.63499 | 0.000643 | 0.118195 | -0.41695 |
| P30626 | SRI | 3.66983 | 7.800031 | 3.698254 | 0.000694 | 0.118195 | -0.47114 |
| P62937 | PPIA | 1.767882 | 11.22551 | 3.531105 | 0.000842 | 0.119522 | -0.66532 |
| P19827 | ITIH1 | 1.48338 | 15.08481 | 3.475464 | 0.000998 | 0.121519 | -0.81684 |
| P03952 | KLKB1 | -2.27976 | 13.33419 | -3.33537 | 0.001525 | 0.162391 | -1.19164 |
| O75947 | ATP5PD | 2.985635 | 8.722357 | 3.248856 | 0.002006 | 0.18026 | -1.38711 |
| P01817 | #N/D | -1.99119 | 11.01698 | -3.22152 | 0.002136 | 0.18026 | -1.48888 |
| Q15365 | PCBP1 | 2.458728 | 11.13094 | 3.198005 | 0.002327 | 0.18026 | -1.56068 |
| P61981 | YWHAG | -2.85253 | 13.78262 | -3.09828 | 0.003055 | 0.187572 | -1.80286 |
| Q99426 | TBCB | 3.624327 | 8.295429 | 3.141189 | 0.003064 | 0.187572 | -1.77299 |
| P68871 | HBB | 2.559711 | 14.57697 | 3.09516 | 0.003082 | 0.187572 | -1.81071 |
| Q9UJU6 | DBNL | 2.152096 | 10.24301 | 3.037981 | 0.003654 | 0.205862 | -1.95793 |
| P31327 | CPS1 | 2.342949 | 9.318571 | 3.013866 | 0.003885 | 0.205862 | -2.01308 |
| A0A075B6. | #N/D | 2.899648 | 10.90519 | 3.001517 | 0.004108 | 0.205862 | -2.05673 |
| P32004 | L1CAM | 2.287076 | 9.227821 | 2.986836 | 0.004378 | 0.207232 | -2.03683 |
| P07196 | NEFL | -1.09959 | 12.62642 | -2.86107 | 0.005945 | 0.2565 | -2.38313 |
| P27797 | CALR | -1.54243 | 10.33747 | -2.85641 | 0.006021 | 0.2565 | -2.39419 |
| Q08211 | DHX9 | 2.121782 | 13.43428 | 2.837661 | 0.006338 | 0.257134 | -2.43858 |
| Q16695 | H3-4 | 1.810439 | 10.26812 | 2.746531 | 0.008108 | 0.27518 | -2.65131 |
| P09104 | ENO2 | -1.4835 | 11.61892 | -2.73624 | 0.008334 | 0.27518 | -2.67502 |
| P55290 | CDH13 | -2.36257 | 9.449083 | -2.74013 | 0.008472 | 0.27518 | -2.62626 |
| Q9Y696 | CLIC4 | -2.93478 | 8.300412 | -2.7337 | 0.008835 | 0.27518 | -2.60138 |
| P61764 | STXBP1 | 1.803525 | 13.40973 | 2.698995 | 0.009202 | 0.27518 | -2.76022 |
| P06576 | ATP5F1B | 1.854789 | 11.92408 | 2.645942 | 0.010582 | 0.27518 | -2.88008 |
| P19823 | ITIH2 | 1.556135 | 14.69691 | 2.645078 | 0.010606 | 0.27518 | -2.88201 |

| | | | | | | | |
|---|---|---|---|---|---|---|---|
| Q9H4G0 | EPB41L1 | 1.734993 | 10.09245 | 2.643191 | 0.010659 | 0.27518 | -2.88624 |
| Q96FC7 | PHYHIPL | 2.809929 | 10.07371 | 2.630384 | 0.011069 | 0.27518 | -2.88478 |
| P49189 | ALDH9A1 | -2.16981 | 8.956597 | -2.63382 | 0.011073 | 0.27518 | -2.91476 |
| P05154 | SERPINA5 | 1.51014 | 11.45435 | 2.609989 | 0.011622 | 0.27518 | -2.96027 |
| O75636 | FCN3 | -1.34323 | 11.6985 | -2.59419 | 0.012108 | 0.27518 | -2.99524 |
| P25398 | RPS12 | 1.761069 | 8.911054 | 2.596323 | 0.012199 | 0.27518 | -2.93728 |
| P07900-2 | #N/D | -1.83752 | 14.11273 | -2.58692 | 0.012338 | 0.27518 | -3.01129 |
| Q96GD0 | PDXP | 1.736438 | 10.49093 | 2.573469 | 0.012773 | 0.27518 | -3.04086 |
| Q12906-7 | #N/D | -1.68036 | 10.81171 | -2.56383 | 0.013093 | 0.27518 | -3.06198 |
| P02533 | KRT14 | 2.020689 | 11.09842 | 2.546941 | 0.013673 | 0.27518 | -3.09886 |
| Q7KZF4 | SND1 | 1.976169 | 9.615375 | 2.550056 | 0.013732 | 0.27518 | -3.03672 |
| P01042-2 | #N/D | -2.60053 | 11.83736 | -2.54083 | 0.013999 | 0.27518 | -3.08231 |
| P08185 | SERPINA6 | -3.01798 | 9.106814 | -2.56935 | 0.014191 | 0.27518 | -2.92019 |
| Q14520 | HABP2 | -2.16694 | 9.292678 | -2.52948 | 0.014468 | 0.27518 | -3.14307 |
| P0DOX2 | #N/D | -1.00758 | 13.86052 | -2.51367 | 0.014883 | 0.27518 | -3.17093 |
| P40925 | MDH1 | 1.387524 | 13.09244 | 2.513002 | 0.014908 | 0.27518 | -3.17237 |
| P62269 | RPS18 | 1.230039 | 10.80738 | 2.511645 | 0.015016 | 0.27518 | -3.14272 |
| P07996 | THBS1 | 1.170318 | 13.73619 | 2.510145 | 0.015017 | 0.27518 | -3.17852 |
| P16112 | #N/D | -1.77778 | 8.198785 | -2.51751 | 0.01518 | 0.27518 | -3.07765 |
| P02538 | KRT6A | 2.931508 | 11.77848 | 2.496951 | 0.015705 | 0.278771 | -3.14916 |
| P35908 | KRT2 | 2.668528 | 14.98502 | 2.48253 | 0.016102 | 0.27998 | -3.23771 |
| P04040 | CAT | 1.915414 | 8.854698 | 2.484371 | 0.01673 | 0.285074 | -3.05364 |
| P12268 | IMPDH2 | 2.521064 | 9.058515 | 2.458895 | 0.017561 | 0.29337 | -3.14581 |
| P49913 | CAMP | -2.02314 | 8.94521 | -2.43526 | 0.018963 | 0.304597 | -3.22501 |
| Q06033 | ITIH3 | 2.879376 | 13.55378 | 2.41081 | 0.01926 | 0.304597 | -3.38902 |
| Q9NSD9 | FARSB | -2.23591 | 9.390867 | -2.41503 | 0.019558 | 0.304597 | -3.23224 |
| P60880 | SNAP25 | 1.615823 | 10.94386 | 2.402434 | 0.019663 | 0.304597 | -3.40646 |
| Q5U7I5 | #N/D | -2.7719 | 12.18705 | -2.38198 | 0.020745 | 0.312599 | -3.45038 |
| O43813 | LANCL1 | 1.723178 | 12.55751 | 2.376701 | 0.020949 | 0.312599 | -3.45973 |
| P68366 | TUBA4A | 1.516229 | 8.182302 | 2.369717 | 0.021834 | 0.312599 | -3.37677 |
| P15313 | ATP6V1B1 | 1.702548 | 8.509029 | 2.363606 | 0.021914 | 0.312599 | -3.42503 |

| | | | | | | | |
|---|---|---|---|---|---|---|---|
| Q04637 | EIF4G1 | -1.97351 | 9.520493 | -2.36033 | 0.022014 | 0.312599 | -3.42997 |
| P0DOY3 | #N/D | -2.01324 | 15.80861 | -2.33423 | 0.023236 | 0.324537 | -3.54665 |
| P78559 | MAP1A | 1.636586 | 10.07764 | 2.331124 | 0.023627 | 0.324676 | -3.51985 |
| P63104 | YWHAZ | 0.98315 | 13.41142 | 2.30452 | 0.024965 | 0.337625 | -3.6067 |
| P07339 | CTSD | 1.782308 | 12.19536 | 2.295426 | 0.025517 | 0.339696 | -3.62495 |
| P23515 | OMG | 1.217953 | 10.03831 | 2.255059 | 0.0281 | 0.363703 | -3.70527 |
| P49006 | MARCKSL1 | -2.62763 | 8.963973 | -2.25097 | 0.028526 | 0.363703 | -3.62535 |
| Q15717 | ELAVL1 | 2.379461 | 11.8003 | 2.253267 | 0.028906 | 0.363703 | -3.54208 |
| Q92597 | NDRG1 | 1.583522 | 10.27498 | 2.235066 | 0.029617 | 0.363703 | -3.70805 |
| P62241 | RPS8 | -1.29755 | 9.619243 | -2.22965 | 0.029842 | 0.363703 | -3.75522 |
| P15104 | GLUL | -1.64365 | 13.57473 | -2.22463 | 0.030197 | 0.363703 | -3.76504 |
| P49368 | CCT3 | 2.318894 | 10.62952 | 2.223062 | 0.030309 | 0.363703 | -3.7681 |
| A0A075B6I | #N/D | -2.34588 | 9.154402 | -2.22693 | 0.030953 | 0.366276 | -3.70167 |
| P08133 | ANXA6 | 0.955735 | 11.75849 | 2.19801 | 0.032144 | 0.370942 | -3.81677 |
| P02765 | AHSG | -1.37747 | 11.76711 | -2.18444 | 0.033178 | 0.370942 | -3.84296 |
| P06312 | #N/D | 1.603211 | 13.06851 | 2.183829 | 0.033225 | 0.370942 | -3.84413 |
| Q9H9Z2 | LIN28A | -1.89319 | 9.50441 | -2.18275 | 0.033388 | 0.370942 | -3.80759 |
| P29762 | CRABP1 | 2.25792 | 11.46855 | 2.17998 | 0.033524 | 0.370942 | -3.85152 |
| Q01105 | SET | 1.236928 | 12.74759 | 2.165742 | 0.034651 | 0.373252 | -3.8788 |
| P38159 | RBMX | 2.15587 | 13.33236 | 2.158354 | 0.035248 | 0.373252 | -3.89289 |
| P02747 | C1QC | 2.205541 | 14.27094 | 2.15482 | 0.035537 | 0.373252 | -3.89961 |
| P61106 | RAB14 | 1.373091 | 9.911827 | 2.149034 | 0.036097 | 0.373252 | -3.87143 |
| A2NJV5 | #N/D | -3.86797 | 8.920818 | -2.15263 | 0.03636 | 0.373252 | -3.72383 |
| O95865 | DDAH2 | -2.08138 | 8.695045 | -2.15261 | 0.036361 | 0.373252 | -3.83697 |
| P04217 | A1BG | 1.20404 | 14.14437 | 2.125083 | 0.038053 | 0.378398 | -3.95585 |
| P46776 | RPL27A | 1.857864 | 10.85159 | 2.125887 | 0.038066 | 0.378398 | -3.91479 |
| P45880 | VDAC2 | -1.16077 | 11.73893 | -2.12346 | 0.038195 | 0.378398 | -3.95891 |
| P24752 | ACAT1 | -1.47248 | 10.23426 | -2.11668 | 0.038791 | 0.379888 | -3.97162 |
| P49321 | NASP | -1.2043 | 13.01531 | -2.11062 | 0.039332 | 0.380802 | -3.98296 |
| Q15102 | PAFAH1B3 | 2.000511 | 9.201983 | 2.107298 | 0.039803 | 0.381034 | -3.89507 |
| P04264 | KRT1 | 1.330826 | 16.88435 | 2.07419 | 0.04272 | 0.401962 | -4.05057 |

| | | | | | | | |
|---|---|---|---|---|---|---|---|
| P18206 | VCL | 0.814422 | 12.66382 | 2.071291 | 0.043 | 0.401962 | -4.05591 |
| P20851-2 | #N/D | -1.59444 | 11.52893 | -2.06438 | 0.043675 | 0.401962 | -4.06861 |
| P31948 | STIP1 | 0.970456 | 10.97486 | 2.062335 | 0.043876 | 0.401962 | -4.07235 |
| P42704 | LRPPRC | 1.88803 | 13.36178 | 2.048148 | 0.045295 | 0.410547 | -4.09829 |
| P05141 | SLC25A5 | 1.639367 | 10.10956 | 2.025675 | 0.047714 | 0.427916 | -4.13972 |
| Q96AE4 | FUBP1 | -1.55494 | 9.61624 | -2.01194 | 0.049097 | 0.432229 | -4.16377 |
| P30086 | PEBP1 | 0.962116 | 12.34105 | 2.010918 | 0.049209 | 0.432229 | -4.16561 |
| Q99747 | NAPG | 1.016536 | 8.259074 | 1.990487 | 0.05157 | 0.448343 | -4.20271 |
| Q96KP4 | CNDP2 | 0.986495 | 11.48894 | 1.979253 | 0.052764 | 0.453135 | -4.22203 |
| Q9UBC3 | DNMT3B | 1.833932 | 11.72397 | 1.975629 | 0.053185 | 0.453135 | -4.22844 |
| P09661 | SNRPA1 | -2.40483 | 11.24558 | -1.9561 | 0.055502 | 0.468192 | -4.26281 |
| Q96F85 | CNRIP1 | -1.11688 | 10.63719 | -1.93852 | 0.05766 | 0.480248 | -4.29349 |
| P20916 | MAG | 1.59501 | 9.172305 | 1.925644 | 0.059913 | 0.480248 | -4.18743 |
| Q00610 | CLTC | -1.53948 | 13.78944 | -1.92074 | 0.059916 | 0.480248 | -4.32427 |
| P0DOX8 | #N/D | 2.208878 | 13.34101 | 1.918398 | 0.060313 | 0.480248 | -4.32879 |
| P11940 | PABPC1 | 1.270697 | 9.661148 | 1.915353 | 0.060808 | 0.480248 | -4.2913 |
| P21333 | FLNA | -1.13035 | 13.07842 | -1.9064 | 0.06179 | 0.480248 | -4.34893 |
| P14866 | HNRNPL | -1.17307 | 12.72325 | -1.9035 | 0.062174 | 0.480248 | -4.35388 |
| Q9Y6R7 | FCGBP | -0.9862 | 13.98588 | -1.90212 | 0.062358 | 0.480248 | -4.35625 |
| Q5IS61 | #N/D | -1.32832 | 8.614239 | -1.90685 | 0.062363 | 0.480248 | -4.30792 |
| Q9UNZ2 | NSFL1C | 1.431329 | 8.71179 | 1.904433 | 0.062568 | 0.480248 | -4.31144 |
| Q8IV08 | PLD3 | 1.392241 | 9.113681 | 1.891565 | 0.063876 | 0.485287 | -4.33117 |
| A0A0A0MS | #N/D | 1.292122 | 10.69979 | 1.883637 | 0.064865 | 0.485287 | -4.38772 |
| P05783 | KRT18 | 1.686804 | 11.19722 | 1.884579 | 0.064933 | 0.485287 | -4.38701 |
| P31146 | CORO1A | 1.742527 | 9.870592 | 1.847371 | 0.070341 | 0.510247 | -4.34516 |
| P01860 | #N/D | -1.49925 | 14.17562 | -1.84428 | 0.070491 | 0.510247 | -4.45384 |
| P02792 | FTL | 1.469724 | 9.643032 | 1.844538 | 0.070759 | 0.510247 | -4.34977 |
| P02675 | FGB | -0.92053 | 14.18233 | -1.83283 | 0.072202 | 0.510247 | -4.47284 |
| O00264 | PGRMC1 | -1.86236 | 9.27944 | -1.83554 | 0.072327 | 0.510247 | -4.25509 |
| P22061 | PCMT1 | 1.051925 | 10.41461 | 1.830732 | 0.07252 | 0.510247 | -4.47631 |
| O75781 | PALM | -1.11353 | 8.620566 | -1.82925 | 0.072946 | 0.510247 | -4.4352 |

| | | | | | | | |
|---|---|---|---|---|---|---|---|
| P07237 | P4HB | 1.747336 | 10.09498 | 1.829182 | 0.073063 | 0.510247 | -4.344 |
| P40227 | CCT6A | -0.97803 | 12.26876 | -1.81679 | 0.074659 | 0.517147 | -4.49929 |
| P11137 | MAP2 | -1.05746 | 10.40085 | -1.80793 | 0.076045 | 0.522501 | -4.51381 |
| P25788 | PSMA3 | -1.36365 | 9.356898 | -1.8013 | 0.077637 | 0.524641 | -4.44319 |
| P25789 | PSMA4 | 1.303116 | 7.883878 | 1.79935 | 0.078197 | 0.524641 | -4.39312 |
| Q92522 | H1-10 | 0.914433 | 9.944587 | 1.791709 | 0.07864 | 0.524641 | -4.54024 |
| P07954 | FH | 1.442931 | 11.84092 | 1.786745 | 0.07955 | 0.524641 | -4.5486 |
| P48539 | PCP4 | 1.091559 | 8.670542 | 1.787383 | 0.07955 | 0.524641 | -4.46433 |
| P30101 | PDIA3 | 1.315248 | 12.87952 | 1.781456 | 0.080318 | 0.524641 | -4.55683 |
| P33993 | MCM7 | -1.21701 | 8.945521 | -1.77786 | 0.081016 | 0.524641 | -4.51803 |
| P02679-2 | #N/D | -0.83375 | 11.17822 | -1.77474 | 0.081434 | 0.524641 | -4.56766 |
| O75390 | CS | 1.50117 | 11.12214 | 1.770679 | 0.082114 | 0.524641 | -4.57419 |
| P07358 | C8B | 1.173368 | 13.46551 | 1.768307 | 0.082514 | 0.524641 | -4.57799 |
| P06733 | ENO1 | 0.732126 | 12.59836 | 1.754104 | 0.084941 | 0.524993 | -4.60069 |
| P35611 | ADD1 | -2.1092 | 8.364787 | -1.77009 | 0.085086 | 0.524993 | -4.28184 |
| P00441 | SOD1 | 1.259499 | 11.21279 | 1.751708 | 0.085458 | 0.524993 | -4.55956 |
| Q9GZV7 | HAPLN2 | -1.01426 | 13.03861 | -1.74631 | 0.086298 | 0.524993 | -4.61307 |
| P04259 | KRT6B | 1.329004 | 11.57937 | 1.744234 | 0.086765 | 0.524993 | -4.61663 |
| P62258 | YWHAE | 0.759529 | 12.79759 | 1.743269 | 0.086833 | 0.524993 | -4.6179 |
| P19338 | NCL | -1.27515 | 14.24 | -1.74299 | 0.086883 | 0.524993 | -4.61834 |
| P02760 | AMBP | 0.81752 | 13.73303 | 1.73699 | 0.087945 | 0.526848 | -4.62782 |
| A0A075B6I | #N/D | -2.17843 | 8.760973 | -1.73492 | 0.088865 | 0.526848 | -4.4471 |
| Q7L099-4 | #N/D | 1.062517 | 10.43573 | 1.730845 | 0.089045 | 0.526848 | -4.63751 |
| Q02543 | RPL18A | -1.71624 | 12.91705 | -1.71704 | 0.091556 | 0.537971 | -4.65915 |
| Q6YN16 | HSDL2 | -1.63741 | 9.242755 | -1.71313 | 0.092713 | 0.538318 | -4.62057 |
| P19367 | HK1 | 0.820265 | 11.89524 | 1.7099 | 0.092879 | 0.538318 | -4.67029 |
| P00742 | F10 | 1.42957 | 12.22352 | 1.703951 | 0.094095 | 0.541683 | -4.67976 |
| Q9UNQ0 | ABCG2 | -2.02851 | 11.29824 | -1.69873 | 0.095415 | 0.545593 | -4.64272 |
| Q96C19 | EFHD2 | 2.034648 | 8.816843 | 1.698055 | 0.09634 | 0.547213 | -4.46527 |
| P15880 | RPS2 | -1.52568 | 12.70417 | -1.66823 | 0.100912 | 0.563202 | -4.73445 |
| Q16555 | DPYSL2 | 0.818715 | 14.86964 | 1.668045 | 0.100949 | 0.563202 | -4.73473 |

| | | | | | | | |
|---|---|---|---|---|---|---|---|
| P08238 | HSP90AB1 | 1.023021 | 13.56843 | 1.667095 | 0.101138 | 0.563202 | -4.73617 |
| P23284 | PPIB | 1.540917 | 11.31656 | 1.664728 | 0.101934 | 0.563949 | -4.59749 |
| O60506 | SYNCRIP | -1.48017 | 8.844164 | -1.65995 | 0.103259 | 0.567591 | -4.55851 |
| P11021 | HSPA5 | -0.83525 | 14.82701 | -1.65204 | 0.104182 | 0.568081 | -4.75898 |
| P31025 | LCN1 | -1.24991 | 14.44478 | -1.64769 | 0.105075 | 0.568081 | -4.76554 |
| P09211 | GSTP1 | 1.86498 | 13.54278 | 1.652414 | 0.105348 | 0.568081 | -4.38571 |
| P29401 | TKT | 0.759468 | 12.31095 | 1.6383 | 0.107026 | 0.573495 | -4.77965 |
| Q12905 | ILF2 | 1.239014 | 9.328399 | 1.630452 | 0.108889 | 0.579836 | -4.74511 |
| Q9H4G4 | GLIPR2 | 1.1232 | 9.978347 | 1.624969 | 0.109948 | 0.581837 | -4.79969 |
| P07741 | APRT | 2.100893 | 8.592073 | 1.617685 | 0.112228 | 0.588364 | -4.66681 |
| Q9Y617 | PSAT1 | -1.0299 | 11.24982 | -1.60799 | 0.113522 | 0.588364 | -4.82467 |
| Q9BY11 | PACSIN1 | -1.44739 | 11.03934 | -1.60765 | 0.113596 | 0.588364 | -4.82517 |
| P50991 | CCT4 | -0.82662 | 13.00041 | -1.60597 | 0.113965 | 0.588364 | -4.82764 |
| Q15257-2 | #N/D | -1.67656 | 9.172995 | -1.60548 | 0.114634 | 0.588364 | -4.74146 |
| P00751 | CFB | 0.952123 | 13.86732 | 1.598254 | 0.115674 | 0.590143 | -4.83897 |
| P07910 | HNRNPC | -0.70837 | 10.97799 | -1.59092 | 0.117316 | 0.590892 | -4.84968 |
| P78371 | CCT2 | 1.197624 | 10.10086 | 1.587266 | 0.118143 | 0.590892 | -4.85501 |
| Q9NRW1 | RAB6B | 1.146801 | 9.368933 | 1.586425 | 0.118546 | 0.590892 | -4.80943 |
| P61978 | HNRNPK | 1.242708 | 11.47478 | 1.584267 | 0.118824 | 0.590892 | -4.85937 |
| P60174 | TPI1 | 1.628026 | 13.33968 | 1.580061 | 0.119785 | 0.590892 | -4.86548 |
| Q15185 | PTGES3 | 1.7637 | 14.08679 | 1.578645 | 0.120214 | 0.590892 | -4.86764 |
| P34932 | HSPA4 | 0.85254 | 12.19409 | 1.576187 | 0.120675 | 0.590892 | -4.87109 |
| P08519 | #N/D | -1.30865 | 15.54961 | -1.5706 | 0.121968 | 0.591454 | -4.87915 |
| Q9Y2W1 | THRAP3 | -1.2005 | 8.282812 | -1.56761 | 0.123106 | 0.591454 | -4.73731 |
| A0A075B6I | #N/D | -0.67924 | 13.46962 | -1.56564 | 0.123127 | 0.591454 | -4.8863 |
| P02790 | HPX | 1.240968 | 14.96138 | 1.563763 | 0.123567 | 0.591454 | -4.88899 |
| P28838 | LAP3 | -0.91675 | 11.12429 | -1.54986 | 0.126868 | 0.601967 | -4.90888 |
| Q9P2U7 | SLC17A7 | 1.447078 | 10.35073 | 1.549461 | 0.127176 | 0.601967 | -4.90963 |
| P27918 | CFP | -1.03459 | 11.216 | -1.5443 | 0.128208 | 0.603498 | -4.91678 |
| Q9Y2T3 | GDA | -1.16805 | 10.08686 | -1.53937 | 0.129617 | 0.605196 | -4.92394 |
| Q15435 | PPP1R7 | 1.505083 | 8.920446 | 1.539799 | 0.129989 | 0.605196 | -4.77612 |

| | | | | | | | |
|---|---|---|---|---|---|---|---|
| P36578 | RPL4 | 0.603343 | 12.0117 | 1.51811 | 0.134672 | 0.613059 | -4.95368 |
| Q14894 | CRYM | -0.93056 | 12.35016 | -1.51672 | 0.135022 | 0.613059 | -4.95562 |
| P30153 | PPP2R1A | -1.18648 | 10.68953 | -1.513 | 0.135963 | 0.613059 | -4.96081 |
| P23471 | PTPRZ1 | -2.06411 | 10.95353 | -1.51287 | 0.136208 | 0.613059 | -4.96113 |
| P01718 | #N/D | 0.997166 | 12.55539 | 1.508805 | 0.137029 | 0.613059 | -4.96664 |
| P10515 | DLAT | 0.973057 | 8.7176 | 1.507879 | 0.137478 | 0.613059 | -4.96806 |
| P39687 | ANP32A | 1.234318 | 9.050789 | 1.507361 | 0.137839 | 0.613059 | -4.82012 |
| Q6PCE3 | PGM2L1 | 1.235084 | 10.75219 | 1.505181 | 0.137957 | 0.613059 | -4.97168 |
| P04114 | APOB | 1.286217 | 15.2199 | 1.502719 | 0.138589 | 0.613059 | -4.97509 |
| P37837 | TALDO1 | 0.779959 | 13.06675 | 1.501576 | 0.138884 | 0.613059 | -4.97667 |
| P05026 | ATP1B1 | 1.439707 | 9.719583 | 1.500999 | 0.139593 | 0.613059 | -4.73771 |
| P0DOX5 | #N/D | -0.79252 | 14.50645 | -1.48744 | 0.142568 | 0.622912 | -4.99615 |
| O94819 | KBTBD11 | 1.006457 | 9.342712 | 1.476724 | 0.145514 | 0.624495 | -4.96268 |
| P15814 | IGLL1 | 2.426766 | 11.9045 | 1.475895 | 0.146191 | 0.624495 | -4.86226 |
| P18124 | RPL7 | -0.79192 | 10.99238 | -1.47086 | 0.146985 | 0.624495 | -5.01877 |
| P55072 | VCP | 0.912515 | 13.88593 | 1.470624 | 0.147049 | 0.624495 | -5.01909 |
| P26583 | HMGB2 | -1.0876 | 9.427067 | -1.4701 | 0.14719 | 0.624495 | -5.0198 |
| P78347 | GTF2I | -0.89544 | 9.347322 | -1.46741 | 0.148021 | 0.624495 | -5.02349 |
| Q99878 | H2AC14 | -1.41046 | 8.686371 | -1.47092 | 0.148061 | 0.624495 | -4.86909 |
| P04181 | OAT | 1.050556 | 9.027395 | 1.461254 | 0.149804 | 0.626015 | -5.03185 |
| P21283 | ATP6V1C1 | 1.382523 | 9.48756 | 1.460936 | 0.149891 | 0.626015 | -4.98397 |
| P11586 | MTHFD1 | 2.835612 | 11.82919 | 1.457259 | 0.150778 | 0.62665 | -5.03719 |
| P01619 | #N/D | -1.0446 | 14.00764 | -1.45119 | 0.152802 | 0.630762 | -4.89467 |
| O76070 | SNCG | -0.76736 | 10.89826 | -1.44801 | 0.153249 | 0.630762 | -5.04957 |
| P51149 | RAB7A | 0.854597 | 8.780581 | 1.430454 | 0.158521 | 0.649328 | -4.9215 |
| P06396 | GSN | -0.8942 | 14.7431 | -1.42067 | 0.161016 | 0.651422 | -5.08584 |
| Q14624-3 | #N/D | -1.35133 | 8.418841 | -1.43308 | 0.161125 | 0.651422 | -4.91903 |
| P06753-2 | #N/D | 1.187925 | 11.55725 | 1.421506 | 0.161326 | 0.651422 | -5.08483 |
| P29966 | MARCKS | 1.131078 | 12.49708 | 1.410685 | 0.163927 | 0.656912 | -5.09893 |
| P02649 | APOE | 0.991462 | 16.1627 | 1.407359 | 0.164906 | 0.656912 | -5.10327 |
| P26373 | RPL13 | -0.82323 | 10.14526 | -1.40664 | 0.165117 | 0.656912 | -5.1042 |

| | | | | | | | |
|---|---|---|---|---|---|---|---|
| P02788 | LTF | 1.647335 | 9.520811 | 1.401829 | 0.166751 | 0.656912 | -4.99309 |
| Q92945 | KHSRP | 1.007717 | 11.19284 | 1.401023 | 0.166783 | 0.656912 | -5.11151 |
| P22626 | HNRNPA2B1 | 0.64708 | 12.6985 | 1.397656 | 0.167787 | 0.656912 | -5.11588 |
| Q9BWD1-2 | #N/D | 1.419905 | 7.847079 | 1.402206 | 0.168305 | 0.656912 | -4.76537 |
| P13645 | KRT10 | -0.82465 | 15.33348 | -1.39409 | 0.168854 | 0.656912 | -5.12048 |
| P08708 | RPS17 | 1.092561 | 7.699005 | 1.389671 | 0.17112 | 0.662701 | -4.90424 |
| Q15366 | PCBP2 | 1.33697 | 12.64187 | 1.381227 | 0.172754 | 0.663658 | -5.13704 |
| Q16658 | FSCN1 | 0.833778 | 13.04135 | 1.376468 | 0.174213 | 0.663658 | -5.14312 |
| P38117 | ETFB | -1.17772 | 8.695554 | -1.37496 | 0.175475 | 0.663658 | -5.09591 |
| P62280 | RPS11 | 0.849537 | 11.12395 | 1.372289 | 0.175503 | 0.663658 | -5.14845 |
| P02745 | C1QA | -1.41934 | 10.24676 | -1.37252 | 0.175859 | 0.663658 | -5.03005 |
| P0C0S5 | H2AZ1 | -1.03768 | 10.02911 | -1.37043 | 0.17608 | 0.663658 | -5.15082 |
| P63000 | RAC1 | 0.929552 | 11.00234 | 1.368046 | 0.17682 | 0.663658 | -5.15385 |
| A0A0C4DH | #N/D | 1.20566 | 14.37611 | 1.362964 | 0.178407 | 0.666678 | -5.16029 |
| Q04695 | KRT17 | -1.10262 | 10.32094 | -1.35935 | 0.179856 | 0.66916 | -5.04642 |
| P0DMV9 | HSPA1A | 0.595388 | 12.81602 | 1.35392 | 0.181258 | 0.671443 | -5.17169 |
| P16152 | CBR1 | -0.7648 | 14.15987 | -1.34158 | 0.185205 | 0.680444 | -5.18715 |
| P09417 | QDPR | 0.976536 | 11.79198 | 1.339356 | 0.185922 | 0.680444 | -5.18991 |
| Q9H115 | NAPB | 0.92488 | 9.074792 | 1.340551 | 0.186323 | 0.680444 | -5.06947 |
| Q9UMF0 | ICAM5 | 0.870793 | 9.084601 | 1.336393 | 0.186882 | 0.680444 | -5.1936 |
| P06748 | NPM1 | 1.527208 | 9.025254 | 1.338739 | 0.187958 | 0.680548 | -5.03592 |
| P12004 | PCNA | -0.66167 | 11.20119 | -1.32905 | 0.189276 | 0.680548 | -5.20269 |
| P04004 | VTN | 0.772947 | 14.98777 | 1.32896 | 0.189307 | 0.680548 | -5.20281 |
| O75475 | PSIP1 | 1.024313 | 10.76389 | 1.323266 | 0.191181 | 0.682949 | -5.20983 |
| P01594 | #N/D | -1.05395 | 13.40637 | -1.32369 | 0.191578 | 0.682949 | -5.08996 |
| Q15424 | SAFB | 0.835907 | 10.29881 | 1.311247 | 0.195281 | 0.689684 | -5.17484 |
| Q14974 | KPNB1 | 1.473955 | 12.05301 | 1.310601 | 0.195398 | 0.689684 | -5.22535 |
| Q96PD5 | PGLYRP2 | 0.73221 | 13.17715 | 1.30912 | 0.195896 | 0.689684 | -5.22715 |
| Q9NUQ9 | CYRIB | -1.04403 | 9.681427 | -1.2989 | 0.199664 | 0.700058 | -5.23947 |
| Q99798 | ACO2 | -0.69318 | 13.35696 | -1.29056 | 0.202214 | 0.705965 | -5.24961 |
| P35637 | FUS | 0.862724 | 9.460308 | 1.288551 | 0.203006 | 0.705965 | -5.252 |

| | | | | | | | |
|---|---|---|---|---|---|---|---|
| P04075 | ALDOA | -0.6143 | 13.09657 | -1.28574 | 0.203881 | 0.706125 | -5.2554 |
| A0A0B4J1Y | #N/D | 1.08647 | 9.589963 | 1.281246 | 0.20621 | 0.708642 | -5.10419 |
| P35858 | IGFALS | -0.88108 | 11.34869 | -1.27887 | 0.206271 | 0.708642 | -5.26361 |
| Q9Y6I3 | EPN1 | 1.370006 | 8.193578 | 1.269779 | 0.210927 | 0.721566 | -4.82252 |
| P01024 | C3 | 0.776486 | 15.31739 | 1.263422 | 0.211727 | 0.721566 | -5.28192 |
| Q8NC51 | SERBP1 | -1.49361 | 11.27405 | -1.25898 | 0.213314 | 0.724079 | -5.28714 |
| A0A0C4DH | #N/D | 1.014923 | 11.05438 | 1.255844 | 0.214442 | 0.725018 | -5.29083 |
| P02689 | PMP2 | 0.920602 | 10.41906 | 1.249836 | 0.216612 | 0.725485 | -5.29785 |
| P39019 | RPS19 | 1.21326 | 12.71185 | 1.248161 | 0.21722 | 0.725485 | -5.2998 |
| O75891 | ALDH1L1 | 0.944219 | 12.21429 | 1.245382 | 0.218232 | 0.725485 | -5.30304 |
| P30084 | ECHS1 | -0.58344 | 10.15609 | -1.24333 | 0.218981 | 0.725485 | -5.30542 |
| O14594 | NCAN | 0.857325 | 9.186149 | 1.243711 | 0.219358 | 0.725485 | -5.14728 |
| P09622 | DLD | 1.433765 | 8.688977 | 1.244922 | 0.219689 | 0.725485 | -4.86842 |
| Q13885 | TUBB2A | -0.89469 | 12.3616 | -1.23665 | 0.221433 | 0.728068 | -5.31316 |
| O94856 | NFASC | 0.768232 | 13.27301 | 1.233564 | 0.222572 | 0.728068 | -5.31672 |
| Q07955 | SRSF1 | -0.7552 | 11.42114 | -1.23231 | 0.223035 | 0.728068 | -5.31816 |
| Q9UBB6 | NCDN | 1.068023 | 8.646641 | 1.228196 | 0.224758 | 0.728723 | -5.2724 |
| P63244 | RACK1 | 0.811842 | 12.16177 | 1.225427 | 0.225597 | 0.728723 | -5.32606 |
| P04179 | SOD2 | 0.755487 | 10.77234 | 1.224881 | 0.225801 | 0.728723 | -5.32669 |
| P05546 | SERPIND1 | 0.79105 | 11.79322 | 1.218203 | 0.228308 | 0.734032 | -5.33431 |
| O94760 | DDAH1 | -0.86239 | 9.840467 | -1.21274 | 0.230466 | 0.737032 | -5.28994 |
| P62899 | RPL31 | 1.27295 | 12.22519 | 1.211167 | 0.230971 | 0.737032 | -5.3423 |
| P05023 | ATP1A1 | -0.82308 | 13.61348 | -1.20826 | 0.232078 | 0.73707 | -5.34559 |
| P62942 | FKBP1A | 1.176334 | 10.83055 | 1.206597 | 0.232713 | 0.73707 | -5.34747 |
| P02751-8 | #N/D | 0.589004 | 15.47018 | 1.203066 | 0.234066 | 0.738607 | -5.35145 |
| Q5IS67 | #N/D | -1.35552 | 8.456854 | -1.1992 | 0.236668 | 0.744064 | -5.19641 |
| Q93050 | ATP6V0A1 | -0.58309 | 10.19313 | -1.18738 | 0.240143 | 0.749876 | -5.36899 |
| P62701 | RPS4X | 0.524021 | 10.53558 | 1.184498 | 0.241271 | 0.749876 | -5.37219 |
| Q9UHY7 | ENOPH1 | -0.81441 | 8.101761 | -1.18378 | 0.242658 | 0.749876 | -5.21314 |
| Q99536 | VAT1 | -0.69303 | 11.20367 | -1.17938 | 0.243287 | 0.749876 | -5.37786 |
| P25311 | AZGP1 | 0.608313 | 12.1082 | 1.178837 | 0.2435 | 0.749876 | -5.37845 |

| | | | | | | | |
|---|---|---|---|---|---|---|---|
| P51649 | ALDH5A1 | 0.541234 | 11.25326 | 1.178084 | 0.243798 | 0.749876 | -5.37928 |
| P05387 | RPLP2 | -0.71625 | 9.837368 | -1.1726 | 0.246064 | 0.751796 | -5.38526 |
| P00367 | GLUD1 | -0.61838 | 13.41429 | -1.16829 | 0.247693 | 0.751796 | -5.39005 |
| P05787 | KRT8 | -0.61819 | 11.88015 | -1.16742 | 0.248041 | 0.751796 | -5.391 |
| P62081 | RPS7 | 0.718127 | 10.42682 | 1.16603 | 0.248596 | 0.751796 | -5.39251 |
| P13861 | PRKAR2A | 0.990465 | 8.585476 | 1.166402 | 0.248834 | 0.751796 | -5.34098 |
| P61026 | RAB10 | -1.1688 | 8.602059 | -1.16587 | 0.250031 | 0.752745 | -5.09154 |
| P02766 | TTR | -1.02464 | 13.95342 | -1.15954 | 0.251207 | 0.75362 | -5.39958 |
| P11217 | PYGM | 0.614258 | 10.22972 | 1.152562 | 0.254038 | 0.756045 | -5.40715 |
| O43301 | HSPA12A | -0.67444 | 10.41348 | -1.14855 | 0.25586 | 0.756045 | -5.41135 |
| P22792 | CPN2 | 0.936888 | 10.29617 | 1.147618 | 0.256057 | 0.756045 | -5.41248 |
| P28072 | PSMB6 | 0.767228 | 9.15565 | 1.147101 | 0.256269 | 0.756045 | -5.41303 |
| P24539 | ATP5PB | -1.06755 | 9.276826 | -1.14868 | 0.256452 | 0.756045 | -5.35976 |
| O00154 | ACOT7 | 0.948432 | 10.65194 | 1.139198 | 0.259521 | 0.758032 | -5.42151 |
| Q16352 | INA | 0.500862 | 13.06932 | 1.137231 | 0.260335 | 0.758032 | -5.42361 |
| Q16623-3 | #N/D | 0.582213 | 9.890803 | 1.137175 | 0.260358 | 0.758032 | -5.42367 |
| P22102 | GART | 1.142325 | 10.28125 | 1.136828 | 0.260685 | 0.758032 | -5.32781 |
| P48643 | CCT5 | -0.76031 | 9.491954 | -1.13299 | 0.262099 | 0.758958 | -5.42813 |
| P84077 | ARF1 | 0.872507 | 9.006425 | 1.131552 | 0.262785 | 0.758958 | -5.37843 |
| P04216 | THY1 | 1.077207 | 9.984486 | 1.127264 | 0.264489 | 0.760927 | -5.43419 |
| P62249 | RPS16 | -0.53398 | 10.28851 | -1.12336 | 0.266129 | 0.760927 | -5.43832 |
| Q13491-4 | #N/D | 0.720095 | 9.101016 | 1.123746 | 0.266146 | 0.760927 | -5.43778 |
| O14576 | DYNC1I1 | -0.922 | 8.850936 | -1.11079 | 0.271545 | 0.762523 | -5.40013 |
| P02749 | APOH | -0.57407 | 12.64195 | -1.10906 | 0.272196 | 0.762523 | -5.45331 |
| Q92777 | SYN2 | 1.060308 | 10.07691 | 1.104051 | 0.274342 | 0.762523 | -5.45851 |
| P43487 | RANBP1 | 1.046917 | 11.29012 | 1.105275 | 0.274393 | 0.762523 | -5.19371 |
| Q9UDR5 | AASS | -1.41497 | 10.55739 | -1.10257 | 0.27498 | 0.762523 | -5.46005 |
| Q92954 | PRG4 | -0.94744 | 10.51058 | -1.10233 | 0.275085 | 0.762523 | -5.4603 |
| P40926 | MDH2 | -0.55488 | 13.17949 | -1.10069 | 0.27579 | 0.762523 | -5.46199 |
| P10636-5 | #N/D | -0.49123 | 11.53737 | -1.09803 | 0.27694 | 0.762523 | -5.46474 |
| Q9NR46 | SH3GLB2 | -0.81166 | 8.407993 | -1.09839 | 0.277054 | 0.762523 | -5.34013 |

| | | | | | | | |
|---|---|---|---|---|---|---|---|
| P31946 | YWHAB | 0.711381 | 9.674481 | 1.093 | 0.279392 | 0.762523 | -5.4697 |
| P23246 | SFPQ | 0.65607 | 12.17656 | 1.092167 | 0.279485 | 0.762523 | -5.47077 |
| P07195 | LDHB | -0.49185 | 13.41831 | -1.09206 | 0.279533 | 0.762523 | -5.47088 |
| Q01813 | PFKP | 0.59738 | 10.6556 | 1.091186 | 0.279913 | 0.762523 | -5.47178 |
| O43761 | SYNGR3 | 1.034676 | 10.57735 | 1.089671 | 0.280574 | 0.762523 | -5.47333 |
| P52758 | RIDA | 0.802395 | 8.569229 | 1.084389 | 0.283155 | 0.762523 | -5.35422 |
| P02511 | CRYAB | -0.51357 | 10.8827 | -1.07915 | 0.285195 | 0.762523 | -5.48405 |
| O43866 | CD5L | 0.530302 | 13.65287 | 1.078452 | 0.285504 | 0.762523 | -5.48476 |
| O43175 | PHGDH | 0.484715 | 13.11601 | 1.077531 | 0.285911 | 0.762523 | -5.4857 |
| P52306-6 | #N/D | -0.81057 | 11.71115 | -1.07319 | 0.287838 | 0.762523 | -5.49009 |
| P10606 | COX5B | -0.90891 | 8.344202 | -1.07018 | 0.289738 | 0.762523 | -5.44107 |
| Q92752 | TNR | -0.49774 | 11.57348 | -1.06809 | 0.290113 | 0.762523 | -5.49523 |
| Q9Y3E1 | HDGFL3 | -1.24905 | 8.28437 | -1.06862 | 0.290138 | 0.762523 | -5.33216 |
| P00568 | AK1 | -0.71185 | 11.14475 | -1.06677 | 0.290703 | 0.762523 | -5.49655 |
| P13667 | PDIA4 | -0.73386 | 11.88336 | -1.0658 | 0.291135 | 0.762523 | -5.49752 |
| P05388 | RPLP0 | -0.62752 | 11.63871 | -1.06454 | 0.291702 | 0.762523 | -5.49879 |
| P04196 | HRG | 1.108563 | 14.14042 | 1.064474 | 0.291731 | 0.762523 | -5.49885 |
| Q16143 | SNCB | -0.64369 | 11.41947 | -1.06425 | 0.291829 | 0.762523 | -5.49907 |
| P02743 | APCS | -1.49526 | 14.22881 | -1.06403 | 0.29193 | 0.762523 | -5.4993 |
| Q9NPH9 | IL26 | -1.35729 | 9.277746 | -1.06323 | 0.292745 | 0.762523 | -5.26228 |
| P15121 | AKR1B1 | -1.18486 | 9.04325 | -1.06266 | 0.293553 | 0.762523 | -5.04384 |
| A0A0C4DH | #N/D | 1.013832 | 10.49513 | 1.057988 | 0.295006 | 0.76326 | -5.45335 |
| P05109 | S100A8 | -1.04693 | 9.04775 | -1.0549 | 0.296308 | 0.76326 | -5.34551 |
| P46821 | MAP1B | -0.62544 | 12.35155 | -1.05033 | 0.298126 | 0.76326 | -5.51293 |
| Q13449 | LSAMP | -0.59546 | 9.843599 | -1.0503 | 0.298139 | 0.76326 | -5.51296 |
| Q969P0 | IGSF8 | 0.630336 | 10.87703 | 1.047761 | 0.299297 | 0.76326 | -5.51546 |
| P01615 | #N/D | 0.805379 | 11.88343 | 1.047844 | 0.300023 | 0.76326 | -5.51473 |
| P13647 | KRT5 | 0.757225 | 13.1342 | 1.044656 | 0.300718 | 0.76326 | -5.51852 |
| Q86YZ3 | HRNR | -1.07746 | 10.14946 | -1.04307 | 0.30189 | 0.76326 | -5.4679 |
| P62753 | RPS6 | -0.7756 | 9.50014 | -1.04208 | 0.3019 | 0.76326 | -5.52105 |
| P53004 | BLVRA | 1.032608 | 7.896539 | 1.038815 | 0.304051 | 0.765838 | -5.25743 |

| | | | | | | | |
|---|---|---|---|---|---|---|---|
| P62873 | GNB1 | 0.638022 | 12.21737 | 1.031187 | 0.306933 | 0.765838 | -5.53169 |
| P01742 | #N/D | 0.817811 | 13.16148 | 1.033737 | 0.306984 | 0.765838 | -5.36498 |
| P42766 | RPL35 | 0.641103 | 9.474201 | 1.030465 | 0.30735 | 0.765838 | -5.48044 |
| Q9Y2J8 | PADI2 | 0.92233 | 10.26827 | 1.029218 | 0.307849 | 0.765838 | -5.5336 |
| P04843 | RPN1 | 0.602629 | 8.447986 | 1.028769 | 0.308313 | 0.765838 | -5.48192 |
| P20671 | H2AC7 | -0.75258 | 8.62645 | -1.02682 | 0.310195 | 0.767564 | -5.48295 |
| Q86VP6 | CAND1 | 0.774739 | 9.469724 | 1.02288 | 0.310809 | 0.767564 | -5.53973 |
| P80108 | GPLD1 | 0.678195 | 9.224428 | 1.020937 | 0.311974 | 0.768212 | -5.54137 |
| O60282 | KIF5C | 0.832397 | 9.337804 | 1.017406 | 0.313634 | 0.770075 | -5.4928 |
| Q14195-2 | #N/D | -0.51639 | 12.44951 | -1.00544 | 0.319056 | 0.781136 | -5.5564 |
| P00338 | LDHA | 0.524731 | 12.44144 | 1.001657 | 0.320863 | 0.783311 | -5.55998 |
| P07477 | PRSS1 | -1.45523 | 13.06236 | -0.99806 | 0.322666 | 0.785461 | -5.5633 |
| Q5T7N2 | L1TD1 | 0.623228 | 8.91276 | 0.995776 | 0.324114 | 0.786151 | -5.46705 |
| P68371 | TUBB4B | 1.002924 | 11.99643 | 0.99348 | 0.324795 | 0.786151 | -5.56767 |
| P36957 | DLST | 1.0433 | 8.614293 | 0.992999 | 0.32609 | 0.786759 | -5.25664 |
| P35080 | PFN2 | 0.731053 | 10.95784 | 0.989305 | 0.326893 | 0.786759 | -5.57151 |
| P20742 | #N/D | 0.692595 | 13.66452 | 0.978921 | 0.331874 | 0.795192 | -5.58122 |
| P00450 | CP | -0.56302 | 14.6821 | -0.97728 | 0.332677 | 0.795192 | -5.58274 |
| P26038 | MSN | 0.941674 | 8.378736 | 0.977468 | 0.333197 | 0.795192 | -5.36136 |
| P01871 | #N/D | -0.64764 | 16.51568 | -0.97042 | 0.336054 | 0.799733 | -5.58905 |
| Q08380 | LGALS3BP | 0.584895 | 14.57471 | 0.967633 | 0.337432 | 0.799733 | -5.5916 |
| Q08722 | CD47 | 0.593669 | 9.746025 | 0.965788 | 0.338346 | 0.799733 | -5.59328 |
| Q9Y4L1 | HYOU1 | 0.611591 | 8.672707 | 0.963755 | 0.339432 | 0.799733 | -5.54274 |
| P06744 | GPI | 0.567537 | 11.87388 | 0.961 | 0.340727 | 0.799733 | -5.59764 |
| P35579 | MYH9 | 0.74204 | 12.21283 | 0.960991 | 0.340731 | 0.799733 | -5.59765 |
| P54652 | HSPA2 | 0.549446 | 13.29815 | 0.955164 | 0.343643 | 0.801149 | -5.60292 |
| P08559-4 | #N/D | -0.64396 | 10.42252 | -0.95315 | 0.344655 | 0.801149 | -5.60474 |
| Q02878 | RPL6 | 0.818135 | 11.19865 | 0.951527 | 0.345469 | 0.801149 | -5.6062 |
| O14967 | CLGN | -0.93951 | 11.39778 | -0.95167 | 0.345992 | 0.801149 | -5.29274 |
| P00739 | HPR | -0.72195 | 13.20664 | -0.9504 | 0.346036 | 0.801149 | -5.60721 |
| O15540 | FABP7 | -0.46061 | 10.93642 | -0.94434 | 0.349096 | 0.806043 | -5.61263 |

| | | | | | | | |
|---|---|---|---|---|---|---|---|
| P80404 | ABAT | -0.6058 | 7.872349 | -0.94067 | 0.35233 | 0.811311 | -5.30132 |
| P62266 | RPS23 | 0.899815 | 9.096047 | 0.935696 | 0.35389 | 0.812708 | -5.34909 |
| P02741 | CRP | -1.13857 | 11.11435 | -0.93015 | 0.3564 | 0.816271 | -5.62513 |
| P62861 | #N/D | 0.766022 | 9.145956 | 0.923704 | 0.359955 | 0.816401 | -5.46478 |
| P13533 | MYH6 | 0.504037 | 10.46117 | 0.920758 | 0.361168 | 0.816401 | -5.63343 |
| P63010 | AP2B1 | 0.683948 | 10.0117 | 0.919085 | 0.362035 | 0.816401 | -5.63488 |
| Q9Y5K8 | ATP6V1D | 0.873767 | 7.792041 | 0.920785 | 0.362495 | 0.816401 | -5.3607 |
| P38606 | ATP6V1A | -0.64475 | 12.55831 | -0.91357 | 0.3649 | 0.816401 | -5.63966 |
| P09936 | UCHL1 | -0.53274 | 12.48925 | -0.91231 | 0.365557 | 0.816401 | -5.64075 |
| Q16798 | ME3 | 0.608252 | 8.140939 | 0.912798 | 0.365693 | 0.816401 | -5.47393 |
| Q13813 | SPTAN1 | -0.33456 | 11.99901 | -0.9114 | 0.366034 | 0.816401 | -5.64154 |
| P55056 | APOC4 | -0.79223 | 9.255911 | -0.91186 | 0.366999 | 0.816401 | -5.47395 |
| P14625 | HSP90B1 | 0.91364 | 13.43176 | 0.907547 | 0.368047 | 0.816401 | -5.64485 |
| P46779 | RPL28 | 0.742178 | 9.65877 | 0.907367 | 0.368213 | 0.816401 | -5.59226 |
| P01876 | #N/D | 0.694342 | 15.78121 | 0.905475 | 0.369134 | 0.816401 | -5.64663 |
| P40939 | HADHA | 0.832285 | 9.753015 | 0.901763 | 0.371157 | 0.816401 | -5.64973 |
| Q6UWR7 | ENPP6 | 0.489162 | 11.19948 | 0.901068 | 0.371452 | 0.816401 | -5.6504 |
| P17600 | SYN1 | -0.68979 | 12.09097 | -0.90086 | 0.37156 | 0.816401 | -5.65057 |
| P35527 | KRT9 | -0.60226 | 15.65986 | -0.89946 | 0.3723 | 0.816401 | -5.65177 |
| P08237-3 | #N/D | -0.53011 | 9.903447 | -0.89844 | 0.373058 | 0.816401 | -5.52488 |
| P29972 | AQP1 | -0.91033 | 8.612235 | -0.89769 | 0.373705 | 0.816401 | -5.38042 |
| Q07020 | RPL18 | 0.85503 | 11.58711 | 0.894039 | 0.375168 | 0.817501 | -5.65637 |
| P12956 | XRCC6 | -0.63763 | 12.11524 | -0.88215 | 0.381504 | 0.826718 | -5.66636 |
| P14618-2 | #N/D | -0.72845 | 11.99726 | -0.88133 | 0.381944 | 0.826718 | -5.66705 |
| P01031 | C5 | -0.33919 | 12.98874 | -0.88065 | 0.382308 | 0.826718 | -5.66762 |
| P01861 | #N/D | 0.531893 | 9.667126 | 0.876937 | 0.384523 | 0.828558 | -5.61764 |
| P43652 | AFM | -0.42898 | 12.25694 | -0.87279 | 0.386545 | 0.828558 | -5.67415 |
| P62913 | RPL11 | 0.607519 | 12.51404 | 0.870396 | 0.387839 | 0.828558 | -5.67613 |
| P02654 | APOC1 | 0.59432 | 11.84732 | 0.868979 | 0.388675 | 0.828558 | -5.62432 |
| P20700 | LMNB1 | -0.66646 | 10.71505 | -0.86849 | 0.388872 | 0.828558 | -5.6777 |
| Q92804 | TAF15 | -0.6282 | 9.172634 | -0.86898 | 0.389147 | 0.828558 | -5.67674 |

| | | | | | | | |
|---|---|---|---|---|---|---|---|
| P22234 | PAICS | -0.51697 | 10.08226 | -0.86546 | 0.390515 | 0.828558 | -5.68018 |
| A0A0C4DH | #N/D | 0.937739 | 10.33687 | 0.860817 | 0.393116 | 0.828558 | -5.68391 |
| P30041 | PRDX6 | 0.813932 | 12.92684 | 0.859712 | 0.393652 | 0.828558 | -5.68488 |
| P0DP25 | CALM1 | -1.17116 | 11.85239 | -0.85866 | 0.394229 | 0.828558 | -5.68574 |
| P60900 | PSMA6 | 0.844916 | 10.92948 | 0.856158 | 0.395598 | 0.828558 | -5.68777 |
| P04406 | GAPDH | 0.589023 | 14.71391 | 0.855899 | 0.395739 | 0.828558 | -5.68798 |
| A0A0B4J1\ | #N/D | -0.60928 | 12.65679 | -0.85579 | 0.395802 | 0.828558 | -5.68808 |
| P26639 | TARS1 | 0.537787 | 9.377104 | 0.84843 | 0.399915 | 0.832174 | -5.64093 |
| P00748 | F12 | -0.4203 | 11.60206 | -0.84811 | 0.400027 | 0.832174 | -5.69428 |
| O60641 | SNAP91 | -0.47187 | 12.93002 | -0.84732 | 0.400459 | 0.832174 | -5.69491 |
| P47914 | RPL29 | 0.701673 | 8.941895 | 0.843476 | 0.402869 | 0.835146 | -5.59777 |
| P05455 | SSB | 0.734765 | 10.60567 | 0.838025 | 0.405683 | 0.838936 | -5.6492 |
| P31943 | HNRNPH1 | -0.52155 | 12.25814 | -0.83569 | 0.40692 | 0.839458 | -5.70419 |
| Q06830 | PRDX1 | 0.439859 | 12.88336 | 0.832729 | 0.408572 | 0.84083 | -5.70653 |
| Q14624 | ITIH4 | -0.74366 | 9.94507 | -0.83032 | 0.410272 | 0.842294 | -5.70807 |
| Q04917 | YWHAH | 0.604749 | 11.45903 | 0.825 | 0.412909 | 0.845669 | -5.71261 |
| P01764 | #N/D | 0.702553 | 9.51328 | 0.818359 | 0.417081 | 0.850911 | -5.54938 |
| Q13228 | SELENBP1 | 0.432943 | 10.91175 | 0.814294 | 0.418962 | 0.850911 | -5.72094 |
| P01008 | SERPINC1 | -0.42022 | 14.22003 | -0.81353 | 0.419397 | 0.850911 | -5.72153 |
| P09972 | ALDOC | 0.42834 | 13.29965 | 0.813412 | 0.419463 | 0.850911 | -5.72162 |
| P06310 | #N/D | 0.542947 | 13.49283 | 0.807587 | 0.422782 | 0.853985 | -5.7261 |
| P50993 | ATP1A2 | -0.69027 | 10.45463 | -0.80585 | 0.423839 | 0.853985 | -5.72737 |
| Q99832 | CCT7 | -0.42568 | 11.03507 | -0.80548 | 0.423985 | 0.853985 | -5.72772 |
| P69905 | HBA1 | 0.403837 | 14.01226 | 0.799334 | 0.42751 | 0.859053 | -5.7324 |
| P50395 | GDI2 | 0.424027 | 11.20433 | 0.797573 | 0.428523 | 0.859063 | -5.73374 |
| Q5IFJ7 | #N/D | 1.030276 | 10.61788 | 0.795798 | 0.429607 | 0.859087 | -5.68171 |
| P28161 | GSTM2 | 0.88439 | 8.502551 | 0.795504 | 0.430552 | 0.859087 | -5.25315 |
| A0A075B6I | #N/D | 0.705948 | 12.27284 | 0.790553 | 0.432575 | 0.860203 | -5.73903 |
| P22087 | FBL | -0.54099 | 8.6313 | -0.78515 | 0.436112 | 0.860203 | -5.57404 |
| O75363-2 | #N/D | 0.56909 | 8.935009 | 0.777213 | 0.440399 | 0.860203 | -5.7489 |
| P07357 | C8A | -0.43551 | 14.30403 | -0.77041 | 0.444331 | 0.860203 | -5.75397 |

| | | | | | | | |
|---|---|---|---|---|---|---|---|
| P49419 | ALDH7A1 | 0.539084 | 13.146 | 0.770132 | 0.444492 | 0.860203 | -5.75417 |
| P04350 | TUBB4A | 0.527521 | 11.70512 | 0.769426 | 0.444908 | 0.860203 | -5.75469 |
| P61970 | NUTF2 | -0.56924 | 8.594163 | -0.76926 | 0.445063 | 0.860203 | -5.75474 |
| P10809 | HSPD1 | -0.3407 | 14.28311 | -0.76686 | 0.446421 | 0.860203 | -5.75656 |
| Q7Z3B1 | NEGR1 | 0.991122 | 7.928181 | 0.764607 | 0.448739 | 0.860203 | -5.27345 |
| P46777 | RPL5 | -0.64045 | 8.455774 | -0.76322 | 0.449277 | 0.860203 | -5.58946 |
| P21579 | SYT1 | 0.529698 | 9.733547 | 0.760515 | 0.450171 | 0.860203 | -5.76117 |
| O75368 | SH3BGRL | 0.822357 | 9.591212 | 0.759104 | 0.451128 | 0.860203 | -5.6325 |
| O94919 | ENDOD1 | -0.5292 | 9.449366 | -0.75451 | 0.453859 | 0.860203 | -5.71185 |
| O00410 | IPO5 | 0.607399 | 9.76488 | 0.752537 | 0.454913 | 0.860203 | -5.76691 |
| P08697 | SERPINF2 | -0.34227 | 12.89207 | -0.75098 | 0.455841 | 0.860203 | -5.76802 |
| P51178-2 | #N/D | 0.74417 | 9.240432 | 0.749327 | 0.457142 | 0.860203 | -5.5996 |
| P45974 | USP5 | -0.43179 | 9.969402 | -0.74659 | 0.458466 | 0.860203 | -5.77114 |
| P46459 | NSF | 0.543026 | 12.65607 | 0.745195 | 0.459303 | 0.860203 | -5.77213 |
| A0A0J9YXX | #N/D | -0.57401 | 10.65969 | -0.74399 | 0.460024 | 0.860203 | -5.77299 |
| P01019 | AGT | 0.609327 | 13.11805 | 0.743691 | 0.460205 | 0.860203 | -5.7732 |
| P17987 | TCP1 | -0.62416 | 10.0471 | -0.74179 | 0.461348 | 0.860203 | -5.77454 |
| A0MZ66 | SHTN1 | 0.715884 | 8.930068 | 0.74182 | 0.46178 | 0.860203 | -5.60465 |
| P19652 | ORM2 | -0.46244 | 9.423031 | -0.7413 | 0.461951 | 0.860203 | -5.64478 |
| Q12931 | TRAP1 | 0.653579 | 11.45079 | 0.739447 | 0.462757 | 0.860203 | -5.77619 |
| Q9NQC3 | RTN4 | 0.436969 | 10.95603 | 0.739224 | 0.462891 | 0.860203 | -5.77635 |
| Q92598-4 | #N/D | 1.035656 | 8.188924 | 0.74101 | 0.463079 | 0.860203 | -5.28828 |
| P62750 | RPL23A | -0.45522 | 11.64551 | -0.73854 | 0.463305 | 0.860203 | -5.77683 |
| Q13363 | CTBP1 | 0.563383 | 9.960849 | 0.737089 | 0.464178 | 0.860203 | -5.77785 |
| P50453 | SERPINB9 | 0.784283 | 9.530376 | 0.736638 | 0.464825 | 0.860203 | -5.60828 |
| Q9Y266 | NUDC | -0.99728 | 9.322208 | -0.7349 | 0.465872 | 0.860203 | -5.29291 |
| P0DJI8 | SAA1 | -0.57295 | 12.87564 | -0.73327 | 0.466486 | 0.860203 | -5.78052 |
| P00736 | C1R | -0.57325 | 13.74472 | -0.73206 | 0.467214 | 0.860203 | -5.78136 |
| P53634 | CTSC | -0.61353 | 8.483897 | -0.73233 | 0.467496 | 0.860203 | -5.61114 |
| P13611 | VCAN | -0.27573 | 12.41909 | -0.72967 | 0.468665 | 0.860203 | -5.78303 |
| P14174 | MIF | 0.636577 | 11.07338 | 0.728818 | 0.469182 | 0.860203 | -5.78362 |

| | | | | | | | |
|---|---|---|---|---|---|---|---|
| P0DJI9 | SAA2 | 0.825487 | 8.735337 | 0.727515 | 0.470488 | 0.860203 | -5.50541 |
| P17174 | GOT1 | 0.492866 | 13.06163 | 0.726467 | 0.47061 | 0.860203 | -5.78524 |
| Q9UHG2 | PCSK1N | 0.934888 | 7.955461 | 0.72729 | 0.47114 | 0.860203 | -5.22816 |
| P60953 | CDC42 | 0.642555 | 9.514288 | 0.7243 | 0.472164 | 0.860203 | -5.68533 |
| Q08209-3 | #N/D | 0.453231 | 10.11222 | 0.723862 | 0.472251 | 0.860203 | -5.73332 |
| P51674 | GPM6A | 0.570437 | 9.977344 | 0.720274 | 0.474384 | 0.860203 | -5.78951 |
| Q15485 | FCN2 | 0.712703 | 12.63067 | 0.717264 | 0.476224 | 0.860203 | -5.79157 |
| Q12860 | CNTN1 | 0.472538 | 11.73525 | 0.717202 | 0.476262 | 0.860203 | -5.79161 |
| P02753 | RBP4 | -0.73997 | 9.614475 | -0.71607 | 0.477191 | 0.860203 | -5.6909 |
| Q02252 | ALDH6A1 | -0.9668 | 13.39322 | -0.71472 | 0.477781 | 0.860203 | -5.7933 |
| P09651 | HNRNPA1 | -0.46703 | 12.21855 | -0.71401 | 0.478217 | 0.860203 | -5.79378 |
| P40429 | RPL13A | 0.445807 | 11.00395 | 0.712762 | 0.478984 | 0.860203 | -5.79463 |
| Q15121 | PEA15 | -0.34571 | 10.46576 | -0.71151 | 0.479755 | 0.860203 | -5.79548 |
| P60981 | DSTN | 0.50419 | 11.21887 | 0.709865 | 0.480819 | 0.860203 | -5.79654 |
| Q01082 | SPTBN1 | 0.278356 | 11.78071 | 0.70852 | 0.481593 | 0.860203 | -5.7975 |
| P02750 | LRG1 | -0.59138 | 10.4896 | -0.70428 | 0.484207 | 0.861714 | -5.80035 |
| P16403 | H1-2 | 0.302067 | 11.57771 | 0.703869 | 0.484461 | 0.861714 | -5.80063 |
| Q5TFQ8 | SIRPB1 | 0.675493 | 8.396877 | 0.702637 | 0.486411 | 0.863353 | -5.47622 |
| P02748 | C9 | 0.569715 | 13.54131 | 0.698473 | 0.487802 | 0.863353 | -5.80423 |
| P09543 | CNP | 0.292154 | 12.18182 | 0.697474 | 0.488423 | 0.863353 | -5.8049 |
| P06454 | PTMA | 0.515657 | 9.310123 | 0.691079 | 0.492401 | 0.867689 | -5.80912 |
| O75874 | IDH1 | 0.646492 | 10.43067 | 0.689738 | 0.493238 | 0.867689 | -5.81001 |
| P02768 | ALB | -0.59503 | 17.61783 | -0.68802 | 0.494311 | 0.867689 | -5.81113 |
| P07451 | CA3 | -0.63411 | 9.536455 | -0.68564 | 0.495848 | 0.867689 | -5.75879 |
| P68104 | EEF1A1 | -0.50966 | 9.086982 | -0.68433 | 0.496674 | 0.867689 | -5.75964 |
| Q12765 | SCRN1 | -0.28748 | 11.08891 | -0.68374 | 0.496986 | 0.867689 | -5.81393 |
| P09012 | SNRPA | -0.83791 | 9.436966 | -0.67482 | 0.503286 | 0.87689 | -5.4939 |
| P48735 | IDH2 | -0.44423 | 11.01398 | -0.67067 | 0.505218 | 0.877429 | -5.82237 |
| P80748 | #N/D | -0.47284 | 13.59399 | -0.66318 | 0.509963 | 0.877429 | -5.82712 |
| P49327 | FASN | -0.35658 | 12.45025 | -0.662 | 0.510715 | 0.877429 | -5.82787 |
| Q16629 | SRSF7 | -0.94266 | 9.577583 | -0.66193 | 0.510969 | 0.877429 | -5.3525 |

| | | | | | | | |
|---|---|---|---|---|---|---|---|
| P0DOX3 | #N/D | -0.28741 | 13.40279 | -0.65989 | 0.51206 | 0.877429 | -5.8292 |
| P09874 | PARP1 | 0.296152 | 12.84743 | 0.659249 | 0.512466 | 0.877429 | -5.8296 |
| P08865 | RPSA | 0.405167 | 13.49287 | 0.657931 | 0.513306 | 0.877429 | -5.83043 |
| A0A075B6I | #N/D | 0.721185 | 11.3767 | 0.656562 | 0.514335 | 0.877429 | -5.77715 |
| P50502 | ST13 | -0.53898 | 12.29466 | -0.65574 | 0.514857 | 0.877429 | -5.83164 |
| P62888 | RPL30 | 0.451449 | 9.168351 | 0.655482 | 0.51492 | 0.877429 | -5.83191 |
| O00499 | BIN1 | -0.58716 | 8.599564 | -0.65536 | 0.515952 | 0.877429 | -5.50524 |
| P61266-2 | #N/D | -0.30435 | 9.516859 | -0.65379 | 0.515953 | 0.877429 | -5.83302 |
| Q86V81 | ALYREF | -0.66598 | 8.628483 | -0.65232 | 0.517156 | 0.877723 | -5.73184 |
| P38646 | HSPA9 | -0.30829 | 14.21459 | -0.64923 | 0.518869 | 0.878627 | -5.83585 |
| P39023 | RPL3 | -0.81037 | 11.33349 | -0.6464 | 0.520688 | 0.878627 | -5.8376 |
| Q99447 | PCYT2 | 0.589296 | 9.931576 | 0.645732 | 0.521118 | 0.878627 | -5.83801 |
| Q9BW30 | TPPP3 | 0.563879 | 7.409834 | 0.647094 | 0.521864 | 0.878627 | -5.27311 |
| A0A0B4J1Y | #N/D | -0.46504 | 11.1449 | -0.64305 | 0.522845 | 0.878627 | -5.83966 |
| P49411 | TUFM | 0.533559 | 11.6554 | 0.640901 | 0.524229 | 0.879218 | -5.84098 |
| P13489 | RNH1 | -0.56223 | 9.869775 | -0.63897 | 0.525477 | 0.87958 | -5.84216 |
| P62277 | RPS13 | -0.38437 | 10.17611 | -0.63498 | 0.528054 | 0.882161 | -5.84458 |
| P04433 | #N/D | -0.55123 | 12.95042 | -0.62833 | 0.532366 | 0.887624 | -5.84858 |
| P29218 | IMPA1 | -0.37407 | 11.50747 | -0.62571 | 0.534076 | 0.888736 | -5.85015 |
| Q13509 | TUBB3 | 0.337849 | 9.79097 | 0.62217 | 0.536429 | 0.890215 | -5.85221 |
| A0A075B6I | #N/D | -0.40197 | 9.618077 | -0.62114 | 0.537054 | 0.890215 | -5.85287 |
| P62917 | RPL8 | 0.661891 | 11.02457 | 0.618163 | 0.539047 | 0.891097 | -5.85458 |
| P36542 | ATP5F1C | -0.56034 | 8.889386 | -0.6165 | 0.540134 | 0.891097 | -5.85556 |
| Q99497 | PARK7 | -0.51844 | 9.50833 | -0.61446 | 0.541623 | 0.891097 | -5.80246 |
| P02686 | MBP | 0.254077 | 12.52744 | 0.613123 | 0.542303 | 0.891097 | -5.85759 |
| P28482 | MAPK1 | -0.31017 | 10.57932 | -0.61241 | 0.542816 | 0.891097 | -5.8038 |
| P35998 | PSMC2 | 0.571967 | 10.28438 | 0.609102 | 0.545088 | 0.89249 | -5.85979 |
| P34897 | SHMT2 | -0.40772 | 10.26867 | -0.60717 | 0.54622 | 0.89249 | -5.86106 |
| P99999 | CYCS | -0.80627 | 7.341751 | -0.60555 | 0.549303 | 0.89249 | -5.18865 |
| Q16653-3 | #N/D | -0.41563 | 9.748252 | -0.60126 | 0.550119 | 0.89249 | -5.86447 |
| P20073 | ANXA7 | -0.35435 | 8.552518 | -0.60104 | 0.550309 | 0.89249 | -5.81034 |

| | | | | | | | |
|---|---|---|---|---|---|---|---|
| O95445 | APOM | -0.64748 | 11.61754 | -0.59991 | 0.551013 | 0.89249 | -5.86524 |
| P49588 | AARS1 | -0.89333 | 10.01352 | -0.59898 | 0.55187 | 0.89249 | -5.73407 |
| P46783 | RPS10 | 0.498047 | 10.88473 | 0.598098 | 0.552212 | 0.89249 | -5.86628 |
| Q9UHD8 | SEPTIN9 | 0.431075 | 7.179257 | 0.598172 | 0.553532 | 0.89249 | -5.53714 |
| O14791 | APOL1 | -0.3969 | 13.07569 | -0.59519 | 0.55414 | 0.89249 | -5.86793 |
| P11279 | LAMP1 | -0.50338 | 9.296891 | -0.59168 | 0.556709 | 0.892609 | -5.61717 |
| P55084 | HADHB | -0.41005 | 8.238028 | -0.59134 | 0.557163 | 0.892609 | -5.73811 |
| P00918 | CA2 | 0.358315 | 9.133649 | 0.589014 | 0.558335 | 0.892609 | -5.87134 |
| P08779 | KRT16 | -0.97866 | 8.523877 | -0.59097 | 0.558404 | 0.892609 | -5.30199 |
| P60842 | EIF4A1 | 0.534753 | 11.09096 | 0.584145 | 0.561493 | 0.894378 | -5.87415 |
| A0A0B4J1U | #N/D | 0.522563 | 9.305291 | 0.580434 | 0.56437 | 0.894378 | -5.59294 |
| P18077 | RPL35A | -0.59497 | 11.39346 | -0.57987 | 0.564444 | 0.894378 | -5.87644 |
| P07737 | PFN1 | 0.638165 | 12.48645 | 0.576034 | 0.567155 | 0.894378 | -5.74672 |
| O43390 | HNRNPR | -0.36032 | 8.163912 | -0.57547 | 0.567533 | 0.894378 | -5.77628 |
| P05091 | ALDH2 | 0.34089 | 12.45982 | 0.574449 | 0.567989 | 0.894378 | -5.87952 |
| P50454 | SERPINH1 | -0.36341 | 9.103501 | -0.57383 | 0.568445 | 0.894378 | -5.87981 |
| P16949 | STMN1 | -0.41887 | 11.41839 | -0.57221 | 0.569497 | 0.894378 | -5.88075 |
| P13798 | APEH | 0.408185 | 11.77021 | 0.571619 | 0.569891 | 0.894378 | -5.88107 |
| P02787 | TF | 0.317651 | 14.24336 | 0.571322 | 0.570091 | 0.894378 | -5.88123 |
| O76021 | RSL1D1 | -0.4606 | 9.23965 | -0.5675 | 0.572795 | 0.894378 | -5.82884 |
| P49591 | SARS1 | 0.61521 | 8.044302 | 0.566838 | 0.573616 | 0.894378 | -5.38597 |
| Q02218 | OGDH | 0.635291 | 8.055839 | 0.566513 | 0.573897 | 0.894378 | -5.3604 |
| Q96AX9-5 | #N/D | -0.50272 | 9.366756 | -0.56524 | 0.574235 | 0.894378 | -5.88449 |
| P14136 | GFAP | -0.27667 | 13.25535 | -0.56366 | 0.575257 | 0.894378 | -5.88538 |
| Q8IXJ6 | SIRT2 | -0.63543 | 11.31293 | -0.56029 | 0.577666 | 0.896487 | -5.7552 |
| P13716-2 | #N/D | 0.508695 | 8.80277 | 0.557836 | 0.579377 | 0.896653 | -5.71606 |
| P62273 | RPS29 | -0.40378 | 8.012306 | -0.55545 | 0.581196 | 0.896653 | -5.83502 |
| Q99729-3 | #N/D | -0.59508 | 12.65612 | -0.55284 | 0.582593 | 0.896653 | -5.89115 |
| A0A0C4DH | #N/D | -0.41388 | 12.36129 | -0.55016 | 0.584418 | 0.896653 | -5.89256 |
| P51148 | RAB5C | -0.57762 | 8.07766 | -0.55104 | 0.584497 | 0.896653 | -5.36761 |
| P04275 | VWF | 0.408793 | 14.63316 | 0.546361 | 0.587011 | 0.896653 | -5.89456 |

| | | | | | | | |
|---|---|---|---|---|---|---|---|
| P48637 | GSS | 0.618704 | 8.001266 | 0.547236 | 0.587302 | 0.896653 | -5.56389 |
| P48147 | PREP | 0.650379 | 7.680943 | 0.546893 | 0.587318 | 0.896653 | -5.36954 |
| P12532 | CKMT1B | 0.367652 | 9.736519 | 0.544626 | 0.588196 | 0.896653 | -5.89546 |
| P08603 | CFH | 0.311098 | 14.2247 | 0.544478 | 0.588297 | 0.896653 | -5.89554 |
| P36871 | PGM1 | -0.30492 | 11.50669 | -0.53673 | 0.593606 | 0.898751 | -5.89954 |
| P60028 | #N/D | -0.38939 | 9.154595 | -0.53488 | 0.594998 | 0.898751 | -5.84591 |
| Q15084 | PDIA6 | 0.315767 | 12.47948 | 0.534015 | 0.595467 | 0.898751 | -5.90093 |
| Q8N163 | CCAR2 | -0.40417 | 8.405871 | -0.53378 | 0.595752 | 0.898751 | -5.84647 |
| Q13151 | HNRNPA0 | 0.390716 | 9.990869 | 0.533055 | 0.596249 | 0.898751 | -5.84684 |
| P26641 | EEF1G | -0.30044 | 10.13888 | -0.53154 | 0.597207 | 0.898751 | -5.90215 |
| P14618-3 | #N/D | -0.37297 | 9.955675 | -0.53109 | 0.597518 | 0.898751 | -5.84791 |
| P14314 | PRKCSH | 0.413247 | 12.48603 | 0.530036 | 0.598325 | 0.898751 | -5.90284 |
| P02671 | FGA | -0.25929 | 12.54848 | -0.52864 | 0.599167 | 0.898751 | -5.90366 |
| P10909 | CLU | -0.20376 | 13.71219 | -0.51581 | 0.608041 | 0.908458 | -5.91007 |
| P01766 | #N/D | 0.46371 | 9.036312 | 0.515175 | 0.608728 | 0.908458 | -5.656 |
| P27348 | YWHAQ | 0.400248 | 11.63346 | 0.513013 | 0.610018 | 0.908458 | -5.91141 |
| P27169 | PON1 | 0.463941 | 14.83311 | 0.511911 | 0.610785 | 0.908458 | -5.91195 |
| P62851 | RPS25 | 0.366837 | 10.64803 | 0.507436 | 0.613862 | 0.908458 | -5.91417 |
| P10768 | ESD | 0.367268 | 10.75161 | 0.506712 | 0.614367 | 0.908458 | -5.91452 |
| P02774-3 | #N/D | -0.39811 | 14.29976 | -0.506 | 0.614863 | 0.908458 | -5.91486 |
| P01700 | #N/D | -0.51736 | 10.59788 | -0.50527 | 0.615408 | 0.908458 | -5.91518 |
| Q04837 | SSBP1 | 0.429267 | 9.844274 | 0.502583 | 0.617285 | 0.908458 | -5.91648 |
| P14415 | ATP1B2 | -0.3489 | 8.426782 | -0.50276 | 0.61732 | 0.908458 | -5.86167 |
| P05160 | F13B | 0.414101 | 11.66103 | 0.50252 | 0.617368 | 0.908458 | -5.91647 |
| P05090 | APOD | 0.28138 | 14.6869 | 0.499701 | 0.619264 | 0.909678 | -5.91789 |
| Q00839 | HNRNPU | 0.261268 | 12.5168 | 0.496911 | 0.621218 | 0.910078 | -5.91922 |
| P63241 | EIF5A | 0.39493 | 8.213411 | 0.496665 | 0.621673 | 0.910078 | -5.74603 |
| P50914 | RPL14 | 0.403607 | 9.06483 | 0.488831 | 0.62704 | 0.916023 | -5.79038 |
| P04080 | CSTB | 0.393233 | 11.10002 | 0.486847 | 0.628287 | 0.916023 | -5.92396 |
| P35520 | CBS | 0.441932 | 8.684517 | 0.485302 | 0.629564 | 0.916023 | -5.63934 |
| P35542 | SAA4 | 0.249391 | 12.48858 | 0.484367 | 0.630035 | 0.916023 | -5.92511 |

| | | | | | | | |
|---|---|---|---|---|---|---|---|
| P00505 | GOT2 | -0.25648 | 12.49377 | -0.48278 | 0.631153 | 0.916086 | -5.92584 |
| P36354 | #N/D | 0.569064 | 8.62113 | 0.476556 | 0.636069 | 0.917235 | -5.59688 |
| P13591 | NCAM1 | 0.31678 | 12.6968 | 0.472245 | 0.638606 | 0.917235 | -5.93066 |
| Q03591 | CFHR1 | 0.37552 | 9.2033 | 0.470587 | 0.640006 | 0.917235 | -5.6979 |
| Q16181 | SEPTIN7 | -0.60908 | 11.66741 | -0.47003 | 0.640177 | 0.917235 | -5.93166 |
| P06703 | S100A6 | 0.536953 | 9.228704 | 0.467433 | 0.642023 | 0.917235 | -5.93282 |
| Q9UPY8 | MAPRE3 | -0.21868 | 11.03301 | -0.46452 | 0.644097 | 0.917235 | -5.93412 |
| Q15848 | ADIPOQ | 0.322773 | 10.82374 | 0.464217 | 0.644311 | 0.917235 | -5.93426 |
| P14649 | MYL6B | 0.250054 | 11.20495 | 0.463241 | 0.645006 | 0.917235 | -5.93469 |
| P20042 | EIF2S2 | 0.472474 | 8.98351 | 0.462419 | 0.645625 | 0.917235 | -5.88033 |
| Q9Y2J2 | EPB41L3 | 0.548456 | 8.157606 | 0.462085 | 0.646328 | 0.917235 | -5.43206 |
| Q9H3S7 | PTPN23 | 0.259847 | 9.197781 | 0.461188 | 0.646469 | 0.917235 | -5.9356 |
| P28070 | PSMB4 | -0.33394 | 9.16426 | -0.46017 | 0.647259 | 0.917235 | -5.88128 |
| Q07954 | LRP1 | 0.402868 | 9.068573 | 0.460199 | 0.647277 | 0.917235 | -5.76248 |
| Q15907 | RAB11B | -0.31139 | 9.911453 | -0.45995 | 0.647383 | 0.917235 | -5.88141 |
| P02652 | APOA2 | -0.29263 | 13.89358 | -0.45891 | 0.648093 | 0.917235 | -5.9366 |
| Q14194 | CRMP1 | 0.229979 | 11.68612 | 0.45068 | 0.653979 | 0.92001 | -5.94019 |
| Q13938 | #N/D | 0.251689 | 11.20708 | 0.448184 | 0.655768 | 0.92001 | -5.94126 |
| P48506 | GCLC | -0.32178 | 8.866158 | -0.44564 | 0.657591 | 0.92001 | -5.94235 |
| P09493-4 | #N/D | -0.29817 | 12.6635 | -0.44407 | 0.658721 | 0.92001 | -5.94302 |
| O14531 | DPYSL4 | 0.484842 | 8.30417 | 0.442586 | 0.660314 | 0.92001 | -5.43959 |
| P23142 | FBLN1 | 0.284725 | 9.472544 | 0.439463 | 0.662167 | 0.92001 | -5.81193 |
| P01023 | A2M | 0.316532 | 15.60344 | 0.438446 | 0.662767 | 0.92001 | -5.94539 |
| A0A0G2JS( | #N/D | -0.53802 | 9.269116 | -0.43754 | 0.663796 | 0.92001 | -5.77175 |
| P60866 | RPS20 | 0.335463 | 8.421034 | 0.433435 | 0.666548 | 0.92001 | -5.84395 |
| Q6TUY0 | #N/D | -0.68968 | 12.02649 | -0.43163 | 0.66775 | 0.92001 | -5.89339 |
| P84103 | SRSF3 | -0.43226 | 7.821272 | -0.43102 | 0.668541 | 0.92001 | -5.44401 |
| P13639 | EEF2 | 0.236982 | 12.11228 | 0.427507 | 0.670667 | 0.92001 | -5.94993 |
| Q9HC38 | GLOD4 | 0.314345 | 9.301142 | 0.427466 | 0.670697 | 0.92001 | -5.94995 |
| Q92686 | NRGN | -0.37284 | 9.047773 | -0.42678 | 0.671289 | 0.92001 | -5.89534 |
| O43776 | NARS1 | 0.260306 | 10.11421 | 0.42629 | 0.671548 | 0.92001 | -5.95043 |

| | | | | | | | |
|---|---|---|---|---|---|---|---|
| Q99623 | PHB2 | -0.39523 | 10.71072 | -0.42372 | 0.67341 | 0.92001 | -5.95147 |
| P69891 | HBG1 | 0.336284 | 10.30411 | 0.421661 | 0.674933 | 0.92001 | -5.95228 |
| P27635 | RPL10 | 0.22643 | 8.546315 | 0.421656 | 0.674937 | 0.92001 | -5.95228 |
| O76054 | SEC14L2 | -0.403 | 8.396433 | -0.42174 | 0.675294 | 0.92001 | -5.61919 |
| O14818 | PSMA7 | -0.27004 | 8.736898 | -0.42023 | 0.675969 | 0.92001 | -5.89803 |
| A0A0C4DH | #N/D | -0.49099 | 9.855679 | -0.41825 | 0.677576 | 0.92001 | -5.66699 |
| P10620 | MGST1 | 0.321333 | 10.88042 | 0.416074 | 0.678963 | 0.92001 | -5.95455 |
| P30044 | PRDX5 | 0.487571 | 11.42963 | 0.413772 | 0.680638 | 0.92001 | -5.95546 |
| Q9NRV9 | HEBP1 | 0.440843 | 10.05472 | 0.4129 | 0.681466 | 0.92001 | -5.78169 |
| Q92823 | NRCAM | -0.20982 | 13.18446 | -0.41203 | 0.681905 | 0.92001 | -5.95615 |
| P01859 | #N/D | 0.32816 | 13.5427 | 0.410866 | 0.682755 | 0.92001 | -5.95661 |
| P61247 | RPS3A | 0.179016 | 12.29605 | 0.410154 | 0.683274 | 0.92001 | -5.95689 |
| P35613-2 | #N/D | 0.216049 | 10.74277 | 0.407198 | 0.685432 | 0.92001 | -5.95805 |
| P37840 | SNCA | 0.334661 | 9.699817 | 0.40712 | 0.685547 | 0.92001 | -5.95803 |
| P23528 | CFL1 | -0.24126 | 12.52033 | -0.40409 | 0.6877 | 0.92001 | -5.95926 |
| Q8B6J5 | #N/D | 0.41316 | 11.86572 | 0.403237 | 0.688666 | 0.92001 | -5.95931 |
| O95336 | PGLS | 0.322877 | 10.2894 | 0.402168 | 0.689108 | 0.92001 | -5.96 |
| P43243 | MATR3 | -0.56844 | 7.788896 | -0.40308 | 0.689247 | 0.92001 | -5.42734 |
| P60709 | ACTB | 0.240043 | 12.29443 | 0.401427 | 0.68965 | 0.92001 | -5.96029 |
| O00425 | IGF2BP3 | -0.39232 | 8.291206 | -0.402 | 0.689706 | 0.92001 | -5.62653 |
| A5YM72 | CARNS1 | -0.4247 | 9.654885 | -0.40024 | 0.690779 | 0.92001 | -5.72617 |
| Q14103 | HNRNPD | -0.41301 | 10.83965 | -0.39887 | 0.691523 | 0.92001 | -5.96127 |
| A0A0C4DH | #N/D | -0.29073 | 9.696796 | -0.39792 | 0.692216 | 0.92001 | -5.96163 |
| P0C0L5 | C4B | 0.229547 | 12.58969 | 0.396444 | 0.693328 | 0.92001 | -5.96217 |
| P30050 | RPL12 | 0.33776 | 10.32925 | 0.394047 | 0.695059 | 0.92001 | -5.9631 |
| P23396 | RPS3 | -0.26932 | 12.74094 | -0.39246 | 0.69622 | 0.92001 | -5.96369 |
| P00740 | F9 | 0.227343 | 11.14519 | 0.391763 | 0.696736 | 0.92001 | -5.96396 |
| O95197 | RTN3 | 0.588635 | 8.031271 | 0.391756 | 0.697404 | 0.92001 | -5.38389 |
| Q15631 | TSN | -0.39637 | 8.819731 | -0.39061 | 0.69767 | 0.92001 | -5.79015 |
| O95394 | PGM3 | -0.29162 | 8.458672 | -0.38907 | 0.698742 | 0.92001 | -5.96494 |
| P49773 | HINT1 | -0.27685 | 8.488339 | -0.38781 | 0.699726 | 0.92001 | -5.91046 |

| | | | | | | | |
|---|---|---|---|---|---|---|---|
| P13010 | XRCC5 | -0.20633 | 12.2267 | -0.3826 | 0.703479 | 0.920626 | -5.96736 |
| P01011 | SERPINA3 | 0.155678 | 14.45594 | 0.382145 | 0.703816 | 0.920626 | -5.96753 |
| P51884 | LUM | 0.283356 | 8.535854 | 0.382176 | 0.704076 | 0.920626 | -5.83396 |
| P68133 | ACTA1 | -0.3463 | 8.536068 | -0.3818 | 0.704516 | 0.920626 | -5.46114 |
| P09960 | LTA4H | 0.321358 | 8.842087 | 0.379845 | 0.705721 | 0.920788 | -5.63456 |
| P00492 | HPRT1 | 0.334132 | 10.76362 | 0.376453 | 0.708044 | 0.92169 | -5.96957 |
| P00738 | HP | 0.190078 | 16.23776 | 0.37358 | 0.710142 | 0.92169 | -5.97063 |
| P52272 | HNRNPM | -0.21043 | 12.38158 | -0.37114 | 0.711951 | 0.92169 | -5.9715 |
| P06730-2 | #N/D | 0.526818 | 8.849688 | 0.366431 | 0.715673 | 0.92169 | -5.46621 |
| P05156 | CFI | -0.24865 | 13.67799 | -0.36401 | 0.717234 | 0.92169 | -5.97401 |
| Q99962 | SH3GL2 | -0.18523 | 12.54019 | -0.36356 | 0.717566 | 0.92169 | -5.97417 |
| P25705 | ATP5F1A | 0.363394 | 13.38486 | 0.363431 | 0.717666 | 0.92169 | -5.97421 |
| P09382 | LGALS1 | 0.263074 | 9.996774 | 0.360856 | 0.719579 | 0.92169 | -5.97511 |
| P05155 | SERPING1 | 0.234308 | 12.77219 | 0.360665 | 0.719721 | 0.92169 | -5.97517 |
| P35232 | PHB | -0.17992 | 11.88214 | -0.36064 | 0.719737 | 0.92169 | -5.97518 |
| Q15392 | DHCR24 | -0.36088 | 8.63106 | -0.35995 | 0.720392 | 0.92169 | -5.80087 |
| P67936 | TPM4 | -0.40581 | 9.11576 | -0.35869 | 0.721457 | 0.92169 | -5.6881 |
| Q9P258 | RCC2 | 0.196418 | 11.42629 | 0.358197 | 0.721557 | 0.92169 | -5.97602 |
| A0A0A0MT | #N/D | -0.55007 | 9.911469 | -0.35715 | 0.722633 | 0.92169 | -5.68858 |
| P01706 | #N/D | -0.28948 | 8.121541 | -0.35709 | 0.722639 | 0.92169 | -5.6421 |
| P46108 | CRK | 0.246273 | 8.591725 | 0.352829 | 0.725659 | 0.921818 | -5.87396 |
| P80723 | BASP1 | -0.31386 | 10.51359 | -0.35158 | 0.726483 | 0.921818 | -5.97827 |
| Q99439 | CNN2 | 0.227458 | 9.43566 | 0.35133 | 0.726698 | 0.921818 | -5.97834 |
| P02746 | C1QB | 0.255019 | 14.57706 | 0.350801 | 0.727068 | 0.921818 | -5.97854 |
| Q9UI12 | ATP6V1H | -0.34096 | 8.594544 | -0.34654 | 0.730297 | 0.924537 | -5.92491 |
| Q96GW7 | BCAN | 0.18975 | 8.242077 | 0.344141 | 0.7322 | 0.924577 | -5.87679 |
| P13637 | ATP1A3 | -0.26676 | 12.48297 | -0.34264 | 0.733165 | 0.924577 | -5.98125 |
| Q14203 | DCTN1 | 0.583541 | 7.611697 | 0.343609 | 0.733585 | 0.924577 | -5.28673 |
| P30405 | PPIF | -0.35432 | 7.967455 | -0.34139 | 0.73483 | 0.924779 | -5.44693 |
| P48047 | ATP5PO | -0.35857 | 12.76757 | -0.33692 | 0.737448 | 0.925277 | -5.98311 |
| P12270 | TPR | -0.30904 | 9.31589 | -0.33635 | 0.737975 | 0.925277 | -5.92819 |

| | | | | | | | |
|---|---|---|---|---|---|---|---|
| P62318 | SNRPD3 | 0.209869 | 9.893724 | 0.335541 | 0.738484 | 0.925277 | -5.98356 |
| P01009 | SERPINA1 | 0.167166 | 14.60704 | 0.329332 | 0.743146 | 0.929751 | -5.98554 |
| A0A0C4DH | #N/D | 0.274072 | 14.43238 | 0.326239 | 0.745518 | 0.930848 | -5.98648 |
| P21926 | CD9 | -0.25652 | 10.94564 | -0.32526 | 0.746208 | 0.930848 | -5.98682 |
| P11166 | SLC2A1 | 0.252533 | 9.8587 | 0.323099 | 0.747836 | 0.931515 | -5.98749 |
| Q13838 | DDX39B | 0.29857 | 13.4483 | 0.318184 | 0.751541 | 0.934375 | -5.989 |
| P17096 | HMGA1 | 0.313269 | 10.9518 | 0.317145 | 0.752325 | 0.934375 | -5.98932 |
| P52597 | HNRNPF | -0.17855 | 9.211955 | -0.31127 | 0.756787 | 0.93551 | -5.93599 |
| A0A087WS | #N/D | -0.28247 | 13.2827 | -0.31118 | 0.756835 | 0.93551 | -5.99112 |
| P01714 | #N/D | 0.269796 | 10.84083 | 0.310173 | 0.757593 | 0.93551 | -5.99142 |
| P01704 | #N/D | -0.19267 | 11.22917 | -0.31012 | 0.757631 | 0.93551 | -5.99143 |
| P27824 | CANX | -0.23991 | 11.05687 | -0.3071 | 0.75992 | 0.935863 | -5.99233 |
| P30038 | ALDH4A1 | -0.15837 | 11.58862 | -0.30601 | 0.760747 | 0.935863 | -5.99265 |
| P27695 | APEX1 | 0.5422 | 9.067064 | 0.305764 | 0.761316 | 0.935863 | -5.48362 |
| P16070 | CD44 | -0.39591 | 8.029181 | -0.30427 | 0.762311 | 0.935863 | -5.48409 |
| P10720 | PF4V1 | -0.2672 | 11.14838 | -0.29717 | 0.767472 | 0.937122 | -5.94008 |
| O00429 | DNM1L | -0.22683 | 7.713053 | -0.29698 | 0.767965 | 0.937122 | -5.75942 |
| P08758 | ANXA5 | 0.188105 | 12.52081 | 0.294912 | 0.769163 | 0.937122 | -5.99585 |
| A0A0C4DH | #N/D | 0.238023 | 9.100349 | 0.294878 | 0.769209 | 0.937122 | -5.99584 |
| Q14240 | EIF4A2 | 0.176693 | 9.225594 | 0.294341 | 0.769597 | 0.937122 | -5.99601 |
| P22314 | UBA1 | 0.193166 | 12.03395 | 0.290196 | 0.772749 | 0.937122 | -5.99717 |
| P04003 | C4BPA | -0.16074 | 14.39715 | -0.28987 | 0.773001 | 0.937122 | -5.99726 |
| P48740 | MASP1 | -0.2242 | 8.81856 | -0.28625 | 0.775797 | 0.937122 | -5.89414 |
| P50990 | CCT8 | -0.13715 | 13.71428 | -0.28618 | 0.775811 | 0.937122 | -5.99828 |
| O00187 | MASP2 | 0.247853 | 13.7813 | 0.285499 | 0.776346 | 0.937122 | -5.99846 |
| P55786 | NPEPPS | 0.174898 | 9.711469 | 0.284133 | 0.777368 | 0.937122 | -5.99884 |
| O43491-4 | #N/D | -0.19163 | 9.677072 | -0.28397 | 0.777495 | 0.937122 | -5.99889 |
| P12036 | NEFH | 0.106298 | 12.50765 | 0.283782 | 0.777635 | 0.937122 | -5.99894 |
| Q9UMS4 | PRPF19 | -0.23327 | 9.289913 | -0.28176 | 0.779179 | 0.937651 | -5.99949 |
| P11142 | HSPA8 | 0.141665 | 13.19735 | 0.27886 | 0.781391 | 0.937651 | -6.00027 |
| P46781 | RPS9 | -0.16524 | 10.5113 | -0.27272 | 0.786081 | 0.937651 | -6.00189 |

| | | | | | | | |
|---|---|---|---|---|---|---|---|
| P13797 | PLS3 | -0.17414 | 9.880516 | -0.27128 | 0.787184 | 0.937651 | -6.00227 |
| Q96F07 | CYFIP2 | 0.348723 | 9.351898 | 0.269886 | 0.788328 | 0.937651 | -5.86861 |
| P07360 | C8G | -0.24319 | 8.367863 | -0.26954 | 0.78892 | 0.937651 | -5.82735 |
| Q15233 | NONO | 0.177709 | 9.777373 | 0.268907 | 0.789 | 0.937651 | -6.00288 |
| P62263 | RPS14 | -0.21061 | 12.25025 | -0.26814 | 0.789586 | 0.937651 | -6.00308 |
| Q99880 | H2BC13 | -0.19531 | 12.3185 | -0.26739 | 0.790158 | 0.937651 | -6.00327 |
| O75083 | WDR1 | -0.13167 | 11.68371 | -0.26665 | 0.790729 | 0.937651 | -6.00346 |
| P15169 | CPN1 | -0.23008 | 9.464541 | -0.26509 | 0.791983 | 0.937651 | -5.89966 |
| P27361 | MAPK3 | 0.163091 | 9.665968 | 0.261038 | 0.795031 | 0.937651 | -6.00488 |
| Q5JNZ5 | #N/D | -0.20243 | 10.43992 | -0.25993 | 0.79588 | 0.937651 | -6.00516 |
| Q9BPU6 | DPYSL5 | -0.1505 | 10.87508 | -0.25977 | 0.796004 | 0.937651 | -6.0052 |
| P08670 | VIM | -0.10102 | 13.17483 | -0.25418 | 0.800294 | 0.937651 | -6.00658 |
| P07384 | CAPN1 | 0.189606 | 8.309043 | 0.253128 | 0.801141 | 0.937651 | -6.00682 |
| P09871 | C1S | -0.13894 | 13.55189 | -0.25028 | 0.803295 | 0.937651 | -6.00752 |
| Q13177 | PAK2 | -0.27645 | 8.338763 | -0.25057 | 0.803326 | 0.937651 | -5.59174 |
| P12277 | CKB | 0.1757 | 13.54112 | 0.249835 | 0.803639 | 0.937651 | -6.00763 |
| Q14697-2 | #N/D | -0.18099 | 9.735768 | -0.24957 | 0.803895 | 0.937651 | -5.90344 |
| P46778 | RPL21 | -0.54082 | 9.748979 | -0.24848 | 0.804771 | 0.937651 | -5.83264 |
| P41250 | GARS1 | -0.19168 | 9.552422 | -0.24767 | 0.805461 | 0.937651 | -5.95286 |
| Q16799 | RTN1 | -0.17312 | 10.36556 | -0.24596 | 0.806626 | 0.937651 | -6.00856 |
| P56747 | CLDN6 | -0.23511 | 8.759588 | -0.2438 | 0.80846 | 0.937651 | -5.67299 |
| Q16775 | HAGH | -0.2102 | 10.09877 | -0.24338 | 0.808608 | 0.937651 | -6.00916 |
| P00915 | CA1 | -0.16627 | 10.62668 | -0.24336 | 0.808626 | 0.937651 | -6.00916 |
| P11766 | ADH5 | 0.212381 | 10.15171 | 0.242534 | 0.809262 | 0.937651 | -6.00936 |
| Q01518 | CAP1 | 0.172042 | 11.32087 | 0.241778 | 0.809845 | 0.937651 | -6.00953 |
| Q9UKX2 | MYH2 | -0.25622 | 9.36748 | -0.2417 | 0.809989 | 0.937651 | -5.90525 |
| P52209 | PGD | -0.10379 | 9.886713 | -0.23687 | 0.813633 | 0.940591 | -6.01066 |
| P78417 | GSTO1 | 0.180551 | 11.81208 | 0.233316 | 0.816376 | 0.94091 | -6.01146 |
| P12814 | ACTN1 | 0.220983 | 7.963896 | 0.232639 | 0.81731 | 0.94091 | -5.72218 |
| P02647 | APOA1 | 0.167754 | 15.5712 | 0.232092 | 0.817322 | 0.94091 | -6.01174 |
| P26599 | PTBP1 | 0.117783 | 10.40181 | 0.230791 | 0.818327 | 0.94091 | -6.01203 |

| | | | | | | | |
|---|---|---|---|---|---|---|---|
| P61254 | RPL26 | 0.141223 | 10.98212 | 0.224119 | 0.823489 | 0.944792 | -6.01348 |
| P11169 | SLC2A3 | 0.20395 | 9.491977 | 0.221892 | 0.825244 | 0.944792 | -5.90963 |
| P02042 | HBD | 0.131335 | 10.61689 | 0.220659 | 0.826169 | 0.944792 | -6.01422 |
| P09471 | GNAO1 | -0.12359 | 11.83785 | -0.21653 | 0.829372 | 0.944792 | -6.01509 |
| P62424 | RPL7A | -0.08806 | 11.51419 | -0.2165 | 0.829391 | 0.944792 | -6.01509 |
| Q9BVA1 | TUBB2B | 0.201201 | 10.28254 | 0.215492 | 0.830175 | 0.944792 | -6.0153 |
| Q05193 | DNM1 | -0.11779 | 10.1149 | -0.21458 | 0.830886 | 0.944792 | -6.01549 |
| P06331 | #N/D | 0.148124 | 11.07987 | 0.214149 | 0.831217 | 0.944792 | -6.01558 |
| P01703 | #N/D | 0.184443 | 13.56328 | 0.208976 | 0.835233 | 0.944792 | -6.01663 |
| P06727 | #N/D | -0.09209 | 14.15249 | -0.20876 | 0.835403 | 0.944792 | -6.01668 |
| O75347 | TBCA | -0.14378 | 9.096358 | -0.20871 | 0.835438 | 0.944792 | -6.01668 |
| P29622 | SERPINA4 | -0.26912 | 8.812058 | -0.2065 | 0.837366 | 0.944792 | -5.47867 |
| P10643 | C7 | 0.101448 | 12.83675 | 0.205602 | 0.837855 | 0.944792 | -6.0173 |
| P53396 | ACLY | 0.209456 | 8.061162 | 0.202053 | 0.840673 | 0.944792 | -5.75947 |
| P01701 | #N/D | 0.17759 | 10.54093 | 0.201898 | 0.840793 | 0.944792 | -5.96273 |
| P62826 | RAN | 0.095811 | 12.49633 | 0.200371 | 0.841924 | 0.944792 | -6.01832 |
| P17844 | DDX5 | -0.09057 | 11.72587 | -0.19938 | 0.842693 | 0.944792 | -6.01851 |
| P07437 | TUBB | -0.15698 | 12.82403 | -0.19809 | 0.843701 | 0.944792 | -6.01876 |
| O94811 | TPPP | -0.09358 | 11.43773 | -0.19604 | 0.845296 | 0.944792 | -6.01915 |
| Q14123 | PDE1C | -0.26569 | 9.472294 | -0.19569 | 0.845597 | 0.944792 | -5.91482 |
| P61626 | LYZ | 0.154713 | 13.1144 | 0.195028 | 0.846084 | 0.944792 | -6.01934 |
| Q15149 | PLEC | 0.086113 | 12.56068 | 0.195009 | 0.846099 | 0.944792 | -6.01934 |
| Q13247 | SRSF6 | 0.125603 | 8.67543 | 0.186672 | 0.852625 | 0.950833 | -6.02086 |
| P01599 | #N/D | -0.16181 | 9.808588 | -0.18299 | 0.855498 | 0.95279 | -5.96621 |
| P61604 | HSPE1 | 0.126747 | 11.60591 | 0.178837 | 0.858718 | 0.955128 | -6.02225 |
| P36955 | SERPINF1 | -0.10387 | 12.75803 | -0.17499 | 0.861725 | 0.957223 | -6.0229 |
| Q16851 | UGP2 | 0.068226 | 12.72477 | 0.17294 | 0.863328 | 0.957755 | -6.02324 |
| O95782 | AP2A1 | -0.19899 | 8.500907 | -0.16746 | 0.867785 | 0.961017 | -5.51187 |
| P07108 | DBI | 0.141255 | 7.299068 | 0.165868 | 0.869031 | 0.961017 | -5.68748 |
| P60201 | PLP1 | -0.09006 | 12.86375 | -0.16215 | 0.871776 | 0.961017 | -6.02498 |
| P31939 | ATIC | 0.117899 | 12.54946 | 0.162012 | 0.871886 | 0.961017 | -6.025 |

| | | | | | | | |
|---|---|---|---|---|---|---|---|
| P08195 | SLC3A2 | 0.123631 | 10.25962 | 0.161996 | 0.871909 | 0.961017 | -6.025 |
| P35268 | RPL22 | 0.090332 | 11.96432 | 0.159055 | 0.874204 | 0.962302 | -6.02546 |
| P32119 | PRDX2 | 0.082843 | 13.16617 | 0.149329 | 0.881836 | 0.966555 | -6.0269 |
| P18669 | PGAM1 | -0.08141 | 13.03714 | -0.14841 | 0.882558 | 0.966555 | -6.02703 |
| Q15334 | LLGL1 | -0.10246 | 8.598936 | -0.1466 | 0.883983 | 0.966555 | -6.02729 |
| P16401 | H1-5 | 0.112213 | 10.04102 | 0.145744 | 0.884653 | 0.966555 | -6.02741 |
| A0A0C4DH | #N/D | 0.08022 | 12.25946 | 0.145385 | 0.884935 | 0.966555 | -6.02746 |
| P39748 | FEN1 | 0.176919 | 9.947938 | 0.144573 | 0.885572 | 0.966555 | -6.02757 |
| P11216 | PYGB | -0.07681 | 10.47934 | -0.14402 | 0.886009 | 0.966555 | -6.02765 |
| P02655 | APOC2 | -0.1068 | 11.22537 | -0.13941 | 0.88965 | 0.968634 | -5.97292 |
| O43143 | DHX15 | 0.117102 | 7.908015 | 0.138084 | 0.890859 | 0.968634 | -5.85261 |
| Q8N573 | OXR1 | 0.173708 | 8.278275 | 0.137561 | 0.891325 | 0.968634 | -5.51578 |
| Q9H299 | SH3BGRL3 | 0.127955 | 8.301393 | 0.133282 | 0.894546 | 0.97011 | -5.739 |
| Q9UQ80 | PA2G4 | 0.08766 | 9.950951 | 0.129911 | 0.897107 | 0.97011 | -6.0295 |
| O43426 | SYNJ1 | -0.26867 | 13.74991 | -0.12839 | 0.89831 | 0.97011 | -6.02969 |
| P14868-2 | #N/D | -0.10298 | 8.099459 | -0.12806 | 0.898653 | 0.97011 | -5.73963 |
| P49418 | AMPH | 0.102786 | 10.79496 | 0.12747 | 0.89903 | 0.97011 | -6.0298 |
| P00734 | F2 | 0.077736 | 14.7427 | 0.125653 | 0.900461 | 0.97011 | -6.03002 |
| P21281 | ATP6V1B2 | 0.086784 | 13.63954 | 0.125409 | 0.900654 | 0.97011 | -6.03005 |
| P61353 | RPL27 | 0.066427 | 9.636353 | 0.123859 | 0.901875 | 0.970199 | -6.03024 |
| Q14204 | DYNC1H1 | 0.058718 | 13.22936 | 0.121725 | 0.903557 | 0.970783 | -6.03049 |
| P52292 | KPNA2 | 0.10458 | 7.997937 | 0.118222 | 0.906413 | 0.972625 | -5.74074 |
| P52565 | ARHGDIA | 0.102512 | 10.94718 | 0.11443 | 0.909318 | 0.973227 | -6.03132 |
| P07225 | PROS1 | -0.04899 | 13.16055 | -0.11368 | 0.909902 | 0.973227 | -6.0314 |
| P61956 | SUMO2 | -0.10466 | 9.670243 | -0.11306 | 0.910401 | 0.973227 | -5.9761 |
| P14324 | FDPS | 0.084953 | 8.575581 | 0.111142 | 0.911983 | 0.973696 | -5.74149 |
| P49207 | RPL34 | 0.082308 | 9.206154 | 0.104446 | 0.9172 | 0.977758 | -6.03237 |
| P0C0L4 | C4A | -0.17821 | 11.8147 | -0.1034 | 0.918082 | 0.977758 | -5.51937 |
| P28074 | PSMB5 | 0.085212 | 8.249193 | 0.097202 | 0.922994 | 0.979968 | -5.74282 |
| P01611 | #N/D | 0.139024 | 10.90728 | 0.097127 | 0.923044 | 0.979968 | -5.51991 |
| Q01484 | ANK2 | -0.08121 | 9.391498 | -0.09544 | 0.924315 | 0.979968 | -5.97785 |

| | | | | | | | |
|---|---|---|---|---|---|---|---|
| P18621 | RPL17 | -0.09798 | 11.78909 | -0.09487 | 0.924758 | 0.979968 | -6.03329 |
| P53680 | AP2S1 | -0.08084 | 7.833872 | -0.08691 | 0.93115 | 0.983261 | -5.69656 |
| P49753 | ACOT2 | 0.058684 | 10.06568 | 0.085466 | 0.932199 | 0.983261 | -6.0341 |
| Q9P2D7-8 | #N/D | 0.072106 | 8.669631 | 0.083889 | 0.933447 | 0.983261 | -6.03423 |
| P20336 | RAB3A | -0.07467 | 9.798318 | -0.0838 | 0.933521 | 0.983261 | -6.03424 |
| A0A075B6I | #N/D | 0.046533 | 14.94383 | 0.08365 | 0.933636 | 0.983261 | -6.03425 |
| P67775 | PPP2CA | -0.07335 | 8.996608 | -0.07952 | 0.936956 | 0.985538 | -5.85863 |
| P07197 | NEFM | -0.03164 | 12.44543 | -0.07454 | 0.940851 | 0.986658 | -6.03494 |
| Q00765 | REEP5 | 0.063452 | 8.66685 | 0.072628 | 0.942408 | 0.986658 | -5.79803 |
| P15531 | NME1 | -0.0788 | 8.297834 | -0.07232 | 0.942677 | 0.986658 | -5.49465 |
| P06681 | C2 | 0.048325 | 10.88017 | 0.071458 | 0.943305 | 0.986658 | -5.93055 |
| P61313 | RPL15 | -0.03134 | 10.74977 | -0.0708 | 0.94381 | 0.986658 | -6.0352 |
| P50213 | IDH3A | 0.044035 | 10.5341 | 0.063564 | 0.949546 | 0.991437 | -6.03567 |
| P17655 | CAPN2 | 0.057692 | 9.117694 | 0.061333 | 0.951352 | 0.992108 | -5.74544 |
| Q96FW1 | OTUB1 | -0.0619 | 7.493366 | -0.05361 | 0.957524 | 0.993821 | -5.44697 |
| Q5IS74 | #N/D | 0.043727 | 8.979446 | 0.052476 | 0.958342 | 0.993821 | -5.98088 |
| P02763 | ORM1 | 0.038667 | 11.84132 | 0.052423 | 0.95838 | 0.993821 | -6.03629 |
| A0A0B4J1X | #N/D | 0.069268 | 10.0226 | 0.051742 | 0.958924 | 0.993821 | -5.98092 |
| P61088 | UBE2N | -0.03653 | 11.45261 | -0.04883 | 0.961236 | 0.993821 | -6.03646 |
| P61020 | RAB5B | -0.02986 | 8.987505 | -0.04625 | 0.963286 | 0.993821 | -5.90198 |
| P35580 | MYH10 | 0.033899 | 9.704816 | 0.045751 | 0.963673 | 0.993821 | -6.03661 |
| Q9UQM7 | CAMK2A | 0.034624 | 11.74729 | 0.043947 | 0.965105 | 0.993821 | -6.03668 |
| P00558 | PGK1 | -0.01852 | 12.86953 | -0.04268 | 0.966109 | 0.993821 | -6.03674 |
| Q9HB71 | CACYBP | 0.04098 | 8.507432 | 0.04035 | 0.967991 | 0.993821 | -5.86084 |
| P32322 | PYCR1 | 0.048655 | 8.954011 | 0.039301 | 0.968797 | 0.993821 | -6.03687 |
| Q5HYA8 | TMEM67 | -0.03098 | 9.59421 | -0.03821 | 0.969658 | 0.993821 | -6.03691 |
| A0A075B6I | #N/D | 0.049523 | 7.939523 | 0.038296 | 0.969666 | 0.993821 | -5.52334 |
| O95989 | NUDT3 | 0.027769 | 9.707781 | 0.036321 | 0.971157 | 0.993821 | -6.03698 |
| P23526 | AHCY | -0.01831 | 12.4871 | -0.03596 | 0.97144 | 0.993821 | -6.03699 |
| P20339 | RAB5A | 0.020853 | 8.711876 | 0.03569 | 0.971658 | 0.993821 | -6.037 |
| P01591 | JCHAIN | 0.018507 | 14.17635 | 0.031673 | 0.974847 | 0.99494 | -6.03713 |

| | | | | | | | |
|---|---|---|---|---|---|---|---|
| P62987 | UBA52 | 0.011056 | 11.77756 | 0.028614 | 0.977275 | 0.99494 | -6.03722 |
| Q13561 | DCTN2 | -0.01793 | 8.671465 | -0.0286 | 0.977288 | 0.99494 | -6.03722 |
| Q9NR30 | DDX21 | 0.01803 | 8.743727 | 0.028434 | 0.977424 | 0.99494 | -6.03722 |
| P25786 | PSMA1 | 0.024582 | 8.552348 | 0.024481 | 0.980576 | 0.996764 | -5.86132 |
| P46782 | RPS5 | -0.01723 | 8.807193 | -0.02299 | 0.981749 | 0.996764 | -5.86136 |
| P09429 | HMGB1 | -0.01036 | 11.33429 | -0.02132 | 0.983068 | 0.996764 | -6.03739 |
| P13671 | C6 | -0.00921 | 13.17346 | -0.01957 | 0.984454 | 0.996764 | -6.03743 |
| Q00325 | SLC25A3 | 0.016602 | 11.25621 | 0.018804 | 0.985065 | 0.996764 | -6.03744 |
| P00747 | PLG | -0.01051 | 12.64273 | -0.01677 | 0.986682 | 0.996901 | -6.03748 |
| P62805 | H4C9 | 0.011893 | 11.74498 | 0.015304 | 0.987845 | 0.996901 | -6.0375 |
| P37108 | SRP14 | -0.0165 | 8.187199 | -0.01329 | 0.989477 | 0.996901 | -5.61962 |
| P51659 | HSD17B4 | -0.0061 | 12.08701 | -0.01203 | 0.990448 | 0.996901 | -6.03754 |
| E9PAV3 | #N/D | 0.009659 | 12.14083 | 0.011268 | 0.991051 | 0.996901 | -6.03755 |
| Q9H0U4 | RAB1B | 0.006572 | 8.154821 | 0.009292 | 0.992627 | 0.997309 | -5.77828 |
| P62829 | RPL23 | 0.009133 | 7.660806 | 0.005953 | 0.995281 | 0.997793 | -5.49686 |
| Q9UN36 | NDRG2 | -0.0042 | 11.61186 | -0.00573 | 0.995451 | 0.997793 | -6.0376 |
| Q9NZL9 | MAT2B | 0.002804 | 9.370227 | 0.003311 | 0.99737 | 0.998542 | -6.03761 |
| P01780 | #N/D | -1.8E-05 | 11.11587 | -2.2E-05 | 0.999983 | 0.999983 | -6.03761 |
| P21796 | VDAC1 | #N/D | 8.076572 | #N/D | #N/D | #N/D | #N/D |
| P48426 | PIP4K2A | #N/D | 8.06123 | #N/D | #N/D | #N/D | #N/D |
| P30048 | PRDX3 | #N/D | 9.178145 | #N/D | #N/D | #N/D | #N/D |
| P83731 | RPL24 | #N/D | 8.556325 | #N/D | #N/D | #N/D | #N/D |
| A0A0B4J1V | #N/D | #N/D | 8.761429 | #N/D | #N/D | #N/D | #N/D |
| P30040 | ERP29 | #N/D | 8.785068 | #N/D | #N/D | #N/D | #N/D |
| P63162 | SNRPN | #N/D | 6.935315 | #N/D | #N/D | #N/D | #N/D |
| P68402 | PAFAH1B2 | #N/D | 6.807079 | #N/D | #N/D | #N/D | #N/D |
| Q7L0J3 | SV2A | #N/D | 9.396699 | #N/D | #N/D | #N/D | #N/D |
| Q13153 | PAK1 | #N/D | 7.407468 | #N/D | #N/D | #N/D | #N/D |
| Q14152 | EIF3A | #N/D | 8.131085 | #N/D | #N/D | #N/D | #N/D |
| Q8WUM4 | PDCD6IP | #N/D | 7.545402 | #N/D | #N/D | #N/D | #N/D |
| O14980 | XPO1 | #N/D | 7.346187 | #N/D | #N/D | #N/D | #N/D |

| | | | | | | | |
|---|---|---|---|---|---|---|---|
| P00403 | COX2 | #N/D | 8.386699 | #N/D | #N/D | #N/D | #N/D |
| Q71U36 | TUBA1A | #N/D | 13.10907 | #N/D | #N/D | #N/D | #N/D |
| O75340 | PDCD6 | #N/D | 7.338176 | #N/D | #N/D | #N/D | #N/D |
| Q99714 | HSD17B10 | #N/D | 7.015178 | #N/D | #N/D | #N/D | #N/D |
| Q9NRX4 | PHPT1 | #N/D | 8.126823 | #N/D | #N/D | #N/D | #N/D |
| Q8WVE0 | EEF1AKMT1 | #N/D | 12.59871 | #N/D | #N/D | #N/D | #N/D |

**Supplemental Table 5.** Differentially expressed proteins in microglial/macrophage extracellular vesicles (m/mEVs) isolated from patients following CAND treatment compared to the pre-CAND condition and the complete list of all proteins identified in the proteomic analysis. Each row corresponds to a protein identified by its UniProt accession number, gene symbol, and protein symbol. Expression changes are reported as $\log_2$ fold change (logFC); positive values indicate upregulation after CAND treatment, whereas negative values indicate downregulation. AveExpr represents the average expression across all samples. The t-statistic and associated P value were derived from moderated t-tests. The B statistic reflects the log-odds that a given protein is differentially expressed.

#### Supplemental Table 5.  Differentially Expressed Proteins in m/mEVs

| UniProt | Gene Symbol | Protein name | logFC | AveExpr | t | P.Value | B |
|---|---|---|---|---|---|---|---|
| Q15435 | PPP1R7 | PP1R7 | 2.534947318 | 8.920445915 | 2.952735327 | 0.00480944 | -4.527624198 |
| Q9UNZ2 | NSFL1C | NSF1C | 2.520283753 | 8.711790029 | 3.353324711 | 0.001521636 | -4.508890721 |
| P67775 | PPP2CA | PP2AA | 2.322136881 | 8.996608308 | 2.632901881 | 0.011393591 | -4.55007742 |
| Q99729-3 | HNRNPAB | ROAA | 2.314112672 | 12.65612226 | 2.149870678 | 0.035945733 | -4.556626472 |
| P37108 | SRP14 | SRP14 | 2.258611349 | 8.187199007 | 2.227007246 | 0.032795541 | -4.571042985 |
| P02652 | APOA2 | APOA2 | 2.086381925 | 13.89357777 | 3.271874042 | 0.001841711 | -4.501181894 |
| P36354 | N | NCAP | 2.06656752 | 8.621130297 | 2.234223072 | 0.03066834 | -4.561957378 |
| Q9H4G0 | EPB41L1 | E41L1 | 2.0274615 | 10.09244966 | 3.088755709 | 0.003139284 | -4.511001909 |
| P49411 | TUFM | EFTU | 1.876021433 | 11.65540175 | 2.253440973 | 0.028207904 | -4.552087288 |
| P60880 | SNAP25 | SNP25 | 1.871082842 | 10.94386326 | 2.781958293 | 0.00737178 | -4.526893083 |
| P01703 | IGLV1-40 | LV140 | 1.84821078 | 13.56328335 | 2.094045794 | 0.040843173 | -4.55901158 |
| A0MZ66 | SHTN1 | SHOT1 | 1.811463761 | 8.930067546 | 2.137163184 | 0.037663073 | -4.561658646 |
| P04433 | IGKV3-11 | KV311 | 1.770918077 | 12.95042165 | 2.018613079 | 0.048377072 | -4.562163168 |
| P61106 | RAB14 | RAB14 | 1.729081762 | 9.911827099 | 2.870355222 | 0.005828909 | -4.522566343 |
| P60953 | CDC42 | CDC42 | 1.711517691 | 9.514287631 | 2.156974943 | 0.035704933 | -4.556549746 |
| P07358 | C8B | CO8B | 1.571495048 | 13.4655143 | 2.368298037 | 0.021384381 | -4.546888284 |
| P20700 | LMNB1 | LMNB1 | 1.549423474 | 10.71505267 | 2.019122623 | 0.048322404 | -4.56214216 |
| P03952 | KLKB1 | KLKB1 | 1.48721322 | 13.33419021 | 2.175842588 | 0.033848101 | -4.555501937 |
| P35268 | RPL22 | RL22 | 1.435786589 | 11.9643162 | 2.528125173 | 0.014345733 | -4.539384924 |
| P16152 | CBR1 | CBR1 | 1.371197738 | 14.15987142 | 2.405303981 | 0.019524027 | -4.545177961 |
| P35542 | SAA4 | SAA4 | 1.318791689 | 12.48858297 | 2.561351945 | 0.013177153 | -4.537788173 |
| Q96PD5 | PGLYRP2 | PGRP2 | 1.202383047 | 13.17714881 | 2.149744003 | 0.035956237 | -4.556631933 |
| P19827 | ITIH1 | ITIH1 | 1.162279118 | 15.08481182 | 2.723145485 | 0.008630431 | -4.529845315 |
| P16112 | ACAN | PGCA | -1.272601077 | 8.198785371 | -2.176279775 | 0.034443484 | -4.555924397 |
| P22087 | FBL | FBRL | -1.292798381 | 8.631299977 | -2.136236421 | 0.037638065 | -4.561635606 |
| P08237-3 | PFKM | PFKAM | -1.32040248 | 9.903447066 | -2.584032694 | 0.012589968 | -4.537045037 |
| Q96F85 | CNRIP1 | CNRP1 | -1.328717271 | 10.63718676 | -2.306203971 | 0.024864221 | -4.549719917 |
| P19823 | ITIH2 | ITIH2 | -1.374399877 | 14.69691094 | -2.336169143 | 0.023126488 | -4.548359494 |

| | | | | | | | |
|---|---|---|---|---|---|---|---|
| P48643 | CCT5 | TCPE | -1.387970379 | 9.491953511 | -2.068292959 | 0.043291812 | -4.56009687 |
| P25398 | RPS12 | RS12 | -1.408102401 | 8.911054274 | -2.320981031 | 0.024210859 | -4.549280468 |
| P63000 | RAC1 | RAC1 | -1.529276926 | 11.00234243 | -2.250677707 | 0.028393508 | -4.552210268 |
| P48539 | PCP4 | PCP4 | -1.563497866 | 8.670541913 | -2.86234989 | 0.005993691 | -4.523149291 |
| P39687 | ANP32A | AN32A | -1.569896551 | 9.050789322 | -2.315205599 | 0.024628028 | -4.549621348 |
| P54652 | HSPA2 | HSP72 | -1.572951859 | 13.29814995 | -2.734436376 | 0.00837446 | -4.52928112 |
| Q9UHG2 | PCSK1N | PCS1N | -1.62226005 | 7.955461425 | -2.185893907 | 0.034530256 | -4.563747278 |
| Q15084 | PDIA6 | PDIA6 | -1.664103119 | 12.47947733 | -2.814284521 | 0.006754242 | -4.525256547 |
| A0A0C4DH38 | IGHV5-51 | HV551 | -1.690624118 | 12.36128945 | -2.247329661 | 0.028619838 | -4.552359138 |
| P17655 | CAPN2 | CAN2 | -1.811555275 | 9.11769381 | -2.637126637 | 0.011270921 | -4.535217381 |
| IGHV5-51 | IGHV5-51 | HV551 | -1.836664333 | 7.847079482 | -2.48360594 | 0.017141699 | -4.55597882 |
| P83731 | RPL24 | RL24 | -1.944873098 | 8.55632495 | -2.48198105 | 0.017107336 | -4.553351196 |
| Q9UBB6 | NCDN | NCDN | -2.016441973 | 8.646641197 | -2.459512676 | 0.017182814 | -4.542833471 |
| P49368 | CCT3 | TCPG | -2.247995622 | 10.62952483 | -2.155093949 | 0.03551495 | -4.556401065 |
| P46783 | RPS10 | RS10 | -2.319166642 | 10.88472705 | -2.785051689 | 0.007310509 | -4.526736897 |
| P16070 | CD44 | CD44 | -2.3686112 | 8.02918107 | -2.432587673 | 0.01899246 | -4.566157982 |
| Q02218 | OGDH | ODO1 | -2.372367518 | 8.055839204 | -2.919706136 | 0.0054849 | -4.553061568 |
| P30626 | SRI | SORCN | -2.396588048 | 7.800030592 | -2.70022077 | 0.010346319 | -4.558319161 |
| Q14152 | EIF3A | EIF3A | -2.44709524 | 8.131085478 | -2.539895843 | 0.015139659 | -4.563926646 |
| P11586 | MTHFD1 | C1TC | -4.088652797 | 11.82918692 | -2.10121402 | 0.040257158 | -4.558750211 |

#### Supplemental Table 5. All Quantified proteins

| UniProt | Gene Symbol | logFC | AveExpr | t | P.Value | adj.P.Val | B |
|---|---|---|---|---|---|---|---|
| Q9UNZ2 | NSFL1C | 2.520284 | 8.71179 | 3.353325 | 0.001522 | 0.624987 | -4.50889 |
| P02652 | APOA2 | 2.086382 | 13.89358 | 3.271874 | 0.001842 | 0.624987 | -4.50118 |
| Q9H4G0 | EPB41L1 | 2.027462 | 10.09245 | 3.088756 | 0.003139 | 0.624987 | -4.511 |
| Q15435 | PPP1R7 | 2.534947 | 8.920446 | 2.952735 | 0.004809 | 0.624987 | -4.52762 |
| Q02218 | OGDH | -2.37237 | 8.055839 | -2.91971 | 0.005485 | 0.624987 | -4.55306 |
| P61106 | RAB14 | 1.729082 | 9.911827 | 2.870355 | 0.005829 | 0.624987 | -4.52257 |
| P48539 | PCP4 | -1.5635 | 8.670542 | -2.86235 | 0.005994 | 0.624987 | -4.52315 |
| Q15084 | PDIA6 | -1.6641 | 12.47948 | -2.81428 | 0.006754 | 0.624987 | -4.52526 |
| P46783 | RPS10 | -2.31917 | 10.88473 | -2.78505 | 0.007311 | 0.624987 | -4.52674 |
| P60880 | SNAP25 | 1.871083 | 10.94386 | 2.781958 | 0.007372 | 0.624987 | -4.52689 |
| P54652 | HSPA2 | -1.57295 | 13.29815 | -2.73444 | 0.008374 | 0.624987 | -4.52928 |
| P19827 | ITIH1 | 1.162279 | 15.08481 | 2.723145 | 0.00863 | 0.624987 | -4.52985 |
| P30626 | SRI | -2.39659 | 7.800031 | -2.70022 | 0.010346 | 0.660069 | -4.55832 |
| P17655 | CAPN2 | -1.81156 | 9.117694 | -2.63713 | 0.011271 | 0.660069 | -4.53522 |
| P67775 | PPP2CA | 2.322137 | 8.996608 | 2.632902 | 0.011394 | 0.660069 | -4.55008 |
| P08237-3 | #N/D | -1.3204 | 9.903447 | -2.58403 | 0.01259 | 0.673585 | -4.53705 |
| P35542 | SAA4 | 1.318792 | 12.48858 | 2.561352 | 0.013177 | 0.673585 | -4.53779 |
| P35268 | RPL22 | 1.435787 | 11.96432 | 2.528125 | 0.014346 | 0.678655 | -4.53938 |
| Q14152 | EIF3A | -2.4471 | 8.131085 | -2.5399 | 0.01514 | 0.678655 | -4.56393 |
| P83731 | RPL24 | -1.94487 | 8.556325 | -2.48198 | 0.017107 | 0.678655 | -4.55335 |
| Q9BWD1-2 | #N/D | -1.83666 | 7.847079 | -2.48361 | 0.017142 | 0.678655 | -4.55598 |
| Q9UBB6 | NCDN | -2.01644 | 8.646641 | -2.45951 | 0.017183 | 0.678655 | -4.54283 |
| P36578 | RPL4 | -0.96052 | 12.0117 | -2.41684 | 0.018975 | 0.678655 | -4.54464 |
| P16070 | CD44 | -2.36861 | 8.029181 | -2.43259 | 0.018992 | 0.678655 | -4.56616 |
| P16152 | CBR1 | 1.371198 | 14.15987 | 2.405304 | 0.019524 | 0.678655 | -4.54518 |
| P07358 | C8B | 1.571495 | 13.46551 | 2.368298 | 0.021384 | 0.710136 | -4.54689 |
| P19823 | ITIH2 | -1.3744 | 14.69691 | -2.33617 | 0.023126 | 0.710136 | -4.54836 |
| P25398 | RPS12 | -1.4081 | 8.911054 | -2.32098 | 0.024211 | 0.710136 | -4.54928 |

| | | | | | | | |
|---|---|---|---|---|---|---|---|
| P39687 | ANP32A | -1.5699 | 9.050789 | -2.31521 | 0.024628 | 0.710136 | -4.54962 |
| Q96F85 | CNRIP1 | -1.32872 | 10.63719 | -2.3062 | 0.024864 | 0.710136 | -4.54972 |
| P49411 | TUFM | 1.876021 | 11.6554 | 2.253441 | 0.028208 | 0.710136 | -4.55209 |
| P63000 | RAC1 | -1.52928 | 11.00234 | -2.25068 | 0.028394 | 0.710136 | -4.55221 |
| A0A0C4DH | #N/D | -1.69062 | 12.36129 | -2.24733 | 0.02862 | 0.710136 | -4.55236 |
| P63104 | YWHAZ | -0.95574 | 13.41142 | -2.24027 | 0.029102 | 0.710136 | -4.55267 |
| P36354 | #N/D | 2.066568 | 8.62113 | 2.234223 | 0.030668 | 0.710136 | -4.56196 |
| P06733 | ENO1 | -0.91826 | 12.59836 | -2.20007 | 0.031989 | 0.710136 | -4.55444 |
| P37108 | SRP14 | 2.258611 | 8.187199 | 2.227007 | 0.032796 | 0.710136 | -4.57104 |
| P03952 | KLKB1 | 1.487213 | 13.33419 | 2.175843 | 0.033848 | 0.710136 | -4.5555 |
| P16112 | #N/D | -1.2726 | 8.198785 | -2.17628 | 0.034443 | 0.710136 | -4.55592 |
| Q9UHG2 | PCSK1N | -1.62226 | 7.955461 | -2.18589 | 0.03453 | 0.710136 | -4.56375 |
| P49368 | CCT3 | -2.248 | 10.62952 | -2.15509 | 0.035515 | 0.710136 | -4.5564 |
| P60953 | CDC42 | 1.711518 | 9.514288 | 2.156975 | 0.035705 | 0.710136 | -4.55655 |
| Q99729-3 | #N/D | 2.314113 | 12.65612 | 2.149871 | 0.035946 | 0.710136 | -4.55663 |
| Q96PD5 | PGLYRP2 | 1.202383 | 13.17715 | 2.149744 | 0.035956 | 0.710136 | -4.55663 |
| P22087 | FBL | -1.2928 | 8.6313 | -2.13624 | 0.037638 | 0.711505 | -4.56164 |
| A0MZ66 | SHTN1 | 1.811464 | 8.930068 | 2.137163 | 0.037663 | 0.711505 | -4.56166 |
| P11586 | MTHFD1 | -4.08865 | 11.82919 | -2.10121 | 0.040257 | 0.739432 | -4.55875 |
| P01703 | #N/D | 1.848211 | 13.56328 | 2.094046 | 0.040843 | 0.739432 | -4.55901 |
| P48643 | CCT5 | -1.38797 | 9.491954 | -2.06829 | 0.043292 | 0.767767 | -4.5601 |
| P20700 | LMNB1 | 1.549423 | 10.71505 | 2.019123 | 0.048322 | 0.808298 | -4.56214 |
| P04433 | #N/D | 1.770918 | 12.95042 | 2.018613 | 0.048377 | 0.808298 | -4.56216 |
| O94811 | TPPP | 0.952806 | 11.43773 | 1.99598 | 0.05086 | 0.808298 | -4.56309 |
| P62241 | RPS8 | -1.15608 | 9.619243 | -1.98655 | 0.051926 | 0.808298 | -4.56348 |
| O00425 | IGF2BP3 | 1.511308 | 8.291206 | 1.999213 | 0.052019 | 0.808298 | -4.56977 |
| P10768 | ESD | -1.43927 | 10.75161 | -1.98573 | 0.05202 | 0.808298 | -4.56351 |
| P46782 | RPS5 | 1.310201 | 8.807193 | 1.990443 | 0.052088 | 0.808298 | -4.56699 |
| P25788 | PSMA3 | 1.338579 | 9.356898 | 1.976884 | 0.053537 | 0.81139 | -4.56407 |
| P05787 | KRT8 | -1.04179 | 11.88015 | -1.96736 | 0.054155 | 0.81139 | -4.56426 |
| P27797 | CALR | 1.044414 | 10.33747 | 1.934144 | 0.058208 | 0.826619 | -4.56559 |

| | | | | | | | |
|---|---|---|---|---|---|---|---|
| P05141 | SLC25A5 | -1.56119 | 10.10956 | -1.92907 | 0.058944 | 0.826619 | -4.56583 |
| Q14520 | HABP2 | 1.751406 | 9.292678 | 1.927503 | 0.059344 | 0.826619 | -4.56912 |
| P35527 | KRT9 | 1.273218 | 15.65986 | 1.901531 | 0.062437 | 0.826619 | -4.56689 |
| P56747 | CLDN6 | 1.423651 | 8.759588 | 1.90584 | 0.062875 | 0.826619 | -4.57254 |
| P0DP09 | #N/D | 1.400567 | 13.65182 | 1.900998 | 0.062914 | 0.826619 | -4.56704 |
| Q9GZV7 | HAPLN2 | -1.09974 | 13.03861 | -1.89349 | 0.063518 | 0.826619 | -4.5672 |
| Q07954 | LRP1 | -1.37289 | 9.068573 | -1.89385 | 0.06377 | 0.826619 | -4.56728 |
| P28482 | MAPK1 | -0.90162 | 10.57932 | -1.88819 | 0.064336 | 0.826619 | -4.56744 |
| Q9UHY7 | ENOPH1 | 1.476945 | 8.101761 | 1.893306 | 0.064699 | 0.826619 | -4.58024 |
| P39748 | FEN1 | -2.29756 | 9.947938 | -1.8775 | 0.065716 | 0.826619 | -4.56783 |
| P01704 | #N/D | 1.161085 | 11.22917 | 1.868909 | 0.066924 | 0.826619 | -4.56816 |
| O14980 | XPO1 | -1.60554 | 7.346187 | -1.88213 | 0.067614 | 0.826619 | -4.5807 |
| P50454 | SERPINH1 | -1.17709 | 9.103501 | -1.85865 | 0.068489 | 0.826619 | -4.56859 |
| P62937 | PPIA | -0.92382 | 11.22551 | -1.8452 | 0.070354 | 0.837499 | -4.56908 |
| Q9NRW1 | RAB6B | 1.252996 | 9.368933 | 1.838473 | 0.071555 | 0.840293 | -4.5694 |
| O75363-2 | #N/D | 1.338141 | 8.935009 | 1.827514 | 0.073108 | 0.847076 | -4.56979 |
| P01023 | A2M | -1.30658 | 15.60344 | -1.80982 | 0.075748 | 0.860314 | -4.57044 |
| P07451 | CA3 | 1.575665 | 9.536455 | 1.807058 | 0.076283 | 0.860314 | -4.57057 |
| Q00765 | REEP5 | -1.29241 | 8.66685 | -1.80057 | 0.078133 | 0.860314 | -4.57362 |
| P13647 | KRT5 | 1.29529 | 13.1342 | 1.786963 | 0.079413 | 0.860314 | -4.5713 |
| P00748 | F12 | -0.88283 | 11.60206 | -1.78143 | 0.080322 | 0.860314 | -4.57151 |
| Q13228 | SELENBP1 | -0.94143 | 10.91175 | -1.77068 | 0.082114 | 0.860314 | -4.57191 |
| P26599 | PTBP1 | -0.90306 | 10.40181 | -1.76951 | 0.082312 | 0.860314 | -4.57195 |
| P0DP25 | CALM1 | -2.41335 | 11.85239 | -1.7694 | 0.08233 | 0.860314 | -4.57196 |
| P18621 | RPL17 | -1.81178 | 11.78909 | -1.75423 | 0.084919 | 0.860314 | -4.57252 |
| P40429 | RPL13A | 1.089736 | 11.00395 | 1.742286 | 0.087006 | 0.860314 | -4.57296 |
| P11137 | MAP2 | 1.016724 | 10.40085 | 1.738297 | 0.087713 | 0.860314 | -4.57311 |
| Q96GW7 | BCAN | 0.986948 | 8.242077 | 1.733143 | 0.089307 | 0.860314 | -4.57874 |
| P49591 | SARS1 | -1.12514 | 8.044302 | -1.72778 | 0.090822 | 0.860314 | -4.57605 |
| Q13838 | DDX39B | 1.610137 | 13.4483 | 1.715912 | 0.091765 | 0.860314 | -4.57393 |
| Q99880 | H2BC13 | -1.25026 | 12.3185 | -1.71167 | 0.09255 | 0.860314 | -4.57408 |

| | | | | | | | |
|---|---|---|---|---|---|---|---|
| P35232 | PHB | -0.84979 | 11.88214 | -1.7034 | 0.094097 | 0.860314 | -4.57438 |
| P27361 | MAPK3 | 1.062647 | 9.665968 | 1.700839 | 0.094579 | 0.860314 | -4.57447 |
| Q92597 | NDRG1 | 1.128364 | 10.27498 | 1.689241 | 0.097003 | 0.860314 | -4.57493 |
| P13861 | PRKAR2A | -1.43518 | 8.585476 | -1.69011 | 0.097062 | 0.860314 | -4.57712 |
| P14618-3 | #N/D | -1.11165 | 9.955675 | -1.67896 | 0.098894 | 0.860314 | -4.57528 |
| P15531 | NME1 | 1.066769 | 8.297834 | 1.68532 | 0.099103 | 0.860314 | -4.57744 |
| P14174 | MIF | -1.46008 | 11.07338 | -1.67165 | 0.100231 | 0.860314 | -4.57552 |
| P06727 | #N/D | -0.73213 | 14.15249 | -1.65972 | 0.10262 | 0.860314 | -4.57595 |
| P51649 | ALDH5A1 | -0.76074 | 11.25326 | -1.65587 | 0.1034 | 0.860314 | -4.57608 |
| P23284 | PPIB | -1.26898 | 11.31656 | -1.65557 | 0.103784 | 0.860314 | -4.57614 |
| Q9HB71 | CACYBP | 1.469301 | 8.507432 | 1.647139 | 0.106433 | 0.860314 | -4.57859 |
| Q06033 | ITIH3 | -1.95718 | 13.55378 | -1.63869 | 0.106944 | 0.860314 | -4.57669 |
| Q9Y4L1 | HYOU1 | -0.97576 | 8.672707 | -1.63089 | 0.108689 | 0.860314 | -4.57698 |
| P14415 | ATP1B2 | -1.06664 | 8.426782 | -1.63026 | 0.109279 | 0.860314 | -4.57706 |
| P07477 | PRSS1 | 2.363862 | 13.06236 | 1.621247 | 0.110746 | 0.860314 | -4.57731 |
| P62753 | RPS6 | 1.202521 | 9.50014 | 1.615686 | 0.111842 | 0.860314 | -4.57749 |
| Q5IS67 | #N/D | 1.749589 | 8.456854 | 1.618759 | 0.112424 | 0.860314 | -4.58185 |
| P0DOX2 | #N/D | 0.64514 | 13.86052 | 1.609469 | 0.113197 | 0.860314 | -4.5777 |
| P38606 | ATP6V1A | 1.125885 | 12.55831 | 1.595311 | 0.116331 | 0.860314 | -4.57819 |
| A0A0A0MS | #N/D | 1.094062 | 10.69979 | 1.594908 | 0.116421 | 0.860314 | -4.57821 |
| O95197 | RTN3 | -1.60156 | 8.031271 | -1.59884 | 0.11805 | 0.860314 | -4.58496 |
| P05783 | KRT18 | 1.419832 | 11.19722 | 1.586305 | 0.118573 | 0.860314 | -4.57853 |
| Q6TUY0 | #N/D | -2.38054 | 12.02649 | -1.5802 | 0.119965 | 0.860314 | -4.57873 |
| P0DJI8 | SAA1 | 1.232771 | 12.87564 | 1.57772 | 0.120322 | 0.860314 | -4.57879 |
| O00264 | PGRMC1 | -1.16044 | 9.27944 | -1.56611 | 0.123579 | 0.860314 | -4.57925 |
| P27169 | PON1 | 1.503288 | 14.83311 | 1.563857 | 0.123649 | 0.860314 | -4.58099 |
| O75390 | CS | -1.32293 | 11.12214 | -1.56044 | 0.12435 | 0.860314 | -4.57938 |
| Q6YN16 | HSDL2 | 1.573702 | 9.242755 | 1.561981 | 0.124427 | 0.860314 | -4.58245 |
| P55786 | NPEPPS | -0.95843 | 9.711469 | -1.55704 | 0.125155 | 0.860314 | -4.57949 |
| Q99439 | CNN2 | -1.00592 | 9.43566 | -1.55374 | 0.126045 | 0.860314 | -4.57961 |
| A0A0B4J1Y | #N/D | -1.12319 | 11.1449 | -1.55312 | 0.126087 | 0.860314 | -4.57962 |

| | | | | | | | |
|---|---|---|---|---|---|---|---|
| P78347 | GTF2I | -0.94295 | 9.347322 | -1.54526 | 0.12808 | 0.860314 | -4.5799 |
| P01718 | #N/D | -1.01784 | 12.55539 | -1.54009 | 0.12923 | 0.860314 | -4.58006 |
| P49913 | CAMP | -1.36121 | 8.94521 | -1.54479 | 0.129493 | 0.860314 | -4.58697 |
| P51178-2 | #N/D | -1.26671 | 9.240432 | -1.5403 | 0.12974 | 0.860314 | -4.58011 |
| P35580 | MYH10 | -1.13388 | 9.704816 | -1.53033 | 0.131624 | 0.860314 | -4.58038 |
| P49006 | MARCKSL1 | 1.546656 | 8.963973 | 1.529923 | 0.131937 | 0.860314 | -4.58042 |
| P08697 | SERPINF2 | 0.692332 | 12.89207 | 1.519057 | 0.134433 | 0.860314 | -4.58076 |
| Q12905 | ILF2 | -1.08864 | 9.328399 | -1.51947 | 0.134543 | 0.860314 | -4.58076 |
| O95394 | PGM3 | -1.13797 | 8.458672 | -1.51825 | 0.134742 | 0.860314 | -4.58079 |
| P67936 | TPM4 | -1.40449 | 9.11576 | -1.5204 | 0.135193 | 0.860314 | -4.58361 |
| O00429 | DNM1L | -1.19025 | 7.713053 | -1.52084 | 0.1359 | 0.860314 | -4.58688 |
| P07357 | C8A | -0.85363 | 14.30403 | -1.51007 | 0.136708 | 0.860314 | -4.58105 |
| O14967 | CLGN | 1.1554 | 11.39778 | 1.510922 | 0.137308 | 0.860314 | -4.58383 |
| P31025 | LCN1 | 1.143102 | 14.44478 | 1.50689 | 0.137519 | 0.860314 | -4.58115 |
| Q5IS61 | #N/D | 1.309104 | 8.614239 | 1.506763 | 0.138238 | 0.860314 | -4.58707 |
| P04080 | CSTB | 1.21303 | 11.10002 | 1.501805 | 0.138825 | 0.860314 | -4.58132 |
| O43301 | HSPA12A | 0.878911 | 10.41348 | 1.49676 | 0.140342 | 0.860314 | -4.5815 |
| P22234 | PAICS | -0.89246 | 10.08226 | -1.49409 | 0.140825 | 0.860314 | -4.58157 |
| P27635 | RPL10 | -0.8024 | 8.546315 | -1.49423 | 0.140891 | 0.860314 | -4.58158 |
| P18077 | RPL35A | 1.712812 | 11.39346 | 1.493084 | 0.141299 | 0.860314 | -4.58425 |
| Q13509 | TUBB3 | -0.81012 | 9.79097 | -1.49188 | 0.141505 | 0.860314 | -4.58165 |
| Q93050 | ATP6V0A1 | -0.73091 | 10.19313 | -1.48839 | 0.142316 | 0.860314 | -4.58176 |
| P01619 | #N/D | 0.88606 | 14.00764 | 1.486499 | 0.143254 | 0.860314 | -4.58186 |
| P01817 | #N/D | 0.912796 | 11.01698 | 1.476802 | 0.145389 | 0.860314 | -4.58213 |
| P51659 | HSD17B4 | -0.74807 | 12.08701 | -1.4753 | 0.14579 | 0.860314 | -4.58218 |
| Q16623-3 | #N/D | 0.755169 | 9.890803 | 1.474992 | 0.145874 | 0.860314 | -4.58219 |
| Q16798 | ME3 | -0.8116 | 8.140939 | -1.47083 | 0.147553 | 0.860314 | -4.58237 |
| P13010 | XRCC5 | -0.79096 | 12.2267 | -1.4667 | 0.148109 | 0.860314 | -4.58245 |
| P50993 | ATP1A2 | 1.255442 | 10.45463 | 1.465642 | 0.148501 | 0.860314 | -4.5825 |
| P61353 | RPL27 | -0.77934 | 9.636353 | -1.45314 | 0.151825 | 0.871918 | -4.58289 |
| P55209 | NAP1L1 | -1.38815 | 12.57393 | -1.44985 | 0.152839 | 0.871918 | -4.583 |

| | | | | | | | |
|---|---|---|---|---|---|---|---|
| P21579 | SYT1 | -1.00288 | 9.733547 | -1.43988 | 0.155527 | 0.871918 | -4.58331 |
| Q92823 | NRCAM | 0.730698 | 13.18446 | 1.434913 | 0.156932 | 0.871918 | -4.58346 |
| Q15392 | DHCR24 | -1.19006 | 8.63106 | -1.4334 | 0.157916 | 0.871918 | -4.58355 |
| Q96KP4 | CNDP2 | -0.71305 | 11.48894 | -1.43063 | 0.15815 | 0.871918 | -4.5836 |
| P02655 | APOC2 | 1.033307 | 11.22537 | 1.430607 | 0.158367 | 0.871918 | -4.58361 |
| P02753 | RBP4 | 1.399391 | 9.614475 | 1.427442 | 0.159498 | 0.871918 | -4.58495 |
| P23515 | OMG | -0.77007 | 10.03831 | -1.4258 | 0.159534 | 0.871918 | -4.58375 |
| P99999 | CYCS | 1.032162 | 7.341751 | 1.435422 | 0.161372 | 0.874054 | -4.58726 |
| P10636-5 | #N/D | 0.632677 | 11.53737 | 1.414215 | 0.162893 | 0.874054 | -4.58411 |
| Q16352 | INA | 0.622007 | 13.06932 | 1.412298 | 0.163454 | 0.874054 | -4.58417 |
| Q14624-3 | #N/D | -1.43014 | 8.418841 | -1.41871 | 0.165244 | 0.874054 | -4.58849 |
| Q9Y5K8 | ATP6V1D | -1.13036 | 7.792041 | -1.41257 | 0.165254 | 0.874054 | -4.58692 |
| Q15257-2 | #N/D | 1.509014 | 9.172995 | 1.399156 | 0.167889 | 0.874054 | -4.58717 |
| P15313 | ATP6V1B1 | 0.954456 | 8.509029 | 1.396723 | 0.168497 | 0.874054 | -4.5858 |
| O00499 | BIN1 | 1.001903 | 8.599564 | 1.397842 | 0.169786 | 0.874054 | -4.58727 |
| O43776 | NARS1 | -0.84764 | 10.11421 | -1.38814 | 0.170651 | 0.874054 | -4.58491 |
| P02747 | C1QC | -1.41701 | 14.27094 | -1.38442 | 0.17178 | 0.874054 | -4.58502 |
| P35908 | KRT2 | 1.484249 | 14.98502 | 1.380796 | 0.172886 | 0.874054 | -4.58513 |
| P01861 | #N/D | -0.8378 | 9.667126 | -1.3813 | 0.173049 | 0.874054 | -4.58621 |
| P23471 | PTPRZ1 | -1.99476 | 10.95353 | -1.37843 | 0.173818 | 0.874054 | -4.58628 |
| A0A075B6I | #N/D | -1.21769 | 12.27284 | -1.36362 | 0.178201 | 0.874054 | -4.58565 |
| P07108 | DBI | -0.90158 | 7.299068 | -1.36675 | 0.178741 | 0.874054 | -4.58748 |
| P61026 | RAB10 | 1.143522 | 8.602059 | 1.363345 | 0.179802 | 0.874054 | -4.58861 |
| Q9BVA1 | TUBB2B | 1.263497 | 10.28254 | 1.353236 | 0.181475 | 0.874054 | -4.58596 |
| Q9BPU6 | DPYSL5 | -0.78328 | 10.87508 | -1.35201 | 0.181866 | 0.874054 | -4.586 |
| P28838 | LAP3 | 0.797584 | 11.12429 | 1.348393 | 0.183018 | 0.874054 | -4.5861 |
| Q92954 | PRG4 | -1.15778 | 10.51058 | -1.34705 | 0.183448 | 0.874054 | -4.58614 |
| O60641 | SNAP91 | 0.750115 | 12.93002 | 1.346952 | 0.183478 | 0.874054 | -4.58615 |
| P23142 | FBLN1 | 0.754584 | 9.472544 | 1.34485 | 0.184576 | 0.874054 | -4.58623 |
| Q13177 | PAK2 | -0.99621 | 8.338763 | -1.34608 | 0.185263 | 0.874054 | -4.5872 |
| P20336 | RAB3A | -1.19258 | 9.798318 | -1.33848 | 0.186304 | 0.874054 | -4.5864 |

| | | | | | | | |
|---|---|---|---|---|---|---|---|
| O94919 | ENDOD1 | 0.884651 | 9.449366 | 1.337789 | 0.186633 | 0.874054 | -4.58643 |
| Q01105 | SET | 0.763608 | 12.74759 | 1.337005 | 0.186684 | 0.874054 | -4.58644 |
| P04843 | RPN1 | 0.738248 | 8.447986 | 1.336738 | 0.187082 | 0.874054 | -4.58647 |
| Q5TFQ8 | SIRPB1 | 1.069064 | 8.396877 | 1.329123 | 0.19145 | 0.882172 | -4.58933 |
| Q99426 | TBCB | 0.937696 | 8.295429 | 1.32713 | 0.191566 | 0.882172 | -4.58842 |
| P01024 | C3 | 0.811994 | 15.31739 | 1.321198 | 0.191865 | 0.882172 | -4.58691 |
| Q92804 | TAF15 | 1.14867 | 9.172634 | 1.315771 | 0.194448 | 0.885341 | -4.58955 |
| Q13449 | LSAMP | 0.744398 | 9.843599 | 1.313008 | 0.194592 | 0.885341 | -4.58714 |
| Q07955 | SRSF1 | -0.80012 | 11.42114 | -1.30562 | 0.197075 | 0.888378 | -4.58736 |
| P01019 | AGT | -1.06528 | 13.11805 | -1.30019 | 0.198917 | 0.888378 | -4.58752 |
| P05388 | RPLP0 | 0.759009 | 11.63871 | 1.2876 | 0.203237 | 0.888378 | -4.58788 |
| P35611 | ADD1 | 1.250903 | 8.364787 | 1.28572 | 0.206657 | 0.888378 | -4.5908 |
| P09960 | LTA4H | 0.747531 | 8.842087 | 1.275342 | 0.208274 | 0.888378 | -4.58825 |
| P61764 | STXBP1 | -0.84544 | 13.40973 | -1.2652 | 0.211093 | 0.888378 | -4.58851 |
| P10620 | MGST1 | -0.97196 | 10.88042 | -1.25853 | 0.213478 | 0.888378 | -4.5887 |
| P62266 | RPS23 | -0.88442 | 9.096047 | -1.25933 | 0.213712 | 0.888378 | -4.58869 |
| O43426 | SYNJ1 | -2.62511 | 13.74991 | -1.25449 | 0.215015 | 0.888378 | -4.58881 |
| P62888 | RPL30 | 0.863748 | 9.168351 | 1.254121 | 0.215159 | 0.888378 | -4.58882 |
| Q9UPY8 | MAPRE3 | 0.589254 | 11.03301 | 1.251711 | 0.215933 | 0.888378 | -4.58889 |
| Q86VP6 | CAND1 | 0.947508 | 9.469724 | 1.250985 | 0.216196 | 0.888378 | -4.58891 |
| Q14203 | DCTN1 | -1.68864 | 7.611697 | -1.25774 | 0.21838 | 0.888378 | -4.59306 |
| P62249 | RPS16 | 0.591315 | 10.28851 | 1.243977 | 0.218744 | 0.888378 | -4.5891 |
| P08865 | RPSA | -0.76574 | 13.49287 | -1.24344 | 0.218939 | 0.888378 | -4.58912 |
| Q6PCE3 | PGM2L1 | -1.01969 | 10.75219 | -1.24268 | 0.219219 | 0.888378 | -4.58914 |
| P07237 | P4HB | -0.98216 | 10.09498 | -1.24163 | 0.219901 | 0.888378 | -4.58918 |
| P22314 | UBA1 | -0.8245 | 12.03395 | -1.23866 | 0.220692 | 0.888378 | -4.58925 |
| P62805 | H4C9 | 0.961467 | 11.74498 | 1.237172 | 0.22124 | 0.888378 | -4.58929 |
| Q01484 | ANK2 | -0.99169 | 9.391498 | -1.23614 | 0.221715 | 0.888378 | -4.58932 |
| Q9UKX2 | MYH2 | 1.350932 | 9.36748 | 1.233936 | 0.222948 | 0.888378 | -4.59078 |
| P30405 | PPIF | -0.90889 | 7.967455 | -1.23845 | 0.223695 | 0.888378 | -4.59159 |
| P49189 | ALDH9A1 | 1.074589 | 8.956597 | 1.229786 | 0.224269 | 0.888378 | -4.5901 |

| | | | | | | | |
|---|---|---|---|---|---|---|---|
| Q9P258 | RCC2 | -0.67347 | 11.42629 | -1.22817 | 0.224573 | 0.888378 | -4.58954 |
| P80748 | #N/D | 0.873903 | 13.59399 | 1.225686 | 0.225501 | 0.888378 | -4.58961 |
| P09471 | GNAO1 | 0.69893 | 11.83785 | 1.224497 | 0.225945 | 0.888378 | -4.58964 |
| Q86V81 | ALYREF | -1.25151 | 8.628483 | -1.22585 | 0.22595 | 0.888378 | -4.59068 |
| Q9UQ80 | PA2G4 | -0.82604 | 9.950951 | -1.22419 | 0.226061 | 0.888378 | -4.58965 |
| P63010 | AP2B1 | 0.907665 | 10.0117 | 1.219715 | 0.227739 | 0.888378 | -4.58977 |
| P10515 | DLAT | 0.785415 | 8.7176 | 1.217103 | 0.228915 | 0.888378 | -4.58984 |
| Q15631 | TSN | -1.02144 | 8.819731 | -1.21557 | 0.229597 | 0.888378 | -4.58989 |
| Q9UNQ0 | ABCG2 | 1.579538 | 11.29824 | 1.215023 | 0.229908 | 0.888378 | -4.59116 |
| O14594 | NCAN | -0.69108 | 9.186149 | -1.21068 | 0.231662 | 0.888378 | -4.59003 |
| P24752 | ACAT1 | -0.84108 | 10.23426 | -1.20905 | 0.231776 | 0.888378 | -4.59006 |
| P02675 | FGB | 0.604427 | 14.18233 | 1.203446 | 0.23392 | 0.888378 | -4.59021 |
| P51674 | GPM6A | -0.95222 | 9.977344 | -1.20234 | 0.234344 | 0.888378 | -4.59024 |
| Q5IFJ7 | #N/D | -1.46679 | 10.61788 | -1.20169 | 0.234687 | 0.888378 | -4.59026 |
| P11021 | HSPA5 | -0.60587 | 14.82701 | -1.19835 | 0.235883 | 0.888378 | -4.59035 |
| P35998 | PSMC2 | 1.12322 | 10.28438 | 1.196145 | 0.237023 | 0.888378 | -4.59041 |
| O43175 | PHGDH | -0.53734 | 13.11601 | -1.19451 | 0.237365 | 0.888378 | -4.59045 |
| P30041 | PRDX6 | 1.125602 | 12.92684 | 1.188913 | 0.239544 | 0.888378 | -4.5906 |
| P35613-2 | #N/D | -0.62964 | 10.74277 | -1.1867 | 0.240407 | 0.888378 | -4.59066 |
| P29762 | CRABP1 | -1.22879 | 11.46855 | -1.18637 | 0.240536 | 0.888378 | -4.59066 |
| Q8WUM4 | PDCD6IP | 1.906781 | 7.545402 | 1.199639 | 0.242278 | 0.888378 | -4.59415 |
| P40925 | MDH1 | -0.65113 | 13.09244 | -1.17928 | 0.243323 | 0.888378 | -4.59085 |
| P22061 | PCMT1 | -0.67721 | 10.41461 | -1.17858 | 0.2436 | 0.888378 | -4.59087 |
| P35637 | FUS | -0.78829 | 9.460308 | -1.17738 | 0.244169 | 0.888378 | -4.5909 |
| P62913 | RPL11 | 0.818898 | 12.51404 | 1.173239 | 0.245718 | 0.888378 | -4.59101 |
| P60981 | DSTN | -0.83049 | 11.21887 | -1.16927 | 0.247392 | 0.888378 | -4.59112 |
| P01876 | #N/D | -0.895 | 15.78121 | -1.16715 | 0.248147 | 0.888378 | -4.59117 |
| P21333 | FLNA | -0.69187 | 13.07842 | -1.16688 | 0.248257 | 0.888378 | -4.59118 |
| P78371 | CCT2 | 0.877888 | 10.10086 | 1.163505 | 0.24961 | 0.888378 | -4.59127 |
| P13716-2 | #N/D | 0.878733 | 8.80277 | 1.163682 | 0.249925 | 0.888378 | -4.59127 |
| P52292 | KPNA2 | 0.908174 | 7.997937 | 1.164099 | 0.250463 | 0.888378 | -4.59243 |

| | | | | | | | |
|---|---|---|---|---|---|---|---|
| O00187 | MASP2 | -1.00078 | 13.7813 | -1.1528 | 0.254033 | 0.894042 | -4.59155 |
| P01042-2 | #N/D | 1.110769 | 11.83736 | 1.151103 | 0.254817 | 0.894042 | -4.59159 |
| P07900-2 | #N/D | -0.81675 | 14.11273 | -1.14984 | 0.255147 | 0.894042 | -4.59162 |
| P42704 | LRPPRC | -1.05281 | 13.36178 | -1.1421 | 0.258324 | 0.898144 | -4.59182 |
| P22792 | CPN2 | 0.932263 | 10.29617 | 1.141952 | 0.258384 | 0.898144 | -4.59182 |
| O95336 | PGLS | -0.91364 | 10.2894 | -1.13801 | 0.260013 | 0.900206 | -4.59192 |
| Q12931 | TRAP1 | -1.00013 | 11.45079 | -1.13153 | 0.262704 | 0.903543 | -4.59209 |
| P30040 | ERP29 | -1.27386 | 8.785068 | -1.13391 | 0.263057 | 0.903543 | -4.59278 |
| P49207 | RPL34 | -0.88816 | 9.206154 | -1.12704 | 0.264672 | 0.90551 | -4.5922 |
| P09417 | QDPR | 0.819545 | 11.79198 | 1.124036 | 0.265843 | 0.905953 | -4.59228 |
| Q08722 | CD47 | 0.68866 | 9.746025 | 1.12032 | 0.267409 | 0.907729 | -4.59237 |
| P52565 | ARHGDIA | -0.99694 | 10.94718 | -1.11284 | 0.27067 | 0.910112 | -4.59256 |
| P30050 | RPL12 | 0.951541 | 10.32925 | 1.110113 | 0.271744 | 0.910112 | -4.59263 |
| P00736 | C1R | -0.86883 | 13.74472 | -1.10954 | 0.271991 | 0.910112 | -4.59264 |
| Q9UMS4 | PRPF19 | 0.914766 | 9.289913 | 1.104918 | 0.273969 | 0.910112 | -4.59276 |
| Q14194 | CRMP1 | -0.56374 | 11.68612 | -1.10474 | 0.274045 | 0.910112 | -4.59276 |
| Q04837 | SSBP1 | 0.943062 | 9.844274 | 1.104131 | 0.274395 | 0.910112 | -4.59278 |
| P62987 | UBA52 | -0.42545 | 11.77756 | -1.10116 | 0.275588 | 0.910594 | -4.59285 |
| P22102 | GART | -0.98511 | 10.28125 | -1.09609 | 0.277958 | 0.914944 | -4.59298 |
| P01008 | SERPINC1 | 0.564679 | 14.22003 | 1.093207 | 0.279033 | 0.915016 | -4.59305 |
| P02656 | APOC3 | -0.82817 | 12.92227 | -1.08243 | 0.283751 | 0.921537 | -4.59331 |
| P07197 | NEFM | 0.459386 | 12.44543 | 1.082193 | 0.283854 | 0.921537 | -4.59332 |
| A0A0C4DH | #N/D | -0.95536 | 14.37611 | -1.08001 | 0.284817 | 0.921537 | -4.59337 |
| P06681 | C2 | -0.64917 | 10.88017 | -1.07323 | 0.288083 | 0.921537 | -4.59354 |
| Q7L0J3 | SV2A | 1.086366 | 9.396699 | 1.074196 | 0.289107 | 0.921537 | -4.59389 |
| P13637 | ATP1A3 | -0.83258 | 12.48297 | -1.06942 | 0.289517 | 0.921537 | -4.59363 |
| Q16851 | UGP2 | -0.42077 | 12.72477 | -1.06657 | 0.290792 | 0.921537 | -4.5937 |
| Q14697-2 | #N/D | -0.69196 | 9.735768 | -1.06676 | 0.290968 | 0.921537 | -4.59369 |
| Q01518 | CAP1 | -0.75737 | 11.32087 | -1.06437 | 0.291779 | 0.921537 | -4.59375 |
| P31943 | HNRNPH1 | -0.66376 | 12.25814 | -1.06356 | 0.29214 | 0.921537 | -4.59377 |
| P02654 | APOC1 | 0.68446 | 11.84732 | 1.061484 | 0.293159 | 0.921537 | -4.59382 |

| | | | | | | | |
|---|---|---|---|---|---|---|---|
| P08708 | RPS17 | -0.76199 | 7.699005 | -1.06171 | 0.293746 | 0.921537 | -4.59419 |
| Q9NQC3 | RTN4 | -0.6249 | 10.95603 | -1.05714 | 0.295034 | 0.922248 | -4.59392 |
| Q99497 | PARK7 | 0.83966 | 9.50833 | 1.055544 | 0.296111 | 0.922297 | -4.59396 |
| Q15334 | LLGL1 | -0.73525 | 8.598936 | -1.05193 | 0.297399 | 0.922999 | -4.59405 |
| P05109 | S100A8 | -0.86081 | 9.04775 | -1.04744 | 0.299704 | 0.926843 | -4.59415 |
| A2NJV5 | #N/D | 1.452229 | 8.920818 | 1.043388 | 0.301951 | 0.927376 | -4.59433 |
| O00154 | ACOT7 | -0.85974 | 10.65194 | -1.03266 | 0.306247 | 0.927376 | -4.5945 |
| P20671 | H2AC7 | -0.79714 | 8.62645 | -1.0318 | 0.307881 | 0.927376 | -4.59462 |
| A0A087WS | #N/D | 0.93303 | 13.2827 | 1.027835 | 0.308493 | 0.927376 | -4.59461 |
| A0A075B6I | #N/D | -0.99999 | 8.760973 | -1.02815 | 0.308788 | 0.927376 | -4.59461 |
| P00338 | LDHA | -0.53766 | 12.44144 | -1.02634 | 0.309191 | 0.927376 | -4.59465 |
| P05026 | ATP1B1 | -0.71858 | 9.719583 | -1.02584 | 0.309862 | 0.927376 | -4.59466 |
| A0A0C4DH | #N/D | 0.981911 | 10.49513 | 1.024677 | 0.310313 | 0.927376 | -4.59472 |
| P27348 | YWHAQ | -0.79585 | 11.63346 | -1.02007 | 0.31221 | 0.927376 | -4.59479 |
| Q5JNZ5 | #N/D | 0.793107 | 10.43992 | 1.018415 | 0.312907 | 0.927376 | -4.59483 |
| P02686 | MBP | -0.42148 | 12.52744 | -1.01709 | 0.313533 | 0.927376 | -4.59486 |
| P29972 | AQP1 | 0.796463 | 8.612235 | 1.013946 | 0.315548 | 0.927376 | -4.59494 |
| Q04637 | EIF4G1 | -0.80353 | 9.520493 | -1.01301 | 0.31571 | 0.927376 | -4.59496 |
| P50395 | GDI2 | 0.538241 | 11.20433 | 1.012402 | 0.315746 | 0.927376 | -4.59497 |
| P68366 | TUBA4A | 0.53696 | 8.182302 | 1.013449 | 0.315884 | 0.927376 | -4.59495 |
| P11279 | LAMP1 | -0.64955 | 9.296891 | -1.00999 | 0.317322 | 0.928461 | -4.59503 |
| P60866 | RPS20 | -0.69447 | 8.421034 | -1.0032 | 0.320557 | 0.93478 | -4.59518 |
| Q96FW1 | OTUB1 | -0.59603 | 7.493366 | -0.99968 | 0.323721 | 0.940848 | -4.59526 |
| P00740 | F9 | 0.572901 | 11.14519 | 0.987236 | 0.327818 | 0.947718 | -4.59554 |
| P20042 | EIF2S2 | 0.94774 | 8.98351 | 0.983836 | 0.329551 | 0.947718 | -4.59562 |
| Q9P2U7 | SLC17A7 | -0.97317 | 10.35073 | -0.98243 | 0.330318 | 0.947718 | -4.59558 |
| P14324 | FDPS | 0.663002 | 8.575581 | 0.983528 | 0.330447 | 0.947718 | -4.59544 |
| P00734 | F2 | 0.604491 | 14.7427 | 0.977104 | 0.332764 | 0.95101 | -4.59577 |
| P05154 | SERPINA5 | 0.564152 | 11.45435 | 0.975028 | 0.333784 | 0.95101 | -4.59581 |
| Q9H4G4 | GLIPR2 | -0.66353 | 9.978347 | -0.95994 | 0.34133 | 0.965156 | -4.59614 |
| Q13151 | HNRNPA0 | 0.662336 | 9.990869 | 0.958441 | 0.342241 | 0.965156 | -4.59618 |

| | | | | | | | |
|---|---|---|---|---|---|---|---|
| P49753 | ACOT2 | 0.656012 | 10.06568 | 0.955405 | 0.343523 | 0.965156 | -4.59624 |
| P00558 | PGK1 | 0.414157 | 12.86953 | 0.954334 | 0.344059 | 0.965156 | -4.59627 |
| P60842 | EIF4A1 | -0.8732 | 11.09096 | -0.95385 | 0.344302 | 0.965156 | -4.59628 |
| P05155 | SERPING1 | 0.617788 | 12.77219 | 0.950947 | 0.345761 | 0.965945 | -4.59634 |
| P28072 | PSMB6 | -0.63077 | 9.15565 | -0.94308 | 0.349732 | 0.965945 | -4.59651 |
| P27918 | CFP | 0.628805 | 11.216 | 0.938597 | 0.352011 | 0.965945 | -4.59661 |
| P07196 | NEFL | 0.360605 | 12.62642 | 0.938276 | 0.352174 | 0.965945 | -4.59661 |
| P08559-4 | #N/D | -0.63386 | 10.42252 | -0.93821 | 0.35221 | 0.965945 | -4.59661 |
| P08238 | HSP90AB1 | -0.56817 | 13.56843 | -0.92587 | 0.358527 | 0.965945 | -4.59688 |
| P34932 | HSPA4 | -0.50034 | 12.19409 | -0.92504 | 0.358956 | 0.965945 | -4.59689 |
| P02787 | TF | -0.51361 | 14.24336 | -0.92376 | 0.359616 | 0.965945 | -4.59692 |
| P37837 | TALDO1 | 0.479462 | 13.06675 | 0.923058 | 0.359979 | 0.965945 | -4.59694 |
| P02745 | C1QA | 0.875308 | 10.24676 | 0.921481 | 0.361103 | 0.965945 | -4.59675 |
| P04040 | CAT | -0.42656 | 8.854698 | -0.9221 | 0.36135 | 0.965945 | -4.59674 |
| P10643 | C7 | -0.45297 | 12.83675 | -0.91803 | 0.362581 | 0.965945 | -4.59704 |
| P09871 | C1S | -0.5084 | 13.55189 | -0.91579 | 0.363746 | 0.965945 | -4.59709 |
| P36542 | ATP5F1C | 0.831712 | 8.889386 | 0.915078 | 0.364188 | 0.965945 | -4.5971 |
| P01860 | #N/D | -0.74071 | 14.17562 | -0.91118 | 0.36615 | 0.965945 | -4.59718 |
| Q71U36 | TUBA1A | 0.937965 | 13.10907 | 0.915162 | 0.367306 | 0.965945 | -4.59644 |
| Q8NC51 | SERBP1 | 1.077696 | 11.27405 | 0.9084 | 0.367601 | 0.965945 | -4.59724 |
| P31948 | STIP1 | 0.42719 | 10.97486 | 0.907831 | 0.367899 | 0.965945 | -4.59725 |
| Q9Y617 | PSAT1 | 0.577773 | 11.24982 | 0.902075 | 0.370922 | 0.965945 | -4.59737 |
| P00492 | HPRT1 | 0.798045 | 10.76362 | 0.899125 | 0.372548 | 0.965945 | -4.59743 |
| Q9HC38 | GLOD4 | 0.660573 | 9.301142 | 0.898289 | 0.372918 | 0.965945 | -4.59745 |
| Q7L099-4 | #N/D | 0.548507 | 10.43573 | 0.89352 | 0.375443 | 0.965945 | -4.59755 |
| P84077 | ARF1 | -0.64386 | 9.006425 | -0.88568 | 0.379689 | 0.965945 | -4.59771 |
| O76070 | SNCG | -0.46924 | 10.89826 | -0.88545 | 0.379738 | 0.965945 | -4.59771 |
| Q02878 | RPL6 | -0.75941 | 11.19865 | -0.88322 | 0.380931 | 0.965945 | -4.59776 |
| P06312 | #N/D | -0.64817 | 13.06851 | -0.88291 | 0.381102 | 0.965945 | -4.59776 |
| P21281 | ATP6V1B2 | -0.60939 | 13.63954 | -0.8806 | 0.382337 | 0.965945 | -4.59781 |
| Q99747 | NAPG | 0.449691 | 8.259074 | 0.880544 | 0.382437 | 0.965945 | -4.59781 |

| | | | | | | | |
|---|---|---|---|---|---|---|---|
| P20742 | #N/D | -0.62256 | 13.66452 | -0.87994 | 0.382695 | 0.965945 | -4.59782 |
| Q00610 | CLTC | 0.70197 | 13.78944 | 0.875816 | 0.384911 | 0.965945 | -4.59791 |
| Q9H3S7 | PTPN23 | -0.4923 | 9.197781 | -0.87375 | 0.386025 | 0.965945 | -4.59795 |
| Q08211 | DHX9 | -0.65208 | 13.43428 | -0.87209 | 0.386925 | 0.965945 | -4.59798 |
| Q96AX9-5 | #N/D | -0.7749 | 9.366756 | -0.87125 | 0.387443 | 0.965945 | -4.598 |
| Q14894 | CRYM | 0.533858 | 12.35016 | 0.870136 | 0.38798 | 0.965945 | -4.59802 |
| Q9Y266 | NUDC | 0.668328 | 9.322208 | 0.870615 | 0.388174 | 0.965945 | -4.598 |
| P04264 | KRT1 | 0.558023 | 16.88435 | 0.86972 | 0.388205 | 0.965945 | -4.59803 |
| O76054 | SEC14L2 | -0.69396 | 8.396433 | -0.86802 | 0.390157 | 0.965945 | -4.59712 |
| P00441 | SOD1 | -0.58361 | 11.21279 | -0.86092 | 0.393059 | 0.965945 | -4.5982 |
| P07954 | FH | 0.694824 | 11.84092 | 0.860383 | 0.393353 | 0.965945 | -4.59821 |
| Q16799 | RTN1 | 0.602158 | 10.36556 | 0.855502 | 0.395957 | 0.965945 | -4.59831 |
| P49588 | AARS1 | 1.27202 | 10.01352 | 0.852892 | 0.397753 | 0.965945 | -4.59754 |
| P28074 | PSMB5 | -0.6109 | 8.249193 | -0.85347 | 0.39788 | 0.965945 | -4.59769 |
| P30153 | PPP2R1A | 0.664348 | 10.68953 | 0.847176 | 0.400541 | 0.965945 | -4.59847 |
| O75340 | PDCD6 | -0.80116 | 7.338176 | -0.85277 | 0.400678 | 0.965945 | -4.59664 |
| P29401 | TKT | 0.39205 | 12.31095 | 0.845717 | 0.401348 | 0.965945 | -4.5985 |
| P05156 | CFI | 0.575013 | 13.67799 | 0.841787 | 0.403526 | 0.965945 | -4.59857 |
| P04406 | GAPDH | 0.578606 | 14.71391 | 0.840763 | 0.404094 | 0.965945 | -4.59859 |
| P01700 | #N/D | 0.857507 | 10.59788 | 0.837469 | 0.405993 | 0.965945 | -4.59866 |
| P78417 | GSTO1 | 0.646741 | 11.81208 | 0.835747 | 0.406887 | 0.965945 | -4.59869 |
| A0A0B4J1Y | #N/D | 0.622483 | 9.589963 | 0.835785 | 0.407382 | 0.965945 | -4.59828 |
| Q5U7I5 | #N/D | 0.969689 | 12.18705 | 0.833283 | 0.408328 | 0.965945 | -4.59874 |
| P02647 | APOA1 | 0.599755 | 15.5712 | 0.829775 | 0.410226 | 0.965945 | -4.5988 |
| P02774-3 | #N/D | -0.65241 | 14.29976 | -0.82922 | 0.41054 | 0.965945 | -4.59882 |
| Q99878 | H2AC14 | -0.7574 | 8.686371 | -0.82605 | 0.412999 | 0.965945 | -4.59792 |
| Q05193 | DNM1 | 0.452225 | 10.1149 | 0.823833 | 0.413566 | 0.965945 | -4.59892 |
| P15169 | CPN1 | -0.6382 | 9.464541 | -0.82207 | 0.414762 | 0.965945 | -4.59895 |
| P62861 | #N/D | -0.56266 | 9.145956 | -0.81934 | 0.416376 | 0.965945 | -4.599 |
| P68133 | ACTA1 | 0.508121 | 8.536068 | 0.820056 | 0.416769 | 0.965945 | -4.59775 |
| Q16143 | SNCB | -0.49442 | 11.41947 | -0.81746 | 0.417168 | 0.965945 | -4.59904 |

| | | | | | | | |
|---|---|---|---|---|---|---|---|
| Q9BY11 | PACSIN1 | 0.735654 | 11.03934 | 0.817111 | 0.417364 | 0.965945 | -4.59904 |
| P52272 | HNRNPM | -0.46282 | 12.38158 | -0.81628 | 0.417838 | 0.965945 | -4.59906 |
| Q15233 | NONO | 0.539126 | 9.777373 | 0.815799 | 0.418108 | 0.965945 | -4.59907 |
| P01706 | #N/D | -0.48663 | 8.121541 | -0.81691 | 0.418156 | 0.965945 | -4.5986 |
| P00568 | AK1 | 0.540949 | 11.14475 | 0.81066 | 0.421029 | 0.965945 | -4.59916 |
| P62851 | RPS25 | -0.58472 | 10.64803 | -0.80884 | 0.422069 | 0.965945 | -4.5992 |
| P20916 | MAG | 0.586857 | 9.172305 | 0.806672 | 0.423721 | 0.965945 | -4.59877 |
| Q15121 | PEA15 | 0.391537 | 10.46576 | 0.805816 | 0.423794 | 0.965945 | -4.59925 |
| P36871 | PGM1 | -0.45714 | 11.50669 | -0.80468 | 0.424444 | 0.965945 | -4.59928 |
| P15880 | RPS2 | 0.734988 | 12.70417 | 0.803659 | 0.425028 | 0.965945 | -4.59929 |
| P29966 | MARCKS | 0.643319 | 12.49708 | 0.80235 | 0.425778 | 0.965945 | -4.59932 |
| A0A0B4J1X | #N/D | -1.01121 | 10.0226 | -0.80118 | 0.426513 | 0.965945 | -4.59934 |
| P02511 | CRYAB | 0.380928 | 10.8827 | 0.800433 | 0.426878 | 0.965945 | -4.59935 |
| P24539 | ATP5PB | -0.9244 | 9.276826 | -0.79749 | 0.429141 | 0.965945 | -4.59756 |
| P27695 | APEX1 | -0.96575 | 9.067064 | -0.79724 | 0.42986 | 0.965945 | -4.59804 |
| P02790 | HPX | -0.63063 | 14.96138 | -0.79467 | 0.430195 | 0.965945 | -4.59946 |
| O75947 | ATP5PD | -0.62995 | 8.722357 | -0.79153 | 0.432133 | 0.965945 | -4.59951 |
| P28161 | GSTM2 | -0.55565 | 8.502551 | -0.79027 | 0.433572 | 0.965945 | -4.59864 |
| P28070 | PSMB4 | 0.572557 | 9.16426 | 0.788989 | 0.433607 | 0.965945 | -4.59906 |
| P38117 | ETFB | -0.82047 | 8.695554 | -0.78697 | 0.435133 | 0.965945 | -4.5978 |
| O43761 | SYNGR3 | -0.74646 | 10.57735 | -0.78614 | 0.435137 | 0.965945 | -4.59961 |
| P00403 | COX2 | 0.473792 | 8.386699 | 0.78595 | 0.43607 | 0.965945 | -4.5991 |
| P60900 | PSMA6 | -0.77232 | 10.92948 | -0.7826 | 0.437194 | 0.965945 | -4.59968 |
| P46459 | NSF | 0.567939 | 12.65607 | 0.779383 | 0.43907 | 0.965945 | -4.59974 |
| P14866 | HNRNPL | -0.4792 | 12.72325 | -0.77758 | 0.440125 | 0.965945 | -4.59977 |
| P16403 | H1-2 | 0.331857 | 11.57771 | 0.773287 | 0.442639 | 0.965945 | -4.59984 |
| P23246 | SFPQ | 0.463855 | 12.17656 | 0.772184 | 0.443286 | 0.965945 | -4.59986 |
| Q07020 | RPL18 | -0.73394 | 11.58711 | -0.76742 | 0.446088 | 0.965945 | -4.59995 |
| Q13885 | TUBB2A | -0.54972 | 12.3616 | -0.75982 | 0.45058 | 0.965945 | -4.60008 |
| P06331 | #N/D | 0.524666 | 11.07987 | 0.758529 | 0.451349 | 0.965945 | -4.6001 |
| Q13363 | CTBP1 | 0.578714 | 9.960849 | 0.757146 | 0.45217 | 0.965945 | -4.60013 |

| | | | | | | | |
|---|---|---|---|---|---|---|---|
| P02679-2 | #N/D | 0.355341 | 11.17822 | 0.756386 | 0.452622 | 0.965945 | -4.60014 |
| P13667 | PDIA4 | 0.520464 | 11.88336 | 0.755884 | 0.45292 | 0.965945 | -4.60015 |
| P30038 | ALDH4A1 | -0.39117 | 11.58862 | -0.75586 | 0.452935 | 0.965945 | -4.60015 |
| P78559 | MAP1A | -0.5001 | 10.07764 | -0.75555 | 0.453303 | 0.965945 | -4.60015 |
| A0A0A0MT | #N/D | 0.845825 | 9.911469 | 0.751992 | 0.455936 | 0.965945 | -4.60021 |
| A0A075B6I | #N/D | 1.250135 | 9.154402 | 0.750562 | 0.456788 | 0.965945 | -4.59675 |
| P14625 | HSP90B1 | 0.754295 | 13.43176 | 0.749265 | 0.456866 | 0.965945 | -4.60027 |
| P07741 | APRT | -0.92804 | 8.592073 | -0.74733 | 0.458477 | 0.965945 | -4.599 |
| P62873 | GNB1 | -0.46038 | 12.21737 | -0.74408 | 0.459971 | 0.965945 | -4.60035 |
| O75636 | FCN3 | 0.38449 | 11.6985 | 0.742566 | 0.460881 | 0.965945 | -4.60038 |
| P62263 | RPS14 | -0.58093 | 12.25025 | -0.73963 | 0.462648 | 0.965945 | -4.60043 |
| P40227 | CCT6A | -0.39734 | 12.26876 | -0.73809 | 0.463571 | 0.965945 | -4.60046 |
| Q8N163 | CCAR2 | -0.52417 | 8.405871 | -0.73425 | 0.466071 | 0.965945 | -4.60052 |
| P08758 | ANXA5 | -0.46711 | 12.52081 | -0.73234 | 0.467048 | 0.965945 | -4.60055 |
| P06576 | ATP5F1B | 0.512466 | 11.92408 | 0.731056 | 0.467825 | 0.965945 | -4.60058 |
| P06730-2 | #N/D | 0.66459 | 8.849688 | 0.730897 | 0.468439 | 0.965945 | -4.59948 |
| Q15485 | FCN2 | -0.72294 | 12.63067 | -0.72757 | 0.469942 | 0.965945 | -4.60063 |
| P61020 | RAB5B | 0.431334 | 8.987505 | 0.727389 | 0.470287 | 0.965945 | -4.60002 |
| P62269 | RPS18 | -0.33307 | 10.80738 | -0.72137 | 0.473771 | 0.965945 | -4.60074 |
| P35858 | IGFALS | -0.4956 | 11.34869 | -0.71935 | 0.474949 | 0.965945 | -4.60077 |
| P80404 | ABAT | -0.38792 | 7.872349 | -0.71995 | 0.4756 | 0.965945 | -4.59898 |
| P12270 | TPR | 0.619202 | 9.31589 | 0.714799 | 0.477966 | 0.965945 | -4.60084 |
| Q99623 | PHB2 | -0.6655 | 10.71072 | -0.71348 | 0.478544 | 0.965945 | -4.60087 |
| A0A0C4DH | #N/D | 0.575075 | 9.100349 | 0.71244 | 0.479236 | 0.965945 | -4.60089 |
| P52597 | HNRNPF | 0.384778 | 9.211955 | 0.711467 | 0.479834 | 0.965945 | -4.6009 |
| P00367 | GLUD1 | -0.37499 | 13.41429 | -0.70845 | 0.481636 | 0.965945 | -4.60095 |
| P02763 | ORM1 | -0.51951 | 11.84132 | -0.70433 | 0.484179 | 0.965945 | -4.60102 |
| P0DOX8 | #N/D | 0.798546 | 13.34101 | 0.693533 | 0.490925 | 0.965945 | -4.60119 |
| P00738 | HP | -0.35211 | 16.23776 | -0.69204 | 0.491805 | 0.965945 | -4.60122 |
| P02748 | C9 | 0.556701 | 13.54131 | 0.682518 | 0.497755 | 0.965945 | -4.60137 |
| P62277 | RPS13 | -0.41287 | 10.17611 | -0.68206 | 0.498044 | 0.965945 | -4.60138 |

| | | | | | | | |
|---|---|---|---|---|---|---|---|
| P46781 | RPS9 | -0.41295 | 10.5113 | -0.68155 | 0.49836 | 0.965945 | -4.60138 |
| Q96AE4 | FUBP1 | 0.525788 | 9.61624 | 0.680319 | 0.499135 | 0.965945 | -4.6014 |
| P62829 | RPL23 | 0.661877 | 7.660806 | 0.682132 | 0.499248 | 0.965945 | -4.5998 |
| P30044 | PRDX5 | -0.79923 | 11.42963 | -0.67826 | 0.500431 | 0.965945 | -4.60144 |
| Q9UI12 | ATP6V1H | 0.665007 | 8.594544 | 0.675884 | 0.50203 | 0.965945 | -4.60077 |
| A0A0C4DH | #N/D | -0.63378 | 14.43238 | -0.67477 | 0.502733 | 0.965945 | -4.60022 |
| P46778 | RPL21 | -1.21615 | 9.748979 | -0.67476 | 0.502904 | 0.965945 | -4.60149 |
| P02750 | LRG1 | -0.56585 | 10.4896 | -0.67389 | 0.503185 | 0.965945 | -4.6015 |
| P60028 | #N/D | -0.46118 | 9.154595 | -0.67191 | 0.50459 | 0.965945 | -4.60153 |
| Q99536 | VAT1 | 0.394659 | 11.20367 | 0.671621 | 0.504615 | 0.965945 | -4.60154 |
| P37840 | SNCA | -0.63566 | 9.699817 | -0.66969 | 0.50594 | 0.965945 | -4.59996 |
| P62318 | SNRPD3 | -0.4184 | 9.893724 | -0.66894 | 0.506309 | 0.965945 | -4.60158 |
| P50990 | CCT8 | 0.320305 | 13.71428 | 0.668355 | 0.506682 | 0.965945 | -4.60159 |
| P62750 | RPL23A | -0.41176 | 11.64551 | -0.66802 | 0.506895 | 0.965945 | -4.60159 |
| O75475 | PSIP1 | 0.515459 | 10.76389 | 0.665899 | 0.508238 | 0.965945 | -4.60163 |
| P06753-2 | #N/D | -0.58875 | 11.55725 | -0.66422 | 0.509574 | 0.965945 | -4.60092 |
| P17096 | HMGA1 | 0.655647 | 10.9518 | 0.663759 | 0.509597 | 0.965945 | -4.60166 |
| P00918 | CA2 | -0.40327 | 9.133649 | -0.66291 | 0.510238 | 0.965945 | -4.60167 |
| Q16653-3 | #N/D | 0.458061 | 9.748252 | 0.662649 | 0.510303 | 0.965945 | -4.60168 |
| P00751 | CFB | 0.394357 | 13.86732 | 0.661977 | 0.51073 | 0.965945 | -4.60169 |
| Q8IXJ6 | SIRT2 | 0.648546 | 11.31293 | 0.660326 | 0.511936 | 0.965945 | -4.60171 |
| P48735 | IDH2 | 0.436628 | 11.01398 | 0.659185 | 0.512507 | 0.965945 | -4.60173 |
| P62280 | RPS11 | -0.40565 | 11.12395 | -0.65526 | 0.515013 | 0.965945 | -4.60179 |
| Q9Y2J8 | PADI2 | 0.586597 | 10.26827 | 0.654577 | 0.515448 | 0.965945 | -4.6018 |
| P00742 | F10 | 0.545819 | 12.22352 | 0.650579 | 0.518056 | 0.965945 | -4.60186 |
| P30048 | PRDX3 | 0.58813 | 9.178145 | 0.651629 | 0.518329 | 0.965945 | -4.59972 |
| P51148 | RAB5C | -0.43005 | 8.07766 | -0.64868 | 0.520044 | 0.965945 | -4.60019 |
| P22626 | HNRNPA2B1 | 0.29892 | 12.6985 | 0.645651 | 0.52117 | 0.965945 | -4.60193 |
| A0A0C4DH | #N/D | -0.47095 | 9.696796 | -0.64458 | 0.52186 | 0.965945 | -4.60195 |
| P0DJI9 | SAA2 | 0.560738 | 8.735337 | 0.637993 | 0.526547 | 0.965945 | -4.60127 |
| P08670 | VIM | 0.253068 | 13.17483 | 0.636766 | 0.526899 | 0.965945 | -4.60207 |

| | | | | | | |
|---|---|---|---|---|---|---|
| P45974 | USP5 | 0.367749 | 9.969402 | 0.635866 | 0.527481 | 0.965945 | -4.60208 |
| P13798 | APEH | -0.45364 | 11.77021 | -0.63528 | 0.52786 | 0.965945 | -4.60209 |
| O43491-4 | #N/D | 0.423953 | 9.677072 | 0.628222 | 0.532439 | 0.965945 | -4.60219 |
| P68871 | HBB | -0.51853 | 14.57697 | -0.627 | 0.533236 | 0.965945 | -4.60221 |
| P26641 | EEF1G | -0.35375 | 10.13888 | -0.62586 | 0.534023 | 0.965945 | -4.60223 |
| P48047 | ATP5PO | 0.664666 | 12.76757 | 0.624532 | 0.534841 | 0.965945 | -4.60225 |
| P20073 | ANXA7 | -0.34643 | 8.552518 | -0.62324 | 0.535727 | 0.965945 | -4.60226 |
| A0A075B6I | #N/D | 0.402977 | 9.618077 | 0.622691 | 0.536042 | 0.965945 | -4.60227 |
| Q8B6J5 | #N/D | 0.63889 | 11.86572 | 0.623546 | 0.536045 | 0.965945 | -4.60225 |
| P06703 | S100A6 | -0.71481 | 9.228704 | -0.62226 | 0.536322 | 0.965945 | -4.60228 |
| P68371 | TUBB4B | -0.62805 | 11.99643 | -0.62214 | 0.536401 | 0.965945 | -4.60228 |
| Q9Y6R7 | FCGBP | -0.32195 | 13.98588 | -0.62096 | 0.537174 | 0.965945 | -4.6023 |
| Q96FC7 | PHYHIPL | 0.617295 | 10.07371 | 0.612904 | 0.542493 | 0.965945 | -4.60241 |
| O75781 | PALM | 0.351511 | 8.620566 | 0.612472 | 0.542824 | 0.965945 | -4.60242 |
| P31946 | YWHAB | -0.39864 | 9.674481 | -0.61249 | 0.54286 | 0.965945 | -4.60242 |
| P04217 | A1BG | 0.346179 | 14.14437 | 0.610992 | 0.543703 | 0.965945 | -4.60244 |
| Q8N573 | OXR1 | 0.847211 | 8.278275 | 0.61246 | 0.543938 | 0.965945 | -4.59707 |
| P07437 | TUBB | 0.483411 | 12.82403 | 0.609988 | 0.544363 | 0.965945 | -4.60245 |
| P02743 | APCS | -0.85509 | 14.22881 | -0.60848 | 0.545354 | 0.965945 | -4.60247 |
| P49327 | FASN | 0.327452 | 12.45025 | 0.60793 | 0.545718 | 0.965945 | -4.60248 |
| P06396 | GSN | 0.381279 | 14.7431 | 0.605757 | 0.547149 | 0.965945 | -4.60251 |
| Q9UN36 | NDRG2 | 0.443517 | 11.61186 | 0.604419 | 0.548032 | 0.965945 | -4.60253 |
| P07384 | CAPN1 | -0.45274 | 8.309043 | -0.60442 | 0.548122 | 0.965945 | -4.60253 |
| P30101 | PDIA3 | -0.44595 | 12.87952 | -0.60402 | 0.548296 | 0.965945 | -4.60254 |
| P31939 | ATIC | -0.4382 | 12.54946 | -0.60216 | 0.549524 | 0.965945 | -4.60256 |
| P53396 | ACLY | -0.50021 | 8.061162 | -0.60182 | 0.549941 | 0.965945 | -4.60174 |
| O95782 | AP2A1 | -0.45255 | 8.500907 | -0.60216 | 0.550201 | 0.965945 | -4.60107 |
| P69891 | HBG1 | -0.47652 | 10.30411 | -0.59751 | 0.552648 | 0.965945 | -4.60263 |
| P04196 | HRG | -0.62192 | 14.14042 | -0.59719 | 0.552814 | 0.965945 | -4.60263 |
| P26038 | MSN | 0.472011 | 8.378736 | 0.596351 | 0.553719 | 0.965945 | -4.60181 |
| Q8WVE0 | EEF1AKMT1 | 0.802977 | 12.59871 | 0.598083 | 0.555292 | 0.965945 | -4.5987 |

| | | | | | | | |
|---|---|---|---|---|---|---|---|
| P25311 | AZGP1 | -0.30571 | 12.1082 | -0.59243 | 0.55597 | 0.965945 | -4.6027 |
| O43866 | CD5L | -0.29038 | 13.65287 | -0.59053 | 0.557239 | 0.965945 | -4.60272 |
| Q99832 | CCT7 | -0.31114 | 11.03507 | -0.58875 | 0.558423 | 0.965945 | -4.60275 |
| Q16555 | DPYSL2 | -0.28869 | 14.86964 | -0.58817 | 0.558805 | 0.965945 | -4.60276 |
| P02741 | CRP | 0.719041 | 11.11435 | 0.587419 | 0.559352 | 0.965945 | -4.60277 |
| P55072 | VCP | 0.363967 | 13.88593 | 0.586575 | 0.559871 | 0.965945 | -4.60278 |
| Q16658 | FSCN1 | 0.353498 | 13.04135 | 0.583584 | 0.561868 | 0.965945 | -4.60282 |
| Q13247 | SRSF6 | -0.39255 | 8.67543 | -0.58341 | 0.56207 | 0.965945 | -4.60282 |
| P13533 | MYH6 | -0.31901 | 10.46117 | -0.58276 | 0.562421 | 0.965945 | -4.60283 |
| O00410 | IPO5 | 0.469808 | 9.76488 | 0.582069 | 0.562881 | 0.965945 | -4.60284 |
| P06454 | PTMA | 0.433172 | 9.310123 | 0.580533 | 0.563909 | 0.965945 | -4.60286 |
| Q15366 | PCBP2 | 0.560225 | 12.64187 | 0.57877 | 0.56509 | 0.965945 | -4.60288 |
| O60506 | SYNCRIP | 0.420354 | 8.844164 | 0.573779 | 0.56872 | 0.965945 | -4.60208 |
| Q13561 | DCTN2 | 0.381053 | 8.671465 | 0.573211 | 0.568953 | 0.965945 | -4.60209 |
| Q9H115 | NAPB | 0.362633 | 9.074792 | 0.572213 | 0.569825 | 0.965945 | -4.6021 |
| P35520 | CBS | 0.402908 | 8.684517 | 0.571198 | 0.570403 | 0.965945 | -4.60211 |
| P00747 | PLG | 0.356712 | 12.64273 | 0.568986 | 0.571664 | 0.965945 | -4.60301 |
| Q9NRX4 | PHPT1 | 0.435134 | 8.126823 | 0.561925 | 0.57761 | 0.965945 | -4.6015 |
| P14550 | AKR1A1 | -0.39032 | 7.585267 | -0.55974 | 0.57847 | 0.965945 | -4.60224 |
| O95865 | DDAH2 | 0.558543 | 8.695045 | 0.559315 | 0.578525 | 0.965945 | -4.60114 |
| P61981 | YWHAG | 0.511736 | 13.78262 | 0.555824 | 0.580569 | 0.965945 | -4.60319 |
| P00915 | CA1 | -0.37797 | 10.62668 | -0.55323 | 0.582335 | 0.965945 | -4.60322 |
| Q04695 | KRT17 | -0.38565 | 10.32094 | -0.549 | 0.585338 | 0.965945 | -4.60327 |
| A0A0C4DH | #N/D | -0.4708 | 9.855679 | -0.54916 | 0.585366 | 0.965945 | -4.60327 |
| Q5IS74 | #N/D | 0.428408 | 8.979446 | 0.545313 | 0.587766 | 0.965945 | -4.60332 |
| P62081 | RPS7 | 0.334135 | 10.42682 | 0.542538 | 0.589624 | 0.965945 | -4.60336 |
| P48637 | GSS | 0.549281 | 8.001266 | 0.543176 | 0.590067 | 0.965945 | -4.60006 |
| P45880 | VDAC2 | 0.295325 | 11.73893 | 0.540253 | 0.591187 | 0.965945 | -4.60338 |
| P02538 | KRT6A | 0.56611 | 11.77848 | 0.539107 | 0.592096 | 0.965945 | -4.6034 |
| P61978 | HNRNPK | -0.42013 | 11.47478 | -0.53561 | 0.594374 | 0.965945 | -4.60344 |
| P11166 | SLC2A1 | 0.418442 | 9.8587 | 0.535368 | 0.594537 | 0.965945 | -4.60345 |

| | | | | | | | |
|---|---|---|---|---|---|---|---|
| P46777 | RPL5 | 0.412097 | 8.455774 | 0.530446 | 0.598387 | 0.965945 | -4.60183 |
| P02671 | FGA | 0.257871 | 12.54848 | 0.525751 | 0.601159 | 0.965945 | -4.60356 |
| Q8IV08 | PLD3 | -0.36383 | 9.113681 | -0.52431 | 0.602194 | 0.965945 | -4.60358 |
| P68104 | EEF1A1 | -0.36165 | 9.086982 | -0.51504 | 0.608611 | 0.965945 | -4.60369 |
| Q92598-4 | #N/D | -0.49028 | 8.188924 | -0.51351 | 0.610464 | 0.965945 | -4.60101 |
| Q99714 | HSD17B10 | 0.464481 | 7.015178 | 0.516041 | 0.610665 | 0.965945 | -4.59769 |
| Q9NSD9 | FARSB | 0.365683 | 9.390867 | 0.509913 | 0.612429 | 0.965945 | -4.6028 |
| P11216 | PYGB | 0.26869 | 10.47934 | 0.503761 | 0.616426 | 0.965945 | -4.60383 |
| A0A075B6I | #N/D | 0.21845 | 13.46962 | 0.503526 | 0.61659 | 0.965945 | -4.60383 |
| P04179 | SOD2 | 0.309577 | 10.77234 | 0.501921 | 0.617711 | 0.965945 | -4.60385 |
| P19338 | NCL | -0.36619 | 14.24 | -0.50053 | 0.618681 | 0.965945 | -4.60387 |
| P62826 | RAN | -0.23892 | 12.49633 | -0.49966 | 0.619293 | 0.965945 | -4.60388 |
| P01615 | #N/D | -0.42907 | 11.88343 | -0.49931 | 0.619873 | 0.965945 | -4.60213 |
| P02765 | AHSG | -0.3142 | 11.76711 | -0.49827 | 0.620266 | 0.965945 | -4.60389 |
| P04114 | APOB | -0.4254 | 15.2199 | -0.497 | 0.621152 | 0.965945 | -4.60391 |
| P23528 | CFL1 | -0.29666 | 12.52033 | -0.49689 | 0.621233 | 0.965945 | -4.60391 |
| Q15102 | PAFAH1B3 | -0.40648 | 9.201983 | -0.49442 | 0.623039 | 0.965945 | -4.60394 |
| P34897 | SHMT2 | 0.331139 | 10.26867 | 0.493127 | 0.623871 | 0.965945 | -4.60395 |
| P12268 | IMPDH2 | -0.3907 | 9.058515 | -0.49195 | 0.624975 | 0.965945 | -4.60298 |
| P62917 | RPL8 | 0.523761 | 11.02457 | 0.489159 | 0.626695 | 0.965945 | -4.604 |
| P11940 | PABPC1 | -0.30567 | 9.661148 | -0.4887 | 0.627057 | 0.965945 | -4.604 |
| P35579 | MYH9 | 0.377263 | 12.21283 | 0.488581 | 0.627067 | 0.965945 | -4.604 |
| E9PAV3 | #N/D | -0.44355 | 12.14083 | -0.48784 | 0.627625 | 0.965945 | -4.60303 |
| P12004 | PCNA | -0.24254 | 11.20119 | -0.48718 | 0.628053 | 0.965945 | -4.60402 |
| P39019 | RPS19 | -0.47256 | 12.71185 | -0.48616 | 0.628774 | 0.965945 | -4.60403 |
| P0C0L5 | C4B | 0.281004 | 12.58969 | 0.485313 | 0.629403 | 0.965945 | -4.60404 |
| P26639 | TARS1 | 0.289711 | 9.377104 | 0.484784 | 0.629776 | 0.965945 | -4.60405 |
| P62942 | FKBP1A | 0.471361 | 10.83055 | 0.483488 | 0.630654 | 0.965945 | -4.60406 |
| P01011 | SERPINA3 | 0.194301 | 14.45594 | 0.476954 | 0.63527 | 0.965945 | -4.60413 |
| P02792 | FTL | 0.324384 | 9.643032 | 0.47009 | 0.640241 | 0.965945 | -4.60421 |
| P61088 | UBE2N | 0.348029 | 11.45261 | 0.465289 | 0.643616 | 0.965945 | -4.60426 |

| | | | | | | | |
|---|---|---|---|---|---|---|---|
| Q01813 | PFKP | 0.253905 | 10.6556 | 0.463788 | 0.644617 | 0.965945 | -4.60428 |
| P02766 | TTR | -0.40976 | 13.95342 | -0.46371 | 0.64467 | 0.965945 | -4.60428 |
| P07339 | CTSD | 0.355667 | 12.19536 | 0.458062 | 0.648699 | 0.965945 | -4.60434 |
| Q9H299 | SH3BGRL3 | -0.34001 | 8.301393 | -0.45723 | 0.649638 | 0.965945 | -4.60333 |
| P43487 | RANBP1 | -0.33273 | 11.29012 | -0.4535 | 0.652172 | 0.965945 | -4.60336 |
| Q92777 | SYN2 | 0.435041 | 10.07691 | 0.452988 | 0.652326 | 0.965945 | -4.60439 |
| A5YM72 | CARNS1 | -0.41599 | 9.654885 | -0.45268 | 0.652847 | 0.965945 | -4.60255 |
| P69905 | HBA1 | -0.22684 | 14.01226 | -0.449 | 0.655181 | 0.965945 | -4.60444 |
| P09211 | GSTP1 | -0.33027 | 13.54278 | -0.44799 | 0.656293 | 0.965945 | -4.60212 |
| Q7Z3B1 | NEGR1 | 0.43413 | 7.928181 | 0.447545 | 0.656759 | 0.965945 | -4.60047 |
| P53634 | CTSC | -0.32779 | 8.483897 | -0.44548 | 0.65796 | 0.965945 | -4.60344 |
| A0A0G2JS( | #N/D | 0.599249 | 9.269116 | 0.444868 | 0.658528 | 0.965945 | -4.60048 |
| P09382 | LGALS1 | -0.32334 | 9.996774 | -0.44352 | 0.659119 | 0.965945 | -4.60449 |
| P02760 | AMBP | 0.20813 | 13.73303 | 0.442214 | 0.660055 | 0.965945 | -4.60451 |
| P0DOY3 | #N/D | -0.38052 | 15.80861 | -0.44119 | 0.660792 | 0.965945 | -4.60452 |
| P02788 | LTF | -0.4467 | 9.520811 | -0.43893 | 0.662481 | 0.965945 | -4.60454 |
| A0A075B6. | #N/D | 0.4216 | 10.90519 | 0.436412 | 0.66433 | 0.965945 | -4.60456 |
| Q9Y696 | CLIC4 | -0.41934 | 8.300412 | -0.43671 | 0.664344 | 0.965945 | -4.60221 |
| P06744 | GPI | -0.25495 | 11.87388 | -0.4317 | 0.667634 | 0.965945 | -4.60461 |
| Q15424 | SAFB | -0.25945 | 10.29881 | -0.43168 | 0.66768 | 0.965945 | -4.60461 |
| P10809 | HSPD1 | -0.19137 | 14.28311 | -0.43073 | 0.668336 | 0.965945 | -4.60462 |
| P53004 | BLVRA | -0.33016 | 7.896539 | -0.4288 | 0.669972 | 0.965945 | -4.60359 |
| P07225 | PROS1 | -0.18353 | 13.16055 | -0.42591 | 0.671824 | 0.965945 | -4.60467 |
| P05090 | APOD | 0.238169 | 14.6869 | 0.422963 | 0.673959 | 0.965945 | -4.6047 |
| P01859 | #N/D | 0.335411 | 13.5427 | 0.419945 | 0.67615 | 0.965945 | -4.60473 |
| Q92752 | TNR | 0.195157 | 11.57348 | 0.418782 | 0.676994 | 0.965945 | -4.60474 |
| Q5T7N2 | L1TD1 | 0.248743 | 8.91276 | 0.418932 | 0.677045 | 0.965945 | -4.60368 |
| P61970 | NUTF2 | -0.30923 | 8.594163 | -0.41789 | 0.677672 | 0.965945 | -4.60475 |
| Q9UMF0 | ICAM5 | -0.27137 | 9.084601 | -0.41647 | 0.678678 | 0.965945 | -4.60476 |
| Q96C19 | EFHD2 | -0.40457 | 8.816843 | -0.41352 | 0.681172 | 0.965945 | -4.60287 |
| P11217 | PYGM | -0.21977 | 10.22972 | -0.41236 | 0.681663 | 0.965945 | -4.6048 |

| | | | | | | | |
|---|---|---|---|---|---|---|---|
| P48147 | PREP | -0.28506 | 7.680943 | -0.41261 | 0.681973 | 0.965945 | -4.60373 |
| P10909 | CLU | 0.16269 | 13.71219 | 0.411844 | 0.682042 | 0.965945 | -4.60481 |
| P13797 | PLS3 | -0.26427 | 9.880516 | -0.41168 | 0.682159 | 0.965945 | -4.60481 |
| P09936 | UCHL1 | -0.24023 | 12.48925 | -0.41139 | 0.682371 | 0.965945 | -4.60481 |
| Q9Y2T3 | GDA | 0.309744 | 10.08686 | 0.408212 | 0.68475 | 0.965945 | -4.60484 |
| A0A075B6I | #N/D | -0.41002 | 7.939523 | -0.40934 | 0.684754 | 0.965945 | -4.60031 |
| P01780 | #N/D | -0.34428 | 11.11587 | -0.40787 | 0.684943 | 0.965945 | -4.60485 |
| P07910 | HNRNPC | -0.18156 | 10.97799 | -0.40776 | 0.685019 | 0.965945 | -4.60485 |
| Q9UQM7 | CAMK2A | -0.32096 | 11.74729 | -0.40739 | 0.685294 | 0.965945 | -4.60485 |
| P49321 | NASP | -0.23167 | 13.01531 | -0.40602 | 0.686289 | 0.965945 | -4.60487 |
| A0A0C4DH | #N/D | -0.22365 | 12.25946 | -0.40534 | 0.686792 | 0.965945 | -4.60487 |
| Q14195-2 | #N/D | 0.207547 | 12.44951 | 0.404101 | 0.687694 | 0.965945 | -4.60488 |
| Q12906-7 | #N/D | -0.26392 | 10.81171 | -0.40268 | 0.688732 | 0.965945 | -4.6049 |
| P01714 | #N/D | 0.34801 | 10.84083 | 0.400092 | 0.690627 | 0.965945 | -4.60492 |
| O60282 | KIF5C | 0.308349 | 9.337804 | 0.399745 | 0.690969 | 0.965945 | -4.60492 |
| Q14240 | EIF4A2 | 0.238439 | 9.225594 | 0.397201 | 0.692745 | 0.965945 | -4.60495 |
| Q7KZF4 | SND1 | 0.275156 | 9.615375 | 0.396971 | 0.693001 | 0.965945 | -4.60495 |
| P05387 | RPLP2 | 0.241983 | 9.837368 | 0.396162 | 0.693535 | 0.965945 | -4.60496 |
| Q96GD0 | PDXP | 0.266934 | 10.49093 | 0.395607 | 0.693914 | 0.965945 | -4.60496 |
| Q00839 | HNRNPU | -0.20685 | 12.5168 | -0.39342 | 0.695521 | 0.965945 | -4.60498 |
| P43652 | AFM | -0.19258 | 12.25694 | -0.39183 | 0.696689 | 0.965945 | -4.605 |
| Q9Y3E1 | HDGFL3 | 0.379063 | 8.28437 | 0.391635 | 0.696915 | 0.965945 | -4.605 |
| P39023 | RPL3 | 0.489818 | 11.33349 | 0.390711 | 0.697509 | 0.965945 | -4.60501 |
| A0A0B4J1V | #N/D | -0.43322 | 8.761429 | -0.38657 | 0.701106 | 0.965945 | -4.60194 |
| P61313 | RPL15 | 0.170312 | 10.74977 | 0.38474 | 0.701902 | 0.965945 | -4.60506 |
| P17987 | TCP1 | 0.323239 | 10.0471 | 0.384154 | 0.702334 | 0.965945 | -4.60507 |
| Q15907 | RAB11B | 0.241695 | 9.911453 | 0.37866 | 0.706414 | 0.965945 | -4.60512 |
| P01594 | #N/D | 0.260927 | 13.40637 | 0.378402 | 0.706721 | 0.965945 | -4.60512 |
| Q9NZL9 | MAT2B | -0.31863 | 9.370227 | -0.37627 | 0.708179 | 0.965945 | -4.60514 |
| P04075 | ALDOA | -0.17928 | 13.09657 | -0.37524 | 0.708918 | 0.965945 | -4.60515 |
| P02689 | PMP2 | -0.27584 | 10.41906 | -0.37449 | 0.709471 | 0.965945 | -4.60515 |

| | | | | | | | |
|---|---|---|---|---|---|---|---|
| A0A075B6I | #N/D | 0.206921 | 14.94383 | 0.371969 | 0.711334 | 0.965945 | -4.60517 |
| P09429 | HMGB1 | 0.180399 | 11.33429 | 0.371219 | 0.71189 | 0.965945 | -4.60518 |
| P46821 | MAP1B | 0.220243 | 12.35155 | 0.369862 | 0.712895 | 0.965945 | -4.60519 |
| P17174 | GOT1 | 0.250631 | 13.06163 | 0.369421 | 0.713222 | 0.965945 | -4.6052 |
| P53680 | AP2S1 | -0.32559 | 7.833872 | -0.36899 | 0.713974 | 0.965945 | -4.60052 |
| P21796 | VDAC1 | -0.38055 | 8.076572 | -0.36801 | 0.714699 | 0.965945 | -4.60091 |
| Q16181 | SEPTIN7 | 0.471774 | 11.66741 | 0.36407 | 0.717191 | 0.965945 | -4.60524 |
| Q12765 | SCRN1 | -0.1521 | 11.08891 | -0.36177 | 0.718901 | 0.965945 | -4.60526 |
| O75368 | SH3BGRL | -0.33938 | 9.591212 | -0.36174 | 0.718974 | 0.965945 | -4.60526 |
| Q13938 | #N/D | 0.202881 | 11.20708 | 0.361272 | 0.71927 | 0.965945 | -4.60527 |
| Q9NPH9 | IL26 | -0.36989 | 9.277746 | -0.36138 | 0.719325 | 0.965945 | -4.60415 |
| O14576 | DYNC1I1 | 0.281948 | 8.850936 | 0.360283 | 0.72003 | 0.965945 | -4.60527 |
| A0A0B4J1V | #N/D | -0.25634 | 12.65679 | -0.36005 | 0.720179 | 0.965945 | -4.60528 |
| P07996 | THBS1 | -0.16761 | 13.73619 | -0.3595 | 0.72059 | 0.965945 | -4.60528 |
| P09104 | ENO2 | -0.19326 | 11.61892 | -0.35645 | 0.722857 | 0.965945 | -4.60531 |
| Q16775 | HAGH | -0.30296 | 10.09877 | -0.35078 | 0.727081 | 0.965945 | -4.60535 |
| P21926 | CD9 | -0.27653 | 10.94564 | -0.35063 | 0.727192 | 0.965945 | -4.60536 |
| P01871 | #N/D | 0.233973 | 16.51568 | 0.350583 | 0.72723 | 0.965945 | -4.60536 |
| P63241 | EIF5A | -0.24206 | 8.213411 | -0.34659 | 0.730399 | 0.965945 | -4.60425 |
| P17844 | DDX5 | -0.15684 | 11.72587 | -0.34527 | 0.731198 | 0.965945 | -4.6054 |
| O76021 | RSL1D1 | 0.263911 | 9.23965 | 0.34489 | 0.731557 | 0.965945 | -4.6054 |
| P05455 | SSB | 0.283268 | 10.60567 | 0.342674 | 0.733164 | 0.965945 | -4.60542 |
| Q92522 | H1-10 | -0.17447 | 9.944587 | -0.34185 | 0.733758 | 0.965945 | -4.60543 |
| Q9NUQ9 | CYRIB | 0.317155 | 9.681427 | 0.341715 | 0.733932 | 0.965945 | -4.60287 |
| P47914 | RPL29 | 0.254252 | 8.941895 | 0.34171 | 0.733962 | 0.965945 | -4.60543 |
| P17600 | SYN1 | 0.260236 | 12.09097 | 0.339865 | 0.735243 | 0.965945 | -4.60544 |
| P02768 | ALB | 0.29385 | 17.61783 | 0.339773 | 0.735312 | 0.965945 | -4.60544 |
| P29218 | IMPA1 | 0.20208 | 11.50747 | 0.338017 | 0.736628 | 0.965945 | -4.60546 |
| P09651 | HNRNPA1 | -0.22097 | 12.21855 | -0.33783 | 0.736769 | 0.965945 | -4.60546 |
| Q99962 | SH3GL2 | 0.171905 | 12.54019 | 0.337418 | 0.737076 | 0.965945 | -4.60546 |
| Q03591 | CFHR1 | 0.219289 | 9.2033 | 0.334481 | 0.739432 | 0.965945 | -4.60434 |

| | | | | | | | |
|---|---|---|---|---|---|---|---|
| P62258 | YWHAE | 0.14483 | 12.79759 | 0.332413 | 0.740831 | 0.965945 | -4.6055 |
| P11142 | HSPA8 | 0.167991 | 13.19735 | 0.330682 | 0.742132 | 0.965945 | -4.60551 |
| Q15185 | PTGES3 | -0.36846 | 14.08679 | -0.3298 | 0.742819 | 0.965945 | -4.60552 |
| P62899 | RPL31 | -0.34027 | 12.22519 | -0.32375 | 0.747346 | 0.965945 | -4.60557 |
| P60174 | TPI1 | 0.328582 | 13.33968 | 0.318901 | 0.751 | 0.965945 | -4.6056 |
| P01766 | #N/D | -0.21592 | 9.036312 | -0.31734 | 0.752325 | 0.965945 | -4.60561 |
| P48740 | MASP1 | 0.221611 | 8.81856 | 0.316334 | 0.752982 | 0.965945 | -4.60562 |
| P31327 | CPS1 | 0.245359 | 9.318571 | 0.315619 | 0.753477 | 0.965945 | -4.60563 |
| P14618-2 | #N/D | 0.259348 | 11.99726 | 0.313779 | 0.754867 | 0.965945 | -4.60564 |
| P00739 | HPR | 0.236707 | 13.20664 | 0.311609 | 0.756507 | 0.965945 | -4.60566 |
| P08185 | SERPINA6 | -0.299 | 9.106814 | -0.31176 | 0.756909 | 0.965945 | -4.6002 |
| P60201 | PLP1 | 0.172663 | 12.86375 | 0.310879 | 0.757059 | 0.965945 | -4.60566 |
| P21283 | ATP6V1C1 | 0.277413 | 9.48756 | 0.31093 | 0.757064 | 0.965945 | -4.60566 |
| P14649 | MYL6B | 0.166876 | 11.20495 | 0.309149 | 0.758368 | 0.965945 | -4.60568 |
| P02649 | APOE | -0.2176 | 16.1627 | -0.30888 | 0.758569 | 0.965945 | -4.60568 |
| P13591 | NCAM1 | 0.205675 | 12.6968 | 0.306614 | 0.760286 | 0.965945 | -4.60569 |
| P01764 | #N/D | -0.23112 | 9.51328 | -0.30652 | 0.760496 | 0.965945 | -4.60453 |
| Q12860 | CNTN1 | -0.20162 | 11.73525 | -0.30601 | 0.760746 | 0.965945 | -4.6057 |
| P49418 | AMPH | 0.246694 | 10.79496 | 0.305937 | 0.760799 | 0.965945 | -4.6057 |
| O75083 | WDR1 | -0.15057 | 11.68371 | -0.30493 | 0.761563 | 0.965945 | -4.60571 |
| Q08209-3 | #N/D | -0.17973 | 10.11222 | -0.30447 | 0.761934 | 0.965945 | -4.60571 |
| P36955 | SERPINF1 | 0.178993 | 12.75803 | 0.301538 | 0.764133 | 0.965945 | -4.60573 |
| O95989 | NUDT3 | -0.23018 | 9.707781 | -0.30106 | 0.764493 | 0.965945 | -4.60573 |
| P09493-4 | #N/D | 0.202094 | 12.6635 | 0.300985 | 0.764552 | 0.965945 | -4.60573 |
| P16401 | H1-5 | 0.231035 | 10.04102 | 0.300071 | 0.765246 | 0.965945 | -4.60574 |
| P80108 | GPLD1 | -0.20908 | 9.224428 | -0.29675 | 0.767833 | 0.965945 | -4.60459 |
| P13639 | EEF2 | 0.164176 | 12.11228 | 0.296168 | 0.768209 | 0.965945 | -4.60577 |
| Q13813 | SPTAN1 | -0.10866 | 11.99901 | -0.29601 | 0.768331 | 0.965945 | -4.60577 |
| P0DOX5 | #N/D | -0.15745 | 14.50645 | -0.29551 | 0.768709 | 0.965945 | -4.60577 |
| P04259 | KRT6B | -0.22355 | 11.57937 | -0.29339 | 0.770338 | 0.965945 | -4.60579 |
| P30084 | ECHS1 | 0.137086 | 10.15609 | 0.292133 | 0.771276 | 0.965945 | -4.6058 |

| | | | | | | | |
|---|---|---|---|---|---|---|---|
| P07195 | LDHB | 0.130739 | 13.41831 | 0.290282 | 0.772684 | 0.965945 | -4.60581 |
| P61266-2 | #N/D | -0.13505 | 9.516859 | -0.29012 | 0.772807 | 0.965945 | -4.60581 |
| P11766 | ADH5 | -0.25405 | 10.15171 | -0.29012 | 0.77281 | 0.965945 | -4.60581 |
| P27824 | CANX | 0.226372 | 11.05687 | 0.289762 | 0.77308 | 0.965945 | -4.60581 |
| O14818 | PSMA7 | -0.17487 | 8.736898 | -0.28864 | 0.773951 | 0.965945 | -4.60582 |
| P62424 | RPL7A | 0.117327 | 11.51419 | 0.288465 | 0.774067 | 0.965945 | -4.60582 |
| A0A0B4J1U | #N/D | 0.201082 | 9.305291 | 0.288345 | 0.77434 | 0.965945 | -4.60464 |
| Q86YZ3 | HRNR | -0.29661 | 10.14946 | -0.28714 | 0.775181 | 0.965945 | -4.60465 |
| P18124 | RPL7 | -0.1537 | 10.99238 | -0.28547 | 0.776348 | 0.965945 | -4.60584 |
| P55084 | HADHB | -0.18068 | 8.238028 | -0.28367 | 0.777927 | 0.965945 | -4.60467 |
| P01591 | JCHAIN | -0.16311 | 14.17635 | -0.27914 | 0.781174 | 0.965945 | -4.60588 |
| P40939 | HADHA | 0.272057 | 9.753015 | 0.277909 | 0.782135 | 0.965945 | -4.6047 |
| P26583 | HMGB2 | 0.20523 | 9.427067 | 0.277407 | 0.7825 | 0.965945 | -4.60589 |
| P40926 | MDH2 | -0.13942 | 13.17949 | -0.27657 | 0.783142 | 0.965945 | -4.6059 |
| Q5HYA8 | TMEM67 | -0.22338 | 9.59421 | -0.27553 | 0.783934 | 0.965945 | -4.60591 |
| P19367 | HK1 | -0.13133 | 11.89524 | -0.27376 | 0.785289 | 0.965945 | -4.60592 |
| P08195 | SLC3A2 | 0.207907 | 10.25962 | 0.272426 | 0.786326 | 0.965945 | -4.60593 |
| P00450 | CP | 0.156657 | 14.6821 | 0.271921 | 0.786694 | 0.965945 | -4.60593 |
| Q15365 | PCBP1 | -0.21798 | 11.13094 | -0.26731 | 0.790261 | 0.965945 | -4.60476 |
| P25789 | PSMA4 | -0.16817 | 7.883878 | -0.26438 | 0.792612 | 0.965945 | -4.60478 |
| Q9Y2J2 | EPB41L3 | 0.196225 | 8.157606 | 0.261399 | 0.795021 | 0.965945 | -4.60384 |
| Q13491-4 | #N/D | 0.167124 | 9.101016 | 0.260806 | 0.795245 | 0.965945 | -4.606 |
| P48426 | PIP4K2A | -0.18209 | 8.06123 | -0.26113 | 0.79535 | 0.965945 | -4.60095 |
| O75347 | TBCA | -0.17862 | 9.096358 | -0.25928 | 0.796377 | 0.965945 | -4.60601 |
| P0DOX3 | #N/D | -0.11281 | 13.40279 | -0.25901 | 0.796588 | 0.965945 | -4.60601 |
| P32322 | PYCR1 | 0.339552 | 8.954011 | 0.258591 | 0.796945 | 0.965945 | -4.60481 |
| P02751-8 | #N/D | 0.125927 | 15.47018 | 0.257212 | 0.797968 | 0.965945 | -4.60602 |
| P25705 | ATP5F1A | -0.25706 | 13.38486 | -0.25709 | 0.798063 | 0.965945 | -4.60602 |
| P20851-2 | #N/D | -0.19843 | 11.52893 | -0.25692 | 0.798195 | 0.965945 | -4.60602 |
| Q99798 | ACO2 | -0.13728 | 13.35696 | -0.25559 | 0.799217 | 0.965945 | -4.60603 |
| P55056 | APOC4 | 0.236782 | 9.255911 | 0.254933 | 0.800007 | 0.965945 | -4.60169 |

| | | | | | | | |
|---|---|---|---|---|---|---|---|
| P02533 | KRT14 | 0.201284 | 11.09842 | 0.253705 | 0.800663 | 0.965945 | -4.60604 |
| P68402 | PAFAH1B2 | 0.352218 | 6.807079 | 0.254024 | 0.802292 | 0.965945 | -4.59805 |
| P08133 | ANXA6 | -0.10888 | 11.75849 | -0.2504 | 0.803207 | 0.965945 | -4.60606 |
| Q15848 | ADIPOQ | 0.173689 | 10.82374 | 0.249803 | 0.803664 | 0.965945 | -4.60606 |
| P09543 | CNP | -0.10456 | 12.18182 | -0.24962 | 0.803806 | 0.965945 | -4.60607 |
| P50914 | RPL14 | -0.17762 | 9.06483 | -0.2484 | 0.804812 | 0.965945 | -4.60607 |
| P12277 | CKB | 0.174272 | 13.54112 | 0.247805 | 0.805201 | 0.965945 | -4.60608 |
| P09661 | SNRPA1 | 0.303183 | 11.24558 | 0.24661 | 0.806121 | 0.965945 | -4.60608 |
| P15104 | GLUL | -0.18216 | 13.57473 | -0.24655 | 0.80617 | 0.965945 | -4.60608 |
| Q00325 | SLC25A3 | -0.21673 | 11.25621 | -0.24547 | 0.806999 | 0.965945 | -4.60609 |
| P50991 | CCT4 | -0.12532 | 13.00041 | -0.24347 | 0.80854 | 0.965945 | -4.6061 |
| P04003 | C4BPA | -0.13304 | 14.39715 | -0.23991 | 0.811284 | 0.965945 | -4.60612 |
| P18669 | PGAM1 | 0.131281 | 13.03714 | 0.239337 | 0.811727 | 0.965945 | -4.60612 |
| Q9UDR5 | AASS | -0.30696 | 10.55739 | -0.23918 | 0.811845 | 0.965945 | -4.60613 |
| P35080 | PFN2 | 0.175742 | 10.95784 | 0.237824 | 0.812911 | 0.965945 | -4.60613 |
| P46108 | CRK | -0.15733 | 8.591725 | -0.23759 | 0.813143 | 0.965945 | -4.60492 |
| Q99447 | PCYT2 | 0.214983 | 9.931576 | 0.235571 | 0.814634 | 0.965945 | -4.60615 |
| P23526 | AHCY | 0.119856 | 12.4871 | 0.235468 | 0.814714 | 0.965945 | -4.60615 |
| Q92686 | NRGN | 0.223062 | 9.047773 | 0.234534 | 0.815484 | 0.965945 | -4.60342 |
| Q01082 | SPTBN1 | -0.09052 | 11.78071 | -0.23041 | 0.818624 | 0.965945 | -4.60617 |
| Q16695 | H3-4 | -0.1514 | 10.26812 | -0.22969 | 0.819182 | 0.965945 | -4.60618 |
| P0C0L4 | C4A | -0.23714 | 11.8147 | -0.22932 | 0.819603 | 0.965945 | -4.60496 |
| Q9H9Z2 | LIN28A | 0.184004 | 9.50441 | 0.225015 | 0.82281 | 0.965945 | -4.6062 |
| P05023 | ATP1A1 | -0.15276 | 13.61348 | -0.22424 | 0.823395 | 0.965945 | -4.60621 |
| O43143 | DHX15 | 0.190074 | 7.908015 | 0.224132 | 0.823784 | 0.965945 | -4.60275 |
| P43243 | MATR3 | 0.192629 | 7.788896 | 0.223055 | 0.824739 | 0.965945 | -4.60401 |
| P11169 | SLC2A3 | -0.1825 | 9.491977 | -0.22199 | 0.825166 | 0.965945 | -4.60622 |
| P80723 | BASP1 | 0.196586 | 10.51359 | 0.220213 | 0.826514 | 0.965945 | -4.60623 |
| P08603 | CFH | 0.12566 | 14.2247 | 0.219928 | 0.826735 | 0.965945 | -4.60623 |
| P49419 | ALDH7A1 | -0.15258 | 13.146 | -0.21797 | 0.828253 | 0.965945 | -4.60624 |
| Q96F07 | CYFIP2 | -0.24329 | 9.351898 | -0.21742 | 0.828742 | 0.965945 | -4.60624 |

| | | | | | | | |
|---|---|---|---|---|---|---|---|
| P02749 | APOH | -0.10924 | 12.64195 | -0.21105 | 0.833622 | 0.965945 | -4.60627 |
| A0A0J9YXX | #N/D | -0.16231 | 10.65969 | -0.21037 | 0.834149 | 0.965945 | -4.60628 |
| P29622 | SERPINA4 | 0.18139 | 8.812058 | 0.210422 | 0.834323 | 0.965945 | -4.6028 |
| Q969P0 | IGSF8 | -0.12643 | 10.87703 | -0.21015 | 0.834324 | 0.965945 | -4.60628 |
| P19652 | ORM2 | 0.113322 | 9.423031 | 0.209762 | 0.834699 | 0.965945 | -4.60628 |
| Q14204 | DYNC1H1 | -0.10081 | 13.22936 | -0.20899 | 0.835224 | 0.965945 | -4.60628 |
| O43813 | LANCL1 | -0.1506 | 12.55751 | -0.20772 | 0.836211 | 0.965945 | -4.60629 |
| P13645 | KRT10 | 0.122815 | 15.33348 | 0.207623 | 0.836284 | 0.965945 | -4.60629 |
| P18206 | VCL | -0.08071 | 12.66382 | -0.20528 | 0.838108 | 0.965945 | -4.6063 |
| Q02543 | RPL18A | 0.201429 | 12.91705 | 0.201523 | 0.841028 | 0.965945 | -4.60632 |
| P52758 | RIDA | -0.1282 | 8.569229 | -0.20006 | 0.842206 | 0.965945 | -4.60633 |
| P12532 | CKMT1B | 0.133415 | 9.736519 | 0.197637 | 0.844053 | 0.965945 | -4.60634 |
| Q9UBC3 | DNMT3B | 0.183055 | 11.72397 | 0.197199 | 0.844393 | 0.965945 | -4.60634 |
| O14531 | DPYSL4 | 0.147555 | 8.30417 | 0.197172 | 0.844635 | 0.965945 | -4.60284 |
| P12814 | ACTN1 | -0.19505 | 7.963896 | -0.1948 | 0.846602 | 0.965945 | -4.60054 |
| P01031 | C5 | -0.07427 | 12.98874 | -0.19283 | 0.847797 | 0.965945 | -4.60636 |
| Q9UHD8 | SEPTIN9 | 0.137989 | 7.179257 | 0.191477 | 0.849245 | 0.965945 | -4.60055 |
| P32004 | L1CAM | 0.137502 | 9.227821 | 0.187801 | 0.851801 | 0.965945 | -4.60359 |
| P20339 | RAB5A | 0.109504 | 8.711876 | 0.187417 | 0.852018 | 0.965945 | -4.60638 |
| O94819 | KBTBD11 | 0.119948 | 9.342712 | 0.18667 | 0.852613 | 0.965945 | -4.60639 |
| P01009 | SERPINA1 | 0.094219 | 14.60704 | 0.185621 | 0.85342 | 0.965945 | -4.60639 |
| P09622 | DLD | 0.135334 | 8.688977 | 0.185798 | 0.853449 | 0.965945 | -4.60416 |
| P33993 | MCM7 | -0.11895 | 8.945521 | -0.1843 | 0.854463 | 0.965945 | -4.6064 |
| Q15717 | ELAVL1 | -0.164 | 11.8003 | -0.18417 | 0.854668 | 0.965945 | -4.6036 |
| Q08380 | LGALS3BP | -0.11074 | 14.57471 | -0.18321 | 0.855303 | 0.965945 | -4.6064 |
| P01742 | #N/D | -0.13433 | 13.16148 | -0.18341 | 0.855333 | 0.965945 | -4.60416 |
| P01599 | #N/D | 0.152108 | 9.808588 | 0.182458 | 0.855914 | 0.965945 | -4.60641 |
| Q9NRV9 | HEBP1 | -0.16989 | 10.05472 | -0.18117 | 0.856978 | 0.965945 | -4.60517 |
| P01701 | #N/D | 0.166107 | 10.54093 | 0.179152 | 0.858523 | 0.965945 | -4.60418 |
| P52209 | PGD | 0.077923 | 9.886713 | 0.17784 | 0.859497 | 0.965945 | -4.60643 |
| P50453 | SERPINB9 | 0.164995 | 9.530376 | 0.176443 | 0.860667 | 0.965945 | -4.60519 |

| | | | | | | | |
|---|---|---|---|---|---|---|---|
| P05091 | ALDH2 | -0.10337 | 12.45982 | -0.17419 | 0.862349 | 0.965945 | -4.60644 |
| Q14624 | ITIH4 | 0.170391 | 9.94507 | 0.170163 | 0.865563 | 0.965945 | -4.60421 |
| P50213 | IDH3A | -0.11773 | 10.5341 | -0.16994 | 0.865676 | 0.965945 | -4.60646 |
| P42766 | RPL35 | -0.09921 | 9.474201 | -0.16914 | 0.866317 | 0.965945 | -4.60646 |
| P51884 | LUM | 0.148306 | 8.535854 | 0.168678 | 0.866783 | 0.965945 | -4.6012 |
| O43390 | HNRNPR | -0.09928 | 8.163912 | -0.16714 | 0.86793 | 0.965945 | -4.60522 |
| P14136 | GFAP | -0.0813 | 13.25535 | -0.16564 | 0.869044 | 0.965945 | -4.60647 |
| P13671 | C6 | 0.077789 | 13.17346 | 0.165298 | 0.869311 | 0.965945 | -4.60648 |
| P13489 | RNH1 | -0.14497 | 9.869775 | -0.16475 | 0.869739 | 0.965945 | -4.60648 |
| P30086 | PEBP1 | -0.0785 | 12.34105 | -0.16408 | 0.870268 | 0.965945 | -4.60648 |
| P04350 | TUBB4A | -0.11129 | 11.70512 | -0.16233 | 0.871638 | 0.965945 | -4.60649 |
| P32119 | PRDX2 | 0.089566 | 13.16617 | 0.161448 | 0.872328 | 0.965945 | -4.60649 |
| P09972 | ALDOC | 0.084852 | 13.29965 | 0.161133 | 0.872574 | 0.965945 | -4.60649 |
| P04216 | THY1 | -0.15183 | 9.984486 | -0.15889 | 0.874334 | 0.966661 | -4.6065 |
| Q06830 | PRDX1 | 0.082436 | 12.88336 | 0.156066 | 0.876548 | 0.966677 | -4.60651 |
| Q92945 | KHSRP | -0.11223 | 11.19284 | -0.15604 | 0.876572 | 0.966677 | -4.60651 |
| Q9NR46 | SH3GLB2 | -0.09638 | 8.407993 | -0.1506 | 0.880869 | 0.967974 | -4.60653 |
| P62701 | RPS4X | 0.066253 | 10.53558 | 0.149759 | 0.881498 | 0.967974 | -4.60653 |
| P12036 | NEFH | -0.05579 | 12.50765 | -0.14894 | 0.882145 | 0.967974 | -4.60654 |
| P41250 | GARS1 | 0.108073 | 9.552422 | 0.148108 | 0.882886 | 0.967974 | -4.60654 |
| P00505 | GOT2 | 0.07833 | 12.49377 | 0.147442 | 0.883318 | 0.967974 | -4.60654 |
| P07360 | C8G | 0.186373 | 8.367863 | 0.144022 | 0.886217 | 0.969927 | -4.59896 |
| P01611 | #N/D | 0.131824 | 10.90728 | 0.140993 | 0.888485 | 0.97037 | -4.60373 |
| P15121 | AKR1B1 | 0.094021 | 9.04325 | 0.140541 | 0.888854 | 0.97037 | -4.6053 |
| P12956 | XRCC6 | -0.10008 | 12.11524 | -0.13846 | 0.890382 | 0.970563 | -4.60657 |
| P23396 | RPS3 | -0.09424 | 12.74094 | -0.13733 | 0.891265 | 0.970563 | -4.60657 |
| P08519 | #N/D | 0.111811 | 15.54961 | 0.134192 | 0.893737 | 0.972037 | -4.60659 |
| P48506 | GCLC | -0.0918 | 8.866158 | -0.12714 | 0.899289 | 0.975861 | -4.60661 |
| P0C0S5 | H2AZ1 | 0.096069 | 10.02911 | 0.126874 | 0.899499 | 0.975861 | -4.60661 |
| P04275 | VWF | -0.08772 | 14.63316 | -0.11724 | 0.907094 | 0.982874 | -4.60664 |
| P38159 | RBMX | -0.11453 | 13.33236 | -0.11466 | 0.909129 | 0.983852 | -4.60664 |

| | | | | | | | |
|---|---|---|---|---|---|---|---|
| P07737 | PFN1 | -0.11981 | 12.48645 | -0.11169 | 0.91151 | 0.984652 | -4.60436 |
| P08779 | KRT16 | 0.127518 | 8.523877 | 0.108897 | 0.913917 | 0.984652 | -4.60131 |
| Q9NR30 | DDX21 | -0.07265 | 8.743727 | -0.10803 | 0.914384 | 0.984652 | -4.60539 |
| Q14103 | HNRNPD | 0.111809 | 10.83965 | 0.107982 | 0.9144 | 0.984652 | -4.60666 |
| P04004 | VTN | 0.060335 | 14.98777 | 0.103736 | 0.917753 | 0.986062 | -4.60667 |
| O15540 | FABP7 | 0.049944 | 10.93642 | 0.102393 | 0.918814 | 0.986062 | -4.60667 |
| P60709 | ACTB | 0.061002 | 12.29443 | 0.102014 | 0.919114 | 0.986062 | -4.60667 |
| P05160 | F13B | -0.08736 | 11.66103 | -0.09995 | 0.92076 | 0.98661 | -4.60541 |
| P02746 | C1QB | 0.070566 | 14.57706 | 0.09707 | 0.923021 | 0.987613 | -4.60669 |
| P61247 | RPS3A | 0.041844 | 12.29605 | 0.095871 | 0.923969 | 0.987613 | -4.60669 |
| Q9Y6I3 | EPN1 | -0.05811 | 8.193578 | -0.09271 | 0.926559 | 0.989023 | -4.60542 |
| P26373 | RPL13 | 0.053447 | 10.14526 | 0.091324 | 0.927565 | 0.989023 | -4.6067 |
| P50502 | ST13 | 0.076031 | 12.29466 | 0.087212 | 0.930835 | 0.991294 | -4.60543 |
| P55290 | CDH13 | 0.062458 | 9.449083 | 0.08099 | 0.93577 | 0.991691 | -4.60672 |
| Q14123 | PDE1C | -0.09461 | 9.472294 | -0.07791 | 0.938191 | 0.991691 | -4.60673 |
| Q9H0U4 | RAB1B | -0.04971 | 8.154821 | -0.07699 | 0.938971 | 0.991691 | -4.60311 |
| P25786 | PSMA1 | -0.06668 | 8.552348 | -0.07561 | 0.940062 | 0.991691 | -4.60545 |
| P61254 | RPL26 | 0.047255 | 10.98212 | 0.074993 | 0.940491 | 0.991691 | -4.60673 |
| P52306-6 | #N/D | 0.056205 | 11.71115 | 0.074414 | 0.940949 | 0.991691 | -4.60673 |
| Q6UWR7 | ENPP6 | -0.0359 | 11.19948 | -0.06613 | 0.947512 | 0.991691 | -4.60675 |
| P14314 | PRKCSH | -0.04962 | 12.48603 | -0.06364 | 0.949499 | 0.991691 | -4.60675 |
| Q14974 | KPNB1 | -0.07147 | 12.05301 | -0.06355 | 0.949555 | 0.991691 | -4.60675 |
| P14868-2 | #N/D | -0.03907 | 8.099459 | -0.06273 | 0.950252 | 0.991691 | -4.60547 |
| P61956 | SUMO2 | -0.05402 | 9.670243 | -0.0619 | 0.950873 | 0.991691 | -4.60675 |
| P63244 | RACK1 | -0.04056 | 12.16177 | -0.06122 | 0.951404 | 0.991691 | -4.60675 |
| O14791 | APOL1 | 0.040787 | 13.07569 | 0.061163 | 0.95145 | 0.991691 | -4.60675 |
| P38646 | HSPA9 | 0.02824 | 14.21459 | 0.059471 | 0.952791 | 0.991691 | -4.60675 |
| Q9P2D7-8 | #N/D | 0.04925 | 8.669631 | 0.057298 | 0.954514 | 0.991691 | -4.60676 |
| P51149 | RAB7A | -0.02588 | 8.780581 | -0.05232 | 0.958473 | 0.991691 | -4.60676 |
| P06748 | NPM1 | -0.05603 | 9.025254 | -0.05137 | 0.959278 | 0.991691 | -4.60388 |
| P36957 | DLST | 0.035947 | 8.614293 | 0.049384 | 0.960835 | 0.991691 | -4.60677 |

| | | | | | | | |
|---|---|---|---|---|---|---|---|
| P61626 | LYZ | 0.036386 | 13.1144 | 0.045868 | 0.963581 | 0.991691 | -4.60677 |
| P02042 | HBD | 0.02707 | 10.61689 | 0.045481 | 0.963888 | 0.991691 | -4.60677 |
| O75874 | IDH1 | 0.04192 | 10.43067 | 0.044724 | 0.964488 | 0.991691 | -4.60677 |
| P05546 | SERPIND1 | 0.029031 | 11.79322 | 0.044707 | 0.964502 | 0.991691 | -4.60677 |
| Q9BW30 | TPPP3 | -0.02597 | 7.409834 | -0.04363 | 0.965455 | 0.991691 | -4.60181 |
| P61604 | HSPE1 | -0.02784 | 11.60591 | -0.03929 | 0.968802 | 0.991691 | -4.60678 |
| Q16629 | SRSF7 | 0.032145 | 9.577583 | 0.039096 | 0.968965 | 0.991691 | -4.60678 |
| O95445 | APOM | 0.039848 | 11.61754 | 0.03692 | 0.970682 | 0.991691 | -4.60678 |
| O94760 | DDAH1 | -0.02449 | 9.840467 | -0.03653 | 0.970997 | 0.991691 | -4.60678 |
| P49773 | HINT1 | 0.025846 | 8.488339 | 0.036204 | 0.971257 | 0.991691 | -4.6055 |
| P09012 | SNRPA | 0.032757 | 9.436966 | 0.0359 | 0.971523 | 0.991691 | -4.6055 |
| P84103 | SRSF3 | -0.02155 | 7.821272 | -0.03581 | 0.971593 | 0.991691 | -4.6055 |
| O94856 | NFASC | 0.021023 | 13.27301 | 0.033757 | 0.973192 | 0.991691 | -4.60678 |
| P04181 | OAT | 0.023272 | 9.027395 | 0.032369 | 0.974298 | 0.991691 | -4.60678 |
| O75891 | ALDH1L1 | -0.02445 | 12.21429 | -0.03225 | 0.974385 | 0.991691 | -4.60678 |
| P15814 | IGLL1 | -0.04621 | 11.9045 | -0.03199 | 0.974603 | 0.991691 | -4.6055 |
| P46779 | RPL28 | 0.022551 | 9.65877 | 0.029243 | 0.976778 | 0.991691 | -4.60679 |
| P62273 | RPS29 | 0.022163 | 8.012306 | 0.028923 | 0.977047 | 0.991691 | -4.60447 |
| A0A075B6I | #N/D | 0.029585 | 11.3767 | 0.028568 | 0.977317 | 0.991691 | -4.60679 |
| P16949 | STMN1 | -0.02068 | 11.41839 | -0.02825 | 0.977564 | 0.991691 | -4.60679 |
| P09874 | PARP1 | -0.01228 | 12.84743 | -0.02733 | 0.978291 | 0.991691 | -4.60679 |
| A0A0C4DH | #N/D | -0.02976 | 10.33687 | -0.02732 | 0.978304 | 0.991691 | -4.60679 |
| Q04917 | YWHAH | -0.01931 | 11.45903 | -0.02634 | 0.979082 | 0.991691 | -4.60679 |
| P10606 | COX5B | 0.019788 | 8.344202 | 0.023299 | 0.981505 | 0.991691 | -4.60551 |
| P46776 | RPL27A | 0.018955 | 10.85159 | 0.023005 | 0.981731 | 0.991691 | -4.60679 |
| Q9Y2W1 | THRAP3 | 0.014362 | 8.282812 | 0.022648 | 0.982019 | 0.991691 | -4.60679 |
| Q15149 | PLEC | 0.009696 | 12.56068 | 0.021956 | 0.982562 | 0.991691 | -4.60679 |
| A0A0C4DH | #N/D | -0.01152 | 11.05438 | -0.01426 | 0.988675 | 0.996703 | -4.60679 |
| P06310 | #N/D | -0.00597 | 13.49283 | -0.00888 | 0.992948 | 0.999083 | -4.6068 |
| P31146 | CORO1A | 0.006295 | 9.870592 | 0.007706 | 0.993881 | 0.999083 | -4.6068 |
| P13611 | VCAN | 0.001908 | 12.41909 | 0.005048 | 0.99599 | 0.999083 | -4.6068 |

| | | | | | | | |
|---|---|---|---|---|---|---|---|
| Q02252 | ALDH6A1 | 0.006136 | 13.39322 | 0.004536 | 0.996397 | 0.999083 | -4.6068 |
| Q9UJU6 | DBNL | 0.00277 | 10.24301 | 0.00391 | 0.996895 | 0.999083 | -4.6068 |
| P10720 | PF4V1 | 0.001813 | 11.14838 | 0.002138 | 0.998302 | 0.999083 | -4.6068 |
| P0DMV9 | HSPA1A | -0.00051 | 12.81602 | -0.00115 | 0.999083 | 0.999083 | -4.6068 |
| P63162 | SNRPN | #N/D | 6.935315 | #N/D | #N/D | #N/D | #N/D |
| Q13153 | PAK1 | #N/D | 7.407468 | #N/D | #N/D | #N/D | #N/D |

**Proteomic analysis of blood neuronal and glial extracellular vesicles reveals neuroprotective effects of the angiotensin type-1 blocker candesartan in Parkinson's disease patients**

Camacho-Meño L[1,†], Labandeira CM[2,†], Bravo SB[3], Torres M.V[1], Bejr-Kasem H[4,5,6,7,8], Molina-Crespo A[7,9], Atienza M[7,9], Lanciego JL[7,10,11], Cantero JL[7,9], Kulisevsky J[4,5,6,7], Labandeira-García JL[1,7,12] *, Rodríguez-Perez AI[1,7,12] *

### Supplementary Methods

### Supplementary Methods

#### Protein identification and quantification of serum extracellular vesicles by LC-MS/MS using DIA-SWATH.

Once the extracellular vesicles were isolated, the samples were mixed 1:1 with a lysis buffer containing 4% SDS, 100 mM Tris (pH 7.6), and 0.1 mM DTT. The mixture was then incubated at 95 °C for 5 minutes to ensure complete protein denaturation [1]. After this, proteins were loaded onto a 10% SDS-PAGE gel, and electrophoresis was stopped once the dye front had migrated approximately 3 mm into the resolving gel. Protein visualization was performed using Sypro Ruby fluorescent staining (Lonza, Switzerland). The stained protein band was then excised and subjected to in-gel tryptic digestion following a previously described manual protocol [2-5]. Peptides were extracted by carrying out three 20-min incubations in 40 µL of 60% acetonitrile dissolved in 0.5% HCOOH. The resulting peptide extracts were pooled, concentrated in a SpeedVac, and stored at −20 °C.

#### Mass spectrometric analysis for the SWATH spectral library creation.

3µl of digested peptides of all individual samples, per each condition, were used to create one pool per sample type (L1CAM [nEVs]; GLAST [aEVs];, MOG [oEVs]; TMEN [m/mEVs]) and separated using Reverse Phase Chromatography. Gradient was created using a micro liquid chromatography system (Eksigent Technologies nanoLC 400, SCIEX) coupled to high speed Triple TOF 6600 mass spectrometer (SCIEX) with a micro flow source. The chosen analytical column was a silica-based reversed phase column Chrom XP C18 150 × 0.30 mm, 3 mm particle size and 120 Å pore size (Eksigent, SCIEX). The trap column was a YMC-TRIART C18 (YMC Technologies, Teknokroma with a 3 mm particle size and 120 Å pore size, switched on-line with the analytical column. The loading pump delivered a solution of 0.1% formic acid in water at 10 µl/min. The micro-pump generated a flow-rate of 5 µl/min and was operated under gradient elution conditions, using 0.1% formic acid in water as mobile phase A, and 0.1% formic acid in acetonitrile as mobile phase B. Peptides were separated using a 40 minutes gradient ranging from 2% to 90% mobile phase B (mobile phase A: 2% acetonitrile, 0.1% formic acid; mobile phase B: 100% acetonitrile, 0.1% formic acid).

Data acquisition was performed in a TripleTOF 6600 System (SCIEX, Foster City, CA) using a Data dependent workflow. Source and interface conditions were the following: ionspray voltage floating (ISVF) 5500 V, curtain gas (CUR) 25, collision energy (CE) 10 and ion source gas 1 (GS1) 25. Instrument was operated with Analyst TF 1.7.1 software (SCIEX, USA). Switching criteria was set to ions greater than mass to charge ratio (m/z) 350 and smaller than m/z 1400 with charge state of 2–5, mass tolerance 250ppm and an abundance threshold of more than 200 counts (cps). Former target ions were excluded for 15 s. The instrument was automatically calibrated every 4 hours using as external calibrant tryptic peptides from PepCalMix.

##### Data Analysis

After MS/MS analysis, data files were processed using ProteinPilotTM 5.0.1 software from Sciex, which uses the algorithm ParagonTM for database search and ProgroupTM for data grouping. Data were searched using a Human specific Uniprot database. A non-linear fitting method was used to control the false discovery rate, displaying only the results with a global false discovery rate of 1% or lower for both peptides and proteins [6]. The MS/MS spectra of the identified peptides were then used to generate the spectral library for SWATH peak extraction using the add-in for PeakView Software (version 2.2, Sciex) MS/MSALL with SWATH Acquisition MicroApp (version 2.0, Sciex). Only peptides with a confidence score above 99% (as obtained from Protein Pilot database search) were included in the spectral library.

##### Relative quantification by SWATH acquisition

SWATH (Sequential Window Acquisition of all Theoretical Mass Spectra) – MS acquisition was performed on a TripleTOF® 6600 LC-MS/MS system (AB SCIEX). Samples were analyzed using a data-independent acquisition (DIA) method (30 total samples). Each sample (4 µL from a mg/ml solution) was analyzed using the LC-MS equipment and LC gradient described above for building the spectral library, but instead using the SWATH-MS acquisition method. The method consisted of repeating a cycle that consisted of the acquisition of 100 TOF MS/MS scans (400 to 1500 m/z, high sensitivity mode, 50 ms acquisition time) of overlapping sequential precursor isolation windows of variable width (1 m/z overlap) covering the 400 to 1250 m/z mass range with a previous TOF MS scan (400 to 1500 m/z, 50 ms acquisition time) for each cycle. Total cycle time was 6.3 s. For each sample set, the width of the 65 variable windows was optimized according to the ion density found in the DDA runs using a SWATH variable window calculator worksheet from Sciex.

##### Data analysis

The targeted data extraction of the fragment ion chromatogram traces from the SWATH runs was performed by PeakView (version 2.2) using the SWATH Acquisition MicroApp(version 2.0). This application processed the data using the spectral library created from the shotgun data. Up to ten peptides per protein and seven fragments per peptide were selected, based on signal intensity; any shared and modified peptides were excluded from the processing. Five-minute windows and 30 ppm widths were used to extract the ion chromatograms; SWATH quantitation was attempted for all proteins in the ion library that were identified by ProteinPilot with an FDR below 1%. The retention times from the peptides that were selected for each protein were used to realign in each run according to the iRT peptides spiked in each sample and eluted along the whole-time axis. The extracted ion chromatograms were then generated for each selected fragment ion; the peak areas for the peptides were obtained by summing the peak areas from the corresponding fragment ions. PeakView computed an FDR and a score for each assigned peptide according to the chromatographic and spectra components; only peptides with an FDR below 5% were used for protein quantitation. Protein quantitation was calculated by adding the peak areas of the corresponding peptides.

The integrated peak areas (processed. mrkvw files from PeakView) were directly exported to the MarkerView software (AB SCIEX) for relative quantitative analysis. The export will generate three files containing quantitative information about individual ions, the summed intensity of different ions for a particular peptide and the summed intensity of different peptides for a particular protein. MarkerView has been used for analysis of SWATH-MS data reported in other proteomics studies [7-10] because of its data-independent method of quantitation. MarkerView uses processing algorithms that accurately find chromatographic and spectral peaks direct from the raw SWATH data. Data alignment by MarkerView compensates for minor variations in both mass and retention time values, ensuring that identical compounds in different samples are accurately compared to one another. To control for possible uneven sample loss across the different samples during the sample preparation process, we performed a global normalization based on the total sum of all the peak areas extracted from all the peptides and transitions across the replicates of each sample. Unsupervised multivariate statistical analysis using principal component analysis (PCA) was performed to compare the data across

**SWATH Data Analysis**

R (version 4.1.2) was used to perform all bioinformatic analyses. The data was log-transformed, quantile normalized, and differentially enriched proteins were identified using using linear models (limma, version 3.50.3). Proteins were considered differentially enriched if $P < 0.05$ and FC $\geq$1.5 or <0.6. Heatmaps and volcano plots were generated using heatmap3 (version 1.1.9), GraphPad Prism 8 (GraphPad, Inc., San Diego, CA, USA), and Glimma (version 2.4.0) packages. Functional annotation was performed using clusterProfiler (version 4.2.2) and rrvgo (version 1.6.0) packages. Identified proteins were analyzed using STRING v[12.0] (https://string-db.org) to evaluate predicted protein–protein interactions. The analysis was performed for Homo sapiens with a minimum required interaction score of 0.7 (high confidence). Gene Ontology (GO) enrichment analysis was conducted within STRING. Interaction networks were visualized and exported for further interpretation.

**Proteomic analysis of blood neuronal and glial extracellular vesicles reveals neuroprotective effects of the angiotensin type-1 blocker candesartan in Parkinson's disease patients**

Camacho-Meño L[1,†], Labandeira CM[2,†], Bravo SB[3], Torres M.V[1], Bejr-Kasem H[4,5,6,7,8], Molina-Crespo A[7,9], Atienza M[7,9], Lanciego JL[7,10,11], Cantero JL[7,9], Kulisevsky J[4,5,6,7], Labandeira-García JL[1,7,12] *, Rodríguez-Perez AI[1,7,12] *

### Supplementary Material: Data sheet relative to Figure 1

#### Data Sheet and statistical analysis of Figures 1A and 1B

| | Mean | Moda | Concentration |
|---|---|---|---|
| Pre-CAND_ 1 | 120 | 77.5 | 4.32E+12 |
| Pre-CAND_ 2 | 119 | 72.5 | 4.36E+12 |
| Pre-CAND_ 3 | 117 | 77.5 | 3.43E+12 |
| Pre-CAND_ 4 | 110 | 67.5 | 2.99E+12 |
| Pre-CAND_ 5 | 112 | 77.5 | 3.72E+12 |
| | **115.6** | **74.5** | **3.764E+12** |

| | Mean | Moda | Concentration |
|---|---|---|---|
| Post-CAND_ 1 | 124 | 87.5 | 4.49E+12 |
| Post-CAND_ 2 | 122 | 87.5 | 3.62E+12 |
| Post-CAND_ 3 | 114 | 77.5 | 3.28E+12 |
| Post-CAND_ 4 | 123 | 72.5 | 2.76E+12 |
| Post-CAND_ 5 | 113 | 87.5 | 3.84E+12 |
| | **119.2** | **82.5** | **3.598E+12** |

**Mean**

Normality Tes Failed (P < 0,050)

Test execution ended by user request, Rank Sum Test begun

Mann-Whitney Rank Sum Test

Data source: Data 1 in Notebook1

| Group | N | Missing | Median | 25% | 75% |
|---|---|---|---|---|---|
| media | 5 | 0 | 117 | 111.50 | 119 |
| post-media | 5 | 0 | 122 | 113.75 | 123 |

Mann-Whitney U Statistic= 6,000

T = 21,000 n(small)= 5 n(big)= 5 P(est.)= 0,210 P(exact)= 0,222

The difference in the median values between the two groups is not great enough to exclude the possibility that the difference is due to random sampling variability; there is not a statistically significant difference (P = 0,222)

**Mode**

Normality Tes Failed (P < 0,050)

Test execution ended by user request, Rank Sum Test begun

Mann-Whitney Rank Sum Test

Data source: Data 1 in Notebook1

| Group | N | Missing | Median | 25% | 75% |
|---|---|---|---|---|---|
| moda | 5 | 0 | 77.5 | 71.25 | 77.5 |
| post-moda | 5 | 0 | 87.5 | 76.25 | 87.5 |

Mann-Whitney U Statistic= 5,000

T = 20,000 n(small)= 5 n(big)= 5 P(est.)= 0,125 P(exact)= 0,151

The difference in the median values between the two groups is not great enough to exclude the possibility that the difference is due to random sampling variability; there is not a statistically significant difference (P = 0,151)

**Concentration**

Normality Tes Passed (P = 0.766)

Equal Varianc Passed (P = 0.971)

| Group Name | N | Missing | Mean | Std Dev | SEM |
|---|---|---|---|---|---|

| | | | | | |
|---|---|---|---|---|---|
| concentració | 5 | 0 | 3.764E+12 | 5.8671E+11 | 2.62385E+11 |
| post-concent | 5 | 0 | 3.598E+12 | 6.4383E+11 | 2.87931E+11 |

Difference ################

t = 0,426 with 8 degrees of freedom. (P = 0,681)

95 percent confidence interval for difference of means: -732306271373,083 to 1,064E+012

The difference in the mean values of the two groups is not great enough to reject the possibility that the difference is due to random sampling variability. There is not a statistically significant difference between the input groups (P = 0,681).

#### Data Sheet and statistical analysis of Figure 1D

| | | | |
|---|---|---|---|
| Pre-CAND_ 1 | 0.1721 | Post-CAND_ 1 | 0.1545 |
| Pre-CAND_ 2 | 0.177 | Post-CAND_ 2 | 0.1946 |
| Pre-CAND_ 3 | 0.2075 | Post-CAND_ 3 | 0.1732 |

Normality Test: Passed (P = 0.296)
Equal Variance T Passed (P = 0.873)

| Group Name | N | Missing | Mean | Std Dev | SEM |
|---|---|---|---|---|---|
| cd81_pre | 3 | 0 | 0 | 0.0192 | 0.0111 |
| cd81_post | 3 | 0 | 0.174 | 0.0201 | 0.0116 |

Difference 0.0115
t = 0.714 with 4 degrees of freedom. (P = 0.515)
95 percent confidence interval for difference of means: -0,0331 to 0,0560
The difference in the mean values of the two groups is not great enough to reject the possibility that the difference is due to random sampling variability. There is not a statistically significant difference between the input groups (P = 0,515).

| | | | |
|---|---|---|---|
| Pre-CAND_ 1 | 0.172 | Pre-CAND_D1 | 0.0188 |
| Pre-CAND_ 2 | 0.177 | Pre-CAND_D2 | 0.021 |
| Pre-CAND_ 3 | 0.208 | Pre-CAND_D3 | 0.0265 |
| Post-CAND_ 1 | 0.1545 | Post-CAND_ D1 | 0.0196 |
| Post-CAND_ 2 | 0.1946 | Post-CAND_ D2 | 0.0231 |
| Post-CAND_ 3 | 0.1732 | Post-CAND_ D3 | 0.0199 |

Normality Test: Passed (P = 0.189)
Equal Variance T Passed (P = 0.061)

| Group Name | N | Missing | Mean | Std Dev | SEM |
|---|---|---|---|---|---|
| DEPLETED | 6 | 0 | 0.0215 | 0.00287 | 0.00117 |
| EVs_CD81 | 6 | 0 | 0.18 | 0.0187 | 0.00762 |

Difference -0.158
t = -20.551 with 10 degrees of freedom. (P = <0.001)
95 percent confidence interval for difference of means: -0,176 to -0,141
The difference in the mean values of the two groups is greater than would be expected by chance; there is a statistically significant difference between the input groups (P = <0,001).

#### Data Sheet and statistical analysis of Figure 1E

| | | | |
|---|---|---|---|
| Pre-CAND_ 1 | 6.577 | Post-CAND_ 1 | 4.730 |
| Pre-CAND_ 2 | 4.316 | Post-CAND_ 2 | 5.404 |
| Pre-CAND_ 3 | 3.290 | Post-CAND_ 3 | 5.666 |

Normality Test: Passed (P = 0.821)
Equal Variance Test Passed (P = 0.213)

| Group Name | N | Missing | Mean | Std Dev | SEM |
|---|---|---|---|---|---|
| cd63-pre cand | 3 | 0 | 4.7280 | 1.682 | 0.971 |
| cd63-post cand | 3 | 0 | 5.2660 | 0.483 | 0.279 |

Difference -0.539

t = -0.533 with 4 degrees of freedom. (P = 0.622)

95 percent confidence interval for difference of means: -3.343 to 2.266

The difference in the mean values of the two groups is not great enough to reject the possibility that the difference is due to random sampling variability. There is not a statistically significant difference between the input groups (P = 0,622).

| | | | |
|---|---|---|---|
| Pre-CAND_ 1 | 6.577 | Pre-CAND_D1 | 0.0242 |
| Pre-CAND_ 2 | 4.316 | Pre-CAND_D2 | 0.0265 |
| Pre-CAND_ 3 | 3.290 | Pre-CAND_D3 | 0.0179 |
| Post-CAND_ 1 | 4.730 | Post-CAND_ D1 | 0.0166 |
| Post-CAND_ 2 | 5.404 | Post-CAND_ D2 | 0.0197 |
| Post-CAND_ 3 | 5.666 | Post-CAND_ D3 | 0.0238 |

Normality Test: Passed (P = 0.100)

Equal Variance Test Failed (P < 0.050)

Test execution ended by user request, Rank Sum Test begun

Mann-Whitney Rank Sum Test

| Group | N | Missing | Median | 25% | 75% |
|---|---|---|---|---|---|
| depleted | 6.000 | 0.000 | 0.022 | 0.018 | 0.024 |
| EVs_CD63 | 6.000 | 0.000 | 5.067 | 4.316 | 5.666 |

Mann-Whitney U Statistic= 0,000

T = 21.000 n(small)= 6 n(big)= 6 P(est.)= 0.005 P(exact)= 0.002

The difference in the median values between the two groups is greater than would be expected by chance; there is a statistically significant difference (P = 0,002)

#### Data Sheet and statistical analysis of Figure 1F

| | |
|---|---|
| Pre-CAND_ 2 | 0.2004 |
| Pre-CAND_3 | 0.3485 |
| Pre-CAND_ 4 | 0. 1075 |

| | |
|---|---|
| Post-CAND_2 | 0.2298 |
| Post-CAND_ 3 | 0.1734 |
| Post-CAND_ 4 | 0.1501 |

| | | |
|---|---|---|
| Normality Test: | Passed | (P = 0.839) |
| Equal Variance Te: | Passed | (P = 0.129) |

| Group Name | N | Missing | Mean | Std Dev | SEM |
|---|---|---|---|---|---|
| ALIX-pre cand | 3.0000 | 0 | 0.219 | 0.122 | 0.0702 |
| ALIX-post cand | 3.0000 | 0 | 0.184 | 0.041 | 0.0237 |
| Difference | 0,0344 | | | | |

t = 0,464 with 4 degrees of freedom. (P = 0,667)

95 percent confidence interval fc Dio

The difference in the mean values of the two groups is not great enough to reject the possibility that the difference is due to random sampling variability. There is not a statistically significant difference between the input groups (P = 0,667).

| | | | |
|---|---|---|---|
| Pre-CAND_ 2 | 0.2004 | Pre-CAND_ D2 | 0.00061 |
| Pre-CAND_3 | 0.3485 | Pre-CAND_ D3 | 0.0006681 |
| Pre-CAND_ 4 | 0.1075 | Pre-CAND_ D4 | 0.00045255 |
| Post-CAND_2 | 0.2298 | Post-CAND_D2 | 0.00041892 |
| Post-CAND_ 3 | 0.1734 | Post-CAND_D3 | 0.00049799 |
| Post-CAND_ 4 | 0.1501 | Post-CAND_D4 | 0.00060078 |

Normality Test: Failed (P < 0,050)

Mann-Whitney Rank Sum Test

Data source: Data 2 in estadística de caracterización origen EVs.JNB

| Group | N | Missing | Median | 25% | 75% |
|---|---|---|---|---|---|
| depeted alix | 6 | 0 | 0,000549 | 0,000453 | 0,000610 |
| evs alix | 6 | 0 | 0,187 | 0,150 | 0,230 |

Mann-Whitney U Statistic= 0,000

T = 21,000 n(small)= 6 n(big)= 6 P(est.)= 0,005 P(exact)= 0,002

The difference in the median values between the two groups is greater than would be expected by chance; there is a statistically significant difference (P = 0,002)

#### Data Sheet and statistical analysis of Figure 1G

| | |
|---|---|
| Pre-CAND_ 3 | 1.1319 |
| Pre-CAND_ 4 | 0.8874 |
| Pre-CAND_ 5 | 0.7564 |

| | |
|---|---|
| Post-CAND_3 | 1.1664 |
| Post-CAND_4 | 1.0434 |
| Post-CAND_5 | 1.271 |

Normality Test: Passed (P = 0,890)

Equal Variance Te Passed (P = 0,327)

| Group Name | N | Missing | Mean | Std Dev | SEM |
|---|---|---|---|---|---|
| calnexin_pre_car | 3 | 0 | 0.925 | 0.191 | 0.11 |
| calnexin_post | 3 | 0 | 1 | 0.114 | 0.0658 |

Difference -0.235

t = -1.833 with 4 degrees of freedom. (P = 0.141)

95 percent confidence interval for difference of means: -0.591 to 0.121

The difference in the mean values of the two groups is not great enough to reject the possibility that the difference is due to random sampling variability. There is not a statistically significant difference between the input groups (P = 0.141).

| | Depleted | EVT | | Control+: HCM3 |
|---|---|---|---|---|
| Pre-CAND_ 3 | 1.255 | 1.1319 | Sample 1 | 36.0444 |
| Pre-CAND_ 4 | 1.4998 | 0.8874 | Sample 2 | 26.811 |
| Pre-CAND_ 5 | 0.9612 | 0.7564 | Sample 3 | 20.8821 |
| Post-CAND_3 | 0.8206 | 1.1664 | Sample 4 | 25.4723 |
| Post-CAND_4 | 1.0577 | 1.0434 | Sample 5 | 27.5236 |
| Post-CAND_5 | 1.0963 | 1.271 | Sample 6 | 18.1128 |

Normality Test: Failed (P < 0.050)

Test execution ended by user request, ANOVA on Ranks begun

Kruskal-Wallis One Way Analysis of Variance on Ranks

| Group | N | Missing | Median | 25% | 75% |
|---|---|---|---|---|---|
| Depleted | 6 | 0 | 1.077 | 0.961 | 1.255 |
| EVt | 6 | 0 | 1.088 | 0.887 | 1.166 |
| control + | 6 | 0 | 26.142 | 20.882 | 27.524 |

H = 11.415 with 2 degrees of freedom. (P = 0.003)

The differences in the median values among the treatment groups are greater than would be expected by chance; there is a statistically significant difference (P = 0,003)

To isolate the group or groups that differ from the others use a multiple comparison procedure.

All Pairwise Multiple Comparison Procedures (Student-Newman-Keuls Method) :

| Comparison | Diff of Ranks | q | P<0.05 |
|---|---|---|---|
| control + vs EVt | 56.000 | 4.282 | Yes |
| control + vs Depl | 52.000 | 5.888 | Yes |
| Depleted vs EVt | 4.000 | 0.453 | No |

#### Data Sheet and statistical analysis of Figure 1H

| | |
|---|---|
| Pre-CAND_ 5 | 1724.93 |
| Pre-CAND_ 1 | 983.4788 |
| Pre-CAND_ 2 | 1225.935 |

| | |
|---|---|
| Post-CAND_5 | 1074.756 |
| Post-CAND_1 | 1902.462 |
| Post-CAND_2 | 1263.14 |

Normality Test: Passed (P = 0,131)
Equal Variance Test Passed (P = 0,883)

| Group Name | N | Missing | Mean | Std Dev | SEM |
|---|---|---|---|---|---|
| enolase_precand | 3 | 0 | 1.311.448 | 378.050 | 218.267 |
| enolase-postcand | 3 | 0 | 1.413.453 | 433.843 | 250.479 |

Difference -102.005

t = -0.307 with 4 degrees of freedom. (P = 0.774)

95 percent confidence interval for difference of means: -1024.439 to 820.429

The difference in the mean values of the two groups is not great enough to reject the possibility that the difference is due to random sampling variability. There is not a statistically significant difference between the input groups (P = 0.774).

| | serum | $EV_T$ | nEV |
|---|---|---|---|
| Pre-CAND_ 5 | 0.003419 | 4.2694 | 1724.93 |
| Pre-CAND_ 1 | 0.000201 | 7.6083 | 983.4788 |
| Pre-CAND_ 2 | 0.002342 | 10.2906 | 1225.935 |
| Post-CAND_5 | 0.003526 | 8.9565 | 1074.756 |
| Post-CAND_1 | 0.001357 | 5.0016 | 1902.462 |
| Post-CAND_2 | 0.006681 | 1.6904 | 1263.14 |

Normality Test: Failed (P < 0,050)
Test execution ended by user request, ANOVA on Ranks begun

| Group | N | Missing | Median | 25% | 75% |
|---|---|---|---|---|---|
| ENOLASE SUERO | 6 | 0 | 0.00288 | 0.00136 | 0.00353 |
| ENOLASE EVT | 6 | 0 | 6 | 4 | 9 |
| ENOLASE nEVs | 6 | 0 | 1.245 | 1.075 | 1.725 |

H = 15.158 with 2 degrees of freedom. (P = <0.001)

The differences in the median values among the treatment groups are greater than would be expected by chance; there is a statistically significant difference (P = <0.001)

To isolate the group or groups that differ from the others use a multiple comparison procedure.

All Pairwise Multiple Comparison Procedures (Student-Newman-Keuls Method) :

| Comparison | Diff of Ranks | q | P<0.05 |
| --- | --- | --- | --- |
| ENOLASE nEVs vs E | 72.000 | 5.506 | Yes |
| ENOLASE nEVs vs E | 36.000 | 4.076 | Yes |
| ENOLASE EVT vs EN | 36.000 | 4.076 | Yes |

#### Data Sheet and statistical analysis of Figure 1I

| | |
|---|---|
| Pre-CAND_ 4 | 40.7908 |
| Pre-CAND_ 5 | 50.1429 |
| Pre-CAND_ 1 | 61.2482 |

| | |
|---|---|
| Post-CAND_4 | 56.9835 |
| Post-CAND_5 | 64.9833 |
| Post-CAND_1 | 37.6855 |

Normality Test: Passed (P = 0,571)
Equal Variance Te: Passed (P = 0,610)

| Group Name | N | Missing | Mean | Std Dev | SEM |
|---|---|---|---|---|---|
| glast pre-cand | 3 | 0 | 50.727 | 10.241 | 5.913 |
| glast post-cand | 3 | 0 | 53.217 | 14.033 | 8.102 |

Difference -2.490
t = -0,248 with 4 degrees of freedom. (P = 0,816)

95 percent confidence interval for difference of means: -30,338 to 25,358

The difference in the mean values of the two groups is not great enough to reject the possibility that the difference is due to random sampling variability. There is not a statistically significant difference between the input groups (P = 0,816).

| | serum | EVT | nEV |
|---|---|---|---|
| Pre-CAND_ 4 | 0.1061 | 0.22294887 | 40.7908 |
| Pre-CAND_ 5 | 0.141 | 0.16420361 | 50.1429 |
| Pre-CAND_ 1 | 0.144 | 0.23908193 | 61.2482 |
| Post-CAND_4 | 0.1424 | 0.21249469 | 56.9835 |
| Post-CAND_5 | 0.1364 | 0.1786565 | 64.9833 |
| Post-CAND_1 | 0.1365 | 0.22967386 | 37.6855 |

Normality Test: Failed (P < 0,050)
Test execution ended by user request, ANOVA on Ranks begun
Kruskal-Wallis One Way Analysis of Variance on Ranks

Data source: Data 4 in estadística de caracterización origen EVs

| Group | N | Missing | Median | 25% | 75% |
|---|---|---|---|---|---|
| GLAST Suero | 6 | 0 | 0.139 | 0.136 | 0.142 |
| Glast EVt | 6 | 0 | 0.218 | 0.179 | 0.230 |
| Glast nEVs | 6 | 0 | 53.563 | 40.791 | 61.248 |

legrees of freedom. (P = <0.001)

The differences in the median values among the treatment groups are greater than would be expected by chance; there is a statistically significant difference (P = <0.001)

To isolate the group or groups that differ from the others use a multiple comparison procedure.

All Pairwise Multiple Comparison Procedures (Student-Newman-Keuls Method) :

| Comparison | Diff of Ranks | q | P<0.05 |
|---|---|---|---|

| | | | |
|---|---|---|---|
| Glast nEVs vs GLA | 72.000 | 5.506 | Yes |
| Glast nEVs vs Glas | 36.000 | 4.076 | Yes |
| Glast EVt vs GLAS | 36.000 | 4.076 | Yes |

#### Data Sheet and statistical analysis of Figure 1J

| | |
|---|---|
| Pre-CAND_ 3 | 106.5017 |
| Pre-CAND_ 4 | 114.938 |
| Pre-CAND_ 5 | 114.5253 |

| | |
|---|---|
| Post-CAND_3 | 153.5087 |
| Post-CAND_4 | 100.9321 |
| Post-CAND_5 | 82.8059 |

Normality Test: Passed (P = 0.503)
Equal Variance Te Passed (P = 0.220)

| Group Name | N | Missing | Mean | Std Dev | SEM |
|---|---|---|---|---|---|
| mbp pre-cand | 3 | 0 | 111.988 | 4.756 | 2.746 |
| mbp post-cand | 3 | 0 | 112.416 | 36.724 | 21.202 |
| Difference | -0.427 | | | | |

t = -0.0200 with 4 degrees of freedom. (P = 0.985)
95 percent confidence interval for difference of means: -59.786 to 58.932
The difference in the mean values of the two groups is not great enough to reject the possibility that the difference is due to random sampling variability. There is not a statistically significant difference between the input groups (P = 0.985).

| | serum | $EV_T$ | nEV |
|---|---|---|---|
| Pre-CAND_ 3 | 0.0058804 | 0.1579 | 106.5017 |
| Pre-CAND_ 4 | 0.0068139 | 0.0587 | 114.938 |
| Pre-CAND_5 | 0.0045664 | 0.1482 | 114.5253 |
| Post-CAND_3 | 0.0048622 | 0.087 | 153.5087 |
| Post-CAND_4 | 0.0048017 | 0.0641 | 100.9321 |
| Post-CAND_5 | 0.0051574 | 0.0931 | 82.8059 |

Normality Test: Failed (P < 0.050)
Test execution ended by user request. ANOVA on Ranks begun
Kruskal-Wallis One Way Analysis of Variance on Ranks

| Group | N | Missing | Median | 25% | 75% |
|---|---|---|---|---|---|
| mbp suero | 6 | 0 | 0.00501 | 0.0048 | 0.00588 |
| mbp EVt | 6 | 0 | 0.0901 | 0.0641 | 0.148 |
| MBP nEVs | 6 | 0.000 | 110.513 | 100.932 | 114.938 |

H = 15.158 with 2 degrees of freedom. (P = <0.001)
The differences in the median values among the treatment groups are greater than would be expected by chance; there is a statistically significant difference (P = <0.001)
To isolate the group or groups that differ from the others use a multiple comparison procedure.
All Pairwise Multiple Comparison Procedures (Student-Newman-Keuls Method) :

| Comparison | Diff of Ranks | q | P<0.05 |
|---|---|---|---|
| MBP nEVs vs mbp | 72.000 | 5.506 | Yes |
| MBP nEVs vs mbp | 36.000 | 4.076 | Yes |
| mbp EVt vs mbp s | 36.000 | 4.076 | Yes |

#### Data Sheet and statistical analysis of Figure 1K

| | |
|---|---|
| Pre-CAND_ 2 | 3.11723933 |
| Pre-CAND_ 3 | 2.45301743 |
| Pre-CAND_ 4 | 2.13203992 |

| | |
|---|---|
| Post-CAND_2 | 2.66009852 |
| Post-CAND_3 | 2.32603788 |
| Post-CAND_4 | 2.64389424 |

| | | | | | |
|---|---|---|---|---|---|
| Normality Test: | Passed | (P = 0.871) | | | |
| Equal Variance Tes | Passed | (P = 0.290) | | | |
| Group Name | N | Missing | Mean | Std Dev | SEM |
| iba-1 pre-cand | 3 | 0 | 2.567 | 0.502 | 0.290 |
| iba-1 post-cand | 3 | 0 | 2.543 | 0.188 | 0.109 |
| Difference | 0.0241 | | | | |

t = 0.0778 with 4 degrees of freedom. (P = 0.942)

95 percent confidence interval for difference of means: -0.836 to 0.884

The difference in the mean values of the two groups is not great enough to reject the possibility that the difference is due to random sampling variability. There is not a statistically significant difference between the input groups (P = 0.942).

| | serum | EVT | nEV |
|---|---|---|---|
| Pre-CAND_ 2 | 0.0078062 | 0.02003309 | 3.11723933 |
| Pre-CAND_ 3 | 0.0087966 | 0.05283073 | 2.45301743 |
| Pre-CAND_ 4 | 0.0087337 | 0.12543139 | 2.13203992 |
| Post-CAND_2 | 0.0097084 | 0.24698658 | 2.66009852 |
| Post-CAND_3 | 0.0098921 | 0.03158854 | 2.32603788 |
| Post-CAND_4 | 0.01 | 0.0872095 | 2.64389424 |

Normality Test: Failed (P < 0.050)

Test execution ended by user request. ANOVA on Ranks begun

Kruskal-Wallis One Way Analysis of Variance on Ranks

| Group | N | Missing | Median | 25% | 75% |
|---|---|---|---|---|---|
| iba-1 suero | 6 | 0 | 0.00925 | 0.00873 | 0.00989 |
| iba-EVt | 6 | 0 | 0.07 | 0.0316 | 0.125 |
| iba nEVs | 6 | 0 | 3 | 2 | 3 |

H = 15.158 with 2 degrees of freedom. (P = <0.001)

The differences in the median values among the treatment groups are greater than would be expected by chance; there is a statistically significant difference (P = <0.001)

To isolate the group or groups that differ from the others use a multiple comparison procedure.

All Pairwise Multiple Comparison Procedures (Student-Newman-Keuls Method) :

| Comparison | Diff of Ranks | q | P<0.05 |
|---|---|---|---|
| iba nEVs vs iba-1 su | 72.000 | 5.506 | Yes |
| iba nEVs vs iba-EVt | 36.000 | 4.076 | Yes |
| iba-EVt vs iba-1 su | 36.000 | 4.076 | Yes |

**Proteomic analysis of blood neuronal and glial extracellular vesicles reveals neuroprotective effects of the angiotensin type-1 blocker candesartan in Parkinson's disease patients**

Camacho-Meño L[1,†], Labandeira CM[2,†], Bravo SB[3], Torres M.V[1], Bejr-Kasem H[4,5,6,7,8], Molina-Crespo A[7,9], Atienza M[7,9], Lanciego JL[7,10,11], Cantero JL[7,9], Kulisevsky J[4,5,6,7], Labandeira-García JL[1,7,12] *, Rodríguez-Perez AI[1,7,12] *

### Supplementary Figures

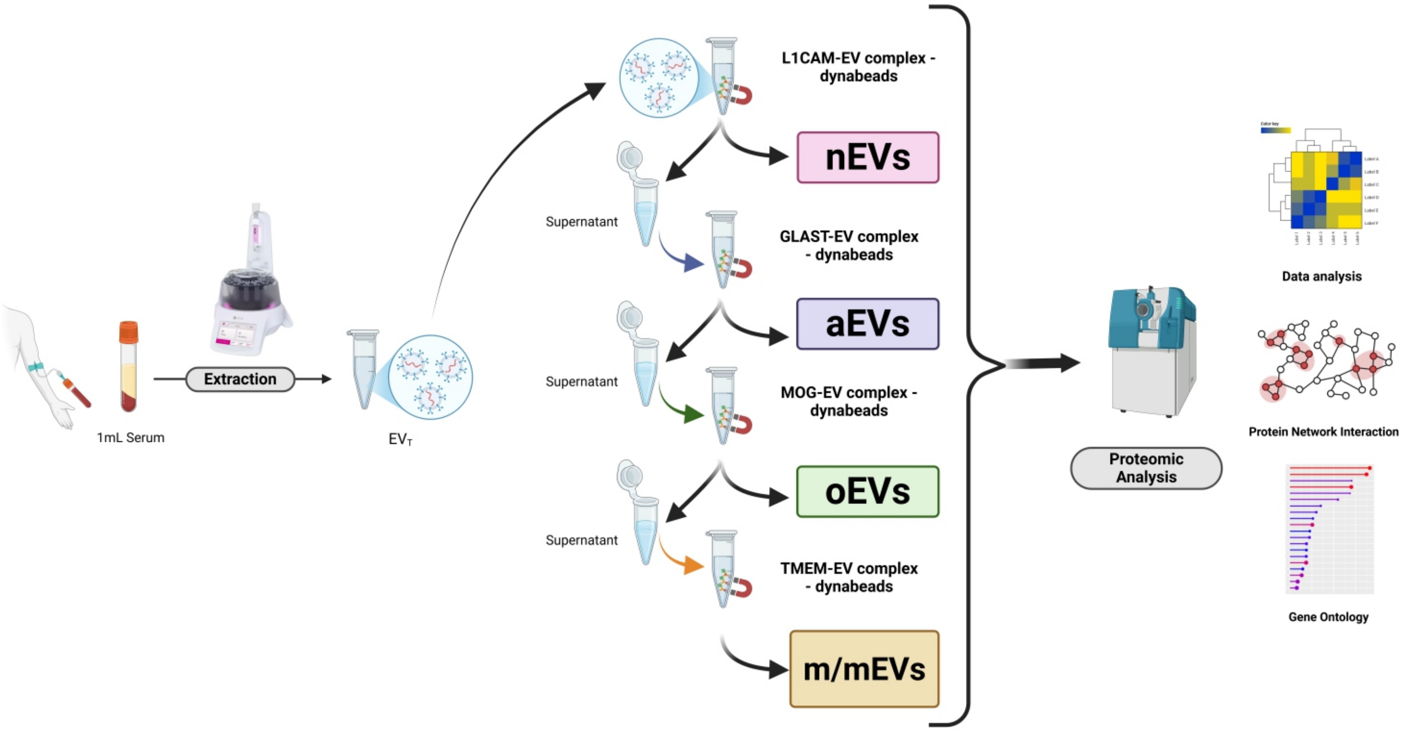


**Figure S1.** Workflow for the isolation and proteomic characterization of nervous system–derived extracellular vesicle (EV) subpopulations from human serum. From 1 mL of serum, total EVs ($EV_T$) are extracted using size-exclusion chromatography (SEC) and subsequently incubated with magnetic beads conjugated with antibodies targeting cell-type-specific surface markers: L1CAM (neuronal EVs, nEVs), GLAST (astrocytic EVs, aEVs), MOG (oligodendrocytic EVs, oEVs), and TMEM (microglial/macrophage EVs, m/mEVs). The resulting enriched EV subpopulations are subjected to mass spectrometry-based proteomic analysis, followed by bioinformatic workflows.

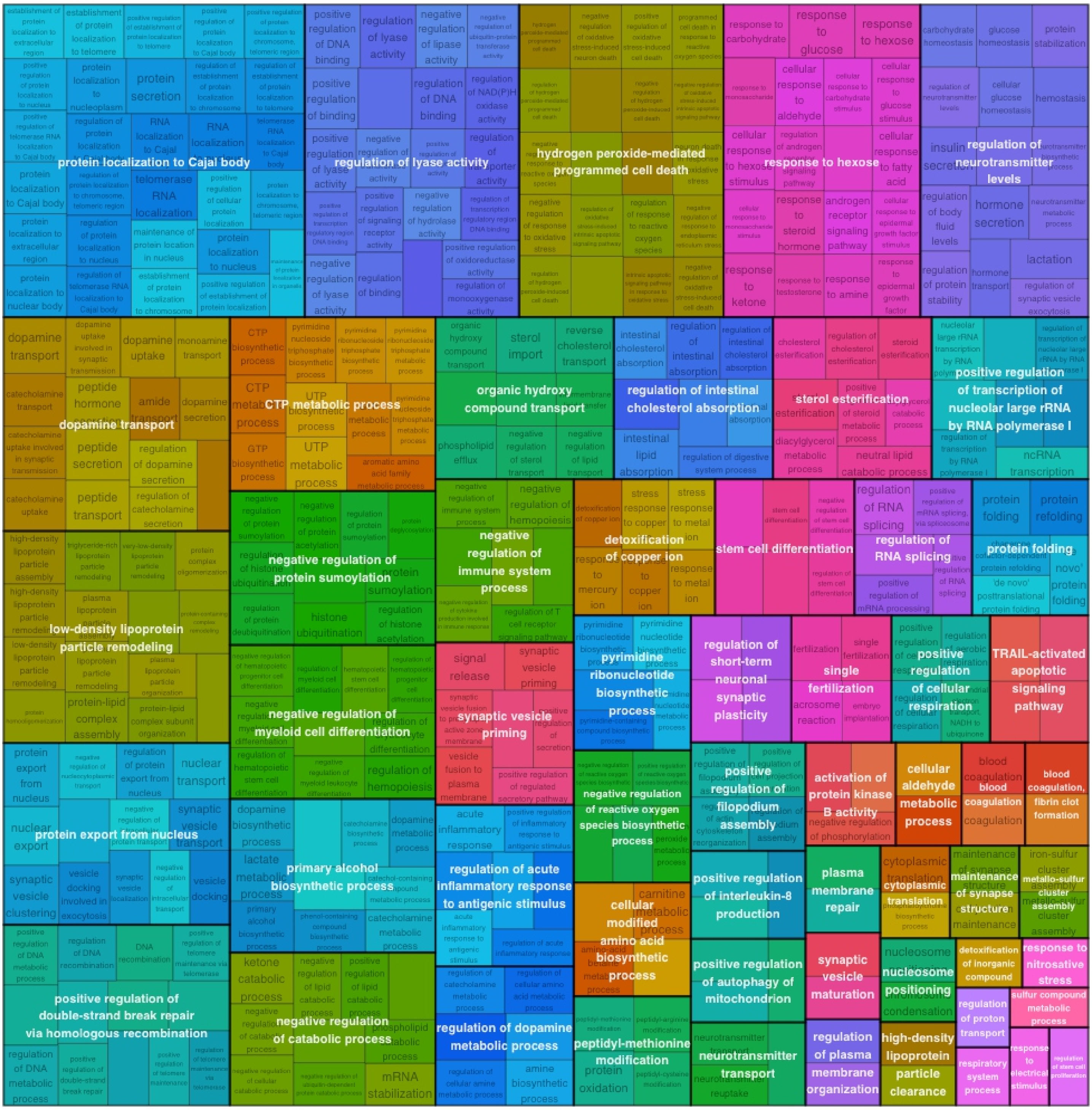


**Figure S2.** Treemap representation of enriched functional categories in upregulated genes from the comparison nEVs post-CAND *vs* nEVs pre-CAND. This treemap illustrates biological processes associated with differentially expressed genes. The size of each box corresponds to the level of enrichment of the respective functional category. Categories include immune response regulation, viral entry and life cycle modulation, cytoskeletal dynamics, neuronal development, metabolic processes, and cell differentiation. The color-coding represents distinct biological functions, grouping related processes for better visualization.

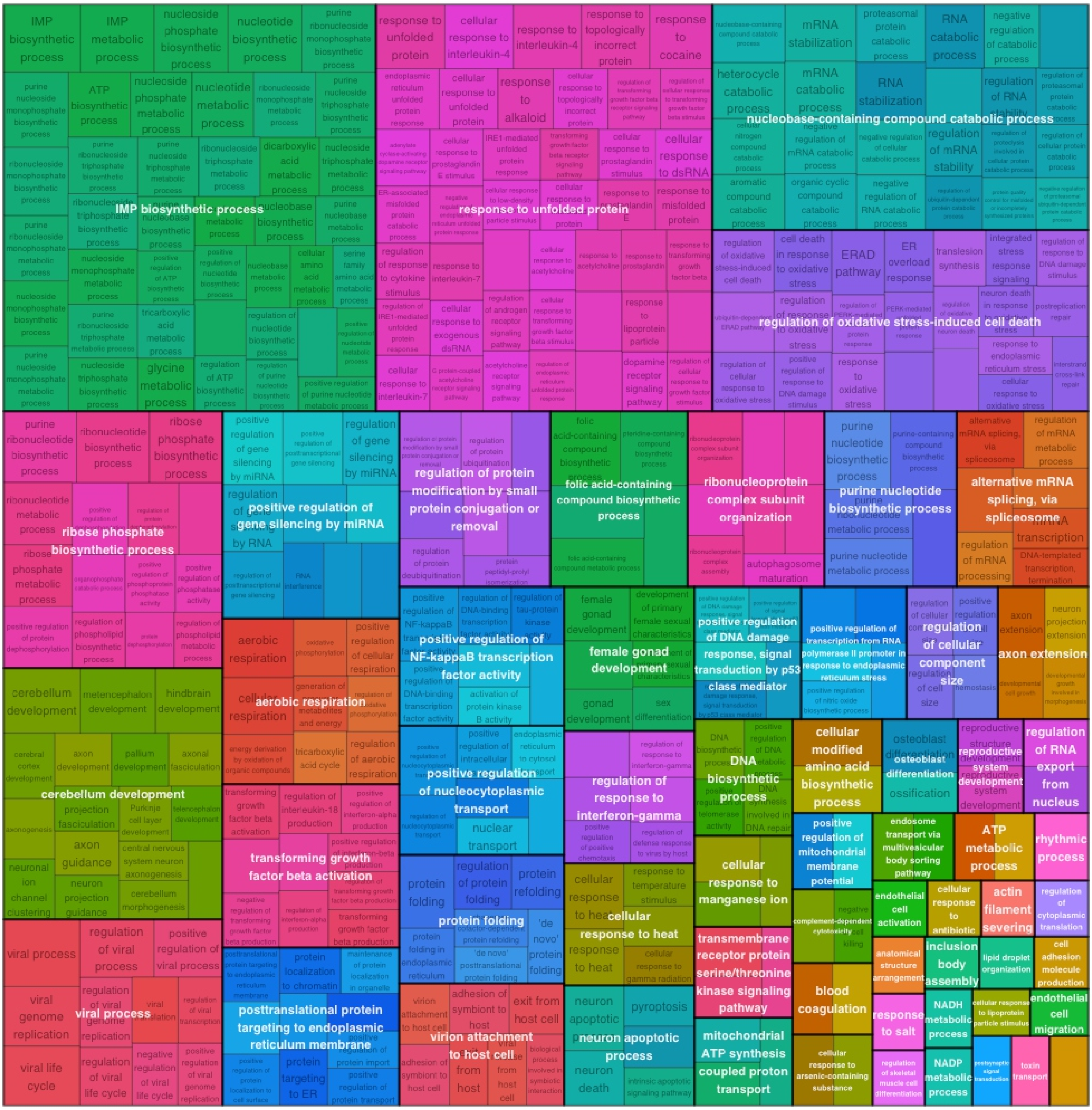


**Figure S3.** Treemap representation of enriched functional categories in downregulated genes from the comparison nEVs post-CAND *vs* nEVs pre-CAND. This treemap illustrates biological processes associated with differentially expressed genes. The size of each box corresponds to the level of enrichment of the respective functional category. The largest downregulated categories include IMP biosynthetic process, response to unfolded protein, nucleobase-containing compound catabolic process, regulation of oxidative stress-induced cell death, ribose phosphate biosynthetic process, and positive regulation of gene silencing by miRNA. The suppression of these pathways suggests a reduction in cellular stress, metabolic overload, neuroinflammation, and proteotoxicity, all of which are key contributors to PD progression. The color-coding represents distinct biological functions, grouping related processes for better visualization.

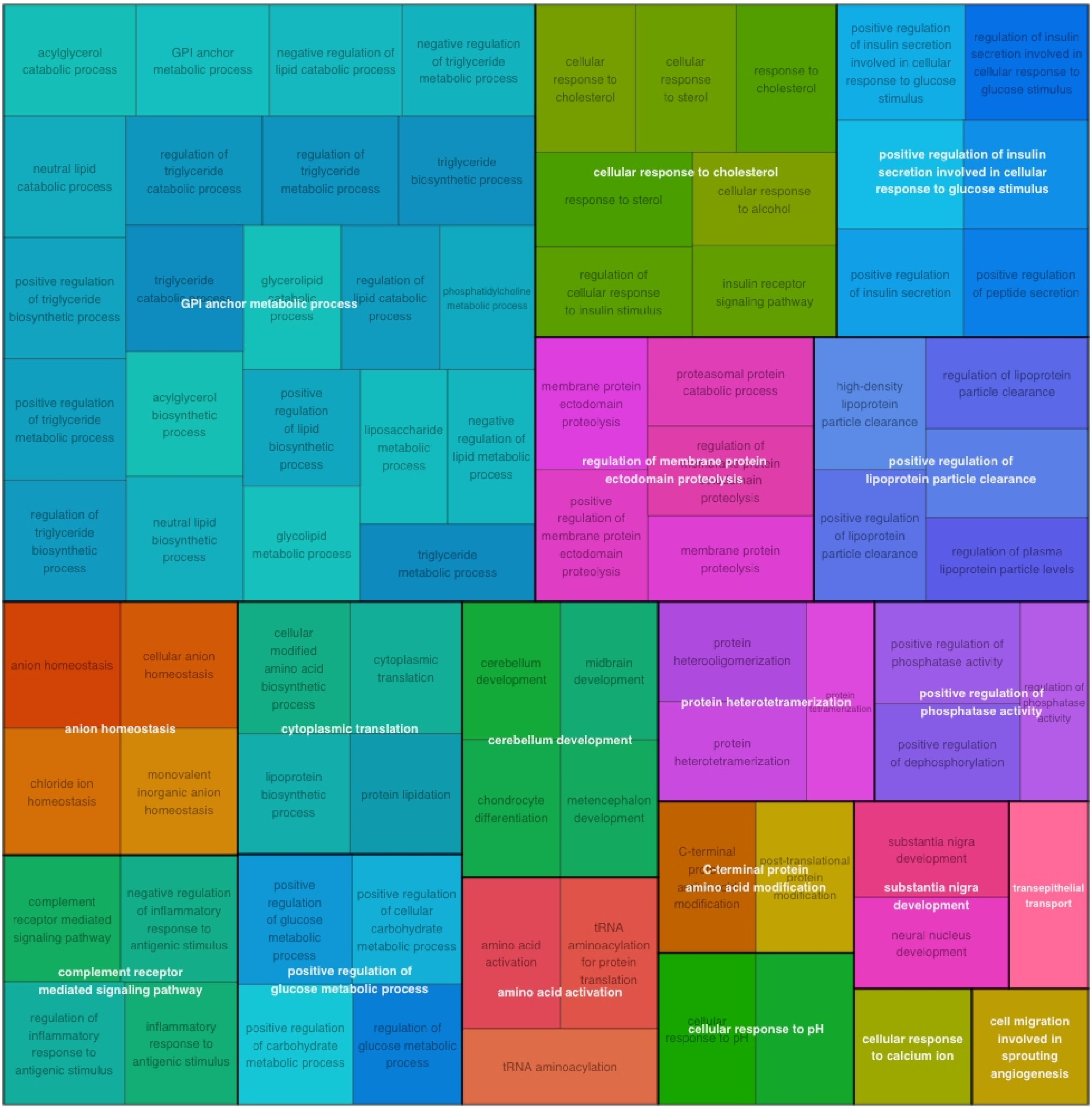


**Figure S4.** Treemap representation of enriched functional categories in upregulated genes from the comparison aEVs post-CAND *vs* aEVs pre-CAND. The size of each box corresponds to the level of enrichment of the respective functional category. Among the categories most enriched highlight biological processes point to an astrocyte-driven neuroprotective response, enhancing metabolic support, lipid and protein homeostasis, neuroimmune regulation, and synaptic plasticity, all of which may help mitigate Parkinson's-related neuronal damage after candesartan treatment. The color-coding represents distinct biological functions, grouping related processes for better visualization.

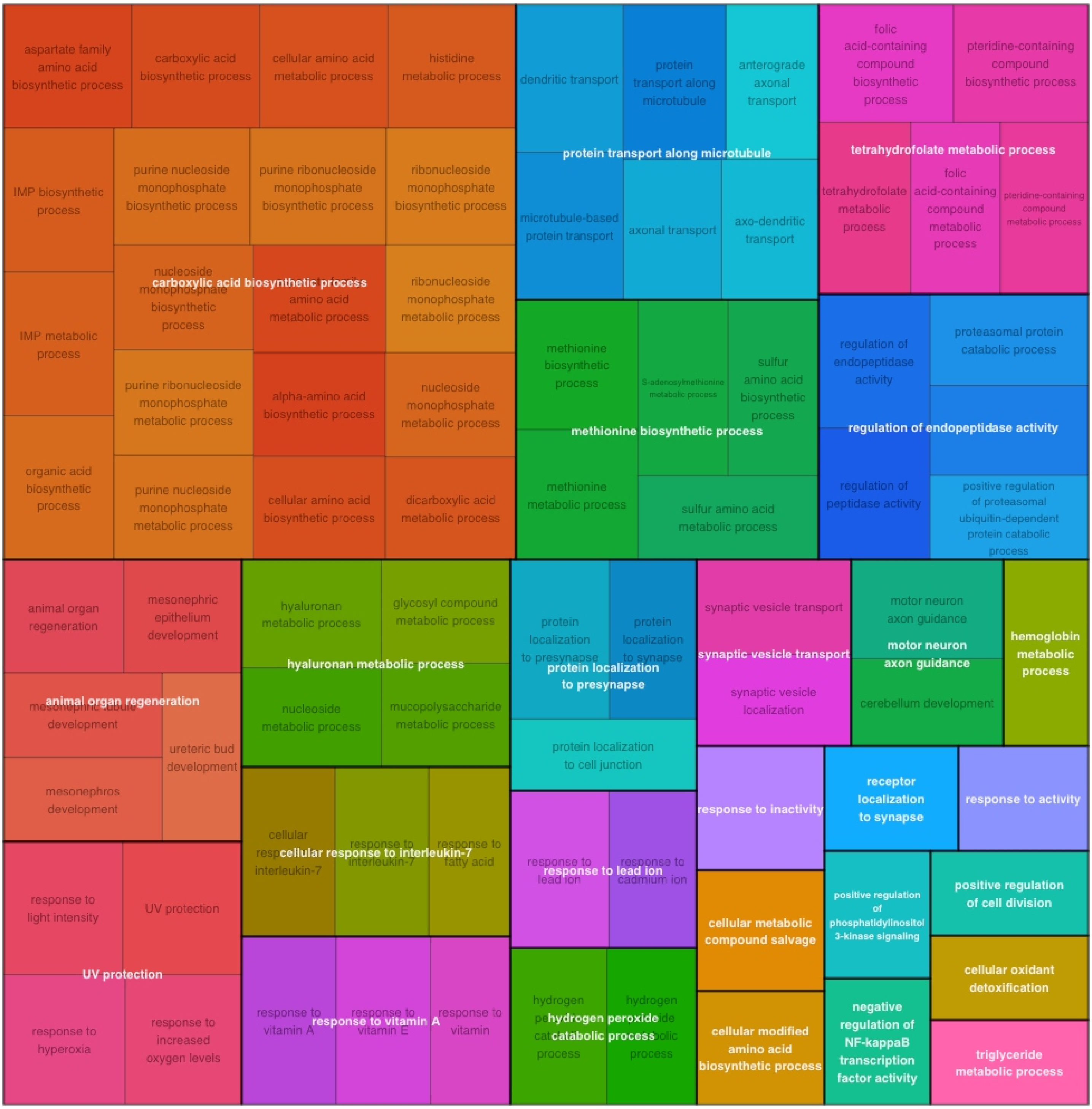


**Figure S5.** Treemap representation of enriched functional categories in downregulated genes from the comparison aEVs post-CAND *vs* aEVs pre-CAND The size of each box corresponds to the level of enrichment of the respective functional category. The largest downregulated categories point to a therapeutic shift towards metabolic efficiency, synaptic stability, reduced neuroinflammation, and oxidative stress control, all of which contribute to a more neuroprotective astrocytic environment following treatment in Parkinson's disease. The color-coding represents distinct biological functions, grouping related processes for better visualization.

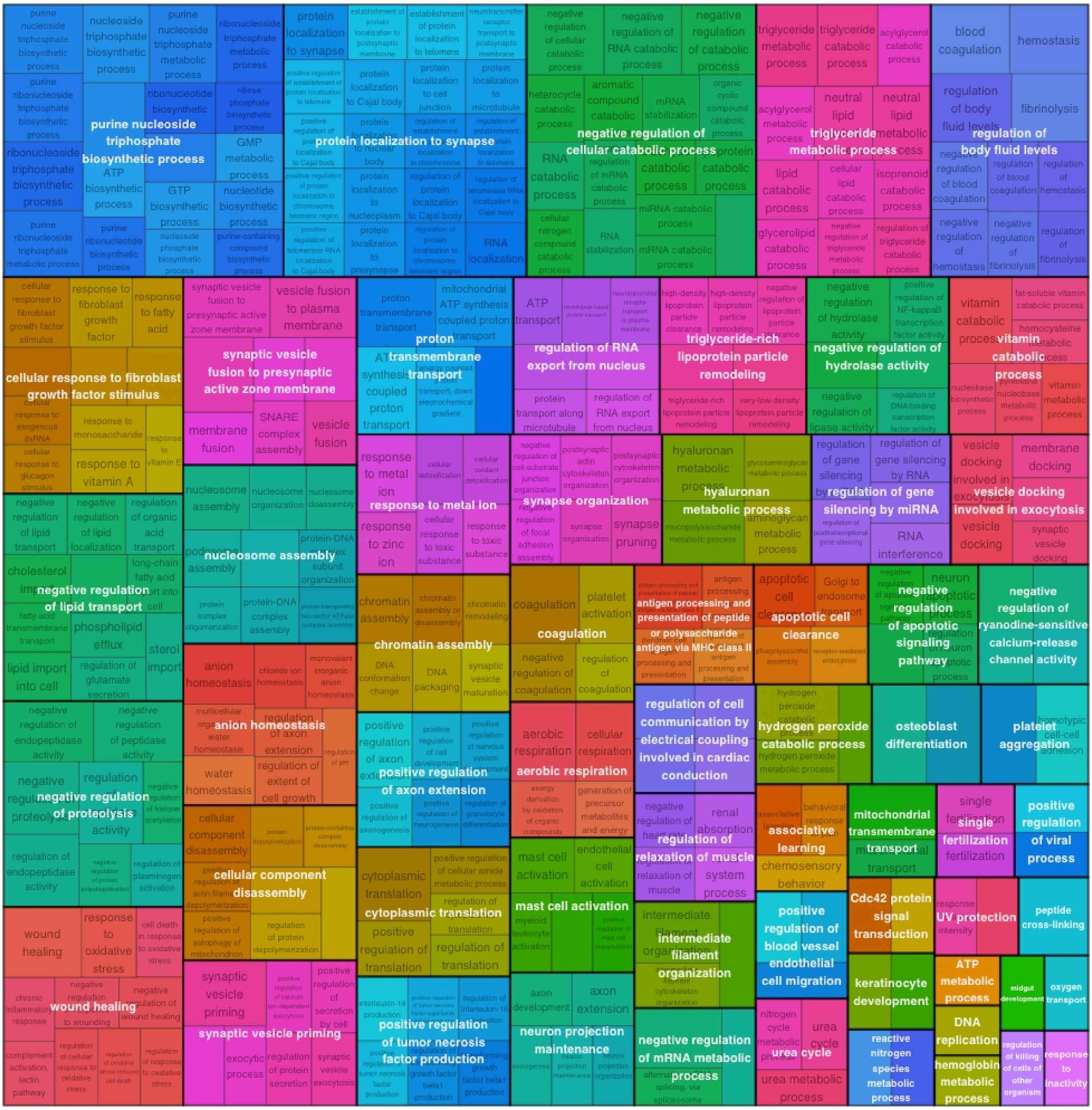


**Figure S6.** Treemap representation of enriched functional categories in upregulated genes from the comparison oEVs post-CAND *vs* oEVs pre-CAND. The size of each box corresponds to the level of enrichment of the respective functional category. Among the categories most enriched highlight biological processes that suggest an adaptive and protective role of oligodendrocytes in PD, enhancing positive regulation of axon extension, synaptic function, metabolic efficiency, and neuroimmune balance. The color-coding represents distinct biological functions, grouping related processes for better visualization.

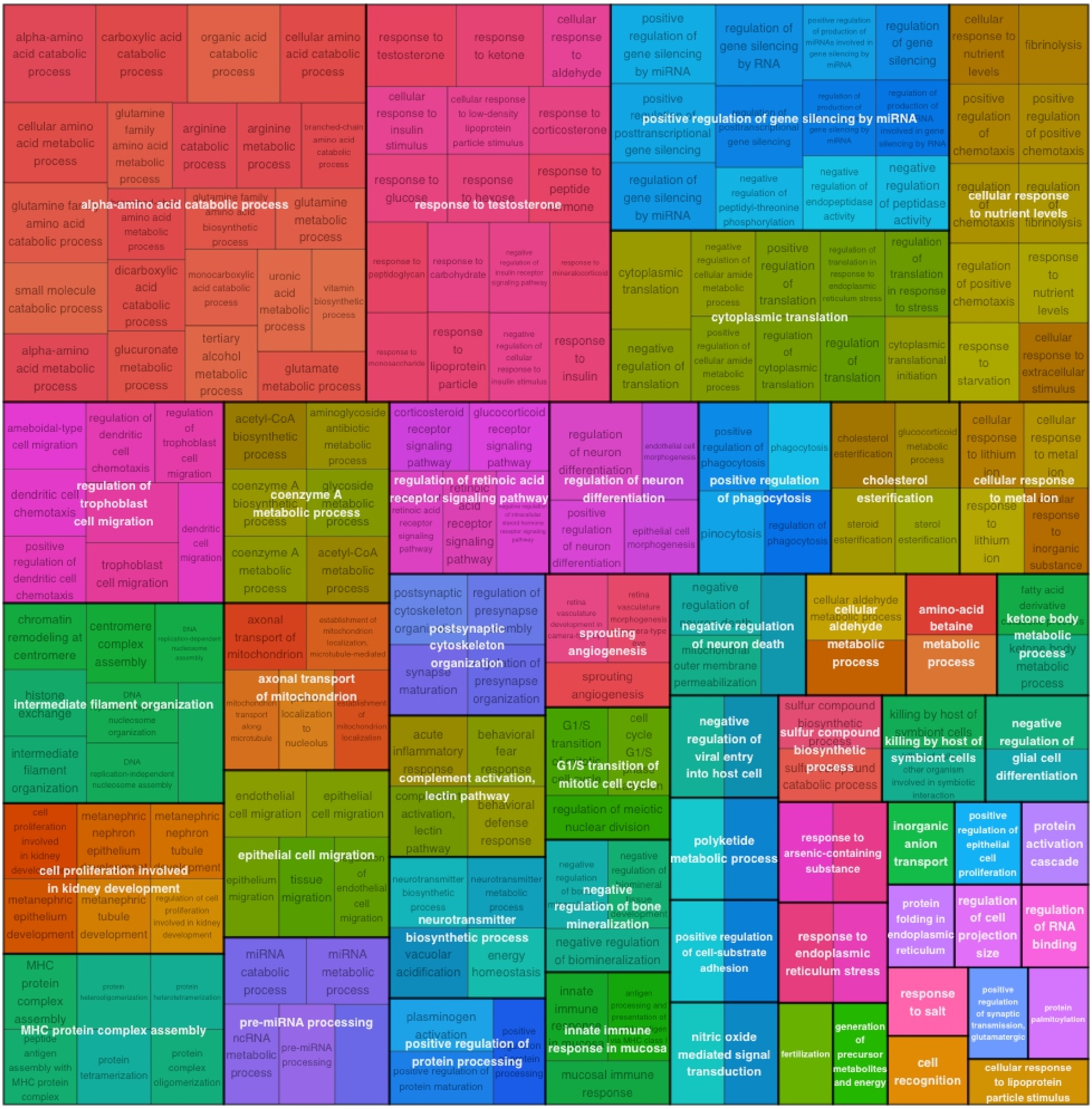


**Figure S7.** Treemap representation of enriched functional categories in downregulated genes from the comparison oEVs post-CAND *vs* oEVs pre-CAND. The size of each box corresponds to the level of enrichment of the respective functional category. The largest downregulated categories include biological processes potentially involved in neuroprotective adaptation in oligodendrocytes, reducing metabolic and oxidative stress, stabilizing gene expression, limiting excessive immune responses, and preventing unnecessary cell proliferation. The color-coding represents distinct biological functions, grouping related processes for better visualization.

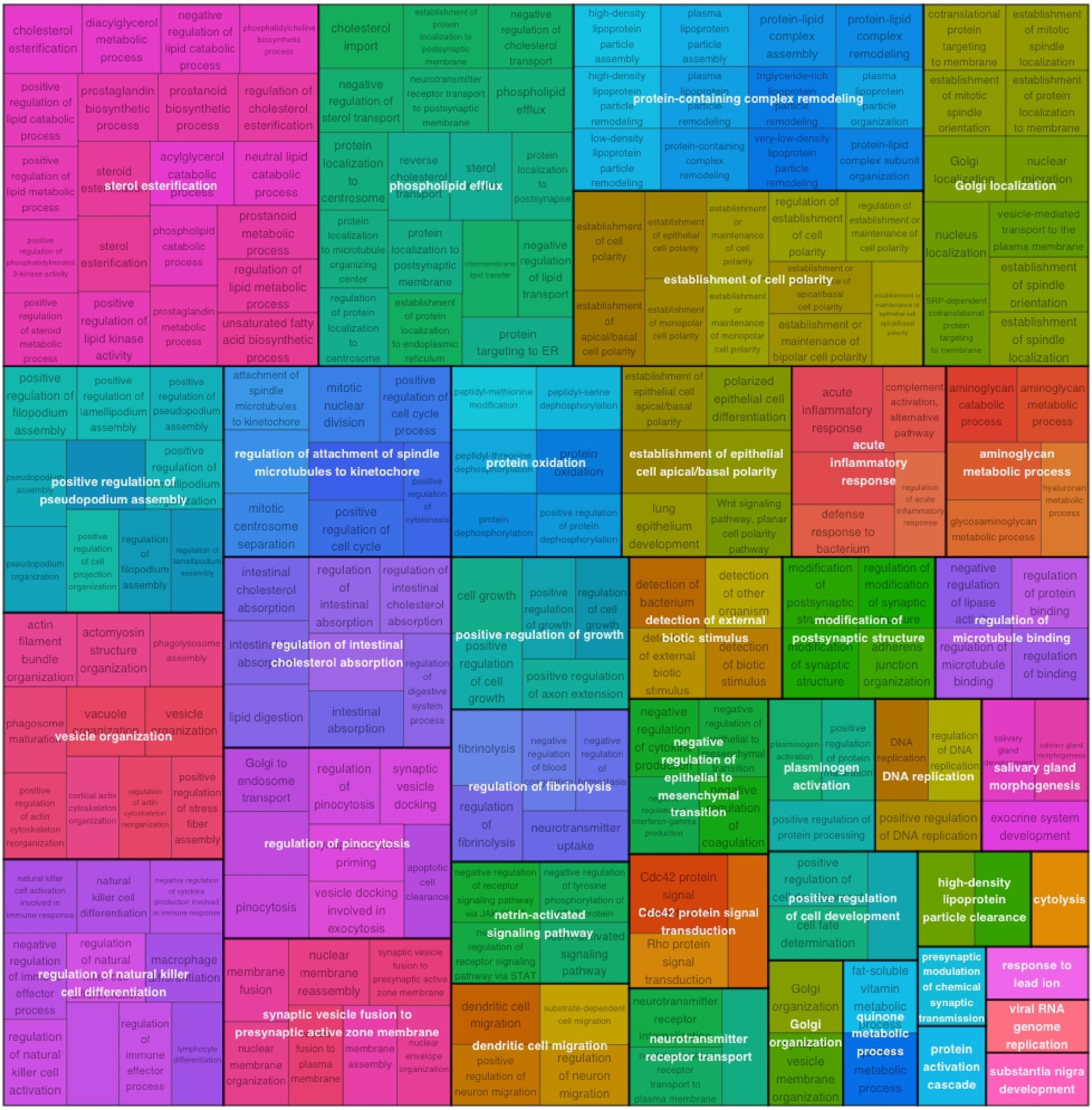


**Figure S8.** Treemap representation of enriched functional categories in upregulated genes from the comparison m/mEVs post-CAND *vs* m/mEVs pre-CAND. The size of each box corresponds to the level of enrichment of the respective functional category. Among the categories most enriched highlight biological processes that suggest that microglia/macrophages are transitioning toward a homeostatic and neuroprotective phenotype, enhancing cellular clearance, lipid homeostasis, synaptic stability, and neuronal support, ultimately contributing to functional recovery and neuroprotection in PD following treatment. The color-coding represents distinct biological functions, grouping related processes for better visualization.

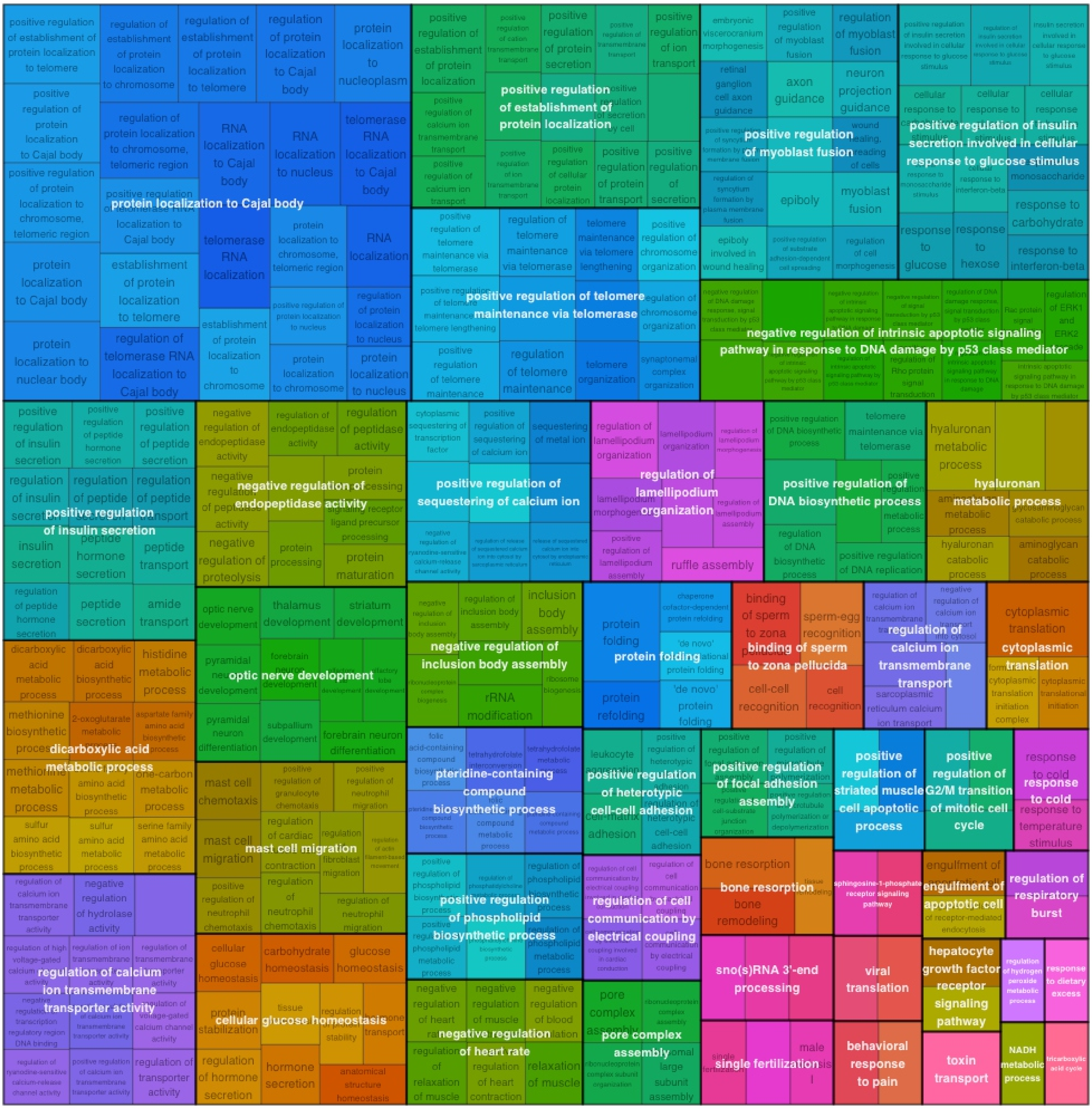


**Figure S9.** Treemap representation of enriched functional categories in downregulated genes from the comparison m/mEVs post-CAND *vs* m/mEVs pre-CAND. The size of each box corresponds to the level of enrichment of the respective functional category. The largest downregulated categories include biological processes, which suggest a transition of microglia/macrophages toward a homeostatic, neuroprotective state, reducing excessive immune activation, metabolic stress, and extracellular remodeling, ultimately contributing to a stabilized neural environment and enhanced neuronal survival in Parkinson's disease after candesartan treatment. The color-coding represents distinct biological functions, grouping related processes for better visualization.
